# Gray-white matter coactivation patterns, cortical receptor correspondence and cross-tissue coordination in major depression

**DOI:** 10.64898/2026.11.27.26369119

**Authors:** Hongzhou Wu, Michael Maes, Jinming Xiao, Yiran Shao, Yuxuan Xing, Chengxiao Yang, Elijah Agoalikum, Drozdstoj Stoyanov, Cheng Luo, Dezhong Yao, Bharat B. Biswal, Benjamin Klugah-Brown

**Author notes:** **Corresponding Author(s): Benjamin Klugah-Brown**, **Bharat B. Biswal**.

## Abstract

Brain function depends on coordinated activity across gray matter (GM) and white matter (WM), yet how this coordination varies with anatomical depth, how it relates to neurochemical architecture, and how it changes in depression remain incompletely understood. We analyzed resting-state fMRI from 2,551 participants at 22 DIRECT Phase II sites, including 1,417 with major depressive disorder (MDD) and 1,134 normal controls. Across 400 parcels, we sampled GM 1 mm outward and WM 1-7 mm inward from their boundary. Joint coactivation pattern (CAP) analysis identified seven shared configurations, including two nearly sign-inverted pairs. Meta-analytic decoding distinguished configurations associated with affective and executive functions, autobiographical memory, social cognition and sensorimotor processing. Similarity across 19 normative cortical receptor and transporter maps explained variation in CAP depth-profile similarity beyond spatial distance in 3.8-49.0% of parcels, depending on the state. Receptor distributions also correlated with mean CAP configurations and their variation with depth. Separate GM and WM state sequences showed small, state-specific differences in co-occurrence and lagged coupling in MDD. Depth-integrated matching above chance was lower in MDD (Hedges’ g = −0.21), and mismatch was higher at 1-5 mm. Normalized mutual information and the adjusted Rand index remained lower through 7 mm. MDD-related shifts in CAP expression corresponded to serotonergic and μ-opioid distributions in a tissue- and state-dependent manner. These findings relate cross-tissue states to normative cortical neurochemistry and identify reduced coordination in depression extending beyond the immediate cortical boundary.

## Introduction

Understanding brain function requires explaining how activity is coordinated across tissue compartments and distributed brain regions [1, 2]. Gray matter (GM) supports local neuronal processing, whereas white matter (WM) provides the axonal pathways through which distributed regions communicate, placing their interface at the intersection of local processing and large-scale integration [3]. Although WM microstructure and its variability have long been linked to cognition and disease [4], and a WM functional-connectome literature has grown steadily [5], most fMRI analyses still treat WM blood oxygenation level-dependent (BOLD) signals as nuisance [6, 7]. Yet WM BOLD signals exhibit reproducible network organization, task-related responses and coordinated fluctuations with GM activity [8–10].

Boundary-aligned analyses identified distinct spatial distributions of GM-WM temporal synchrony and BOLD power contrast [3]. Complementary connectome mapping demonstrated that GM-WM connectivity follows the cortical unimodal-transmodal hierarchy [11]. Incorporating WM BOLD signals changes the inferred community organization of heteromodal cortical systems [12], while GM networks constructed from their connectivity profiles with WM predict individual differences in language performance [13]. In major depressive disorder (MDD), reduced GM-WM connectivity and altered gray-white-gray connectivity have been associated with symptom severity and antidepressant response [14, 15]. Together, these findings motivate a more specific question: how consistently do GM and WM express corresponding activity states over time? Correlations estimated across an entire recording do not reveal whether corresponding spatial configurations occur simultaneously, how often they diverge, or whether one tends to precede the other. This temporal question also has an anatomical dimension: superficial WM networks show stronger correspondence with cortical networks than deeper WM networks [8].

Coactivation pattern (CAP) analysis addresses this question by identifying recurring spatial configurations at individual fMRI time points [16]. CAP-derived state sequences vary with cognitive demands [17, 18]. Studies of current and remitted MDD have identified alterations in state occupancy and transitions, while recent work links asymmetric transitions between visual-attentional and limbic–default configurations to depressive symptom severity [19, 20]. These findings motivate examining whether depression-related state dysregulation also involves the coordination of activity across tissue compartments.

A molecular perspective may further help contextualize the spatial organization of these functional states. Neurotransmitter receptor and transporter distributions provide a regional neurochemical context for brain activity [21]. Network control models show how receptor-informed inputs can alter the predicted accessibility of cognitive activation patterns [22]. Empirical studies have also associated CAP dynamics with PET-measured dopamine D receptor availability, and recent research in MDD has linked altered brain-state energetics to serotonergic and cellular spatial signatures [23, 24]. It remains unclear whether receptor fingerprints provide information about CAP depth profiles beyond spatial distance.

In a large multisite cohort of individuals with MDD and normal controls, we sampled spatially corresponding GM and WM signals along boundary normals, with WM sampling depths ranging from 1 to 7 mm. We combined joint and separate CAP analyses to characterize shared configurations and tissue-specific state expression. We first used meta-analytic functional decoding to characterize the cognitive associations of the shared CAPs. Using normal-control maps, we then examined normative receptor associations with mean activation and depth-related variation. Finally, we assessed MDD-related differences in simultaneous state co-occurrence, lagged coupling and chance-adjusted cross-tissue coordination, alongside receptor associations with disease-related CAP spatial differences, and asked whether these effects persisted into deeper WM, where partial-volume contributions from cortex are smaller.

## Materials and Methods

### Participants and image processing

We analyzed DIRECT Phase II resting-state fMRI and T1-weighted images [25, 26]. Sites diagnosed MDD using ICD-10 or DSM-IV criteria. Local institutional review boards approved the studies, participants provided written informed consent, and data were de-identified. Of 3,001 participants, 442 failed quality control, and eight lacked usable time series, leaving 1,417 with MDD and 1,134 NCs across 22 sites. Complete covariates were available for 2,534 (1,404 MDD, 1,130 NCs) before feature-specific exclusions (**Supplementary Figure S1 and Tables S1–S3**).

Using DPABISurf-preprocessed volumetric BOLD images in MNI space [25, 27], we sampled 400 Schaefer parcels along template boundary normals, 1 mm outward into GM and 1-7 mm inward into WM [28]. Valid GM points could vary with their WM partners’ depth. We applied no additional spatial smoothing. Denoising included motion, cerebrospinal fluid and outlier regressors, 0.01-0.08 Hz filtering and frame censoring. We omitted mean WM and global-signal regressors to preserve cross-tissue covariance (**Supplementary Methods and Table S4**).

### CAP estimation and multisite harmonization

At each depth, k-means pooled 50 random frames per participant. Joint models identified shared configurations across 800 GM and WM values. Separate 400-parcel models yielded tissue-specific sequences. The elbow criterion supported seven states at every depth (**Supplementary Figure S2**). We assigned all retained frames to their nearest centroid and aligned labels to the 1-mm joint solution using centroid correlations and Hungarian matching. For each joint state, we standardized each tissue map across parcels and averaged the two maps per participant. ComBat harmonization modeled site as a batch, preserving diagnosis, age, sex, education, and mean framewise displacement (FD) [29, 30].

### Functional decoding and receptor associations

We estimated NC maps adjusted for age, sex, education, mean FD and site at each depth and summarized shared activation by its mean across depths (A) and its linear slope per millimeter (L). Both quantify relative expression. We decoded A using Spearman correlations with 123 Neurosynth term maps [31, 32]. We assessed significance with 10,000 hemisphere-preserving spins and Benjamini-Hochberg false discovery rate (FDR) correction across all 861 tests [33].

For each CAP and seed parcel, we modeled pairwise similarity of seven-depth profiles as a function of linear Euclidean distance, then added the correlation between the parcels’ 19-target cortical receptor and transporter profiles (**Supplementary Figure S3**) [21]. Gain was the increase in adjusted R². We tested positive gains against a spatial null and corrected across 2,800 CAP-by-parcel tests. We also correlated A and L with individual receptor maps, correcting across 133 tests per phenotype. Partial least squares (PLS) regression identified multireceptor associations [34]. Whole-cortex components were spin-tested with FDR correction within each target. Local correlations, spatial-block bootstrap estimates and fivefold spatial cross-validation assessed localization, weight stability and generalization (**Supplementary Methods**).

### Cross-tissue coordination and group comparisons

From aligned separate sequences, we estimated contemporaneous co-occurrence, mismatch conditional on the state in either tissue, and one-frame-lag probabilities in both directions.

Directional asymmetry was the WM-leading minus GM-leading probability for the same ordered state pair. Lags used original acquisition indices and excluded censoring gaps. Overall coordination was assessed by matched-state concordance, mismatch, chance matching from marginal occupancies, matching above chance, Cohen’s κ, normalized mutual information (NMI) and the adjusted Rand index (ARI). NMI and ARI are invariant to label permutations.

We summarized coordination by the trapezoidal area under the depth curve (AUC) divided by the observed depth span (6 mm for a complete profile) and by the linear coefficient of an individual quadratic fit to uncentered depth. Robust models adjusted for age, sex, education and site. Repeated-depth models used participant-clustered covariance and categorical depth. State-pair comparisons used HC3 robust covariance. We report two-sided MDD-minus-NC contrasts, adjusted Hedges’ g and FDR-adjusted q values. Correction families are specified in **Supplementary Methods**.

Finally, we correlated signed MDD-minus-NC Hedges’ g maps for depth-averaged separate GM and WM activation with the normative cortical receptor maps. These parcel-wise group models additionally adjusted for mean FD. Inference used 10,000 spins and FDR correction across 133 tests per tissue. We repeated this analysis with absolute effects to assess correspondence with effect magnitude.

## Results

### Shared configurations and tissue-specific sequences

Joint clustering identified seven shared configurations. Separately clustered GM and WM sequences could diverge at individual frames (Figure 1). ComBat reduced site effects (median reduction in site partial η², 0.0351), preserving the diagnosis coefficient’s direction for most features (**Supplementary Figure S4**).

**Figure 1.**
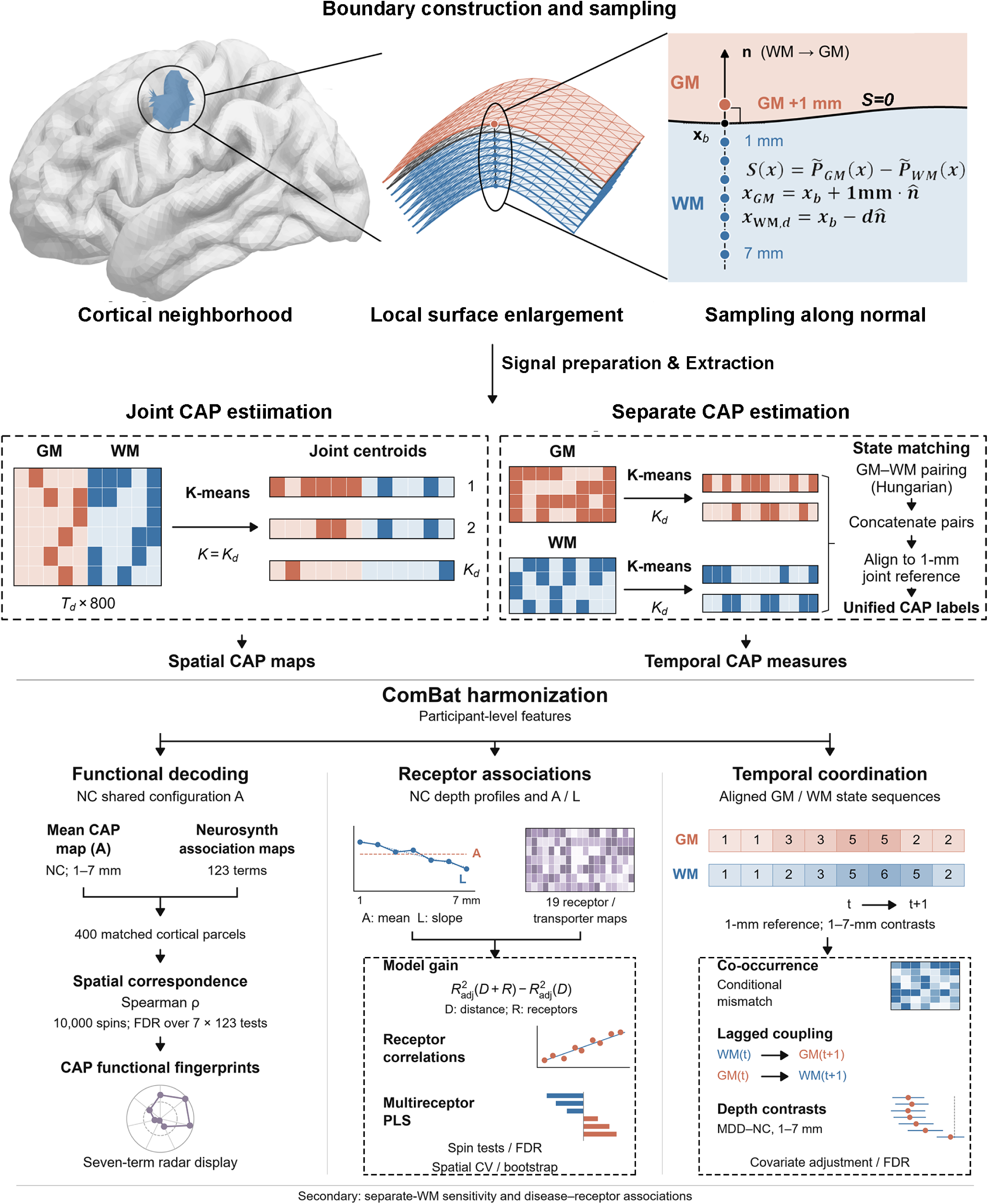
Workflow for gray-white matter CAP analysis and functional, molecular, and temporal characterization. Workflow arrows indicate analysis steps; arrows between time points denote temporal ordering. A, mean configuration across depths; L, linear depth slope; NC, normal controls; MDD, major depressive disorder; PLS, partial least squares; FDR, false discovery rate.

### Spatial and functional organization of GM-WM coactivation patterns

Seven depth-averaged NC maps included two near-inverse pairs (Figure 2). CAP1 showed positive expression in the frontal operculum and temporal regions and negative expression in many occipital parcels. CAP2 showed the reverse (r = −0.990), consistent with previous CAP studies [35, 36]. CAP3 emphasized temporal, parietal and prefrontal default mode regions. CAP4 emphasized visual cortex, and CAP5 emphasized parietal control and opercular regions (r = −0.479). CAP6 emphasized dorsal attention, sensorimotor and visual regions. CAP7 emphasized posterior cingulate, precuneus and lateral default mode regions, nearly inverting CAP6 (r = −0.995) and resembling CAP3 (r = 0.840).

**Figure 2.**
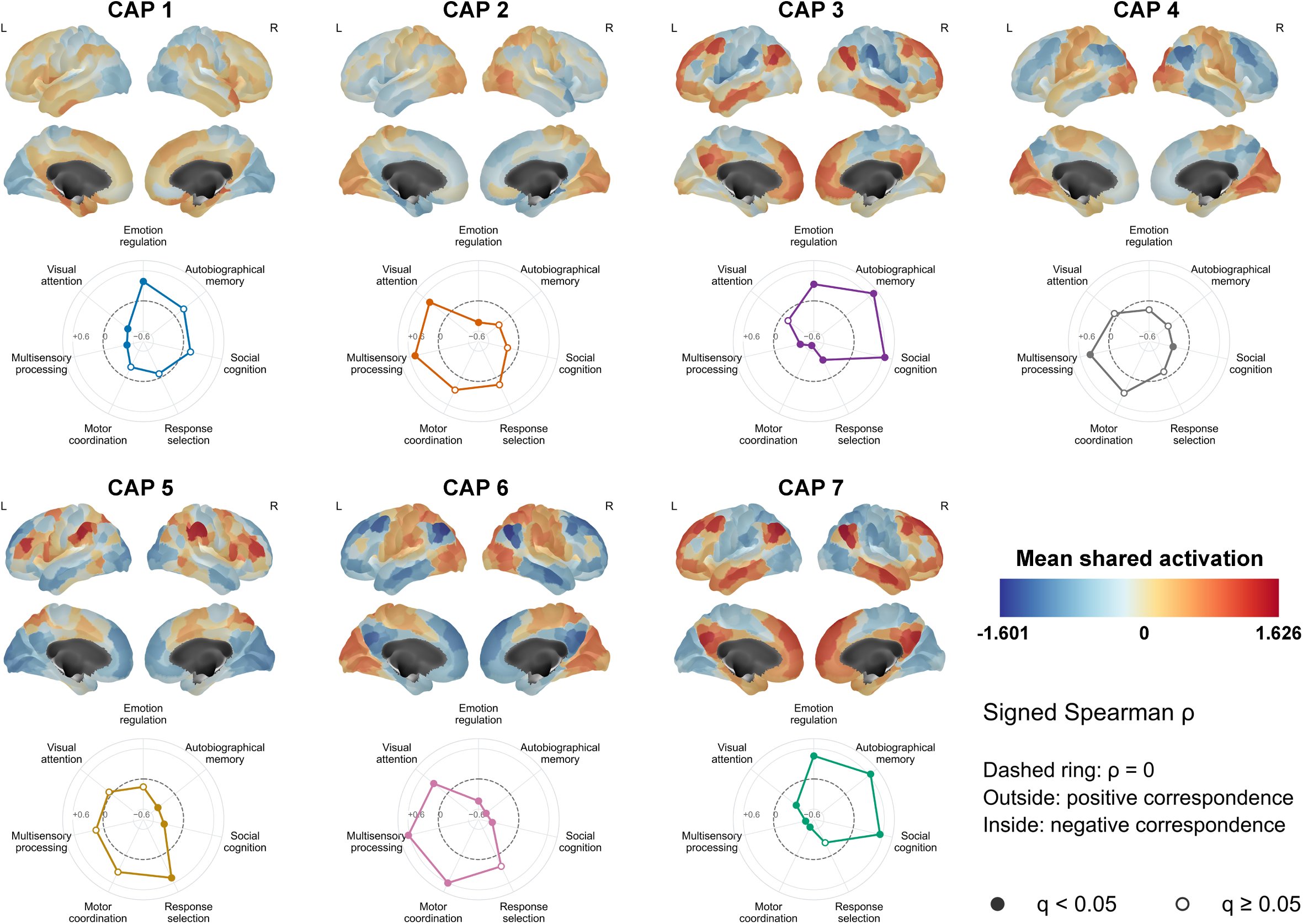
Spatial configurations and meta-analytic functional profiles of shared GM-WM CAPs. For each CAP, cortical maps show covariate-adjusted normal-control mean shared GM-WM activation across WM sampling depths of 1-7 mm (phenotype A). Values represent relative spatial expression after standardizing participant maps across parcels and harmonizing with ComBat, adjusted for age, sex, education, mean framewise displacement, and site. Lateral views are shown above medial views; all maps share the displayed color scale. Radar plots show signed Spearman correlations between each CAP map and seven representative Neurosynth meta-analytic association-test maps across 400 Schaefer parcels. The axes indicate emotion regulation, autobiographical memory, social cognition, response selection, motor coordination (reference term: coordination), multisensory processing (multisensory), and visual attention. The dashed ring indicates ρ = 0; points outside and inside this ring indicate positive and negative correspondence, respectively. All plots use the same scale (ρ = −0.8 to 0.8). Filled markers indicate q < 0.05 after 10,000 two-sided, null-mean-centered spatial spin permutations and BH-FDR correction across all 7 × 123 CAP-by-term tests; open markers indicate q ≥ 0.05. L, left; R, right.

Mean correspondence between matched GM and WM centroids declined with depth. Every pair exceeded the prespecified correlation threshold of 0.30 (**Supplementary Figures S5–S9**). Decoding identified 267 significant associations across 861 tests (Supplementary Tables S5–S6). CAP1 correlated positively with emotion regulation (ρ = 0.386) and negatively with multisensory processing and visual attention (ρ = −0.466 and −0.407). CAP2 showed the reverse. CAP3 correlated with autobiographical memory and social cognition (ρ = 0.717 and 0.645) and inversely with motor coordination (ρ = −0.705). CAP4 favored perception and multisensory processing over cognitive control and memory retrieval. CAP5 correlated positively with response selection (ρ = 0.491) and negatively with autobiographical memory and social cognition. CAP6 emphasized multisensory processing and motor coordination; CAP7 emphasized autobiographical memory.

Across all terms, CAP1-CAP2 and CAP6-CAP7 were nearly inverse (r = −0.993 and −0.997), and CAP3 resembled CAP7 (r = 0.950). Despite partial spatial opposition, CAP4 and CAP5 had dissimilar functional profiles (r = 0.051; **Supplementary Figures S10–S11**). The elbow criterion selected seven states at all seven depths in each site- and diagnosis-stratified subsample (10%, 30%, and 50% of participants). Of the 147 joint centroids estimated across subsamples and depths, 129 correlated most strongly with the full-sample centroid of the same state. Six CAPs retained the direction of all seven representative Neurosynth associations in every subsample (126/126 comparisons), and Spearman correlations between the subsample and full-sample 123-term profiles for CAP4 ranged from 0.563 to 0.709 (**Supplementary Figure S12 and Table S7**).

### Multireceptor contributions to CAP depth-profile similarity

All seven states showed significant gains over the distance-only model (Figure 3). Gains occurred in 196/400 parcels for CAP3 (49.0%), 187 for CAP5 (46.8%), 178 for CAP2 (44.5%), 152 for CAP7 (38.0%), 126 for CAP1 (31.5%), 46 for CAP6 (11.5%) and 15 for CAP4 (3.8%) (**Supplementary Figure S13**).

**Figure 3.**
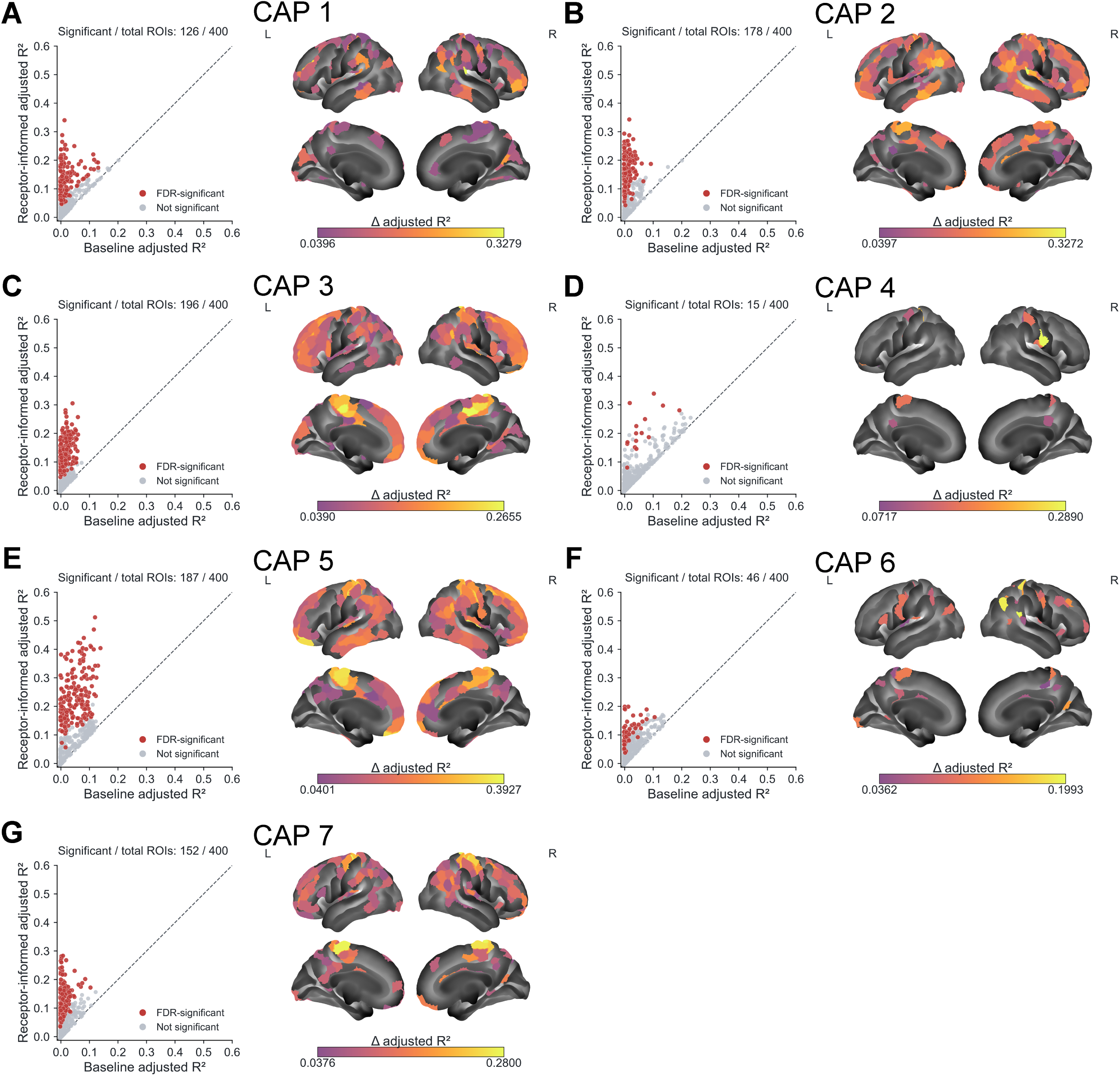
Multireceptor information improves regional models of CAP depth-profile similarity beyond spatial distance. (A-G) Results for coactivation patterns CAP1-CAP7, respectively. Left: Each point represents one seed parcel, comparing adjusted R² between the two models. The dashed line indicates equal model fit. Red points indicate significant positive model gains; gray points indicate parcels that do not meet this criterion. Labels report the number of significant parcels out of 400. Model gain was defined as Δadjusted R² = adjusted *R*²*_receptor-informed_* – adjusted *R*²*_baseline_*. Significance was assessed using 10,000 hemisphere-preserving spatial spin permutations with a one-sided test, followed by Benjamini-Hochberg false discovery rate correction across all 2,800 CAP-by-parcel tests (q < 0.05). Right: Cortical maps display significant positive model gains, with lateral views above and medial views below. Colors from purple to yellow indicate increasing gain; gray regions do not meet the significance criterion. Color limits are determined separately for each CAP from the minimum and maximum gains among its significant parcels. L, left hemisphere; R, right hemisphere.

Mean gain was largest in CAP5 (adjusted R², 0.049 to 0.150), followed by CAP2 (gain, 0.083) and CAP3 (0.070). CAP5 peaked in the left lateral prefrontal control cortex (gain, 0.393). CAP1, CAP2, CAP3 and CAP7 peaked in the somatomotor cortex (**Supplementary Tables S8–S9**).

CAP2 and CAP3 each showed significant gains in 42/52 control-network parcels (80.8% per CAP). CAP5 gains involved 54/77 somatomotor and 55/91 default mode parcels. Somatomotor coverage reached 51/77 in CAP7. In CAP4, 11 of the 15 significant parcels were somatomotor (**Supplementary Table S10**).

### Receptor associations with shared GM-WM CAP organization

Depth-averaged expression (A) showed 17 receptor associations across six CAPs (Figure 4A and Supplementary Table S11). The μ-opioid receptor (MOR) correlated positively with CAP1 and CAP7 (ρ = 0.589 and 0.472) and negatively with CAP2, CAP4 and CAP6 (ρ = −0.576, −0.502 and −0.517). CAP4 and CAP6 also correlated negatively with 5-HT2A, 5-HT4 and CB1. CAP3 and CAP7 correlated positively with 5-HT2A and negatively with the norepinephrine transporter (NET), with the opposite pattern for CAP6. No individual association survived correction for CAP5-A.

**Figure 4.**
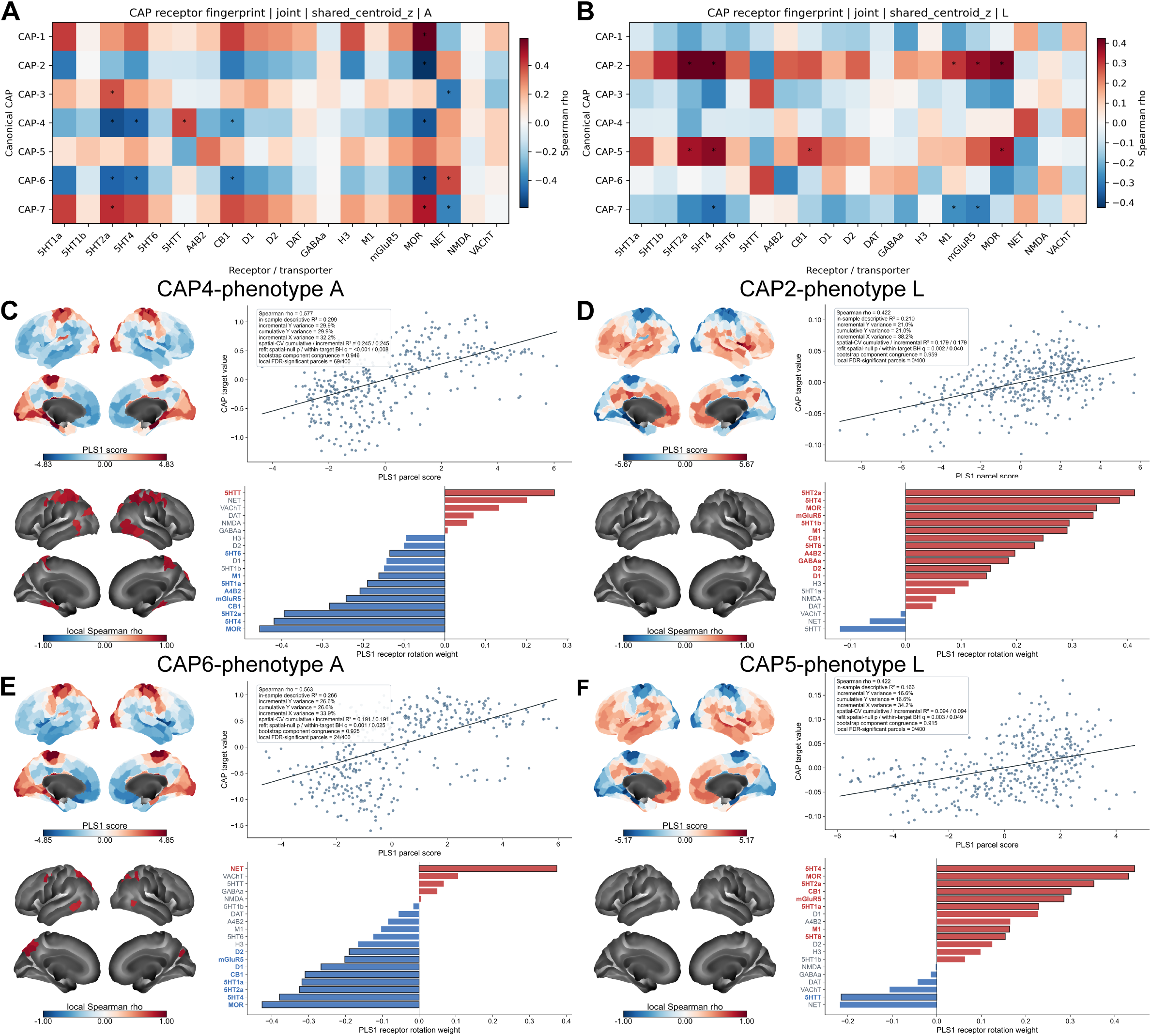
Receptor fingerprints and multireceptor associations of shared GM-WM CAP organization. Analyses used covariate-adjusted NC maps. Phenotype A represents mean shared gray matter-white matter (GM-WM) activation across sampling depths of 1-7 mm, whereas phenotype L represents the linear slope with depth. (A, B) Spearman correlations between seven coactivation patterns (CAPs) and 19 receptor/transporter maps for phenotypes A and L, respectively. Asterisks indicate significance following spatial spin testing and false discovery rate (FDR) correction (q < 0.05). (C-F) First partial least squares components (PLS1) for CAP4-A, CAP2-L, CAP6-A, and CAP5-L, respectively. Each panel displays unthresholded component-score maps (upper left), score-target associations across 400 cortical parcels (upper right), FDR-significant local correlations (lower left), and receptor/transporter rotation weights (lower right). Red and blue bars indicate positive and negative weights; bold labels and outlined bars identify bootstrap-stable weights (q < 0.05).

The depth slope (L) showed 12 associations, all in CAP2, CAP5 and CAP7 (Figure 4B). CAP2 and CAP5 correlated positively with 5-HT2A, 5-HT4 and MOR (ρ = 0.353–0.424). Additional positive associations involved M1 and mGluR5 for CAP2 and CB1 for CAP5. CAP7 correlated negatively with 5-HT4, M1 and mGluR5. The positive CAP1–MOR and opposing CAP6/CAP7–5-HT2A associations persisted at every depth (**Supplementary Figures S14–S16**).

The first PLS component (PLS1) explained 29.92% of CAP4-A and 26.63% of CAP6-A variance (q = 0.0076 and 0.0247, spatial cross-validated R² = 0.245 and 0.191, respectively) (Figure 4C–F). Both showed stable negative weights for MOR, 5-HT4 and 5-HT2A. Positive weights included the serotonin transporter (5-HTT) for CAP4-A and NET for CAP6-A. For CAP2-L and CAP5-L, corresponding values were 20.96% and 16.60% (q = 0.0399 and 0.0494, cross-validated R² = 0.179 and 0.094), with stable positive weights for 5-HT2A, 5-HT4 and MOR.

Local correlations survived correction at 69 centers for CAP4-A (mainly dorsal attention, somatomotor and visual cortex) and 24 for CAP6-A (including posterior parietal, visual and posteromedial cortex). Neither slope component had significant local centers. Fourteen components met the whole-cortex criterion (**Supplementary Figure S17 and Table S12**).

### Altered GM-WM CAP co-occurrence and directional coupling in MDD

At 1 mm, co-occurrence differed between groups for 12 state pairs (Figure 5A and **Supplementary Table S13**). Same-state co-occurrence was lower in MDD for CAP1 and CAP2 (g = −0.248 and −0.259) and higher for CAP3 and CAP4 (g = 0.147 and 0.158). GM CAP5 co-occurred less often with WM CAP1 (g = −0.213), whereas WM CAP5 co-occurred more often with GM CAP3, CAP4 and CAP6 (g = 0.177–0.199).

**Figure 5.**
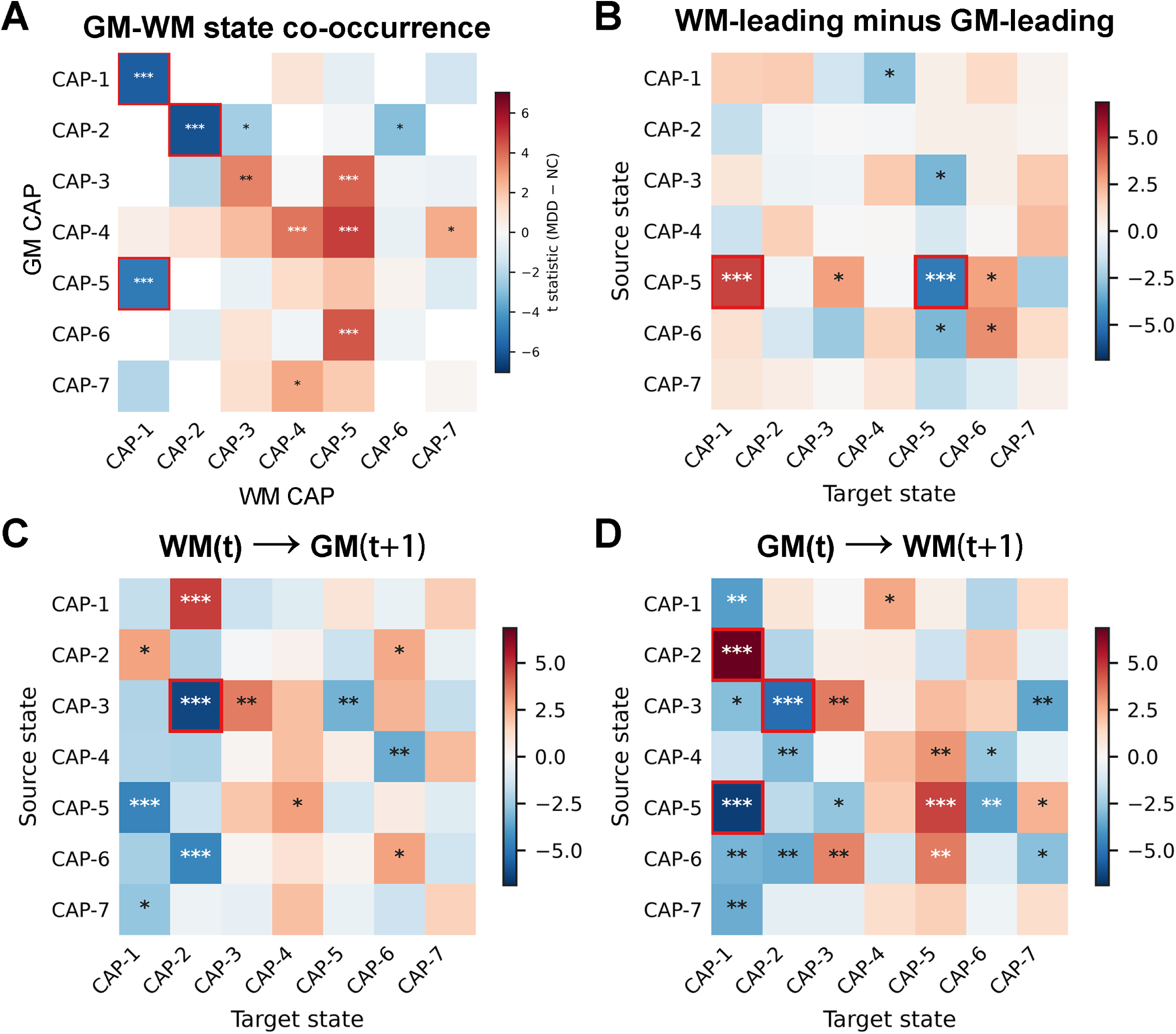
Group differences in gray-white matter CAP co-occurrence and lagged coupling. Heatmaps compare MDD and NC at a WM sampling depth of 1 mm. (A) Simultaneous co-occurrence frequencies of GM and WM CAPs. (B) Directional asymmetry, defined as WM-leading minus GM-leading conditional probabilities. (C, D) One-frame-lag conditional probabilities for WM(t) → GM(t+1) and GM(t) → WM(t+1), respectively; rows indicate source states and columns indicate target states. Colors represent MDD-NC t-statistics after ComBat harmonization and adjustment for age, sex, education, and site; red and blue indicate higher and lower MDD values, respectively. Asterisks indicate Benjamini-Hochberg FDR significance (q < 0.05, q < 0.01, and q < 0.001 for one, two, and three asterisks, respectively). Red outlines identify effects with |Hedges’ g| > 0.20. Arrows indicate temporal ordering rather than causal influence.

At one frame, 12 WM-leading and 21 GM-leading probabilities differed (Figure 5C–D and Supplementary Table S14B–C). CAP3→CAP2 coupling decreased (WM-leading g = −0.264, GM-leading g = −0.219), as did CAP5→CAP1 coupling (−0.192 and −0.276). GM CAP2→WM CAP1 and GM CAP5→WM CAP5 coupling increased (g = 0.248 and 0.196). Asymmetry differed for eight pairs (Figure 5B and Supplementary Table S14A). For CAP5→CAP1, the GM-leading probability fell more, increasing asymmetry (g = 0.200). For CAP5→CAP5, increased GM-leading coupling reduced asymmetry (g = −0.204). Both effects persisted across depths, as did reductions in CAP1 same-state and GM CAP5–WM CAP1 co-occurrence and in WM CAP3→GM CAP2 and GM CAP5→WM CAP1 lagged coupling (**Supplementary Figures S18–S19**).

CAP5 mismatch decreased in MDD conditional on GM CAP5 at 2–4 and 6–7 mm, but increased conditional on WM CAP5 at 1–3 mm. Frame-weighted analyses confirmed both findings (**Supplementary Figure S20 and Table S15**).

### Reduced GM-WM state coordination across sampling depths in MDD

Depth-integrated matching above chance was lower in MDD (adjusted AUC, 0.3859 versus 0.3965 in NCs, g = −0.205, q = 4.22 × 10□□, Figure 6A). Concordance decreased, and mismatch increased (g = −0.148 and 0.148, both q = 4.69 × 10□). Cohen’s κ, NMI and ARI were also lower (g = −0.181, −0.183 and −0.172; all q ≤ 6.43 × 10□□) (**Supplementary Table S16**).

**Figure 6.**
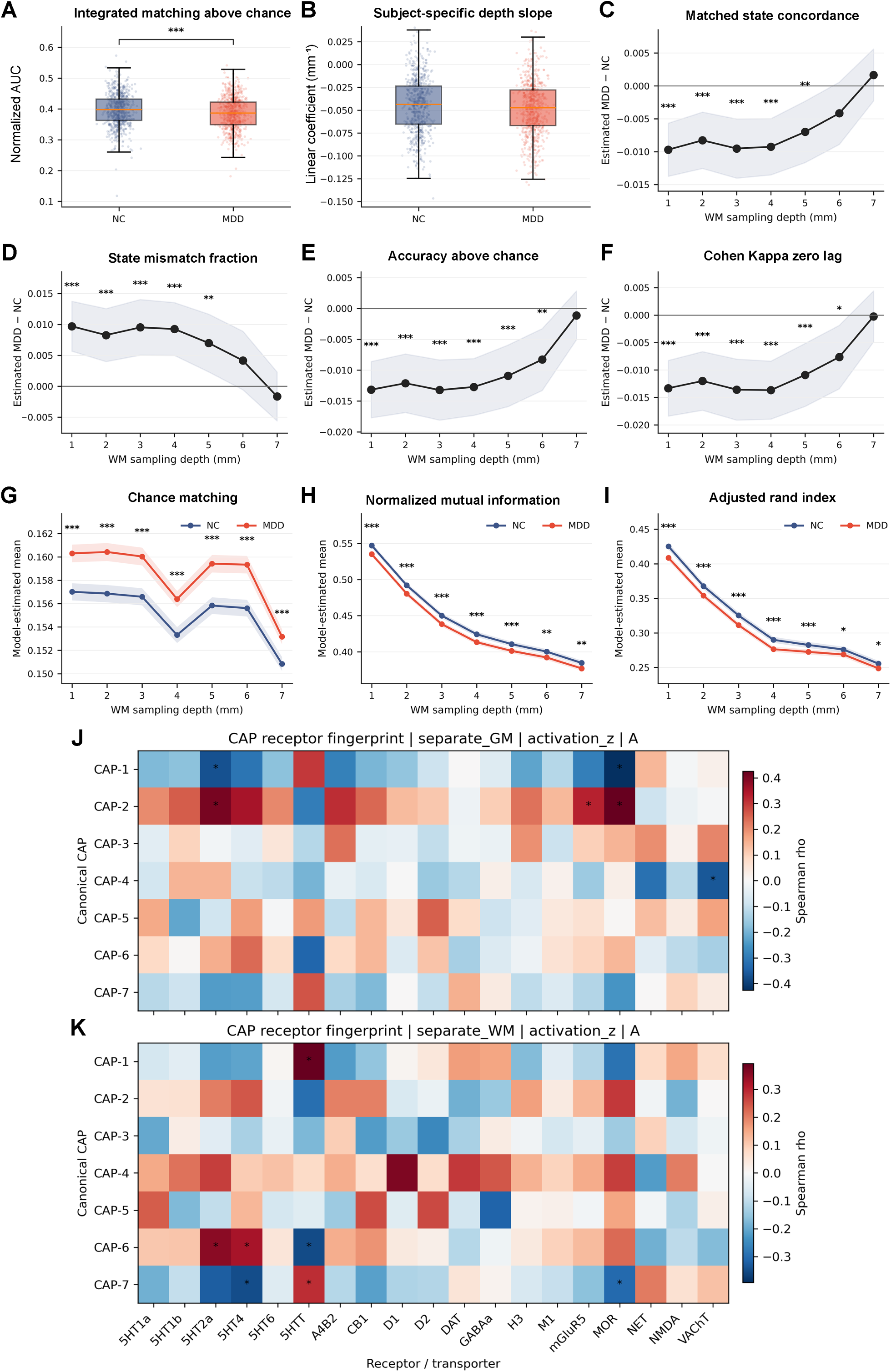
GM–WM state coordination across sampling depths and receptor associations of CAP group differences. (A, B) Normalized area under the curve for matching above chance and the linear coefficient from individual quadratic depth fits across 1–7 mm, respectively. Boxes show medians and interquartile ranges, whiskers extend to observations within 1.5 times the interquartile range, and dots represent participants. (C–F) Depth-specific MDD-minus-NC differences in matched-state concordance, mismatch fraction, matching above chance and zero-lag Cohen’s κ. (G–I) Adjusted group means for chance matching, normalized mutual information and adjusted Rand index (blue for NC, red for MDD). Shading indicates 95% confidence intervals. Panels A–I use ComBat-harmonized measures adjusted for age, sex, education and site (*q < 0.05, **q < 0.01, ***q < 0.001). Depth-specific tests were corrected across seven depths per metric. (J, K) Spearman correlations of signed, depth-averaged CAP group differences (phenotype A, expressed as Hedges’ g for MDD minus NC) in GM (J) and WM (K) with 19 normative cortical receptor/transporter maps across 400 parcels. Group effects were additionally adjusted for mean framewise displacement. Red and blue indicate positive and negative correlations, respectively. Asterisks denote q < 0.05 after 10,000 spatial-spin permutations and BH-FDR correction across 7 CAPs × 19 targets within each tissue.

Concordance was lower and mismatch higher at 1–5 mm. At 1 mm, adjusted concordance was 0.6421 in MDD and 0.6518 in NCs (difference −0.0097, 95% CI −0.0137 to −0.0057). Matching above chance and κ remained lower in MDD through 6 mm. Chance matching was higher at all depths (AUC g = 0.270, q = 8.42 × 10 ¹□□). NMI and ARI declined with depth in both groups and remained lower in MDD at 7 mm (adjusted differences −0.0076 and −0.0069, q = 0.0026 and 0.0159, respectively) (Figure 6C–I and **Supplementary Table S17**).

Diagnosis-by-depth interactions survived correction for six of seven outcomes (F□, □□□□ = 4.41–7.72, all q ≤ 2.26 × 10□□), with NMI the exception (q = 0.1483). Adjustment for site alone yielded the same six interactions (**Supplementary Figure S21**). No group difference in the linear coefficient of individual quadratic depth fits survived correction (all q ≥ 0.0785, Figure 6B and **Supplementary Table S16**).

### Receptor associations of GM and WM CAP alterations in MDD

Signed depth-averaged MDD-minus-NC maps showed six associations in GM and seven in WM (Figure 6J–K and Supplementary Table S18). GM CAP1 effects correlated negatively with 5-HT2A and MOR (ρ = −0.370 and −0.425), and CAP2 positively (ρ = 0.403 and 0.425, all q = 0.0199), reflecting two poles of one spatial mode. CAP2 also correlated positively with mGluR5 (ρ = 0.328), and CAP4 correlated negatively with the vesicular acetylcholine transporter (VAChT, ρ = −0.360), both with q = 0.0310.

In WM, the receptor maps describe the overlying cortical parcels. CAP1 effects correlated positively with 5-HTT (ρ = 0.392). CAP6 effects correlated positively with 5-HT2A and 5-HT4 and negatively with 5-HTT (ρ = 0.355, 0.323 and −0.351). CAP7 showed the opposite associations with 5-HT4 and 5-HTT (ρ = −0.349 and 0.288) and a negative MOR association (ρ = −0.303).

Opposing GM CAP1/CAP2 associations with 5-HT2A and MOR persisted across all depths. GM CAP2–mGluR5 and CAP4–VAChT associations survived at every depth except 2 mm. WM CAP1–5-HTT correspondence persisted at 2–7 mm and negative CAP7–5-HT4 correspondence at 3–7 mm (Supplementary Figure S22). Absolute effect maps yielded no significant depth-averaged associations (**Supplementary Table S18**).

## Discussion

We used sampling across the gray-white matter boundary at multiple depths to characterize seven shared coactivation patterns. Mean CAP configurations and their depth-related variation showed distinct, state-specific receptor associations. MDD was associated with small but consistent changes in state co-occurrence and lagged coupling, alongside lower overall GM-WM coordination.

### Functional organization of shared GM-WM coactivation patterns

Functional contrast across the GM-WM boundary follows patterns of sensorimotor organization, myelination, and neurochemical architecture [3]. Our findings extend this perspective by resolving the boundary signal into recurring spatial states. This complements the original CAP framework, in which brief instances of coordinated activity reveal configurations obscured by temporal averaging [16, 18].

The posterior cingulate/precuneus, lateral parietal, and temporal default mode regions showed increased relative activation in CAP3 and CAP7. These territories overlap systems implicated in autobiographical reconstruction and representations of self and others [37, 38]. Conversely, the dorsal attention, visual, and somatomotor expression of CAP6 aligns it with externally oriented processing. The near-inverse CAP6-CAP7 pair likely reflects the two poles of a single sensorimotor-to-default axis [39], a sign symmetry commonly produced by CAP clustering [35].

CAP1 and CAP2, the second near-inverse pair, expressed a complementary distinction between affective regulation and external sensory attention. This contrast is compatible with evidence implicating frontal opercular regions in emotional processing [40], lateral prefrontal and temporal regions in cognitive reappraisal [41], and occipital visual cortex in attentional modulation [42]. Subsampling preserved both the affective–sensory contrast between CAP1 and CAP2 and the memory–sensorimotor contrast between CAP7 and CAP6. Despite their partial spatial opposition, CAP4 and CAP5 showed little similarity across their functional fingerprints, distinguishing perceptual organization from response selection and planning. CAP5’s parietal control and opercular/attention-related expression are compatible with the contribution of frontoparietal systems to executive performance [43]. The shared expression of these states across tissues motivates examining whether regions with similar receptor profiles exhibit corresponding CAP organization as sampling proceeds into WM.

### Receptor associations with CAP spatial organization and depth variation

Multireceptor similarity contributed information about CAP depth-profile similarity beyond the linear Euclidean-distance baseline in every state (Figure 3). Because receptor similarity decays non-linearly with distance, these gains are best regarded as an upper bound on the receptor-specific contribution. Receptor fingerprints correspond to structural connectivity, functional coupling, and cognitive topographies [21], while network-control modeling has demonstrated how receptor-informed inputs can alter modeled transitions between cognitive configurations [22]. Our results extend this molecular perspective to depth. CAP5 showed widespread model gains despite having no FDR-significant individual receptor association for its mean configuration A. Its depth slope L, however, was associated with 5-HT2A, 5-HT4, CB1, and MOR. Examining depth variation can reveal associations not apparent in the average map.

The opposite MOR associations of CAP1 and CAP2 provide an opioid-related spatial context for their contrast between emotion regulation and sensory attention. Human PET experiments have linked endogenous opioid responses to emotional recovery following social rejection [44]. CAP3 and CAP7 shared positive 5-HT2A and negative norepinephrine-transporter associations, whereas CAP6 combined positive NET correspondence with negative associations involving 5-HT2A. These profiles distinguish the molecular environments of internally oriented memory/social configurations from those of externally oriented sensorimotor configurations. Pharmacological fMRI studies show that noradrenaline-reuptake manipulation changes network topology according to cognitive context [45].

In the local PLS results, significant neighborhoods for CAP4-A were concentrated in dorsal attention, somatomotor, and visual territories. CAP6-A included posterior parietal regions and parts of the precuneus/posterior cingulate cortex. Posterior parietal regions coordinate spatial orienting and detect behaviorally relevant stimuli [46], while posteromedial default mode regions support internally generated representations [37, 38]. Molecular correspondence at these locations suggests that the distinction between sensory selection and internal representation is aligned with regional neurochemical gradients.

### Altered GM-WM coordination and receptor-associated CAP differences in MDD

Against this shared functional and molecular organization, MDD showed reduced coordination between GM and WM state sequences (Figure 6A–I). Chance matching was higher in MDD, indicating that the marginal state distributions differed between groups. However, observed concordance was lower, and the label-invariant NMI and ARI were also reduced, indicating that the joint organization of the sequences changed. These effects were small, as in large multisite MDD studies [25]. Earlier CAP studies linked altered default mode and frontoparietal dynamics to rumination [19], and recent work related imbalanced transitions between visual-attentional and limbic-default configurations to depression severity [20]. Our findings add a cross-tissue dimension to this account.

Depth-resolved analysis revealed distinct aspects of this difference. Increased mismatch and reduced concordance were evident at 1–5 mm, while NMI and ARI remained lower across all seven depths. Boundary-adjacent WM samples show the strongest GM-WM spatial correspondence (**Supplementary Figure S9**), so effects confined to shallow depths could partly reflect group differences in how much cortical signal is mixed into WM samples. The persistence of NMI and ARI differences at 7 mm is less readily explained in this way.

This overall reduction was accompanied by selective redistribution. Same-state co-occurrence decreased for CAP1 and CAP2 but increased for CAP3 and CAP4 (Figure 5). The concurrent reductions for CAP1 and CAP2 indicate that GM and WM less often occupied the same pole of this emotion-regulation/sensory-attention axis. This redistribution fits a view of depression as a reorganization of relationships among functional systems. Precision mapping has likewise identified altered allocation of cortical territory among salience and neighboring networks in depression [47].

Lagged coupling added temporal specificity to this redistribution. Reduced WM CAP3→GM CAP2 and GM CAP5→WM CAP1 probabilities persisted across 1–7 mm (Supplementary Figure S19). The CAP5→CAP1 asymmetry arose because coupling decreased more strongly in the GM-leading direction. Because WM hemodynamic responses are delayed and attenuated relative to GM [9], these asymmetries describe the ordering of sampled BOLD states rather than the direction of neural influence. This temporal ordering complements evidence for asymmetric state trajectories in depression [20].

The spatial CAP differences associated with MDD supplied a complementary molecular perspective (Figure 6J–K). In GM, CAP1 and CAP2 effect maps had opposite associations with 5-HT2A and MOR. In WM, the opposing 5-HT4 and serotonin-transporter associations of CAP6 and CAP7 connected disease-related spatial changes to another complementary state pair. Because absolute effect maps showed no significant depth-averaged associations, this correspondence with normative receptor distributions concerns the signed pattern of change.

PET evidence links altered MOR responses in MDD to impaired recovery from social rejection [44], and meta-analysis supports altered cortical 5-HT2A binding in depression [48]. Psilocybin treatment studies associate increased network integration with antidepressant improvement [49]. For 5-HT4, lower binding in untreated MDD is associated with poorer verbal memory, while longitudinal treatment data link binding changes to memory outcomes [50, 51]. The serotonin transporter is a target of commonly used antidepressants [52]. Additional GM associations with mGluR5 and VAChT broaden this molecular context to glutamatergic and cholinergic systems. The mGluR5 finding is consistent with human PET and postmortem evidence for altered glutamatergic receptor expression in depression [53]. This framework motivates testing whether treatment-related changes in cross-tissue coordination track improvements in the affective and cognitive processes these states represent.

### Limitations

Template-based sampling leaves shallow WM measurements susceptible to registration error, partial-volume mixing and group differences in cortical thickness. Persistence at 7 mm reduces this concern. Weaker WM BOLD signals, physiological noise and tissue differences in hemodynamic timing complicate interpretation of depth and lag [9]. CAP solutions also depend on model choice. The elbow criterion selected seven states; two pairs were near-inverse, and CAP3 and CAP7 overlapped. Decoding uses independent task-based maps and cannot identify participants’ ongoing mental processes. The receptor atlas measures normative cortical distributions, precluding inference about WM receptor expression or molecular changes in individual patients. Multiple testing was controlled within analytic families, and several molecular associations were close to threshold. The small group effects require replication. Medication, illness stage and comorbidity were not modeled, and the cross-sectional design cannot establish when altered coordination emerges. Higher-resolution and longitudinal studies with individual molecular and behavioral measurements could clarify these relationships.

## Conclusion

Shared GM-WM states showed distinct cognitive and cortical receptor associations. In MDD, altered co-occurrence and temporal ordering accompanied lower cross-tissue coordination. Exact mismatch increased at shallower depths, while reduced dependence between tissue sequences persisted through 7 mm. Serotonergic and opioid distributions also tracked the direction of regional shifts in state expression.

## Supporting information

bCAP_19_whz_20261127_BB_BKB2_MM.docx

## Author contributions

**H.W.** led the study conception and design, developed the analysis code, performed the statistical analyses, interpreted the results, prepared the figures, and wrote the first draft of the manuscript. **J.X.** contributed to the initial study concept and provided the data. **M.M., Y.S., Y.X., C.Y., E.A., D.S., C.L**., and **D.Y.** reviewed and commented on the manuscript. **B.B.B.** provided scientific guidance and critically revised successive drafts. **B.K.-B.** helped shape the initial research question, supervised the study throughout, secured funding, and critically revised the manuscript at each stage.

## Data availability

DIRECT Phase II imaging data are available from Science Data Bank (https://doi.org/10.57760/sciencedb.psych.00689). Access to the archive password and phenotypic data requires a signed data use agreement (https://rfmri.org/DIRECTPhase2). Normative receptor and transporter maps are available at https://github.com/netneurolab/hansen_receptors. The parcellated Neurosynth term maps used for functional decoding are available at https://github.com/netneurolab/hansen_brainstemfc.

## Code availability

The corresponding authors provide scripts on request.

## Acknowledgments

We thank the DIRECT Consortium, contributing sites and participants. The China MOST2030 Brain Project supported B.K.-B. (grant 2022ZD0208500).

## Supplementary information

Supplementary information is available at MP’s website. The Word supplement includes detailed methods, equations, tables and depth-specific results.

## Consent for publication

Not applicable. No identifiable individual participant information is presented in this manuscript.

