## Supplementary material for "Gray-white matter coactivation patterns, cortical receptor correspondence and cross-tissue coordination in major depression": bCAP_19_whz_20261127_BB_BKB2_MM.docx

**Supplementary Methods**

**Participants**

This study used resting-state fMRI (rs-fMRI) and T1-weighted structural MRI data from Phase II of the DIRECT dataset [1, 2]. Contributing sites diagnosed MDD according to criteria in either the International Classification of Diseases, 10th Revision (ICD-10), or the Diagnostic and Statistical Manual of Mental Disorders, Fourth Edition (DSM-IV) [3, 4]. Clinical assessments available for the present analyses included the Hamilton Depression Rating Scale (HAMD) and the Hamilton Anxiety Rating Scale (HAMA), which assess the severity of depressive and anxiety symptoms, respectively [5, 6]. All participants provided written informed consent. Each contributing cohort study received approval from its local Institutional Review Board. All data were de-identified and anonymized.

Participants required a recorded MDD or NC diagnosis, resting-state fMRI and valid extracted time series. Of 3,001 participants with diagnostic information and imaging data, 442 failed the motion or time-series length criteria given below, and eight lacked usable extraction outputs. The CAP sample included 2,551 participants from 22 sites (1,417 MDD, 1,134 NCs). Missing clinical scores did not preclude inclusion. Fully adjusted analyses required complete age, sex, education, and mean FD (1,404 MDD, 1,130 NCs), with sample sizes further determined by feature availability (Figure S1 and Tables S1, S2 and S3).

**Image preprocessing**

rs-fMRI and structural T1 data were preprocessed at each participating site using the harmonized Data Processing & Analysis for Brain Imaging on Surface (DPABISurf) protocol described in the original DIRECT Phase II report [1, 7]. DPABISurf incorporates fMRIPrep for anatomical and functional image preprocessing [8]. The present analysis used the released volumetric BOLD images in the MNI152NLin2009cAsym standard space. No additional initial volumes were discarded, and no spatial smoothing was applied to the BOLD images in our analysis, in order to limit further signal mixing across the gray-white matter boundary.

After extracting paired GM and WM time series, we performed temporal denoising using Nilearn [9]. Because the released data did not include complete fMRIPrep confound files, nuisance regressors were reconstructed from the available realignment parameters and image data. The nuisance model included 24 motion regressors (the six rigid-body motion parameters, their first temporal derivatives, and the squared terms of both sets), together with a mean cerebrospinal fluid (CSF) signal and one indicator regressor for each outlier volume. The CSF signal was sampled from high-probability CSF locations (probability ≥ 0.90) defined using the MNI template tissue-probability maps. The extracted signals underwent linear detrending, Butterworth band-pass filtering (0.01-0.08 Hz), and nuisance regression, with filtering based on each participant’s repetition time. Temporal filtering was also applied to the nuisance regressors within the same cleaning procedure. Mean WM and global-signal regressors were deliberately omitted because WM signals were the signal of interest, and removing them would alter the GM-WM covariance structure under study.

Outlier volumes were identified using the supplied Power framewise displacement (FD) values and standardized DVARS recomputed from a fixed sample of template-defined brain locations. Volumes with FD > 0.5 mm or standardized DVARS > 1.5 were modeled with individual indicator regressors and subsequently removed from the denoised time series. Participants were excluded if their mean FD before censoring exceeded 0.20 mm, more than 20% of volumes were censored, or fewer than 150 valid volumes remained. Finally, each retained GM and WM parcel time series was standardized to zero mean and unit sample variance within each participant before CAP analysis.

**Template-based GM-WM boundary construction and sampling at multiple depths**

A common sampling geometry was constructed from the GM and WM tissue probability maps of the MNI152NLin2009cAsym template obtained through TemplateFlow [10]. Tissue probability maps were aligned to the 2-mm template grid containing the 400-parcel, seven-network Schaefer atlas [11], using linear interpolation where resampling was required. For boundary estimation, the GM and WM probability maps were separately smoothed with a Gaussian kernel of 2 mm full width at half maximum. A signed tissue contrast field was then defined as:

$S\left( x \right)=\tilde{P}_{GM}\left( x \right)-\tilde{P}_{WM}\left( x \right)$	(1)

where the tilde denotes the smoothed probability maps. The GM–WM boundary was defined by the zero level set of S, with positive and negative values indicating the GM-dominant and WM-dominant sides, respectively.

Boundary points were identified by detecting sign changes between adjacent voxel centers along the three image axes. Candidate edges were retained when the GM-side voxel had a valid Schaefer parcel label, and the summed GM and WM probabilities were at least 0.50 at both endpoints. For an edge connecting $v_{0}$ and $v_{1}$, the subvoxel boundary location was estimated by linear interpolation:

$t=\frac{S(v_{0})}{S(v_{0})-S(v_{1})}$	(2)

$x_{b}=v_{0}+t(v_{1}-v_{0})$	(3)

Duplicate boundary detections within each parcel were removed using coordinates rounded to a 0.25 mm grid, while the original subvoxel coordinates were retained for sampling. Each boundary point inherited the parcel label of the GM-side voxel.

At each boundary point, a unit normal directed from WM toward GM was calculated from the spatial gradient of $S$ in physical coordinates. Paired sampling locations were positioned 1 mm toward GM and at seven distances toward WM along the same normal:

$n\left( x_{b} \right)=\frac{\nabla S(x_{b})}{\left\| \nabla S(x_{b}) \right\|}$	(4)

$x_{\mathrm{GM}}=x_{b}+(1 mm)\cdot n\left( x_{b} \right)$	(5)

$x_{WM,d}=x_{b}-d\cdot n\left( x_{b} \right)$	(6)

where $d=1, 2,\ldots, 7$ mm. Sampling validity was assessed using tissue probability maps without the additional smoothing used for boundary estimation. A pair was retained only when both locations were within the image bounds, the GM probability at the GM location was at least 0.20, and the WM probability at the WM location was at least 0.50. At least three valid pairs were required for each parcel at each depth. Pair selection was performed independently at each WM depth. Consequently, although the GM sampling offset was fixed at 1 mm, the GM sampling subset could differ across depths according to the validity of its paired WM locations. Supplementary Table S4 reports the number of effective sampling points at each depth.

The resulting template sampling geometry was applied consistently across participants. BOLD signals were extracted from each participant’s MNI-space functional images by trilinear interpolation at the retained coordinates and averaged with equal weights across sampling points within each parcel, separately for GM and WM. Both members of each pair shared the same cortical parcel label, thereby defining corresponding GM and underlying WM signals. Temporal denoising, censoring, and within-participant z-standardization were subsequently applied as described above. This procedure yielded paired GM and WM time-series matrices, each containing 400 parcels, at every WM sampling depth for the CAP analyses.

**Coactivation pattern (CAP) analysis**

The paired GM and WM parcel time series obtained at each sampling depth were used to identify recurring spatial patterns of BOLD activity through CAP analysis [12], implemented using NeuroCAPs (NeuroCAPs: A Python Package for Performing Co-Activation Patterns Analyses on Resting-State and Task-Based fMRI Data) and custom Python scripts. Two complementary representations were analyzed at each depth. Separate CAP models were fitted independently to the 400-dimensional GM and WM signals, allowing each tissue to have its own state sequence. A joint CAP model was fitted to the concatenated 800-dimensional GM-WM signals to characterize their shared spatial configurations. Participants with MDD and NCs were pooled during model fitting to establish common state definitions for subsequent group comparisons.

To equalize participants’ contributions to model fitting, 50 retained frames were randomly sampled without replacement from each participant, yielding 127,550 frames per model. Corresponding GM and WM frames were sampled using the same indices. Candidate cluster numbers from 2 to 20 were evaluated for the joint representation at each depth using a further participant-balanced subset of 48,469 frames (19 per participant). Clustering used k-means with Euclidean distance. We evaluated cluster solutions using the within-cluster sum-of-squares elbow criterion, which indicated seven clusters at every depth (Supplementary Figure S2). We retained the seven-cluster solution at each depth and used the same number of clusters for the corresponding separate GM and WM models. Candidate solutions used 10 initializations, and final models used 20 initializations, with a maximum of 500 iterations. Each cluster centroid represented the characteristic spatial pattern of a CAP. Following model fitting, all retained frames from each participant were assigned to the nearest centroid to reconstruct individual CAP sequences.

CAP labels were aligned across tissues and depths using Pearson spatial correlations between centroids and the Hungarian assignment algorithm, which maximized the total correlation under a one-to-one matching constraint. The joint CAP model at a WM depth of 1 mm served as the common reference. Joint models at the remaining depths were matched directly to this reference. For the separate models, GM and WM centroids were first paired according to their correlations across the 400 corresponding parcels. Each matched GM-WM pair was then concatenated into an 800-dimensional vector and matched to the reference joint centroids. Both members of each pair received the same reference CAP label, establishing a common reference-based labeling scheme across representations and depths.

Individual CAP dynamics were characterized by temporal fraction, persistence, occurrence counts, and transition measures. Temporal fraction was defined as the proportion of retained frames assigned to each CAP. In addition to the standard NeuroCAPs outputs, temporal measures accounting for censoring gaps were calculated using the original acquisition indices. In these calculations, censored intervals interrupted state episodes, and transitions were counted only between consecutive retained frames that were also consecutive in the original acquisition. Mean dwell time was calculated as the average duration of uninterrupted state episodes, converted to seconds using each participant’s repetition time. State-entry rates were expressed as the number of episodes per minute of retained data. Transition probabilities were obtained by normalizing each state’s outgoing transition counts, including self-transitions.

Participant-specific CAP maps were generated by averaging the standardized parcel signals over all retained frames assigned to each state. Joint-model maps used a shared state sequence for GM and WM, whereas separate-model maps used the respective tissue-specific sequences. When a CAP was not observed in a participant, its spatial activation values were treated as missing. These maps and the aligned state sequences provided the inputs for subsequent analyses of GM-WM correspondence and its variation with sampling depth. Main Figure 1 depicts the overall computational workflow.

**ComBat harmonization of CAP-derived features**

Following CAP estimation and state alignment, site-related variation in participant-level CAP features was corrected using ComBat, implemented in the Python package neuroCombat (version 0.2.12) [13, 14]. The acquisition site was specified as the batch variable. Diagnosis and sex were included as categorical covariates, while age, education, and mean framewise displacement were included as continuous covariates to preserve their modeled associations with the imaging features. Harmonization adjusted for both additive and multiplicative site effects.

For CAP summary measures, features were aggregated at the participant level before harmonization. Features were grouped by measurement source and further partitioned according to their complete-case patterns, allowing features with the same available participants to be harmonized together without imputing missing observations. Each fitting block required at least 20 participants overall and at least five participants per included site. Parametric empirical Bayes estimation was applied to blocks containing at least 10 nonconstant features.

Participant-specific parcel-level CAP maps were harmonized separately for each combination of representation, metric, CAP state, and sampling depth, with the 400 parcels constituting the features and participants constituting the observations. Within each representation, metric, and CAP state, participants were required to have complete parcel-level data across all seven depths and complete covariate information. The same participant subset was therefore retained across depths, preserving the paired structure of subsequent depth comparisons. Analysis-specific sample sizes are reported in Supplementary Table S2. ComBat parameters were estimated separately at each depth, with empirical Bayes information pooled across the 400 parcels. The resulting harmonized features were used in subsequent group-level and molecular analyses.

**Acquisition and processing of neurotransmitter receptor and transporter maps**

Normative neurotransmitter receptor and transporter maps were obtained from the publicly available atlas compiled by Hansen et al. [15], which aggregates molecular imaging data from more than 1,200 healthy individuals. The current study used the released Schaefer-400 parcel-wise matrix [11], comprising 19 molecular targets across nine neurotransmitter systems: serotonin (5-HT1A, 5-HT1B, 5-HT2A, 5-HT4, 5-HT6, and 5-HTT), dopamine (D1, D2, and DAT), acetylcholine (α4β2, M1, and VAChT), norepinephrine (NET), glutamate (NMDA and mGluR5), γ-aminobutyric acid (GABA_A), histamine (H3), cannabinoid (CB1), and opioid (MOR). The source maps were primarily derived from positron emission tomography (PET), with the DAT map derived from single-photon emission computed tomography (SPECT).

The pre-calculated receptor matrix was downloaded from a fixed version of the authors’ repository and checked for file integrity, completeness, and parcel ordering (<https://github.com/netneurolab/hansen_receptors>). All 400 cortical parcels, comprising 200 parcels per hemisphere, were retained and aligned with the corresponding parcel identifiers used in the CAP analyses, yielding a 400 × 19 molecular feature matrix. For partial least squares analyses, each receptor or transporter map was subsequently centered and scaled to unit variance across parcels during model fitting. In cross-validated multivariate models, scaling parameters were estimated within the training folds and applied to the corresponding held-out parcels. The cortical distributions of the 19 receptor and transporter maps are shown in Supplementary Figure S3.

**Receptor contributions to CAP depth profile similarity beyond spatial distance**

We assessed whether regional receptor similarity explained CAP depth profile similarity beyond interregional spatial distance. This analysis used covariate-adjusted maps from NC participants, based on the joint GM-WM CAP states defined in the pooled MDD and NC sample. For each participant i, CAP c, and depth d, the individual activation maps were z-standardized across the 400 parcels separately for GM and WM:

$Z_{i,c,d,p}^{\tau}=\frac{A_{i,c,d,p}^{\tau}-\overline{A_{i,c,d}^{\tau}}}{s_{i,c,d}^{\tau}}, \tau\in\{GM, WM\}$	(7)

Here, A denotes the parcel-wise mean activation of frames assigned to the CAP; the overbar and s denote the mean and sample standard deviation across parcels, respectively. The shared GM-WM activation map was defined as:

$Y_{i,c,d,p}=\frac{Z_{i,c,d,p}^{\mathrm{GM}}+Z_{i,c,d,p}^{\mathrm{WM}}}{2}$	(8)

Following ComBat harmonization, NC marginal mean maps adjusted for age, sex, education, mean framewise displacement, and site were estimated for each CAP and depth. Denoting the adjusted NC mean for parcel p at depth d by $\mu_{c,p,d}$, its seven-depth profile was:

$C_{c,p}=(\mu_{c,p,1}, \mu_{c,p,2}, \ldots, \mu_{c,p,7})$	(9)

For each CAP, interparcel depth profile similarity was calculated as the Pearson correlation across the seven depths (1–7 mm):

$T_{p,q}^{c}=corr(C_{c,p}, C_{c,q})$	(10)

Using the 19 receptor and transporter maps described above, each molecular map was z-standardized across the 400 parcels. Molecular similarity was then defined as the Pearson correlation between the 19-dimensional receptor fingerprints $z_{p}$ and $z_{q}$:

$S_{p,q}=corr(z_{p}, z_{q})$	(11)

For each CAP c and seed parcel p, two ordinary least squares models were fitted across target parcels q. The baseline model included the Euclidean distance $D_{p,q}$ between parcel coordinates in MNI space, and the full model additionally included receptor similarity:

$T_{p,q}^{c}=\alpha_{0,c,p}+\alpha_{1,c,p}D_{p,q}+\varepsilon_{c,p,q}$	(12)

$T_{p,q}^{c}=\beta_{0,c,p}+\beta_{1,c,p}D_{p,q}+\beta_{2,c,p}S_{p,q}+\eta_{c,p,q}$	(13)

Each model used all 400 target parcels, including the seed parcel itself. Model fit was quantified by adjusted R²:

$R_{\mathrm{adj}}^{2}=1-\frac{(1-R^{2})(n-1)}{n-K-1}$	(14)

Here, n = 400 and K is the number of predictors excluding the intercept (K = 1 for the distance-only model and K = 2 for the full model). Receptor model gain for each CAP and seed parcel was defined as:

$\Delta_{c,p}=R_{adj,full,c,p}^{2}-R_{adj,distance,c,p}^{2}$	(15)

Positive gain indicated additional in-sample variance explained by receptor similarity. When summarized across the cortex, gain was averaged over the 400 seed parcels within each CAP.

Statistical significance was assessed using 10,000 hemisphere-preserving spatial spin permutations with bijective parcel matching. For each seed, only the target locations in its receptor similarity vector were permuted, replacing $S_{p,q}$ with $S_{p,P_{b}(q)}$, where $P_{b}$ denotes the target permutation; the seed, CAP similarity, and distance vector remained fixed. The full model and its gain were recomputed for each permutation. One-sided empirical P values were calculated as:

$P_{c,p}=\frac{1+\sum_{b=1}^{B} \mathbb{1[}\Delta_{c,p}^{b}\geq\Delta_{c,p}^{\mathrm{obs}}]}{B+1}, B=10,000$	(16)

Here, $\Delta^{\mathrm{obs}}$ and $\Delta^{b}$ denote the observed and permuted gains, respectively, and 𝟙 is the indicator function. Benjamini–Hochberg false discovery rate (FDR) correction was applied across the 2,800 tests (seven CAPs × 400 seed parcels) in the joint shared-activation analysis. Significant receptor contributions required FDR-adjusted P < 0.05 and positive model gain.

Mean spatial configuration and depth phenotypes

The shared GM-WM activation maps were summarized by two complementary parcel-wise phenotypes. Let $\tilde{Y}_{i,c,d,p}$ denote the ComBat-adjusted shared activation of participant i, CAP c, depth d, and parcel p. The mean configuration A was defined as the arithmetic mean across the seven WM depths:

$A_{i,c,p}=\frac{1}{7}\sum_{d=1}^{7} \tilde{Y}_{i,c,d,p}$	(17)

The depth phenotype L was the ordinary least squares slope of shared activation against the actual WM sampling distance in millimeters:

$L_{i,c,p}=\frac{\sum_{d=1}^{7} (d-4)(\tilde{Y}_{i,c,d,p}-A_{i,c,p})}{\sum_{d=1}^{7} (d-4)^{2}}$	(18)

Thus, A described the average spatial configuration, whereas L quantified its linear redistribution with depth; positive and negative slopes indicated increasing and decreasing relative shared activation, respectively. Both phenotypes required complete seven-depth profiles. Because GM and WM maps had been standardized across parcels, these phenotypes described relative spatial expression rather than absolute BOLD amplitude.

For each CAP and phenotype, NC reference maps were estimated from parcel-wise ordinary least squares models within the NC group, adjusting for age, sex, education, mean FD, and site. Marginal means were evaluated at a common NC covariate distribution, defined across unique NC participants, to make the maps comparable across CAPs. The main molecular analyses used the joint shared-activation representation. The same summaries were also calculated for signed tissue differences (standardized WM minus GM activation) and for separate GM and WM activation (Supplementary Figures S14B–D and S15B–D). Signed tissue differences describe spatial activation differences and are distinct from temporal state mismatch. Network means and the locations of prominent parcels were summarized using the Schaefer seven-network annotations for descriptive anatomical interpretation.

**Meta-analytic functional decoding of CAP spatial maps**

We correlated signed adjusted NC maps for mean shared activation (A) with 123 Neurosynth association-test maps using Spearman correlation across 400 parcels [16]. The terms were selected from the Cognitive Atlas [17].

The reference matrix was atl-schaefer2018_res-400_neurosynth.csv from the public Hansen et al. brainstem functional-connectivity repository (https://github.com/netneurolab/hansen_brainstemfc; fixed commit 522a60f2736f09b5c29cab9b9ffd174e4006e6af; 10.1038/s41593-024-01787-0). It contains 123 Neurosynth association-test maps for Cognitive Atlas terms, already parcellated into the 400 Schaefer regions. We checked parcel order and retained all 400 parcels, comprising 200 in each hemisphere. The target maps were the same adjusted NC joint shared-activation phenotype A maps shown in Figure 2 and used for the mean-configuration receptor analysis.

The released spins_schaefer400_hungarian.npy ensemble supplied 10,000 bijective, hemisphere-preserving permutations [18]. Each permutation reassigned a CAP map across parcel locations before correlation with each reference map. For each CAP–term pair, the two-sided P value was [1 + number of permutations for which |ρperm − mean(ρperm)| ≥ |ρobserved − mean(ρperm)|]/10,001. Thus, this decoding analysis centered each comparison on its own spatial-null mean; it did not use ordinary correlation P values. All 861 primary tests entered one BH-FDR family before any term selection. The seven radar coordinates retained their original coefficients and q values; radial position was ρ + 0.8, with zero at radius 0.8. No CAP-specific rescaling was applied.

The compact fingerprint heatmap displays the union of the three strongest positive and three strongest negative correlations for each CAP (20 distinct terms). The exhaustive heatmap displays all 123 terms in alphabetical order. CAP spatial similarity was the Pearson correlation of the 400-element target maps; functional fingerprint similarity was the Pearson correlation of the complete, unthresholded 123-element coefficient vectors. These two similarity matrices are descriptive and were not assigned significance tests. The seven representative radar terms are descriptive coordinates, not orthogonal or independently derived functional axes. The reference maps originate from task-based literature; their spatial correspondence with resting-state CAPs does not identify participants’ mental states or establish task activation in WM.

After inference, we displayed emotion regulation, autobiographical memory, social cognition, response selection, coordination, multisensory processing and visual attention. Tables S5–S6 report the summary and complete results.

**Participant subsampling of joint CAPs and meta-analytic functional decoding**

From the 2,551 participants who passed CAP quality control, we drew one random sample without replacement at each of three proportions (10%, 30% and 50%). We stratified by site and diagnosis and used the same participants at all seven WM depths within each proportion. At each depth, we refitted joint k-means models to the 800 GM and WM parcel features. We sampled up to 50 retained frames per participant and assessed K = 2–20 on a participant-balanced set capped at 50,000 frames (50, 50 and 39 frames per participant, respectively). We applied the original elbow criterion to the within-cluster sums of squares using 10 initializations per candidate K. We also fitted seven-state models with 20 initializations on all 50 sampled frames per participant.

We aligned the seven subsample centroids at 1 mm to the full-cohort centroids at 1 mm using Hungarian matching on signed spatial Pearson correlations. Centroids at the other depths were aligned to the 1-mm centroids from the same subsample. We then compared corresponding subsample and full-cohort centroids at each depth. Within each subsample, we assigned all retained frames to the nearest centroid, reconstructed individual shared-activation maps and repeated ComBat harmonization and covariate adjustment to estimate the NC mean phenotype A. For each CAP, we correlated this map with the same 123 Neurosynth term maps and compared the resulting signed correlation profile with its full-cohort counterpart using Spearman correlation. We also compared the signs of the seven representative associations shown in Figure 2. Each subsample was drawn from the full cohort used as the reference (Figure S12 and Table S7).

**Spatial associations with individual receptor and transporter maps**

For each CAP, the adjusted NC A and L maps were correlated with each of the 19 receptor and transporter maps across the 400 cortical parcels using Spearman rank correlation. Statistical inference used 10,000 hemisphere-preserving, bijective spatial spin permutations of parcel locations [18]. The same spin ensemble was used for all molecular analyses. Two-sided empirical P values were obtained from the proportion of absolute permuted correlations at least as large as the absolute observed correlation, with one added to both the exceedance count and the number of permutations. Benjamini–Hochberg FDR correction was applied across 133 tests (seven CAPs × 19 molecular targets), separately for A and L and for each representation, activation metric, effect type, and receptor-data version.

**Multireceptor partial least squares analysis**

Partial least squares (PLS) regression was used to identify combinations of receptor and transporter maps that covaried with the NC CAP topography [19]. Each model treated the 400 parcels as observations, the 19 molecular maps as predictors, and one adjusted NC CAP map as the response. Models were fitted separately for each CAP to A, L, and the individual depth-specific maps. Predictors and responses were centered and scaled to unit sample variance across parcels. Up to 19 components of a single-response PLS (PLS1) model were extracted sequentially, with predictor and response residuals deflated after each component. Component signs were oriented so that the parcel score correlated positively with the target map. Receptor contributions were represented by the rotation coefficients that map the original standardized predictors to each component score. The incremental and cumulative response variance explained, and the incremental predictor variance explained, were recorded.

Whole-cortex component significance was assessed using the component-specific increment in response variance explained across all 400 cortical parcels. Here, “whole-cortex” refers to the spatial extent of the component-level test, distinguishing it from the local neighborhood tests described below. For each target, the entire PLS model was refitted after rotating the target map in each of 10,000 spatial permutations, and a one-sided permutation P value was obtained for each component. Benjamini–Hochberg FDR correction was applied across the 19 components separately within each CAP–phenotype–depth target, without additional correction across targets. Components meeting this within-target criterion (q < 0.05) were displayed; explained variance, spatial cross-validation, and bootstrap stability were reported as complementary model diagnostics.

Spatial localization was assessed using neighborhood Spearman correlations between the observed target map and each component score. Each neighborhood comprised the focal parcel and its 24 nearest parcels in the same hemisphere, based on Euclidean distances between parcel coordinates. Local two-sided P values were derived from the corresponding correlations obtained after refitting the PLS model for every rotated target. FDR correction was applied across the 400 neighborhood centers separately for each component. Significant centers were mapped and summarized by their anatomical and network assignments.

Model stability was evaluated using 1,000 spatial-block bootstrap samples. Ten spatial blocks were constructed by k-means clustering of parcel coordinates after standardizing the coordinate axes; blocks were resampled with replacement. Each bootstrap PLS solution was matched to the original solution by maximum absolute cosine similarity of the rotation vectors under one-to-one Hungarian assignment, followed by sign alignment. Receptor bootstrap ratios were calculated as the original rotation coefficient divided by its bootstrap standard deviation. Two-sided normal-approximation P values were FDR-corrected across the 19 receptor weights within each component and used as stability summaries. Spatial generalization was evaluated by five-fold cross-validation in which groups of these spatial blocks were held out, with at least 100 training parcels per fold. Scaling and model fitting were repeated using training parcels only. Cross-validated R² was calculated from the pooled held-out parcel predictions. These analyses evaluated the spatial prediction of NC reference maps.

**Spatial correspondence of GM and WM CAPs**

To characterize the spatial correspondence underlying the separate state sequences, Pearson correlations were calculated between the matched GM and WM CAP centroids across the 400 corresponding parcels at each WM depth. The mean and minimum correlations across the seven matched states were summarized as functions of depth. A correlation of 0.30 served as a matching-quality reference for flagging weak correspondence, rather than as an additional participant exclusion criterion. All temporal coordination analyses below used the aligned separate GM and WM state sequences, allowing the tissues to occupy different CAPs at the same time.

**Overall cross-tissue state agreement**

For a participant and depth, let $G_{t}$ and $W_{t}$ denote the aligned GM and WM state labels at a common retained frame, and let T be the number of common valid frames. The observed matching rate $p_{o}$ and the chance matching rate $p_{e}$ were defined as:

$p_{o}=\frac{1}{T}\sum_{t=1}^{T} \delta_{G_{t},W_{t}}, p_{e}=\sum_{k=1}^{7} p_{G,k}p_{W,k}$	(19)

Here, $\delta_{G_{t},W_{t}}$ denotes the Kronecker delta, which equals 1 when the GM and WM state labels are identical and 0 otherwise. $p_{G,k}$ and $p_{W,k}$ are the marginal proportions of frames assigned to state k in GM and WM, respectively. Chance matching therefore assumed independent tissue labels while retaining their observed state occupancies. We calculated the raw mismatch rate (1 − $p_{o}$), excess matching above chance E, and Cohen’s κ:

$E=p_{o}-p_{e}, \kappa=\frac{p_{o}-p_{e}}{1-p_{e}}$	(20)

Excess matching served as the principal summary of coordination beyond that expected from marginal occupancy. Normalized mutual information (NMI), the adjusted Rand index (ARI), and chance matching were examined as secondary agreement measures. With $p_{kl}$ denoting the joint proportion of GM state k and WM state l, mutual information and NMI were defined as:

$I(G;W)=\sum_{k,l=1}^{7} p_{kl}\log\frac{p_{kl}}{p_{G,k}p_{W,l}}, NMI=\frac{2I(G;W)}{H_{G}+H_{W}}$	(21)

$H_{G}$ and $H_{W}$ are the Shannon entropies of the corresponding marginal state distributions, with zero-probability terms omitted. ARI quantified agreement between the GM and WM partitions of retained frames, adjusted for the expected agreement of random partitions with fixed cluster sizes. NMI and ARI were calculated using scikit-learn. Unlike same-label matching, both measures are invariant to permutations of state labels. Measures were computed within runs and, where multiple runs were available, aggregated using the number of valid paired frames as weights.

**State co-occurrence and conditional mismatch**

We characterized the contemporaneous joint distribution of GM and WM states with a 7 × 7 co-occurrence matrix. Its entry $C_{kl}$ = $n_{kl}$/T was the fraction of all common valid frames in which GM occupied state k and WM occupied state l. This joint proportion was distinguished from state-conditional mismatch. Let $n_{G,k}$ and $n_{W,k}$ denote the numbers of frames in which the respective tissue occupied state k. GM-anchored and WM-anchored conditional mismatch rates were:

$M_{GM,k}=1-\frac{n_{kk}}{n_{G,k}}, M_{WM,k}=1-\frac{n_{kk}}{n_{W,k}}$	(22)

The two measures have different denominators: the first describes WM mismatch conditional on a GM state, whereas the second describes GM mismatch conditional on a WM state. When an anchor state was not observed, the corresponding value was treated as missing. The primary analysis included only participant-state-depth observations with at least 20 anchor frames. Across runs, conditional measures were combined using anchor-frame counts as weights.

A sensitivity analysis included all valid observations with at least one anchor frame and used weighted least squares models. Each observation was weighted by its anchor-frame count, capped at the 95th percentile within the corresponding tissue-anchor, CAP, and comparison-model sample. Both primary and weighted models accounted for repeated depths using participant-clustered robust covariance estimates. Depth-specific contrasts were FDR-corrected jointly across the two tissue anchors, seven states, and seven depths (98 contrasts) separately for each analysis and covariate model; diagnosis-by-depth interaction tests were corrected across the 14 anchor-state models.

**Lagged cross-tissue state relationships**

Temporal ordering was assessed using the separate GM and WM state sequences at their original acquisition indices. For a positive lag ℓ, the WM-leading and GM-leading conditional probabilities were defined as:

$P_{WM\to GM}^{\mathcal{l}}(k,l)=Pr(G_{t\mathcal{+l}}=l | W_{t}=k)$	(23)

$P_{GM\to WM}^{\mathcal{l}}(k,l)=Pr(W_{t\mathcal{+l}}=l | G_{t}=k)$	(24)

Each probability was estimated by dividing the corresponding transition count by the number of valid lagged pairs with the specified source state. Original frame indices were preserved after censoring. A lagged pair was retained only if both tissues had valid labels at the endpoints and at every intervening frame, preventing artificially adjacent observations across censoring gaps. At least 30 valid paired observations were required for a run–depth–lag estimate; conditional entries with an unobserved source state were treated as missing. The main directional analysis focused on the 1-mm WM sampling depth and a lag of one frame. One frame represented one repetition time, so its duration in seconds depended on the acquisition protocol.

Directional asymmetry was calculated within each participant for the same ordered source-target pair:

$\Delta_{kl}^{\mathcal{l}}=P_{WM\to GM}^{\mathcal{l}}(k,l)-P_{GM\to WM}^{\mathcal{l}}(k,l)$	(25)

Thus, positive asymmetry indicated a greater conditional probability for the WM-leading relation than for the GM-leading relation with the same source and target labels. Each direction and its within-participant difference were analyzed separately. In the equivalent signed-lag convention comparing WM at t with GM at t + ℓ, positive lags denoted WM-leading relationships and negative lags denoted GM-leading relationships. Group contrasts were defined as MDD minus NC. FDR correction covered the 49 ordered state pairs separately for each depth, lag, direction or direction-difference measure, and covariate model. These measures describe temporal ordering in sampled BOLD state sequences and do not establish neuronal transmission delays or causal direction.

**Receptor correspondence of MDD-related spatial effects**

To relate disease-associated spatial effects to the independent molecular atlas, adjusted MDD-minus-NC effect maps were estimated for phenotype A of the separately estimated GM and WM CAP activation maps. A separate ordinary least squares model was fitted at each parcel, including diagnosis, age, sex, education, mean FD, and categorical site. The adjusted standardized effect was:

$g_{c,p}=J(\nu_{c,p})\frac{\hat{\beta}_{group,c,p}}{\sigma_{res,c,p}}$	(26)

Here, the numerator is the fitted diagnosis coefficient, $\sigma_{\mathrm{res}}$ is the model residual standard deviation, and J(ν) is the Hedges small-sample correction for the residual degrees of freedom. A positive g indicates greater activation in the corresponding tissue in MDD. Signed g maps were correlated with each molecular map using Spearman correlation across parcels, with significance assessed using 10,000 spatial-spin permutations with two-sided inference and BH-FDR correction across 133 CAP-by-receptor tests separately for GM and WM. Absolute effect maps were assessed separately as a sensitivity analysis of effect magnitude, with their own correction family.

**Statistical analysis**

Continuous demographic and clinical variables were summarized as mean ± sample standard deviation, with the number of available observations reported for each variable. Between-group demographic comparisons used two-sided Welch t tests for continuous variables and Pearson χ² tests for sex. Clinical scales and illness-history measures, which were unavailable for NCs, were summarized within the MDD group only. Missing demographic, clinical, or imaging values were not imputed; each model used participants with complete values for its outcome and covariates. Analyses were conducted in Python using NumPy, SciPy, pandas, scikit-learn, neuroCombat, NeuroCAPs, and custom scripts.

Participant-level co-occurrence, lagged probabilities, and other temporal CAP features were compared using ordinary least squares with HC3 heteroskedasticity-robust standard errors. The main model included diagnosis, age, sex, education, and categorical site. A site-adjusted model including diagnosis and site was examined as a sensitivity analysis. Mean FD was included as a protected covariate during ComBat harmonization of these features but was not included in the temporal outcome regressions. Molecular parcel-wise models additionally adjusted for mean FD, as specified in the main text. Effect estimates were reported as MDD minus NC, together with test statistics, 95% confidence intervals, and adjusted Hedges’ g obtained by dividing the model contrast by the residual standard deviation and applying a small-sample correction.

Repeated-depth analyses of overall agreement and state-conditional mismatch included categorical depth and its interaction with diagnosis. For an outcome $u_{id}$ measured at depth d in participant i, the main model was:

$u_{id}=\beta_{0}+\beta_{G}G_{i}+\sum_{h=2}^{7} \alpha_{h}D_{dh}+\sum_{h=2}^{7} \delta_{h}G_{i}D_{dh}+\gamma^{T}X_{i}+\varepsilon_{id}$	(27)

$G_{i}$ indicated MDD status, $D_{dh}$ indicated sampling depth h (1 mm as the reference), and $X_{i}$ included age, sex, education, and site. Covariance estimates were clustered by participant to account for repeated observations, with finite-sample correction. Diagnosis-by-depth interactions were tested jointly using a robust Wald F test. Adjusted group means and depth-specific contrasts were evaluated over the observed covariate distribution of unique participants. Primary conditional-mismatch models used ordinary least squares, and the anchor-frame sensitivity models used weighted least squares. A corresponding site-adjusted model was also fitted.

For each of the seven overall agreement measures, the depth curve was additionally summarized within participants using the trapezoidal area under the curve, normalized by the observed depth span:

${AUC}_{i}=\frac{\sum_{j=1}^{m-1} \frac{u_{i,d_{j}}+u_{i,d_{j+1}}}{2}(d_{j+1}-d_{j})}{d_{m}-d_{1}}$	(28)

Here, m is the number of valid depths, ordered from shallow to deep; at least five were required. For complete profiles, the normalization span was 6 mm. Individual curve shape was also characterized by a quadratic polynomial in the actual depth coordinate:

$u_{id}=a_{i}+b_{i}d+c_{i}d^{2}+e_{id}$	(29)

The coefficient $b_{i}$ was the linear term of this quadratic fit, and $c_{i}$ represented curvature. Group comparisons of individual depth-curve shape used $b_{i}$. This coefficient was distinct from the simple linear slope L of parcel-wise shared activation defined in Eq. 18 above. Normalized depth AUC and $b_{i}$ were compared using ordinary least squares with participant-clustered, small-sample-corrected covariance (equivalent to HC1 for these participant-level summaries), adjusted for age, sex, education and site.

All conventional group tests were two-sided. The significance threshold was FDR-adjusted P < 0.05. For overall agreement curves, depth-specific contrasts were corrected across the seven depths within each outcome, while omnibus interaction tests were corrected across the seven outcomes. Depth-AUC and curve-coefficient tests were corrected across the seven outcomes separately for each summary and covariate model. Co-occurrence matrices were corrected across the 49 state pairs within each depth, measure, and covariate model. The larger conditional-mismatch family, the lag-specific families, and the molecular testing families are defined in the preceding sections of these Supplementary Methods. Primary and sensitivity analyses were corrected separately. Permutation P values used a finite-sample correction in which one was added to both the exceedance count and the permutation denominator; molecular correlations were tested two-sided, whereas receptor model gain and PLS component increments were tested one-sided. Benjamini–Hochberg correction was used throughout.

In effect matrices, colors represented the adjusted group-comparison t-statistics, and significance symbols indicated the corresponding FDR level. Borders marking |g| > 0.20 were descriptive effect-size annotations and were not additional significance criteria. Spatial receptor correlations and PLS treated cortical parcels as the observations, whereas demographic and temporal group comparisons treated participants as the independent sampling units. Network coverage, regional rankings, and representative anatomical locations were descriptive summaries.

**Supplementary Tables**

Table S1. Site-specific demographic and clinical characteristics. Panel A. Sample size, sex and age in the full CAP cohort (N = 2551).

| **Site** | **MDD n** | **NC n** | **MDD male/female (female %)** | **NC male/female (female %)** | **Sex test** | **P sex** | **MDD age (years)** | **NC age (years)** | **Age test** | **P age** |
| --- | --- | --- | --- | --- | --- | --- | --- | --- | --- | --- |
| IS001 | 241 | 198 | 86/155 (64.3%) | 74/124 (62.6%) | χ² = 0.134 | 0.714 | 37.56 ± 13.27 [241] | 36.07 ± 15.12 [198] | t = 1.091 | 0.276 |
| IS002 | 138 | 61 | 35/99 (73.9%) | 22/39 (63.9%) | χ² = 2.005 | 0.157 | 18.94 ± 3.87 [130] | 20.72 ± 3.12 [61] | t = -3.403 | <0.001 |
| IS003 | 15 | 17 | 5/10 (66.7%) | 3/14 (82.4%) | OR = 2.333 | 0.423 | 33.93 ± 9.35 [15] | 23.24 ± 4.45 [17] | t = 4.044 | <0.001 |
| IS004 | 28 | 25 | 12/16 (57.1%) | 7/18 (72.0%) | χ² = 1.268 | 0.260 | 29.93 ± 9.31 [28] | 30.00 ± 9.98 [25] | t = -0.027 | 0.979 |
| IS005 | 122 | 53 | 56/66 (54.1%) | 31/22 (41.5%) | χ² = 2.342 | 0.126 | 33.79 ± 9.94 [122] | 37.04 ± 9.32 [53] | t = -2.077 | 0.040 |
| IS006 | 44 | 87 | 21/23 (52.3%) | 37/50 (57.5%) | χ² = 0.320 | 0.572 | 30.70 ± 9.61 [44] | 23.77 ± 5.32 [87] | t = 4.455 | <0.001 |
| IS007 | 18 | 27 | 4/14 (77.8%) | 7/20 (74.1%) | OR = 0.816 | 1.000 | 20.39 ± 6.98 [18] | 20.33 ± 3.62 [27] | t = 0.031 | 0.975 |
| IS009 | 44 | 24 | 18/26 (59.1%) | 10/14 (58.3%) | χ² = 0.004 | 0.952 | 44.59 ± 14.61 [44] | 36.71 ± 13.83 [24] | t = 2.201 | 0.032 |
| IS010 | 73 | 62 | 33/40 (54.8%) | 29/33 (53.2%) | χ² = 0.033 | 0.855 | 33.68 ± 11.67 [73] | 35.56 ± 12.55 [62] | t = -0.896 | 0.372 |
| IS011 | 107 | 62 | 51/56 (52.3%) | 25/37 (59.7%) | χ² = 0.855 | 0.355 | 37.88 ± 12.78 [107] | 35.92 ± 11.93 [62] | t = 1.002 | 0.318 |
| IS012 | 62 | 32 | 21/41 (66.1%) | 15/17 (53.1%) | χ² = 1.510 | 0.219 | 30.47 ± 7.20 [62] | 29.59 ± 5.00 [32] | t = 0.688 | 0.494 |
| IS013 | 25 | 25 | 11/14 (56.0%) | 9/16 (64.0%) | χ² = 0.333 | 0.564 | 38.80 ± 14.53 [25] | 33.96 ± 13.32 [25] | t = 1.228 | 0.226 |
| IS014 | 127 | 130 | 44/83 (65.4%) | 56/74 (56.9%) | χ² = 1.921 | 0.166 | 43.06 ± 11.34 [127] | 37.27 ± 11.80 [130] | t = 4.009 | <0.001 |
| IS015 | 33 | 33 | 6/27 (81.8%) | 22/11 (33.3%) | χ² = 15.880 | <0.001 | 22.21 ± 4.69 [33] | 21.33 ± 4.45 [33] | t = 0.781 | 0.438 |
| IS016 | 82 | 79 | 36/46 (56.1%) | 43/36 (45.6%) | χ² = 1.784 | 0.182 | 25.99 ± 7.67 [82] | 25.37 ± 7.08 [79] | t = 0.534 | 0.594 |
| IS017 | 30 | 42 | 12/18 (60.0%) | 15/27 (64.3%) | χ² = 0.137 | 0.711 | 33.77 ± 8.98 [30] | 34.45 ± 9.41 [42] | t = -0.313 | 0.755 |
| IS018 | 32 | 30 | 17/15 (46.9%) | 11/19 (63.3%) | χ² = 1.694 | 0.193 | 35.72 ± 12.70 [32] | 33.07 ± 11.69 [30] | t = 0.856 | 0.395 |
| IS019 | 30 | 29 | 4/26 (86.7%) | 15/14 (48.3%) | χ² = 9.954 | 0.002 | 26.67 ± 10.91 [30] | 30.59 ± 7.12 [29] | t = -1.640 | 0.107 |
| IS020 | 20 | 20 | 4/16 (80.0%) | 5/15 (75.0%) | OR = 0.750 | 1.000 | 27.80 ± 8.99 [20] | 31.40 ± 10.99 [20] | t = -1.134 | 0.264 |
| IS021 | 61 | 53 | 16/45 (73.8%) | 29/24 (45.3%) | χ² = 9.633 | 0.002 | 32.21 ± 11.23 [61] | 32.28 ± 10.84 [53] | t = -0.034 | 0.973 |
| IS022 | 32 | 21 | 7/25 (78.1%) | 7/14 (66.7%) | χ² = 0.856 | 0.355 | 36.06 ± 10.36 [32] | 36.29 ± 13.60 [21] | t = -0.064 | 0.949 |
| IS023 | 53 | 24 | 5/48 (90.6%) | 12/12 (50.0%) | χ² = 15.802 | <0.001 | 13.96 ± 1.53 [53] | 13.62 ± 2.24 [24] | t = 0.670 | 0.508 |

Age is mean ± SD with valid n in square brackets. Sex is male/female (female %), using observed sex. Sex was compared using Pearson χ² tests or, when expected counts were below 5, Fisher exact tests (OR shown). Age was tested using a two-sided Welch t-test; t is MDD minus NC. P values are unadjusted. IS008 is absent from the final CAP cohort.

Table S1. continued. Panel B. Education and head motion by site.

| **Site** | **MDD education (years)** | **NC education (years)** | **Education test** | **P** | **MDD mean FD (mm)** | **NC mean FD (mm)** | **FD test** | **P** |
| --- | --- | --- | --- | --- | --- | --- | --- | --- |
| IS001 | 12.12 ± 3.46 [239] | 13.53 ± 3.76 [198] | t = -4.044 | <0.001 | 0.121 ± 0.038 [241] | 0.121 ± 0.037 [198] | t = 0.143 | 0.887 |
| IS002 | 11.95 ± 2.55 [128] | 13.72 ± 1.80 [60] | t = -5.458 | <0.001 | 0.093 ± 0.039 [138] | 0.073 ± 0.027 [61] | t = 4.036 | <0.001 |
| IS003 | 14.80 ± 3.32 [15] | 15.41 ± 1.18 [17] | t = -0.677 | 0.507 | 0.133 ± 0.031 [15] | 0.113 ± 0.032 [17] | t = 1.771 | 0.087 |
| IS004 | 12.79 ± 4.23 [28] | 12.91 ± 4.03 [23] | t = -0.110 | 0.913 | 0.139 ± 0.028 [28] | 0.133 ± 0.034 [25] | t = 0.666 | 0.509 |
| IS005 | 12.96 ± 3.46 [122] | 14.34 ± 2.57 [53] | t = -2.926 | 0.004 | 0.078 ± 0.040 [122] | 0.093 ± 0.042 [53] | t = -2.213 | 0.029 |
| IS006 | 11.93 ± 3.01 [44] | 13.93 ± 2.32 [87] | t = -3.866 | <0.001 | 0.118 ± 0.039 [44] | 0.128 ± 0.034 [87] | t = -1.524 | 0.132 |
| IS007 | 10.83 ± 3.05 [18] | 12.33 ± 2.29 [27] | t = -1.778 | 0.086 | 0.073 ± 0.023 [18] | 0.089 ± 0.030 [27] | t = -2.006 | 0.051 |
| IS009 | 10.89 ± 4.65 [44] | 13.50 ± 4.09 [24] | t = -2.399 | 0.020 | 0.102 ± 0.037 [44] | 0.116 ± 0.043 [24] | t = -1.324 | 0.193 |
| IS010 | 13.11 ± 3.15 [72] | 14.69 ± 2.23 [61] | t = -3.366 | 0.001 | 0.116 ± 0.040 [73] | 0.124 ± 0.041 [62] | t = -1.128 | 0.262 |
| IS011 | 10.92 ± 3.47 [107] | 13.27 ± 3.46 [62] | t = -4.267 | <0.001 | 0.085 ± 0.039 [107] | 0.087 ± 0.044 [62] | t = -0.318 | 0.751 |
| IS012 | 13.73 ± 3.39 [62] | 14.59 ± 2.82 [32] | t = -1.319 | 0.191 | 0.035 ± 0.010 [62] | 0.037 ± 0.010 [32] | t = -1.138 | 0.259 |
| IS013 | 12.04 ± 4.11 [25] | 11.72 ± 4.13 [25] | t = 0.275 | 0.785 | 0.119 ± 0.044 [25] | 0.113 ± 0.028 [25] | t = 0.577 | 0.567 |
| IS014 | 9.09 ± 3.62 [127] | 13.36 ± 4.16 [130] | t = -8.796 | <0.001 | 0.101 ± 0.038 [127] | 0.102 ± 0.035 [130] | t = -0.240 | 0.811 |
| IS015 | 11.91 ± 2.40 [33] | 12.61 ± 1.46 [33] | t = -1.425 | 0.160 | 0.079 ± 0.021 [33] | 0.092 ± 0.033 [33] | t = -1.942 | 0.057 |
| IS016 | 13.77 ± 2.59 [82] | 15.63 ± 2.68 [79] | t = -4.488 | <0.001 | 0.083 ± 0.030 [82] | 0.082 ± 0.027 [79] | t = 0.200 | 0.842 |
| IS017 | 12.80 ± 3.75 [30] | 14.31 ± 4.17 [42] | t = -1.606 | 0.113 | 0.131 ± 0.034 [30] | 0.136 ± 0.030 [42] | t = -0.557 | 0.580 |
| IS018 | 12.66 ± 4.74 [32] | 12.67 ± 3.06 [30] | t = -0.010 | 0.992 | 0.092 ± 0.033 [32] | 0.102 ± 0.044 [30] | t = -0.994 | 0.325 |
| IS019 | 12.83 ± 3.01 [30] | 13.97 ± 2.78 [29] | t = -1.502 | 0.139 | 0.104 ± 0.032 [30] | 0.111 ± 0.031 [29] | t = -0.821 | 0.415 |
| IS020 | 13.60 ± 4.36 [20] | 16.10 ± 3.21 [20] | t = -2.066 | 0.046 | 0.091 ± 0.044 [20] | 0.087 ± 0.025 [20] | t = 0.363 | 0.719 |
| IS021 | 12.33 ± 4.14 [61] | 16.11 ± 4.54 [53] | t = -4.624 | <0.001 | 0.072 ± 0.028 [61] | 0.085 ± 0.039 [53] | t = -1.959 | 0.053 |
| IS022 | 13.09 ± 3.72 [32] | 13.86 ± 2.87 [21] | t = -0.841 | 0.404 | 0.130 ± 0.036 [32] | 0.131 ± 0.025 [21] | t = -0.158 | 0.875 |
| IS023 | 7.96 ± 1.53 [53] | 7.62 ± 2.24 [24] | t = 0.670 | 0.508 | 0.113 ± 0.037 [53] | 0.106 ± 0.038 [24] | t = 0.729 | 0.470 |

Values are mean ± SD [valid n]. FD is the mean framewise displacement after dummy-volume removal, as recorded in the analysis covariate file. Comparisons use two-sided Welch t tests, with t representing MDD minus NC. P values are unadjusted.

Table S1. continued. Panel C. Clinical variables used in the analyses (MDD participants).

| **Site** | **HAMD total** | **HAMA total** | **Age at onset (years)** | **Illness duration (months)** |
| --- | --- | --- | --- | --- |
| IS001 | 20.88 ± 5.35 [218] | 22.68 ± 8.84 [220] | 34.96 ± 13.28 [200] | 46.98 ± 61.54 [211] |
| IS002 | 17.85 ± 9.24 [131] | 17.50 ± 6.48 [20] | 16.20 ± 4.13 [74] | 16.85 ± 19.52 [72] |
| IS003 | 30.07 ± 7.18 [14] | 21.21 ± 8.00 [14] | 33.78 ± 9.97 [9] | 16.50 ± 32.56 [8] |
| IS004 | 21.92 ± 5.92 [26] | — | 26.32 ± 10.41 [28] | 44.48 ± 61.32 [27] |
| IS005 | 26.61 ± 3.78 [122] | 14.97 ± 4.36 [122] | — | — |
| IS006 | 20.00 ± 6.91 [44] | 16.11 ± 9.28 [44] | 27.74 ± 9.78 [43] | 49.99 ± 66.56 [43] |
| IS007 | 23.67 ± 12.20 [18] | 10.89 ± 8.71 [18] | 19.17 ± 7.69 [18] | 15.71 ± 16.20 [18] |
| IS009 | — | — | 41.73 ± 14.46 [44] | 35.40 ± 53.21 [44] |
| IS010 | 14.04 ± 8.04 [72] | 13.76 ± 8.58 [71] | 25.99 ± 10.42 [72] | 88.29 ± 87.78 [72] |
| IS011 | 23.79 ± 9.23 [107] | — | 32.19 ± 12.49 [107] | 70.29 ± 84.12 [107] |
| IS012 | 21.27 ± 3.47 [62] | — | 29.63 ± 7.11 [62] | 6.10 ± 4.22 [62] |
| IS013 | 18.40 ± 9.59 [25] | 11.84 ± 6.05 [25] | 34.36 ± 14.12 [25] | — |
| IS014 | 30.00 ± 10.52 [126] | — | 37.07 ± 11.46 [127] | 67.79 ± 77.84 [127] |
| IS015 | — | — | 21.97 ± 5.20 [33] | 12.91 ± 15.32 [33] |
| IS016 | 25.42 ± 3.78 [33] | 13.49 ± 7.59 [49] | 22.89 ± 8.07 [82] | 30.48 ± 44.43 [82] |
| IS017 | 23.80 ± 5.90 [30] | 22.20 ± 6.85 [30] | 33.57 ± 9.08 [30] | 3.99 ± 2.63 [30] |
| IS018 | 22.34 ± 4.48 [32] | — | — | 19.94 ± 21.89 [32] |
| IS019 | 22.77 ± 5.47 [30] | 20.97 ± 6.46 [30] | 25.12 ± 10.65 [30] | 13.53 ± 14.63 [30] |
| IS020 | 24.85 ± 6.47 [20] | — | 26.65 ± 9.21 [20] | 14.55 ± 15.14 [20] |
| IS021 | 22.89 ± 2.68 [36] | 17.16 ± 6.60 [19] | 31.16 ± 11.67 [44] | 35.79 ± 45.44 [42] |
| IS022 | 22.31 ± 5.83 [32] | 21.50 ± 7.77 [32] | 31.25 ± 8.21 [20] | 5.85 ± 15.67 [20] |
| IS023 | — | — | — | — |

Values are mean ± sample SD [valid n]. HAMD and HAMA totals correspond to the supplied HAMDTotal17 and HAMA-Total fields used in the clinical analyses, without recalculation from individual items. The numbers of MDD participants with valid data are 1178, 694, 1068 and 1080 for HAMD, HAMA, age at onset and illness duration, respectively.

NC data for these clinical variables were unavailable/not applicable; consequently, neither NC estimates nor MDD–NC tests are reported. — indicates unavailable. Non-numeric entries were treated as missing, and no values were imputed.

Abbreviations: MDD, major depressive disorder; NC, normal controls; SD, standard deviation; FD, framewise displacement; HAMD, Hamilton Depression Rating Scale; HAMA, Hamilton Anxiety Rating Scale.

Table S2. Analysis-specific sample sizes for regional molecular analyses.

| **Representation** | **CAP** | **NC, n** | **MDD, n** | **Total, n** | **Parcels, n** |
| --- | --- | --- | --- | --- | --- |
| Joint shared centroid | CAP-1 | 1126 | 1383 | 2509 | 400 |
| Joint shared centroid | CAP-2 | 1123 | 1390 | 2513 | 400 |
| Joint shared centroid | CAP-3 | 1130 | 1404 | 2534 | 400 |
| Joint shared centroid | CAP-4 | 1130 | 1404 | 2534 | 400 |
| Joint shared centroid | CAP-5 | 1130 | 1404 | 2534 | 400 |
| Joint shared centroid | CAP-6 | 1130 | 1404 | 2534 | 400 |
| Joint shared centroid | CAP-7 | 1130 | 1404 | 2534 | 400 |
| Separate GM activation | CAP-1 | 1125 | 1393 | 2518 | 400 |
| Separate GM activation | CAP-2 | 1126 | 1398 | 2524 | 400 |
| Separate GM activation | CAP-3 | 1130 | 1404 | 2534 | 400 |
| Separate GM activation | CAP-4 | 1130 | 1404 | 2534 | 400 |
| Separate GM activation | CAP-5 | 1130 | 1404 | 2534 | 400 |
| Separate GM activation | CAP-6 | 1130 | 1404 | 2534 | 400 |
| Separate GM activation | CAP-7 | 1130 | 1404 | 2534 | 400 |
| Separate WM activation | CAP-1 | 1120 | 1366 | 2486 | 400 |
| Separate WM activation | CAP-2 | 1119 | 1373 | 2492 | 400 |
| Separate WM activation | CAP-3 | 1130 | 1404 | 2534 | 400 |
| Separate WM activation | CAP-4 | 1130 | 1404 | 2534 | 400 |
| Separate WM activation | CAP-5 | 1130 | 1404 | 2534 | 400 |
| Separate WM activation | CAP-6 | 1128 | 1387 | 2515 | 400 |
| Separate WM activation | CAP-7 | 1130 | 1404 | 2534 | 400 |

Notes. Counts are taken from the parcel-wise phenotype A group-effect models. Each row has the same participant counts across all 400 parcels. Eligibility was defined within each representation and CAP by complete regional data across all seven depth conditions. Counts can therefore differ from the full cohort size. Models were adjusted for age, sex, education, mean framewise displacement and site. Phenotype A is the participant-level mean across depths, whereas phenotype L is the linear depth slope. The temporal coordination analyses in Table S16 used 1,404 MDD and 1,130 NC participants. Conditional state and lagged-transition analyses may have smaller eligible samples, which are reported separately. GM, gray matter; WM, white matter; CAP, coactivation pattern.

**Table S3. Demographic and clinical characteristics. DIRECT Phase II CAP cohort: 2551 participants from 22 sites (MDD, n = 1417; NC, n = 1134).**

| Characteristic | MDD | Valid n | NC | Valid n | Test statistic | P |
| --- | --- | --- | --- | --- | --- | --- |
| Participants, n | 1417 | 1417 | 1134 | 1134 | — | — |
| Sex, male/female | 504/909 | 1413 | 484/650 | 1134 | χ² = 13.028 | <0.001 |
| Age (years) | 32.46 ± 13.05 | 1409 | 31.35 ± 12.33 | 1134 | t = 2.210 | 0.027 |
| Education (years) | 11.94 ± 3.69 | 1404 | 13.80 ± 3.52 | 1130 | t = -12.920 | <0.001 |
| Mean FD (mm) | 0.099 ± 0.042 | 1417 | 0.104 ± 0.041 | 1134 | t = -3.474 | <0.001 |
| HAMD total | 22.52 ± 8.27 | 1178 | — | 0 | — | — |
| HAMA total | 18.17 ± 8.63 | 694 | — | 0 | — | — |
| Age at onset (years) | 30.17 ± 12.52 | 1068 | — | 0 | — | — |
| Illness duration (months) | 41.96 ± 62.58 | 1080 | — | 0 | — | — |

Values are mean ± sample SD unless otherwise indicated. Sex is shown as male/female counts. Summaries use available observations; valid n is shown for each variable. No missing values were imputed. — indicates unavailable/not applicable. Between-group comparisons use two-sided Welch t tests for continuous variables and Pearson χ² for sex. Positive t indicates MDD > NC. P values are unadjusted descriptive comparisons. NC clinical scale and illness-history data were unavailable/not applicable, so no tests were performed for those rows. We used the 17-item HAMD total and the HAMA total provided by the contributing sites. Mean FD is mean framewise displacement after dummy-volume removal. The full CAP cohort is summarized here. Complete age, sex, education, and mean FD were available for 2534 participants (1404 MDD; 1130 NC), constituting the complete-covariate subset for fully adjusted models before any feature-specific missingness. Abbreviations: MDD, major depressive disorder; NC, normal controls; SD, standard deviation; FD, framewise displacement; HAMD, Hamilton Depression Rating Scale; HAMA, Hamilton Anxiety Rating Scale.

Table S4. Effective sampling points at different white matter depths.

| **Depth of WM** | **Valid paired point count** | **Median per brain region** | **Minimum per brain region** |
| --- | --- | --- | --- |
| 1 mm | 54,245 | 130.0 | 12 |
| 2 mm | 53,474 | 128.0 | 12 |
| 3 mm | 50,762 | 119.0 | 9 |
| 4 mm | 45,297 | 106.0 | 8 |
| 5 mm | 39,253 | 92.0 | 5 |
| 6 mm | 34,025 | 78.0 | 4 |
| 7 mm | 29,746 | 66.5 | 3 |

Table S5. Summary of meta-analytic CAP functional decoding.

A. Significant positive and negative associations among all 123 reference terms.

| **CAP** | **Positive, n** | **Negative, n** | **Total, n / 123** |
| --- | --- | --- | --- |
| CAP-1 | 8 | 9 | 17 |
| CAP-2 | 10 | 9 | 19 |
| CAP-3 | 35 | 20 | 55 |
| CAP-4 | 10 | 30 | 40 |
| CAP-5 | 15 | 14 | 29 |
| CAP-6 | 18 | 33 | 51 |
| CAP-7 | 35 | 21 | 56 |

B. Correlations and adjusted q values for the seven representative radar coordinates.

| **Term** | **CAP1** | **CAP2** | **CAP3** | **CAP4** | **CAP5** | **CAP6** | **CAP7** |
| --- | --- | --- | --- | --- | --- | --- | --- |
| Emotion regulation | **0.386 (0.0107)** | **-0.427 (0.0079)** | **0.331 (0.0268)** | -0.177 (0.3584) | -0.157 (0.5212) | **-0.437 (0.0058)** | **0.458 (0.0038)** |
| Autobiographical memory | 0.226 (0.2129) | -0.282 (0.1141) | **0.717 (0.0012)** | -0.316 (0.0506) | **-0.429 (0.0025)** | **-0.610 (0.0012)** | **0.640 (0.0012)** |
| Social cognition | 0.161 (0.2479) | -0.212 (0.1109) | **0.645 (0.0012)** | **-0.312 (0.0276)** | **-0.376 (0.0025)** | **-0.522 (0.0012)** | **0.547 (0.0012)** |
| Response selection | -0.083 (0.7584) | 0.159 (0.5211) | **-0.387 (0.0016)** | -0.127 (0.3672) | **0.491 (0.0016)** | 0.237 (0.1224) | -0.284 (0.0520) |
| Motor coordination | -0.230 (0.3683) | 0.277 (0.2205) | **-0.705 (0.0012)** | 0.342 (0.0608) | 0.361 (0.0518) | **0.604 (0.0012)** | **-0.628 (0.0012)** |
| Multisensory processing | **-0.466 (0.0025)** | **0.493 (0.0020)** | **-0.523 (0.0012)** | **0.398 (0.0012)** | 0.163 (0.7484) | **0.628 (0.0016)** | **-0.634 (0.0012)** |
| Visual attention | **-0.407 (0.0217)** | **0.441 (0.0083)** | -0.146 (0.5014) | 0.082 (0.6803) | 0.070 (0.8915) | **0.338 (0.0489)** | **-0.352 (0.0258)** |

Notes. Panel A counts q < 0.05 associations across all 123 terms, totaling 267 of 861 CAP-by-term tests. Panel B reports signed Spearman ρ with BH-FDR q in parentheses; bold cells indicate q < 0.05. Each correlation uses 400 corresponding cortical parcels from the covariate-adjusted NC joint shared GM–WM phenotype A map and a normative Neurosynth association-test map. Inference used 10,000 hemisphere-preserving spatial spins, a two-sided null-mean-centered test, and one FDR family of 861 comparisons. The seven terms were selected for visualization after this full analysis; they were not tested as a separate family. “Motor coordination” and “Multisensory processing” correspond to reference terms “coordination” and “multisensory”, respectively. Coefficients were not rescaled within CAPs. NC, normal controls; CAP, coactivation pattern.

Table S6. Complete meta-analytic functional decoding of the seven CAPs. Terms 1–25 of 123.

| **Reference term** | **CAP1** | **CAP2** | **CAP3** | **CAP4** | **CAP5** | **CAP6** | **CAP7** |
| --- | --- | --- | --- | --- | --- | --- | --- |
| action | -0.312 (0.2123) | 0.375 (0.0906) | **-0.475 (0.0012)** | 0.091 (0.6764) | **0.383 (0.0119)** | **0.472 (0.0025)** | **-0.520 (0.0012)** |
| adaptation | -0.285 (0.0937) | 0.317 (0.0630) | -0.126 (0.4935) | 0.071 (0.6650) | 0.086 (0.7598) | 0.244 (0.1453) | -0.254 (0.1306) |
| addiction | 0.249 (0.2313) | -0.282 (0.1517) | 0.231 (0.0914) | -0.047 (0.8401) | -0.208 (0.2212) | -0.257 (0.1425) | 0.287 (0.0718) |
| anticipation | 0.145 (0.6158) | -0.137 (0.6560) | -0.240 (0.0673) | 0.146 (0.3513) | 0.097 (0.6187) | 0.143 (0.4400) | -0.130 (0.4789) |
| anxiety | 0.304 (0.1478) | -0.341 (0.0853) | 0.215 (0.0988) | -0.044 (0.8780) | -0.183 (0.2716) | -0.266 (0.1380) | 0.296 (0.0811) |
| arousal | 0.228 (0.2407) | -0.261 (0.1432) | 0.062 (0.8005) | 0.098 (0.4993) | -0.164 (0.3072) | -0.110 (0.6273) | 0.131 (0.5407) |
| association | 0.201 (0.2122) | -0.254 (0.0919) | **0.227 (0.0445)** | 0.009 (0.9551) | **-0.243 (0.0447)** | -0.245 (0.0924) | **0.281 (0.0381)** |
| attention | -0.271 (0.2947) | 0.329 (0.1895) | -0.147 (0.5967) | -0.147 (0.3751) | 0.279 (0.2059) | 0.229 (0.3516) | -0.266 (0.2260) |
| autobiographical memory | 0.226 (0.2129) | -0.282 (0.1141) | **0.717 (0.0012)** | -0.316 (0.0506) | **-0.429 (0.0025)** | **-0.610 (0.0012)** | **0.640 (0.0012)** |
| balance | 0.123 (0.4462) | -0.134 (0.3869) | -0.109 (0.6187) | 0.039 (0.8305) | 0.048 (0.8923) | -0.011 (0.9208) | 0.023 (0.8564) |
| belief | 0.132 (0.2541) | -0.157 (0.1770) | **0.416 (0.0012)** | **-0.388 (0.0016)** | -0.092 (0.5578) | **-0.407 (0.0012)** | **0.402 (0.0012)** |
| categorization | -0.146 (0.4073) | 0.161 (0.3705) | -0.116 (0.5911) | 0.171 (0.1453) | 0.030 (0.9851) | 0.187 (0.2755) | -0.179 (0.3138) |
| cognitive control | 0.328 (0.0679) | -0.298 (0.1189) | 0.282 (0.1572) | **-0.636 (0.0012)** | **0.288 (0.0375)** | **-0.481 (0.0016)** | **0.438 (0.0033)** |
| communication | -0.003 (0.9489) | -0.032 (0.8564) | 0.187 (0.1907) | -0.033 (0.7893) | -0.169 (0.3619) | -0.135 (0.3291) | 0.138 (0.3275) |
| competition | -0.145 (0.3498) | 0.175 (0.2169) | 0.026 (0.9912) | -0.116 (0.3349) | 0.113 (0.2497) | 0.055 (0.6786) | -0.075 (0.5513) |
| concept | 0.180 (0.0556) | **-0.210 (0.0238)** | **0.224 (0.0189)** | -0.165 (0.0993) | -0.097 (0.5121) | **-0.262 (0.0138)** | **0.256 (0.0104)** |
| consciousness | -0.045 (0.7848) | 0.042 (0.8281) | 0.004 (0.7611) | -0.032 (0.8305) | 0.002 (0.8399) | 0.011 (0.9784) | -0.012 (0.9654) |
| consolidation | -0.013 (0.8828) | -0.031 (0.8923) | 0.165 (0.1759) | 0.061 (0.6187) | **-0.265 (0.0074)** | -0.103 (0.3751) | 0.131 (0.2273) |
| context | 0.293 (0.0560) | **-0.322 (0.0271)** | **0.425 (0.0016)** | -0.230 (0.0669) | -0.200 (0.3143) | **-0.473 (0.0016)** | **0.485 (0.0016)** |
| coordination | -0.230 (0.3683) | 0.277 (0.2205) | **-0.705 (0.0012)** | 0.342 (0.0608) | 0.361 (0.0518) | **0.604 (0.0012)** | **-0.628 (0.0012)** |
| decision | 0.163 (0.4114) | -0.171 (0.4105) | **0.421 (0.0025)** | **-0.320 (0.0029)** | -0.113 (0.7894) | **-0.402 (0.0012)** | **0.413 (0.0012)** |
| decision making | 0.143 (0.6082) | -0.156 (0.5940) | **0.349 (0.0436)** | **-0.280 (0.0381)** | -0.100 (0.8305) | -0.329 (0.0803) | 0.343 (0.0608) |
| detection | -0.259 (0.1365) | 0.301 (0.0622) | -0.253 (0.1129) | 0.030 (0.8768) | 0.273 (0.0997) | **0.307 (0.0436)** | **-0.328 (0.0227)** |
| discrimination | -0.158 (0.4360) | 0.205 (0.2916) | **-0.344 (0.0086)** | 0.149 (0.2984) | 0.224 (0.3015) | **0.342 (0.0233)** | **-0.363 (0.0119)** |
| distraction | **0.200 (0.0411)** | -0.173 (0.0891) | 0.086 (0.2158) | **-0.246 (0.0020)** | 0.182 (0.0697) | **-0.192 (0.0245)** | **0.179 (0.0375)** |

Notes. Cells show signed Spearman ρ (BH-FDR q); bold indicates q < 0.05. All 861 tests used 400 parcels and 10,000 hemisphere-preserving spatial spins with a two-sided null-mean-centered test. FDR correction precedes term selection and covers the complete 7 × 123 matrix. All coefficients, including nonsignificant associations, are shown. Terms retain the reference vocabulary and are ordered alphabetically. Full-precision coefficients and empirical P and adjusted q values are provided in Meta_CAP_Neurosynth_all123.csv.

**Table S6. continued. Terms 26–50 of 123.**

| **Reference term** | **CAP1** | **CAP2** | **CAP3** | **CAP4** | **CAP5** | **CAP6** | **CAP7** |
| --- | --- | --- | --- | --- | --- | --- | --- |
| eating | 0.152 (0.5968) | -0.196 (0.4637) | 0.159 (0.5280) | 0.156 (0.2694) | -0.286 (0.1129) | -0.124 (0.7210) | 0.163 (0.5648) |
| efficiency | -0.080 (0.6690) | 0.111 (0.5319) | 0.050 (0.7400) | **-0.260 (0.0058)** | 0.157 (0.2129) | -0.058 (0.6273) | 0.028 (0.8214) |
| effort | 0.056 (0.7815) | -0.012 (0.9425) | **-0.278 (0.0327)** | -0.121 (0.3585) | **0.380 (0.0012)** | 0.087 (0.6990) | -0.129 (0.4998) |
| emotion | 0.303 (0.2358) | -0.364 (0.1129) | **0.456 (0.0033)** | -0.093 (0.6868) | **-0.359 (0.0460)** | **-0.450 (0.0029)** | **0.490 (0.0020)** |
| emotion regulation | **0.386 (0.0107)** | **-0.427 (0.0079)** | **0.331 (0.0268)** | -0.177 (0.3584) | -0.157 (0.5212) | **-0.437 (0.0058)** | **0.458 (0.0038)** |
| empathy | 0.257 (0.0712) | -0.263 (0.0542) | 0.027 (0.7772) | -0.039 (0.8089) | -0.017 (0.8546) | -0.132 (0.3219) | 0.121 (0.3738) |
| encoding | -0.054 (0.8031) | 0.060 (0.7607) | 0.269 (0.1517) | -0.140 (0.3824) | -0.124 (0.6803) | -0.131 (0.5637) | 0.129 (0.5724) |
| episodic memory | 0.158 (0.3699) | -0.200 (0.2635) | **0.646 (0.0012)** | **-0.326 (0.0411)** | -0.351 (0.0640) | **-0.524 (0.0012)** | **0.545 (0.0012)** |
| expectancy | 0.214 (0.0797) | -0.167 (0.2129) | 0.007 (0.9549) | **-0.324 (0.0020)** | **0.302 (0.0016)** | -0.170 (0.2741) | 0.137 (0.4068) |
| expertise | -0.155 (0.3007) | 0.166 (0.2499) | -0.043 (0.5119) | 0.084 (0.5580) | -0.025 (0.9939) | 0.151 (0.2617) | -0.156 (0.2129) |
| extinction | 0.186 (0.4498) | -0.225 (0.2910) | 0.190 (0.2123) | 0.060 (0.6580) | -0.259 (0.0637) | -0.195 (0.2577) | 0.228 (0.1447) |
| face recognition | -0.066 (0.8217) | 0.039 (0.8973) | 0.296 (0.0688) | 0.050 (0.7294) | **-0.365 (0.0041)** | -0.083 (0.8305) | 0.115 (0.7400) |
| facial expression | -0.162 (0.5637) | 0.146 (0.6504) | -0.072 (0.7149) | **0.357 (0.0016)** | -0.220 (0.1224) | 0.243 (0.2573) | -0.209 (0.3620) |
| familiarity | 0.110 (0.5637) | -0.118 (0.5453) | **0.389 (0.0033)** | -0.266 (0.1165) | -0.153 (0.3763) | **-0.341 (0.0200)** | **0.332 (0.0214)** |
| fear | 0.213 (0.4323) | -0.249 (0.3414) | 0.167 (0.3524) | 0.102 (0.5086) | -0.236 (0.2281) | -0.161 (0.4757) | 0.196 (0.3498) |
| fixation | **-0.429 (0.0074)** | **0.458 (0.0046)** | -0.149 (0.2721) | 0.165 (0.3630) | 0.009 (0.9449) | **0.390 (0.0012)** | **-0.394 (0.0012)** |
| focus | -0.016 (0.9111) | 0.007 (0.9820) | 0.028 (0.5512) | -0.050 (0.5145) | -0.001 (0.8305) | -0.029 (0.6803) | 0.021 (0.7143) |
| gaze | **-0.523 (0.0016)** | **0.545 (0.0012)** | **-0.275 (0.0273)** | **0.325 (0.0141)** | -0.006 (0.9586) | **0.523 (0.0020)** | **-0.517 (0.0016)** |
| goal | -0.031 (0.9620) | 0.060 (0.8768) | 0.018 (0.9425) | **-0.293 (0.0029)** | 0.194 (0.1985) | -0.047 (0.7277) | -0.001 (0.9425) |
| hyperactivity | 0.204 (0.2167) | -0.236 (0.1277) | 0.139 (0.3513) | -0.081 (0.6644) | -0.088 (0.6158) | -0.208 (0.2029) | 0.226 (0.1437) |
| imagery | -0.308 (0.2202) | 0.365 (0.0924) | **-0.609 (0.0012)** | 0.282 (0.0893) | 0.344 (0.0888) | **0.581 (0.0012)** | **-0.614 (0.0012)** |
| impulsivity | 0.222 (0.2059) | -0.243 (0.1634) | **0.340 (0.0174)** | -0.245 (0.0560) | -0.133 (0.6202) | **-0.351 (0.0141)** | **0.368 (0.0107)** |
| induction | -0.018 (0.8533) | 0.008 (0.9128) | **-0.242 (0.0458)** | 0.194 (0.1744) | 0.063 (0.8274) | 0.190 (0.2039) | -0.176 (0.2499) |
| inference | 0.002 (0.9449) | -0.049 (0.5967) | **0.310 (0.0016)** | -0.084 (0.3774) | **-0.268 (0.0016)** | **-0.241 (0.0029)** | **0.260 (0.0016)** |
| inhibition | 0.087 (0.8225) | -0.052 (0.8923) | -0.196 (0.2281) | -0.123 (0.5197) | 0.293 (0.0750) | 0.042 (0.9271) | -0.050 (0.8959) |

Notes. Cells show signed Spearman ρ (BH-FDR q); bold indicates q < 0.05. All 861 tests used 400 parcels and 10,000 hemisphere-preserving spatial spins with a two-sided null-mean-centered test. FDR correction precedes term selection and covers the complete 7 × 123 matrix. All coefficients, including nonsignificant associations, are shown. Terms retain the reference vocabulary and are ordered alphabetically. Full-precision coefficients and empirical P and adjusted q values are provided in Meta_CAP_Neurosynth_all123.csv.

**Table S6. continued. Terms 51–75 of 123.**

| **Reference term** | **CAP1** | **CAP2** | **CAP3** | **CAP4** | **CAP5** | **CAP6** | **CAP7** |
| --- | --- | --- | --- | --- | --- | --- | --- |
| insight | 0.108 (0.6678) | -0.138 (0.5071) | 0.055 (0.5724) | 0.089 (0.4627) | -0.183 (0.0891) | -0.044 (0.8294) | 0.067 (0.7059) |
| integration | -0.177 (0.4998) | 0.165 (0.5724) | -0.164 (0.5086) | **0.248 (0.0445)** | -0.057 (0.6504) | 0.226 (0.2778) | -0.225 (0.2890) |
| intelligence | -0.012 (0.9128) | 0.001 (0.8848) | 0.274 (0.0875) | **-0.249 (0.0373)** | -0.059 (0.9864) | -0.212 (0.1281) | 0.207 (0.1468) |
| intention | 0.009 (0.8005) | -0.027 (0.7092) | **0.528 (0.0012)** | **-0.402 (0.0020)** | -0.187 (0.1949) | **-0.397 (0.0016)** | **0.392 (0.0020)** |
| interference | -0.087 (0.6306) | 0.160 (0.3435) | -0.246 (0.0669) | -0.181 (0.2050) | **0.440 (0.0012)** | 0.181 (0.1677) | -0.225 (0.0756) |
| judgment | 0.116 (0.3284) | -0.112 (0.3588) | **0.509 (0.0016)** | **-0.450 (0.0016)** | -0.090 (0.7893) | **-0.456 (0.0016)** | **0.436 (0.0020)** |
| knowledge | 0.145 (0.2029) | -0.163 (0.1654) | **0.472 (0.0012)** | **-0.359 (0.0012)** | -0.149 (0.3895) | **-0.430 (0.0012)** | **0.426 (0.0012)** |
| language | 0.058 (0.8546) | -0.064 (0.8564) | 0.194 (0.2762) | -0.111 (0.4360) | -0.039 (0.9912) | -0.194 (0.3895) | 0.186 (0.4185) |
| language comprehension | 0.202 (0.1042) | -0.230 (0.0557) | **0.291 (0.0071)** | -0.179 (0.1426) | -0.106 (0.6081) | **-0.363 (0.0012)** | **0.367 (0.0012)** |
| learning | -0.025 (0.7528) | 0.046 (0.6187) | -0.104 (0.2990) | 0.071 (0.5418) | 0.045 (0.6786) | 0.090 (0.3269) | -0.087 (0.3206) |
| listening | **0.330 (0.0449)** | **-0.354 (0.0230)** | -0.051 (0.9864) | -0.024 (0.8485) | 0.090 (0.8230) | -0.198 (0.1591) | 0.196 (0.1581) |
| localization | **-0.283 (0.0445)** | **0.289 (0.0349)** | **-0.306 (0.0101)** | **0.302 (0.0171)** | 0.047 (0.8305) | **0.374 (0.0016)** | **-0.366 (0.0020)** |
| loss | 0.159 (0.4620) | -0.219 (0.2570) | 0.165 (0.3034) | 0.155 (0.3098) | **-0.293 (0.0335)** | -0.157 (0.4014) | 0.208 (0.1949) |
| maintenance | 0.059 (0.7368) | -0.006 (0.9912) | -0.006 (0.9784) | **-0.357 (0.0054)** | **0.358 (0.0041)** | -0.110 (0.4436) | 0.065 (0.6603) |
| manipulation | -0.122 (0.5319) | 0.191 (0.2782) | **-0.292 (0.0349)** | -0.128 (0.3683) | **0.420 (0.0012)** | 0.230 (0.1399) | **-0.279 (0.0375)** |
| meaning | 0.034 (0.8923) | -0.060 (0.8401) | **0.284 (0.0227)** | -0.097 (0.3542) | -0.173 (0.3980) | -0.225 (0.1042) | 0.232 (0.0853) |
| memory | 0.210 (0.1677) | -0.196 (0.2281) | **0.469 (0.0029)** | **-0.515 (0.0012)** | -0.002 (0.8281) | **-0.491 (0.0012)** | **0.468 (0.0012)** |
| memory retrieval | **0.366 (0.0054)** | **-0.385 (0.0050)** | **0.618 (0.0012)** | **-0.603 (0.0012)** | -0.079 (0.7965) | **-0.695 (0.0012)** | **0.681 (0.0012)** |
| mental imagery | -0.208 (0.3138) | 0.250 (0.1973) | -0.190 (0.1141) | 0.044 (0.7532) | 0.156 (0.3866) | 0.246 (0.0802) | **-0.272 (0.0333)** |
| monitoring | 0.117 (0.7498) | -0.080 (0.8305) | -0.001 (0.8923) | **-0.382 (0.0020)** | **0.331 (0.0381)** | -0.160 (0.4649) | 0.125 (0.5881) |
| mood | 0.211 (0.4480) | -0.266 (0.2942) | **0.500 (0.0016)** | -0.111 (0.6331) | **-0.416 (0.0025)** | **-0.426 (0.0041)** | **0.476 (0.0016)** |
| morphology | 0.113 (0.6498) | -0.156 (0.5084) | 0.035 (0.9912) | 0.141 (0.3524) | -0.169 (0.4360) | -0.077 (0.8031) | 0.110 (0.6843) |
| motor control | -0.210 (0.4993) | 0.254 (0.3574) | **-0.627 (0.0012)** | 0.289 (0.1163) | 0.347 (0.0944) | **0.520 (0.0020)** | **-0.543 (0.0016)** |
| movement | -0.341 (0.1759) | 0.387 (0.0909) | **-0.715 (0.0012)** | **0.392 (0.0349)** | 0.339 (0.1517) | **0.672 (0.0012)** | **-0.692 (0.0012)** |
| multisensory | **-0.466 (0.0025)** | **0.493 (0.0020)** | **-0.523 (0.0012)** | **0.398 (0.0012)** | 0.163 (0.7484) | **0.628 (0.0016)** | **-0.634 (0.0012)** |

Notes. Cells show signed Spearman ρ (BH-FDR q); bold indicates q < 0.05. All 861 tests used 400 parcels and 10,000 hemisphere-preserving spatial spins with a two-sided null-mean-centered test. FDR correction precedes term selection and covers the complete 7 × 123 matrix. All coefficients, including nonsignificant associations, are shown. Terms retain the reference vocabulary and are ordered alphabetically. Full-precision coefficients and empirical P and adjusted q values are provided in Meta_CAP_Neurosynth_all123.csv.

**Table S6. continued. Terms 76–100 of 123.**

| **Reference term** | **CAP1** | **CAP2** | **CAP3** | **CAP4** | **CAP5** | **CAP6** | **CAP7** |
| --- | --- | --- | --- | --- | --- | --- | --- |
| naming | -0.182 (0.3778) | 0.183 (0.3719) | -0.155 (0.1778) | **0.290 (0.0101)** | -0.068 (0.8305) | 0.262 (0.0504) | **-0.265 (0.0399)** |
| navigation | -0.217 (0.4303) | 0.237 (0.3778) | 0.043 (0.9426) | 0.010 (0.8915) | -0.054 (0.9018) | 0.123 (0.5753) | -0.117 (0.5968) |
| object recognition | **-0.464 (0.0211)** | **0.480 (0.0171)** | -0.064 (0.7210) | 0.230 (0.1956) | -0.134 (0.6803) | 0.375 (0.0612) | -0.365 (0.0686) |
| pain | 0.266 (0.2570) | -0.265 (0.2541) | **-0.364 (0.0424)** | 0.125 (0.5016) | 0.206 (0.4836) | 0.101 (0.7149) | -0.101 (0.7210) |
| perception | -0.365 (0.0782) | 0.384 (0.0626) | **-0.476 (0.0020)** | **0.513 (0.0016)** | 0.037 (0.9425) | **0.602 (0.0012)** | **-0.591 (0.0012)** |
| planning | -0.255 (0.4305) | 0.322 (0.2260) | **-0.482 (0.0012)** | 0.062 (0.7815) | **0.417 (0.0020)** | **0.425 (0.0041)** | **-0.471 (0.0016)** |
| priming | -0.070 (0.7532) | 0.052 (0.8281) | **0.385 (0.0016)** | -0.096 (0.5145) | **-0.287 (0.0436)** | -0.196 (0.1778) | 0.224 (0.1042) |
| psychosis | 0.165 (0.2169) | -0.213 (0.0919) | **0.376 (0.0020)** | -0.086 (0.5911) | **-0.296 (0.0050)** | **-0.311 (0.0101)** | **0.351 (0.0054)** |
| reading | -0.194 (0.5119) | 0.201 (0.4993) | 0.066 (0.7893) | 0.060 (0.8305) | -0.078 (0.8281) | 0.067 (0.8051) | -0.072 (0.7894) |
| reasoning | 0.087 (0.5086) | -0.091 (0.5222) | **0.523 (0.0012)** | **-0.476 (0.0016)** | -0.073 (0.9147) | **-0.442 (0.0012)** | **0.430 (0.0012)** |
| recall | **0.360 (0.0171)** | **-0.400 (0.0086)** | **0.519 (0.0012)** | **-0.412 (0.0029)** | -0.161 (0.4303) | **-0.626 (0.0012)** | **0.629 (0.0012)** |
| recognition | 0.026 (0.9487) | -0.036 (0.9425) | **0.469 (0.0033)** | -0.224 (0.2917) | -0.236 (0.3275) | -0.301 (0.1655) | 0.308 (0.1333) |
| rehearsal | -0.056 (0.8031) | 0.108 (0.5911) | **-0.315 (0.0067)** | -0.009 (0.9364) | **0.348 (0.0071)** | 0.185 (0.2448) | -0.231 (0.1129) |
| reinforcement learning | 0.071 (0.7265) | -0.063 (0.7680) | 0.172 (0.1813) | -0.069 (0.6655) | -0.073 (0.6827) | -0.111 (0.4480) | 0.123 (0.3895) |
| response inhibition | 0.128 (0.6246) | -0.083 (0.7941) | -0.071 (0.7149) | **-0.278 (0.0174)** | **0.313 (0.0029)** | -0.074 (0.7484) | 0.046 (0.8399) |
| response selection | -0.083 (0.7584) | 0.159 (0.5211) | **-0.387 (0.0016)** | -0.127 (0.3672) | **0.491 (0.0016)** | 0.237 (0.1224) | -0.284 (0.0520) |
| retention | -0.109 (0.2389) | 0.117 (0.2029) | 0.002 (0.9784) | -0.094 (0.3457) | 0.072 (0.4436) | 0.012 (0.9310) | -0.031 (0.7815) |
| retrieval | 0.221 (0.1163) | -0.232 (0.1130) | **0.582 (0.0012)** | **-0.506 (0.0016)** | -0.111 (0.6970) | **-0.576 (0.0012)** | **0.566 (0.0012)** |
| reward anticipation | -0.008 (0.9057) | -0.005 (0.9426) | 0.068 (0.9861) | 0.146 (0.2723) | -0.207 (0.2721) | 0.050 (0.5982) | -0.014 (0.7400) |
| rhythm | -0.047 (0.8546) | 0.059 (0.8305) | **-0.520 (0.0038)** | **0.348 (0.0012)** | 0.167 (0.5371) | **0.383 (0.0086)** | **-0.388 (0.0101)** |
| risk | 0.248 (0.2680) | -0.290 (0.1772) | **0.448 (0.0041)** | -0.255 (0.1097) | -0.233 (0.3476) | **-0.441 (0.0071)** | **0.471 (0.0046)** |
| rule | 0.006 (0.9208) | 0.026 (0.9425) | 0.180 (0.4360) | **-0.408 (0.0012)** | 0.199 (0.0517) | -0.194 (0.1956) | 0.157 (0.3524) |
| salience | 0.209 (0.2456) | -0.252 (0.1513) | **0.384 (0.0016)** | -0.146 (0.3620) | -0.268 (0.0877) | **-0.325 (0.0255)** | **0.348 (0.0090)** |
| search | -0.192 (0.1967) | 0.192 (0.2044) | 0.163 (0.1964) | -0.057 (0.6786) | -0.101 (0.4998) | 0.005 (0.9939) | -0.008 (0.9879) |
| selective attention | **-0.280 (0.0086)** | **0.320 (0.0041)** | **-0.276 (0.0025)** | 0.100 (0.3866) | **0.216 (0.0461)** | **0.352 (0.0016)** | **-0.381 (0.0016)** |

Notes. Cells show signed Spearman ρ (BH-FDR q); bold indicates q < 0.05. All 861 tests used 400 parcels and 10,000 hemisphere-preserving spatial spins with a two-sided null-mean-centered test. FDR correction precedes term selection and covers the complete 7 × 123 matrix. All coefficients, including nonsignificant associations, are shown. Terms retain the reference vocabulary and are ordered alphabetically. Full-precision coefficients and empirical P and adjusted q values are provided in Meta_CAP_Neurosynth_all123.csv.

**Table S6. continued. Terms 101–123 of 123.**

| **Reference term** | **CAP1** | **CAP2** | **CAP3** | **CAP4** | **CAP5** | **CAP6** | **CAP7** |
| --- | --- | --- | --- | --- | --- | --- | --- |
| semantic memory | 0.299 (0.1179) | -0.345 (0.0520) | **0.532 (0.0012)** | -0.275 (0.0782) | -0.261 (0.1677) | **-0.550 (0.0012)** | **0.563 (0.0012)** |
| sentence comprehension | 0.150 (0.1189) | -0.138 (0.1695) | 0.196 (0.0555) | **-0.295 (0.0050)** | 0.089 (0.3895) | **-0.268 (0.0025)** | **0.243 (0.0041)** |
| skill | -0.270 (0.0742) | **0.300 (0.0412)** | -0.214 (0.0997) | 0.051 (0.7400) | 0.121 (0.5121) | 0.244 (0.0650) | -0.255 (0.0540) |
| sleep | -0.008 (0.8281) | -0.036 (0.9414) | 0.073 (0.5418) | 0.173 (0.1399) | **-0.238 (0.0399)** | -0.022 (0.9617) | 0.063 (0.7210) |
| social cognition | 0.161 (0.2479) | -0.212 (0.1109) | **0.645 (0.0012)** | **-0.312 (0.0276)** | **-0.376 (0.0025)** | **-0.522 (0.0012)** | **0.547 (0.0012)** |
| spatial attention | **-0.385 (0.0412)** | **0.450 (0.0054)** | **-0.276 (0.0500)** | 0.045 (0.8225) | 0.244 (0.2059) | **0.411 (0.0033)** | **-0.440 (0.0020)** |
| speech perception | 0.016 (0.9310) | -0.028 (0.8848) | -0.142 (0.5171) | 0.176 (0.1964) | 0.035 (0.9425) | 0.093 (0.5911) | -0.081 (0.6678) |
| speech production | 0.039 (0.8324) | -0.038 (0.8390) | -0.298 (0.0814) | 0.255 (0.0687) | 0.106 (0.6917) | 0.180 (0.3335) | -0.180 (0.3439) |
| strategy | **0.284 (0.0033)** | **-0.278 (0.0029)** | **0.368 (0.0016)** | **-0.429 (0.0016)** | 0.054 (0.6029) | **-0.453 (0.0016)** | **0.432 (0.0016)** |
| strength | 0.082 (0.5081) | -0.098 (0.3934) | 0.031 (0.6981) | -0.006 (0.9530) | -0.056 (0.5119) | -0.066 (0.5648) | 0.078 (0.4729) |
| stress | 0.308 (0.1244) | **-0.368 (0.0411)** | 0.188 (0.2123) | 0.063 (0.7281) | -0.263 (0.0919) | -0.270 (0.1163) | **0.307 (0.0441)** |
| sustained attention | **0.230 (0.0453)** | -0.218 (0.0686) | -0.124 (0.4480) | -0.093 (0.3286) | 0.188 (0.1949) | -0.073 (0.4993) | 0.058 (0.5753) |
| task difficulty | 0.019 (0.9489) | 0.025 (0.8305) | -0.018 (0.9926) | **-0.277 (0.0126)** | **0.289 (0.0104)** | -0.057 (0.6602) | 0.020 (0.8564) |
| thought | **0.272 (0.0074)** | **-0.290 (0.0041)** | **0.452 (0.0029)** | **-0.357 (0.0020)** | -0.126 (0.5386) | **-0.490 (0.0012)** | **0.492 (0.0012)** |
| uncertainty | 0.141 (0.2260) | -0.131 (0.2637) | **0.248 (0.0235)** | **-0.367 (0.0016)** | 0.069 (0.5988) | **-0.289 (0.0016)** | **0.274 (0.0020)** |
| updating | -0.018 (0.9310) | 0.070 (0.7309) | 0.031 (0.9018) | **-0.276 (0.0122)** | 0.210 (0.0825) | -0.054 (0.7400) | 0.027 (0.8848) |
| utility | 0.089 (0.5287) | -0.136 (0.3082) | 0.180 (0.0579) | 0.015 (0.9425) | **-0.235 (0.0098)** | -0.170 (0.1770) | 0.199 (0.0715) |
| valence | 0.266 (0.3705) | -0.325 (0.2059) | **0.433 (0.0016)** | -0.034 (0.9040) | **-0.394 (0.0033)** | **-0.397 (0.0054)** | **0.438 (0.0025)** |
| verbal fluency | 0.078 (0.5724) | -0.079 (0.6021) | 0.056 (0.7893) | -0.072 (0.4800) | 0.046 (0.5603) | -0.110 (0.3488) | 0.104 (0.3865) |
| visual attention | **-0.407 (0.0217)** | **0.441 (0.0083)** | -0.146 (0.5014) | 0.082 (0.6803) | 0.070 (0.8915) | **0.338 (0.0489)** | **-0.352 (0.0258)** |
| visual perception | **-0.425 (0.0020)** | **0.436 (0.0025)** | -0.220 (0.2230) | **0.338 (0.0033)** | -0.097 (0.5024) | **0.455 (0.0012)** | **-0.446 (0.0012)** |
| word recognition | -0.060 (0.8305) | 0.065 (0.8213) | 0.088 (0.5641) | -0.032 (0.8289) | -0.028 (0.9390) | -0.025 (0.9425) | 0.018 (0.9586) |
| working memory | 0.024 (0.8564) | 0.037 (0.8289) | -0.028 (0.8234) | **-0.366 (0.0038)** | **0.375 (0.0012)** | -0.072 (0.7026) | 0.020 (0.9425) |

Notes. Cells show signed Spearman ρ (BH-FDR q); bold indicates q < 0.05. All 861 tests used 400 parcels and 10,000 hemisphere-preserving spatial spins with a two-sided null-mean-centered test. FDR correction precedes term selection and covers the complete 7 × 123 matrix. All coefficients, including nonsignificant associations, are shown. Terms retain the reference vocabulary and are ordered alphabetically. Full-precision coefficients and empirical P and adjusted q values are provided in Meta_CAP_Neurosynth_all123.csv.

Table S7. Sensitivity of joint GM–WM CAPs and their functional annotations to participant subsampling.

A. Cluster number, spatial matching and direction of functional associations.

| **Measure** | **10% sample** | **30% sample** | **50% sample** |
| --- | --- | --- | --- |
| Participants, n (MDD / NC) | 255 (141 / 114) | 765 (425 / 340) | 1,276 (710 / 566) |
| Depths selecting K = 7, n / 7 | 7/7 | 7/7 | 7/7 |
| Centroids closest to the same full-cohort state, n / 49 | 46/49 | 38/49 | 45/49 |
| Representative associations with the same sign, n / 49 | 44/49 | 46/49 | 46/49 |
| Same sign among full-cohort FDR-significant associations, n / 29 | 28/29 | 28/29 | 28/29 |

B. Spatial and functional similarity of each CAP to its full-cohort counterpart.

|  | **Centroid similarity, r** | | | **123-term profile similarity, ρ** | | |
| --- | --- | --- | --- | --- | --- | --- |
| **CAP** | **10%** | **30%** | **50%** | **10%** | **30%** | **50%** |
| CAP1 | 0.957 | 0.950 | 0.959 | 0.976 | 0.983 | 0.984 |
| CAP2 | 0.978 | 0.950 | 0.983 | 0.995 | 0.987 | 0.993 |
| CAP3 | 0.905 | 0.855 | 0.812 | 0.983 | 0.881 | 0.966 |
| CAP4 | 0.805 | 0.768 | 0.661 | 0.586 | 0.563 | 0.709 |
| CAP5 | 0.911 | 0.572 | 0.809 | 0.881 | 0.722 | 0.917 |
| CAP6 | 0.987 | 0.939 | 0.991 | 0.997 | 0.956 | 0.995 |
| CAP7 | 0.956 | 0.984 | 0.980 | 0.980 | 0.995 | 0.995 |

In panel A, the 49 centroid comparisons cover seven CAPs at seven depths. The 49 functional associations cover seven CAPs and the seven terms displayed in Figure 2. FDR significance refers to the full-cohort analysis (q < 0.05 across 861 CAP–term tests). In panel B, spatial similarity is the median signed Pearson correlation across the seven depths. Functional similarity is the Spearman correlation between the two complete 123-term profiles. Direction agreed in all 126 comparisons for CAP1–3 and CAP5–7 across the three samples. For CAP4, the correlations with autobiographical memory and social cognition changed from negative in the full cohort to weakly positive in each subsample.

Table S8. Regional receptor-model gain beyond spatial distance.

| **CAP** | **Baseline mean adjusted R²** | **Full model mean adjusted R²** | **Mean Δadjusted R²** | **Median gain [Q1, Q3]** | **Significant n / 400 (%)** |
| --- | --- | --- | --- | --- | --- |
| CAP-1 | 0.022 | 0.073 | 0.051 | 0.031 [0.004, 0.070] | 126 / 400 (31.5) |
| CAP-2 | 0.013 | 0.096 | 0.083 | 0.065 [0.015, 0.144] | 178 / 400 (44.5) |
| CAP-3 | 0.014 | 0.083 | 0.070 | 0.058 [0.014, 0.118] | 196 / 400 (49.0) |
| CAP-4 | 0.040 | 0.065 | 0.025 | 0.011 [0.001, 0.032] | 15 / 400 (3.8) |
| CAP-5 | 0.049 | 0.150 | 0.101 | 0.068 [0.016, 0.166] | 187 / 400 (46.8) |
| CAP-6 | 0.025 | 0.059 | 0.033 | 0.025 [0.004, 0.051] | 46 / 400 (11.5) |
| CAP-7 | 0.014 | 0.075 | 0.061 | 0.043 [0.011, 0.092] | 152 / 400 (38.0) |

Notes. Related to Figure 3 and Figure S13. For each CAP and each of 400 seed parcels, models explained shared GM–WM CAP depth-profile similarity to all 400 parcels using distance alone (baseline) or distance plus multireceptor fingerprint similarity (full model). Gain equals full-model minus baseline adjusted R². Means, medians and quartiles summarize all 400 seed parcels, including nonsignificant and negative gains. Significant parcels had positive gains and one-sided spatial-spin BH-FDR q < 0.05 across 2,800 CAP-by-parcel tests (10,000 permutations). Q1 and Q3 denote the 25th and 75th percentiles, not confidence bounds. Full parcel-level estimates, spin p values and corrected q values are supplied in Receptor_gain_all_parcels.csv.

Table S9. Significant regional gains of receptor-informed CAP depth-profile models.

| **CAP-1: 126 / 400 significant parcels (31.50%)** | | | | | | |
| --- | --- | --- | --- | --- | --- | --- |
| **ROI ID** | **Atlas parcel** | **Baseline adjusted R²** | **Full model adjusted R²** | **Gain (Δadj. R²)** | **Spin p** | **FDR q** |
| 240 | RH_SomMot_10 | 0.01236 | 0.34031 | 0.32795 | 0.00039996 | 0.00324605 |
| 106 | LH_SalVentAttn_PFCl_1 | 0.00471 | 0.28816 | 0.28345 | 0.00039996 | 0.00324605 |
| 259 | RH_SomMot_29 | 0.00448 | 0.26475 | 0.26027 | 0.00039996 | 0.00324605 |
| 342 | RH_Cont_PFCl_2 | 0.01479 | 0.25426 | 0.23947 | 0.00039996 | 0.00324605 |
| 59 | LH_SomMot_28 | −0.00048 | 0.23869 | 0.23917 | 0.00039996 | 0.00324605 |
| 363 | RH_Default_Par_2 | 0.00018 | 0.23786 | 0.23768 | 9.999e−5 | 0.0022762 |
| 53 | LH_SomMot_22 | 0.01222 | 0.24087 | 0.22865 | 9.999e−5 | 0.0022762 |
| 38 | LH_SomMot_7 | 0.02016 | 0.23750 | 0.21734 | 0.00039996 | 0.00324605 |
| 224 | RH_Vis_24 | 0.00123 | 0.21856 | 0.21734 | 9.999e−5 | 0.0022762 |
| 336 | RH_Cont_Par_5 | 0.00026 | 0.21437 | 0.21411 | 0.00039996 | 0.00324605 |
| 95 | LH_SalVentAttn_ParOper_4 | 0.00137 | 0.21452 | 0.21315 | 0.00049995 | 0.003535 |
| 103 | LH_SalVentAttn_FrOper_7 | −0.00216 | 0.21077 | 0.21294 | 0.00039996 | 0.00324605 |
| 255 | RH_SomMot_25 | −0.00230 | 0.20752 | 0.20982 | 0.00069993 | 0.00439418 |
| 93 | LH_SalVentAttn_ParOper_2 | 0.00980 | 0.21939 | 0.20959 | 9.999e−5 | 0.0022762 |
| 365 | RH_Default_Par_4 | 0.00042 | 0.20786 | 0.20743 | 0.00069993 | 0.00439418 |
| 252 | RH_SomMot_22 | 0.04703 | 0.24963 | 0.20260 | 0.00039996 | 0.00324605 |
| 51 | LH_SomMot_20 | 0.00613 | 0.20307 | 0.19694 | 0.00039996 | 0.00324605 |
| 304 | RH_SalVentAttn_FrOper_3 | 0.00127 | 0.19769 | 0.19642 | 0.00079992 | 0.00479609 |
| 343 | RH_Cont_PFCl_3 | 0.01714 | 0.20854 | 0.19140 | 0.00049995 | 0.003535 |
| 130 | LH_Cont_Par_4 | 0.00545 | 0.19506 | 0.18961 | 0.00109989 | 0.0059684 |
| 134 | LH_Cont_PFCl_1 | 0.00574 | 0.19445 | 0.18872 | 0.00039996 | 0.00324605 |
| 371 | RH_Default_Temp_5 | 0.00169 | 0.18642 | 0.18472 | 9.999e−5 | 0.0022762 |
| 151 | LH_Default_Temp_3 | −0.00213 | 0.18226 | 0.18438 | 9.999e−5 | 0.0022762 |
| 138 | LH_Cont_PFCl_5 | 0.00948 | 0.19290 | 0.18342 | 0.00089991 | 0.00521687 |
| 366 | RH_Default_Par_5 | −0.00221 | 0.17730 | 0.17951 | 0.00089991 | 0.00521687 |
| 270 | RH_SomMot_40 | 0.01199 | 0.18802 | 0.17603 | 0.00039996 | 0.00324605 |
| 333 | RH_Cont_Par_2 | 0.00850 | 0.17742 | 0.16892 | 0.00029997 | 0.00324605 |
| 132 | LH_Cont_Par_6 | −0.00251 | 0.16597 | 0.16848 | 0.00039996 | 0.00324605 |
| 280 | RH_DorsAttn_Post_10 | −0.00198 | 0.16552 | 0.16751 | 0.00029997 | 0.00324605 |
| 133 | LH_Cont_Temp_1 | 0.02778 | 0.19091 | 0.16313 | 9.999e−5 | 0.0022762 |
| 338 | RH_Cont_Temp_1 | −0.00083 | 0.16161 | 0.16244 | 0.00049995 | 0.003535 |
| 127 | LH_Cont_Par_1 | 0.00276 | 0.16037 | 0.15761 | 0.00029997 | 0.00324605 |
| 25 | LH_Vis_25 | 0.00364 | 0.15845 | 0.15480 | 0.00069993 | 0.00439418 |
| 344 | RH_Cont_PFCl_4 | 0.02304 | 0.17695 | 0.15391 | 0.00039996 | 0.00324605 |
| 161 | LH_Default_Temp_13 | 0.00849 | 0.16088 | 0.15239 | 0.00169983 | 0.00819195 |
| 248 | RH_SomMot_18 | 0.00639 | 0.14971 | 0.14332 | 0.00079992 | 0.00479609 |
| 341 | RH_Cont_PFCl_1 | 0.00366 | 0.14400 | 0.14034 | 9.999e−5 | 0.0022762 |
| 104 | LH_SalVentAttn_FrOper_8 | −0.00156 | 0.13722 | 0.13878 | 0.00039996 | 0.00324605 |
| 100 | LH_SalVentAttn_FrOper_4 | 0.00991 | 0.14808 | 0.13817 | 0.00119988 | 0.00631516 |
| 310 | RH_SalVentAttn_PFCl_1 | 0.03996 | 0.17659 | 0.13663 | 0.00039996 | 0.00324605 |
| 349 | RH_Cont_PFCl_9 | 0.02306 | 0.15555 | 0.13249 | 0.0019998 | 0.00944256 |
| 282 | RH_DorsAttn_Post_12 | −0.00229 | 0.12912 | 0.13142 | 0.00039996 | 0.00324605 |
| 187 | LH_Default_PFC_22 | 0.01042 | 0.14012 | 0.12971 | 0.00059994 | 0.00398064 |
| 308 | RH_SalVentAttn_FrOper_7 | 0.00363 | 0.13279 | 0.12916 | 9.999e−5 | 0.0022762 |
| 15 | LH_Vis_15 | 0.03009 | 0.15831 | 0.12822 | 0.00139986 | 0.00702439 |
| 294 | RH_SalVentAttn_TempOccPar_1 | −0.00247 | 0.12529 | 0.12776 | 0.00079992 | 0.00479609 |
| 299 | RH_SalVentAttn_TempOccPar_6 | 0.00992 | 0.13692 | 0.12700 | 0.0009999 | 0.00561066 |
| 63 | LH_SomMot_32 | 0.07029 | 0.19699 | 0.12670 | 0.00079992 | 0.00479609 |
| 351 | RH_Cont_PFCl_11 | 0.06213 | 0.18626 | 0.12412 | 9.999e−5 | 0.0022762 |
| 345 | RH_Cont_PFCl_5 | 0.03284 | 0.15577 | 0.12292 | 0.00019998 | 0.00324605 |
| 142 | LH_Cont_PFCl_9 | 0.01029 | 0.13259 | 0.12230 | 0.00039996 | 0.00324605 |
| 257 | RH_SomMot_27 | 0.03952 | 0.16181 | 0.12229 | 0.00079992 | 0.00479609 |
| 361 | RH_Cont_PFCmp_2 | 0.01970 | 0.14087 | 0.12117 | 0.00089991 | 0.00521687 |
| 285 | RH_DorsAttn_Post_15 | −0.00051 | 0.11933 | 0.11983 | 0.00019998 | 0.00324605 |
| 139 | LH_Cont_PFCl_6 | 0.00442 | 0.12398 | 0.11956 | 0.00089991 | 0.00521687 |
| 131 | LH_Cont_Par_5 | −0.00250 | 0.11407 | 0.11657 | 0.00039996 | 0.00324605 |
| 26 | LH_Vis_26 | 0.00301 | 0.11799 | 0.11498 | 0.00119988 | 0.00631516 |
| 278 | RH_DorsAttn_Post_8 | 0.03410 | 0.14751 | 0.11341 | 0.00049995 | 0.003535 |
| 251 | RH_SomMot_21 | 0.01122 | 0.12276 | 0.11154 | 0.00049995 | 0.003535 |
| 340 | RH_Cont_PFCv_1 | −0.00060 | 0.10952 | 0.11012 | 0.00019998 | 0.00324605 |
| 205 | RH_Vis_5 | 0.00246 | 0.11243 | 0.10997 | 0.00869913 | 0.0305616 |
| 90 | LH_DorsAttn_PrCv_1 | −0.00166 | 0.10780 | 0.10947 | 0.00509949 | 0.0199143 |
| 137 | LH_Cont_PFCl_4 | −0.00072 | 0.10792 | 0.10864 | 0.0009999 | 0.00561066 |
| 265 | RH_SomMot_35 | 0.02238 | 0.13090 | 0.10852 | 0.00029997 | 0.00324605 |
| 220 | RH_Vis_20 | 0.01771 | 0.12558 | 0.10787 | 0.00139986 | 0.00702439 |
| 62 | LH_SomMot_31 | 0.11319 | 0.22071 | 0.10753 | 0.00019998 | 0.00324605 |
| 60 | LH_SomMot_29 | 0.04737 | 0.15433 | 0.10696 | 0.00139986 | 0.00702439 |
| 12 | LH_Vis_12 | 0.02850 | 0.13448 | 0.10598 | 0.00369963 | 0.015554 |
| 188 | LH_Default_PFC_23 | 0.01072 | 0.11583 | 0.10511 | 0.00259974 | 0.0115728 |
| 74 | LH_DorsAttn_Post_6 | −0.00246 | 0.10234 | 0.10479 | 0.00349965 | 0.0149375 |
| 94 | LH_SalVentAttn_ParOper_3 | 0.00002 | 0.10470 | 0.10468 | 0.00249975 | 0.0112529 |
| 45 | LH_SomMot_14 | 0.00215 | 0.10591 | 0.10376 | 0.00469953 | 0.0186913 |
| 144 | LH_Cont_pCun_1 | 0.00176 | 0.10421 | 0.10245 | 0.00789921 | 0.0282475 |
| 339 | RH_Cont_Temp_2 | −0.00244 | 0.09905 | 0.10149 | 0.00249975 | 0.0112529 |
| 170 | LH_Default_PFC_5 | −0.00248 | 0.09676 | 0.09924 | 0.00239976 | 0.0109793 |
| 84 | LH_DorsAttn_Post_16 | 0.03058 | 0.12576 | 0.09518 | 0.00059994 | 0.00398064 |
| 176 | LH_Default_PFC_11 | 0.00133 | 0.09488 | 0.09355 | 0.0069993 | 0.0256855 |
| 6 | LH_Vis_6 | 0.05678 | 0.15018 | 0.09339 | 0.00459954 | 0.0183982 |
| 129 | LH_Cont_Par_3 | −0.00239 | 0.09066 | 0.09304 | 0.00179982 | 0.00859982 |
| 89 | LH_DorsAttn_FEF_4 | 0.05650 | 0.14869 | 0.09219 | 0.00069993 | 0.00439418 |
| 289 | RH_DorsAttn_Post_19 | 0.08419 | 0.17494 | 0.09074 | 0.00049995 | 0.003535 |
| 225 | RH_Vis_25 | 0.00081 | 0.09071 | 0.08990 | 0.00839916 | 0.0296566 |
| 335 | RH_Cont_Par_4 | 0.03737 | 0.12598 | 0.08862 | 0.00059994 | 0.00398064 |
| 337 | RH_Cont_Par_6 | 0.01889 | 0.10626 | 0.08737 | 0.00569943 | 0.0218309 |
| 354 | RH_Cont_PFCl_14 | 0.11622 | 0.20197 | 0.08575 | 0.00019998 | 0.00324605 |
| 296 | RH_SalVentAttn_TempOccPar_3 | 0.01272 | 0.09535 | 0.08263 | 0.00059994 | 0.00398064 |
| 228 | RH_Vis_28 | −0.00250 | 0.07737 | 0.07987 | 0.0153985 | 0.048174 |
| 83 | LH_DorsAttn_Post_15 | 0.00051 | 0.07978 | 0.07927 | 0.0116988 | 0.0388113 |
| 36 | LH_SomMot_5 | −0.00110 | 0.07780 | 0.07890 | 0.00069993 | 0.00439418 |
| 28 | LH_Vis_28 | 0.01002 | 0.08789 | 0.07787 | 0.00819918 | 0.0290604 |
| 48 | LH_SomMot_17 | 0.01201 | 0.08766 | 0.07564 | 0.00709929 | 0.0259504 |
| 390 | RH_Default_PFCm_12 | 0.02014 | 0.09549 | 0.07535 | 0.00319968 | 0.0138471 |
| 148 | LH_Cont_PFCmp_1 | 0.02407 | 0.09829 | 0.07422 | 0.00969903 | 0.0333628 |
| 80 | LH_DorsAttn_Post_12 | 0.02471 | 0.09788 | 0.07317 | 0.00039996 | 0.00324605 |
| 50 | LH_SomMot_19 | −0.00247 | 0.07040 | 0.07288 | 0.00369963 | 0.015554 |
| 186 | LH_Default_PFC_21 | 0.00597 | 0.07647 | 0.07051 | 0.0111989 | 0.0375082 |
| 246 | RH_SomMot_16 | −0.00116 | 0.06853 | 0.06969 | 0.00889911 | 0.0311469 |
| 64 | LH_SomMot_33 | 0.12458 | 0.19417 | 0.06959 | 0.00069993 | 0.00439418 |
| 4 | LH_Vis_4 | 0.03574 | 0.10516 | 0.06942 | 0.010299 | 0.0351672 |
| 297 | RH_SalVentAttn_TempOccPar_4 | 0.01717 | 0.08621 | 0.06904 | 0.00959904 | 0.0331001 |
| 196 | LH_Default_PCC_7 | −0.00166 | 0.06652 | 0.06817 | 0.00819918 | 0.0290604 |
| 332 | RH_Cont_Par_1 | 0.01731 | 0.08373 | 0.06642 | 0.00249975 | 0.0112529 |
| 68 | LH_SomMot_37 | 0.10495 | 0.16994 | 0.06498 | 0.00089991 | 0.00521687 |
| 165 | LH_Default_Temp_17 | −0.00249 | 0.06240 | 0.06489 | 0.00569943 | 0.0218309 |
| 381 | RH_Default_PFCm_3 | −0.00216 | 0.06234 | 0.06450 | 0.0127987 | 0.0413815 |
| 97 | LH_SalVentAttn_FrOper_1 | 0.00937 | 0.07270 | 0.06333 | 0.0127987 | 0.0413815 |
| 277 | RH_DorsAttn_Post_7 | 0.01209 | 0.07538 | 0.06330 | 0.00119988 | 0.00631516 |
| 57 | LH_SomMot_26 | 0.06157 | 0.12482 | 0.06324 | 0.00179982 | 0.00859982 |
| 263 | RH_SomMot_33 | 0.12996 | 0.19244 | 0.06248 | 0.00109989 | 0.0059684 |
| 394 | RH_Default_PCC_3 | 0.00132 | 0.05946 | 0.05814 | 0.00589941 | 0.022566 |
| 206 | RH_Vis_6 | 0.01783 | 0.07561 | 0.05779 | 0.0115988 | 0.038571 |
| 254 | RH_SomMot_24 | 0.07646 | 0.13337 | 0.05691 | 0.0039996 | 0.0164448 |
| 353 | RH_Cont_PFCl_13 | 0.07942 | 0.13583 | 0.05640 | 0.00489951 | 0.0193493 |
| 260 | RH_SomMot_30 | 0.05778 | 0.11217 | 0.05439 | 0.0105989 | 0.0358418 |
| 276 | RH_DorsAttn_Post_6 | 0.05602 | 0.11036 | 0.05434 | 0.00379962 | 0.0158553 |
| 256 | RH_SomMot_26 | 0.08985 | 0.14405 | 0.05420 | 0.00169983 | 0.00819195 |
| 262 | RH_SomMot_32 | 0.09530 | 0.14888 | 0.05359 | 0.00839916 | 0.0296566 |
| 269 | RH_SomMot_39 | 0.11568 | 0.16829 | 0.05261 | 0.00159984 | 0.00783138 |
| 298 | RH_SalVentAttn_TempOccPar_5 | 0.00916 | 0.06107 | 0.05192 | 0.010299 | 0.0351672 |
| 239 | RH_SomMot_9 | 0.02772 | 0.07754 | 0.04982 | 0.00349965 | 0.0149375 |
| 373 | RH_Default_Temp_7 | 0.00206 | 0.04740 | 0.04535 | 0.0142986 | 0.0454438 |
| 67 | LH_SomMot_36 | 0.03128 | 0.07601 | 0.04473 | 0.0113989 | 0.0379962 |
| 154 | LH_Default_Temp_6 | 0.00263 | 0.04615 | 0.04352 | 0.0153985 | 0.048174 |
| 268 | RH_SomMot_38 | 0.13047 | 0.17369 | 0.04322 | 0.0127987 | 0.0413815 |
| 85 | LH_DorsAttn_Post_17 | 0.01167 | 0.05307 | 0.04140 | 0.010499 | 0.0356328 |
| 283 | RH_DorsAttn_Post_13 | 0.01662 | 0.05623 | 0.03961 | 0.00469953 | 0.0186913 |

Table S9. Continued. B. Significant regional gains of receptor-informed CAP depth-profile models.

| **CAP-2: 178 / 400 significant parcels (44.50%)** | | | | | | |
| --- | --- | --- | --- | --- | --- | --- |
| **ROI ID** | **Atlas parcel** | **Baseline adjusted R²** | **Full model adjusted R²** | **Gain (Δadj. R²)** | **Spin p** | **FDR q** |
| 240 | RH_SomMot_10 | 0.01591 | 0.34311 | 0.32720 | 0.00039996 | 0.00324605 |
| 371 | RH_Default_Temp_5 | −0.00186 | 0.29342 | 0.29528 | 9.999e−5 | 0.0022762 |
| 162 | LH_Default_Temp_14 | 0.01598 | 0.28769 | 0.27171 | 0.00049995 | 0.003535 |
| 103 | LH_SalVentAttn_FrOper_7 | −0.00119 | 0.26490 | 0.26609 | 0.00039996 | 0.00324605 |
| 59 | LH_SomMot_28 | 0.00487 | 0.26643 | 0.26156 | 0.00039996 | 0.00324605 |
| 163 | LH_Default_Temp_15 | 0.01033 | 0.26785 | 0.25752 | 9.999e−5 | 0.0022762 |
| 259 | RH_SomMot_29 | −0.00228 | 0.25258 | 0.25486 | 0.00059994 | 0.00398064 |
| 270 | RH_SomMot_40 | −0.00213 | 0.24673 | 0.24886 | 9.999e−5 | 0.0022762 |
| 236 | RH_SomMot_6 | 0.00106 | 0.24955 | 0.24850 | 0.00049995 | 0.003535 |
| 365 | RH_Default_Par_4 | −0.00251 | 0.24469 | 0.24720 | 0.00069993 | 0.00439418 |
| 64 | LH_SomMot_33 | 0.00876 | 0.25352 | 0.24476 | 9.999e−5 | 0.0022762 |
| 373 | RH_Default_Temp_7 | −0.00142 | 0.24315 | 0.24456 | 9.999e−5 | 0.0022762 |
| 62 | LH_SomMot_31 | 0.02974 | 0.27411 | 0.24436 | 0.00039996 | 0.00324605 |
| 151 | LH_Default_Temp_3 | 0.01384 | 0.25479 | 0.24095 | 0.00039996 | 0.00324605 |
| 364 | RH_Default_Par_3 | −0.00105 | 0.23490 | 0.23595 | 0.00049995 | 0.003535 |
| 278 | RH_DorsAttn_Post_8 | −0.00206 | 0.22156 | 0.22362 | 0.00049995 | 0.003535 |
| 234 | RH_SomMot_4 | −0.00184 | 0.22161 | 0.22345 | 0.00029997 | 0.00324605 |
| 252 | RH_SomMot_22 | 0.00928 | 0.23099 | 0.22172 | 0.00039996 | 0.00324605 |
| 134 | LH_Cont_PFCl_1 | 0.01121 | 0.23200 | 0.22078 | 0.00049995 | 0.003535 |
| 341 | RH_Cont_PFCl_1 | 0.01717 | 0.23784 | 0.22067 | 0.00029997 | 0.00324605 |
| 263 | RH_SomMot_33 | 0.00801 | 0.22762 | 0.21961 | 9.999e−5 | 0.0022762 |
| 93 | LH_SalVentAttn_ParOper_2 | 0.00481 | 0.22133 | 0.21652 | 0.00049995 | 0.003535 |
| 363 | RH_Default_Par_2 | 0.00062 | 0.21650 | 0.21588 | 0.00079992 | 0.00479609 |
| 384 | RH_Default_PFCm_6 | −0.00170 | 0.21313 | 0.21483 | 0.00069993 | 0.00439418 |
| 51 | LH_SomMot_20 | 0.00151 | 0.21491 | 0.21340 | 9.999e−5 | 0.0022762 |
| 262 | RH_SomMot_32 | 0.00551 | 0.21516 | 0.20965 | 0.00049995 | 0.003535 |
| 106 | LH_SalVentAttn_PFCl_1 | 0.01405 | 0.22349 | 0.20945 | 0.00149985 | 0.00740667 |
| 138 | LH_Cont_PFCl_5 | 0.01995 | 0.22937 | 0.20942 | 0.00139986 | 0.00702439 |
| 276 | RH_DorsAttn_Post_6 | 0.00499 | 0.21350 | 0.20851 | 0.00019998 | 0.00324605 |
| 254 | RH_SomMot_24 | −0.00238 | 0.20462 | 0.20701 | 9.999e−5 | 0.0022762 |
| 165 | LH_Default_Temp_17 | 0.00364 | 0.20973 | 0.20609 | 0.00079992 | 0.00479609 |
| 132 | LH_Cont_Par_6 | −0.00161 | 0.20252 | 0.20414 | 0.00079992 | 0.00479609 |
| 63 | LH_SomMot_32 | 0.00516 | 0.20920 | 0.20404 | 0.00069993 | 0.00439418 |
| 131 | LH_Cont_Par_5 | 0.00146 | 0.20535 | 0.20390 | 0.0009999 | 0.00561066 |
| 53 | LH_SomMot_22 | 0.00653 | 0.20988 | 0.20335 | 0.00059994 | 0.00398064 |
| 342 | RH_Cont_PFCl_2 | 0.01946 | 0.22048 | 0.20102 | 0.00069993 | 0.00439418 |
| 187 | LH_Default_PFC_22 | 0.00520 | 0.20365 | 0.19845 | 0.00119988 | 0.00631516 |
| 71 | LH_DorsAttn_Post_3 | 0.01599 | 0.21322 | 0.19723 | 0.00039996 | 0.00324605 |
| 164 | LH_Default_Temp_16 | 0.01542 | 0.21204 | 0.19662 | 0.00019998 | 0.00324605 |
| 133 | LH_Cont_Temp_1 | 0.02298 | 0.21876 | 0.19578 | 0.00019998 | 0.00324605 |
| 38 | LH_SomMot_7 | 0.03558 | 0.22926 | 0.19368 | 0.00039996 | 0.00324605 |
| 171 | LH_Default_PFC_6 | 0.01346 | 0.20527 | 0.19180 | 0.00169983 | 0.00819195 |
| 255 | RH_SomMot_25 | −0.00241 | 0.18824 | 0.19065 | 0.00039996 | 0.00324605 |
| 233 | RH_SomMot_3 | −0.00150 | 0.18860 | 0.19010 | 0.00019998 | 0.00324605 |
| 304 | RH_SalVentAttn_FrOper_3 | −0.00209 | 0.18723 | 0.18932 | 0.00249975 | 0.0112529 |
| 336 | RH_Cont_Par_5 | −0.00229 | 0.18645 | 0.18874 | 0.0009999 | 0.00561066 |
| 354 | RH_Cont_PFCl_14 | 0.02884 | 0.21505 | 0.18620 | 0.00159984 | 0.00783138 |
| 68 | LH_SomMot_37 | 0.00679 | 0.18923 | 0.18244 | 0.00059994 | 0.00398064 |
| 118 | LH_Limbic_OFC_5 | 0.01490 | 0.19533 | 0.18043 | 0.00039996 | 0.00324605 |
| 116 | LH_Limbic_OFC_3 | 0.01621 | 0.19601 | 0.17980 | 0.00069993 | 0.00439418 |
| 343 | RH_Cont_PFCl_3 | 0.02211 | 0.20119 | 0.17908 | 0.00079992 | 0.00479609 |
| 95 | LH_SalVentAttn_ParOper_4 | −0.00054 | 0.17826 | 0.17879 | 0.00139986 | 0.00702439 |
| 49 | LH_SomMot_18 | 0.00222 | 0.18081 | 0.17858 | 0.00039996 | 0.00324605 |
| 359 | RH_Cont_Cing_2 | 0.03832 | 0.21673 | 0.17841 | 9.999e−5 | 0.0022762 |
| 390 | RH_Default_PFCm_12 | 0.01832 | 0.19671 | 0.17839 | 0.00079992 | 0.00479609 |
| 251 | RH_SomMot_21 | −0.00039 | 0.17689 | 0.17728 | 0.00029997 | 0.00324605 |
| 296 | RH_SalVentAttn_TempOccPar_3 | −0.00248 | 0.17282 | 0.17530 | 0.00109989 | 0.0059684 |
| 127 | LH_Cont_Par_1 | 0.01021 | 0.18464 | 0.17443 | 0.00169983 | 0.00819195 |
| 130 | LH_Cont_Par_4 | 0.00345 | 0.17777 | 0.17432 | 0.00159984 | 0.00783138 |
| 268 | RH_SomMot_38 | 0.00754 | 0.18140 | 0.17387 | 0.00019998 | 0.00324605 |
| 137 | LH_Cont_PFCl_4 | 0.01412 | 0.18768 | 0.17355 | 0.00119988 | 0.00631516 |
| 257 | RH_SomMot_27 | −0.00193 | 0.17095 | 0.17288 | 0.00069993 | 0.00439418 |
| 256 | RH_SomMot_26 | −0.00166 | 0.16956 | 0.17123 | 0.00059994 | 0.00398064 |
| 348 | RH_Cont_PFCl_8 | 0.01478 | 0.18570 | 0.17092 | 0.00129987 | 0.00672761 |
| 366 | RH_Default_Par_5 | −0.00101 | 0.16980 | 0.17080 | 0.00119988 | 0.00631516 |
| 362 | RH_Default_Par_1 | −0.00124 | 0.16937 | 0.17061 | 0.00039996 | 0.00324605 |
| 337 | RH_Cont_Par_6 | −0.00234 | 0.16762 | 0.16996 | 0.00149985 | 0.00740667 |
| 388 | RH_Default_PFCm_10 | 0.02873 | 0.19855 | 0.16982 | 0.00089991 | 0.00521687 |
| 351 | RH_Cont_PFCl_11 | 0.02472 | 0.19403 | 0.16931 | 0.00139986 | 0.00702439 |
| 297 | RH_SalVentAttn_TempOccPar_4 | −0.00229 | 0.16692 | 0.16921 | 0.00109989 | 0.0059684 |
| 340 | RH_Cont_PFCv_1 | 0.00501 | 0.17279 | 0.16778 | 0.00079992 | 0.00479609 |
| 308 | RH_SalVentAttn_FrOper_7 | −0.00239 | 0.16532 | 0.16771 | 9.999e−5 | 0.0022762 |
| 181 | LH_Default_PFC_16 | 0.03630 | 0.20290 | 0.16660 | 0.00129987 | 0.00672761 |
| 310 | RH_SalVentAttn_PFCl_1 | 0.02607 | 0.19184 | 0.16577 | 0.00109989 | 0.0059684 |
| 80 | LH_DorsAttn_Post_12 | 0.01103 | 0.17581 | 0.16478 | 9.999e−5 | 0.0022762 |
| 128 | LH_Cont_Par_2 | −0.00026 | 0.16422 | 0.16449 | 0.00109989 | 0.0059684 |
| 50 | LH_SomMot_19 | 0.04390 | 0.20629 | 0.16239 | 9.999e−5 | 0.0022762 |
| 60 | LH_SomMot_29 | 0.00299 | 0.16454 | 0.16155 | 0.00049995 | 0.003535 |
| 105 | LH_SalVentAttn_FrOper_9 | 0.00112 | 0.15992 | 0.15880 | 0.00139986 | 0.00702439 |
| 142 | LH_Cont_PFCl_9 | 0.01492 | 0.17349 | 0.15858 | 0.00139986 | 0.00702439 |
| 312 | RH_SalVentAttn_Med_2 | 0.00574 | 0.16400 | 0.15827 | 0.00039996 | 0.00324605 |
| 248 | RH_SomMot_18 | −0.00180 | 0.15623 | 0.15803 | 0.00059994 | 0.00398064 |
| 148 | LH_Cont_PFCmp_1 | −0.00088 | 0.15562 | 0.15649 | 0.00069993 | 0.00439418 |
| 368 | RH_Default_Temp_2 | −0.00235 | 0.15401 | 0.15635 | 0.0009999 | 0.00561066 |
| 129 | LH_Cont_Par_3 | −0.00139 | 0.15393 | 0.15531 | 0.00189981 | 0.00904671 |
| 45 | LH_SomMot_14 | −0.00130 | 0.15270 | 0.15400 | 0.00059994 | 0.00398064 |
| 161 | LH_Default_Temp_13 | 0.01349 | 0.16735 | 0.15386 | 0.00249975 | 0.0112529 |
| 352 | RH_Cont_PFCl_12 | 0.03577 | 0.18960 | 0.15383 | 0.00089991 | 0.00521687 |
| 339 | RH_Cont_Temp_2 | 0.00127 | 0.15457 | 0.15330 | 0.00059994 | 0.00398064 |
| 387 | RH_Default_PFCm_9 | 0.00283 | 0.15415 | 0.15132 | 9.999e−5 | 0.0022762 |
| 104 | LH_SalVentAttn_FrOper_8 | −0.00200 | 0.14906 | 0.15106 | 9.999e−5 | 0.0022762 |
| 280 | RH_DorsAttn_Post_10 | −0.00074 | 0.14917 | 0.14991 | 0.00139986 | 0.00702439 |
| 338 | RH_Cont_Temp_1 | −0.00031 | 0.14844 | 0.14875 | 0.00059994 | 0.00398064 |
| 108 | LH_SalVentAttn_Med_2 | 0.00304 | 0.15022 | 0.14718 | 0.00049995 | 0.003535 |
| 265 | RH_SomMot_35 | −0.00246 | 0.14456 | 0.14702 | 0.00119988 | 0.00631516 |
| 273 | RH_DorsAttn_Post_3 | 0.00032 | 0.14697 | 0.14665 | 0.00039996 | 0.00324605 |
| 333 | RH_Cont_Par_2 | −0.00210 | 0.14385 | 0.14595 | 0.00089991 | 0.00521687 |
| 321 | RH_Limbic_OFC_3 | 0.01299 | 0.15858 | 0.14558 | 0.00049995 | 0.003535 |
| 332 | RH_Cont_Par_1 | −0.00251 | 0.14195 | 0.14446 | 0.00079992 | 0.00479609 |
| 345 | RH_Cont_PFCl_5 | 0.02479 | 0.16900 | 0.14421 | 0.00229977 | 0.010626 |
| 56 | LH_SomMot_25 | 0.00081 | 0.14474 | 0.14393 | 0.00069993 | 0.00439418 |
| 355 | RH_Cont_PFCl_15 | 0.02914 | 0.17238 | 0.14323 | 0.00069993 | 0.00439418 |
| 100 | LH_SalVentAttn_FrOper_4 | 0.00367 | 0.14645 | 0.14278 | 0.00259974 | 0.0115728 |
| 382 | RH_Default_PFCm_4 | 0.01661 | 0.15937 | 0.14276 | 0.0009999 | 0.00561066 |
| 89 | LH_DorsAttn_FEF_4 | 0.00998 | 0.14836 | 0.13838 | 0.00139986 | 0.00702439 |
| 122 | LH_Limbic_TempPole_4 | 0.00481 | 0.14318 | 0.13837 | 0.00269973 | 0.0119231 |
| 48 | LH_SomMot_17 | −0.00136 | 0.13612 | 0.13748 | 0.00289971 | 0.0126664 |
| 250 | RH_SomMot_20 | 0.01295 | 0.14933 | 0.13639 | 0.00049995 | 0.003535 |
| 253 | RH_SomMot_23 | 0.00193 | 0.13776 | 0.13582 | 0.00109989 | 0.0059684 |
| 135 | LH_Cont_PFCl_2 | 0.00914 | 0.14434 | 0.13520 | 0.00149985 | 0.00740667 |
| 334 | RH_Cont_Par_3 | −0.00216 | 0.13287 | 0.13502 | 0.00219978 | 0.0102828 |
| 303 | RH_SalVentAttn_FrOper_2 | 0.00516 | 0.13571 | 0.13055 | 0.00959904 | 0.0331001 |
| 344 | RH_Cont_PFCl_4 | 0.01891 | 0.14945 | 0.13054 | 0.00209979 | 0.00986479 |
| 335 | RH_Cont_Par_4 | −0.00015 | 0.13010 | 0.13025 | 0.00139986 | 0.00702439 |
| 302 | RH_SalVentAttn_FrOper_1 | −0.00144 | 0.12845 | 0.12989 | 0.0069993 | 0.0256855 |
| 294 | RH_SalVentAttn_TempOccPar_1 | −0.00121 | 0.12860 | 0.12980 | 0.00109989 | 0.0059684 |
| 28 | LH_Vis_28 | 0.01946 | 0.14842 | 0.12896 | 0.00339966 | 0.0146673 |
| 12 | LH_Vis_12 | 0.05975 | 0.18869 | 0.12894 | 0.0069993 | 0.0256855 |
| 183 | LH_Default_PFC_18 | 0.03580 | 0.16428 | 0.12848 | 0.00049995 | 0.003535 |
| 269 | RH_SomMot_39 | 0.00599 | 0.13444 | 0.12845 | 0.0009999 | 0.00561066 |
| 285 | RH_DorsAttn_Post_15 | −0.00085 | 0.12750 | 0.12836 | 0.00209979 | 0.00986479 |
| 57 | LH_SomMot_26 | 0.00402 | 0.13172 | 0.12769 | 0.00069993 | 0.00439418 |
| 146 | LH_Cont_Cing_1 | 0.03329 | 0.15947 | 0.12618 | 0.00039996 | 0.00324605 |
| 186 | LH_Default_PFC_21 | 0.01617 | 0.14134 | 0.12517 | 0.00369963 | 0.015554 |
| 32 | LH_SomMot_1 | 0.00141 | 0.12481 | 0.12340 | 0.00319968 | 0.0138471 |
| 224 | RH_Vis_24 | 0.03754 | 0.16026 | 0.12272 | 0.00219978 | 0.0102828 |
| 361 | RH_Cont_PFCmp_2 | 0.00348 | 0.12527 | 0.12179 | 0.00039996 | 0.00324605 |
| 179 | LH_Default_PFC_14 | 0.01890 | 0.13930 | 0.12040 | 0.00569943 | 0.0218309 |
| 74 | LH_DorsAttn_Post_6 | −0.00233 | 0.11648 | 0.11882 | 0.00639936 | 0.024019 |
| 92 | LH_SalVentAttn_ParOper_1 | 0.00028 | 0.11889 | 0.11862 | 0.00459954 | 0.0183982 |
| 272 | RH_DorsAttn_Post_2 | 0.00282 | 0.12082 | 0.11800 | 0.00149985 | 0.00740667 |
| 282 | RH_DorsAttn_Post_12 | −0.00072 | 0.11711 | 0.11783 | 0.00279972 | 0.0123452 |
| 309 | RH_SalVentAttn_FrOper_8 | 0.02138 | 0.13459 | 0.11321 | 0.00389961 | 0.0161762 |
| 170 | LH_Default_PFC_5 | 0.00260 | 0.11253 | 0.10993 | 0.00369963 | 0.015554 |
| 139 | LH_Cont_PFCl_6 | 0.01418 | 0.12377 | 0.10959 | 0.00629937 | 0.0238033 |
| 109 | LH_SalVentAttn_Med_3 | 0.04169 | 0.15045 | 0.10876 | 0.00079992 | 0.00479609 |
| 349 | RH_Cont_PFCl_9 | 0.01942 | 0.12744 | 0.10802 | 0.0105989 | 0.0358418 |
| 26 | LH_Vis_26 | 0.04075 | 0.14797 | 0.10722 | 0.00749925 | 0.0270591 |
| 36 | LH_SomMot_5 | 0.01255 | 0.11919 | 0.10664 | 0.00039996 | 0.00324605 |
| 400 | RH_Default_PCC_9 | 0.00143 | 0.10736 | 0.10593 | 0.00369963 | 0.015554 |
| 235 | RH_SomMot_5 | −0.00209 | 0.10122 | 0.10331 | 0.0049995 | 0.0196059 |
| 214 | RH_Vis_14 | 0.03479 | 0.13731 | 0.10252 | 0.00379962 | 0.0158553 |
| 357 | RH_Cont_pCun_2 | 0.00335 | 0.10509 | 0.10174 | 0.00459954 | 0.0183982 |
| 353 | RH_Cont_PFCl_13 | 0.00349 | 0.10486 | 0.10137 | 0.0069993 | 0.0256855 |
| 188 | LH_Default_PFC_23 | 0.01381 | 0.11317 | 0.09936 | 0.00729927 | 0.0265084 |
| 144 | LH_Cont_pCun_1 | 0.00753 | 0.10657 | 0.09904 | 0.0118988 | 0.0392885 |
| 94 | LH_SalVentAttn_ParOper_3 | −0.00194 | 0.09568 | 0.09762 | 0.00789921 | 0.0282475 |
| 298 | RH_SalVentAttn_TempOccPar_5 | −0.00176 | 0.09550 | 0.09726 | 0.00429957 | 0.017347 |
| 67 | LH_SomMot_36 | −0.00229 | 0.09490 | 0.09719 | 0.00919908 | 0.0319968 |
| 136 | LH_Cont_PFCl_3 | 0.00952 | 0.10658 | 0.09706 | 0.0130987 | 0.0421083 |
| 277 | RH_DorsAttn_Post_7 | 0.01295 | 0.10884 | 0.09589 | 0.00069993 | 0.00439418 |
| 88 | LH_DorsAttn_FEF_3 | 0.09156 | 0.18740 | 0.09584 | 0.00049995 | 0.003535 |
| 299 | RH_SalVentAttn_TempOccPar_6 | 0.00011 | 0.09387 | 0.09375 | 0.00749925 | 0.0270591 |
| 97 | LH_SalVentAttn_FrOper_1 | 0.00258 | 0.09514 | 0.09256 | 0.0153985 | 0.048174 |
| 300 | RH_SalVentAttn_TempOccPar_7 | −0.00076 | 0.09155 | 0.09231 | 0.00819918 | 0.0290604 |
| 288 | RH_DorsAttn_Post_18 | −0.00217 | 0.08985 | 0.09202 | 0.010399 | 0.0353363 |
| 159 | LH_Default_Temp_11 | −0.00047 | 0.09121 | 0.09168 | 0.00019998 | 0.00324605 |
| 209 | RH_Vis_9 | −0.00235 | 0.08927 | 0.09162 | 0.00039996 | 0.00324605 |
| 274 | RH_DorsAttn_Post_4 | 0.00059 | 0.09150 | 0.09092 | 0.00529947 | 0.0205805 |
| 84 | LH_DorsAttn_Post_16 | 0.00064 | 0.09085 | 0.09021 | 0.00509949 | 0.0199143 |
| 145 | LH_Cont_pCun_2 | −0.00193 | 0.08494 | 0.08686 | 0.00569943 | 0.0218309 |
| 35 | LH_SomMot_4 | −0.00194 | 0.08481 | 0.08675 | 0.00659934 | 0.024572 |
| 154 | LH_Default_Temp_6 | 0.01033 | 0.09433 | 0.08400 | 0.00659934 | 0.024572 |
| 157 | LH_Default_Temp_9 | 0.00105 | 0.08429 | 0.08324 | 0.00029997 | 0.00324605 |
| 185 | LH_Default_PFC_20 | −0.00146 | 0.07904 | 0.08050 | 0.0147985 | 0.0469262 |
| 182 | LH_Default_PFC_17 | 0.00445 | 0.08491 | 0.08046 | 0.0150985 | 0.0476079 |
| 167 | LH_Default_PFC_2 | 0.00461 | 0.08383 | 0.07922 | 0.0127987 | 0.0413815 |
| 307 | RH_SalVentAttn_FrOper_6 | 0.00139 | 0.07950 | 0.07811 | 0.0089991 | 0.0313792 |
| 231 | RH_SomMot_1 | −0.00158 | 0.07634 | 0.07791 | 0.0107989 | 0.0364301 |
| 377 | RH_Default_PFCv_3 | 0.00066 | 0.07285 | 0.07218 | 0.00479952 | 0.019008 |
| 330 | RH_Limbic_TempPole_6 | 0.00007 | 0.06820 | 0.06813 | 9.999e−5 | 0.0022762 |
| 289 | RH_DorsAttn_Post_19 | 0.01834 | 0.08501 | 0.06667 | 0.00709929 | 0.0259504 |
| 239 | RH_SomMot_9 | 0.00069 | 0.06585 | 0.06517 | 0.00039996 | 0.00324605 |
| 20 | LH_Vis_20 | 0.06539 | 0.12656 | 0.06117 | 0.0132987 | 0.0427022 |
| 34 | LH_SomMot_3 | −0.00125 | 0.05483 | 0.05608 | 0.00119988 | 0.00631516 |
| 260 | RH_SomMot_30 | 0.02792 | 0.08263 | 0.05472 | 0.00139986 | 0.00702439 |
| 395 | RH_Default_PCC_4 | 0.00241 | 0.04256 | 0.04016 | 0.00319968 | 0.0138471 |
| 196 | LH_Default_PCC_7 | 0.02444 | 0.06417 | 0.03974 | 0.00559944 | 0.0216254 |

Table S9. Continued. C. Significant regional gains of receptor-informed CAP depth-profile models.

| **CAP-3: 196 / 400 significant parcels (49.00%)** | | | | | | |
| --- | --- | --- | --- | --- | --- | --- |
| **ROI ID** | **Atlas parcel** | **Baseline adjusted R²** | **Full model adjusted R²** | **Gain (Δadj. R²)** | **Spin p** | **FDR q** |
| 55 | LH_SomMot_24 | 0.01585 | 0.28130 | 0.26545 | 9.999e−5 | 0.0022762 |
| 254 | RH_SomMot_24 | 0.04069 | 0.30556 | 0.26487 | 0.00039996 | 0.00324605 |
| 270 | RH_SomMot_40 | 0.01649 | 0.27270 | 0.25621 | 0.00039996 | 0.00324605 |
| 260 | RH_SomMot_30 | 0.01950 | 0.25919 | 0.23969 | 0.00039996 | 0.00324605 |
| 259 | RH_SomMot_29 | 0.01933 | 0.23961 | 0.22028 | 0.00039996 | 0.00324605 |
| 49 | LH_SomMot_18 | 0.01940 | 0.23923 | 0.21983 | 0.00039996 | 0.00324605 |
| 264 | RH_SomMot_34 | 0.01867 | 0.23536 | 0.21669 | 0.00039996 | 0.00324605 |
| 64 | LH_SomMot_33 | 0.00697 | 0.21859 | 0.21162 | 9.999e−5 | 0.0022762 |
| 365 | RH_Default_Par_4 | 0.00417 | 0.21176 | 0.20759 | 0.00039996 | 0.00324605 |
| 106 | LH_SalVentAttn_PFCl_1 | 0.02195 | 0.22096 | 0.19900 | 0.00039996 | 0.00324605 |
| 354 | RH_Cont_PFCl_14 | 0.05937 | 0.25750 | 0.19813 | 0.00039996 | 0.00324605 |
| 262 | RH_SomMot_32 | 0.00820 | 0.20388 | 0.19568 | 9.999e−5 | 0.0022762 |
| 66 | LH_SomMot_35 | 0.00653 | 0.20193 | 0.19540 | 9.999e−5 | 0.0022762 |
| 171 | LH_Default_PFC_6 | 0.00883 | 0.20387 | 0.19504 | 0.00029997 | 0.00324605 |
| 146 | LH_Cont_Cing_1 | 0.03863 | 0.23175 | 0.19312 | 0.00039996 | 0.00324605 |
| 341 | RH_Cont_PFCl_1 | 0.01189 | 0.20294 | 0.19104 | 0.00039996 | 0.00324605 |
| 233 | RH_SomMot_3 | −0.00070 | 0.18643 | 0.18713 | 0.00049995 | 0.003535 |
| 187 | LH_Default_PFC_22 | 0.02231 | 0.20779 | 0.18548 | 0.00039996 | 0.00324605 |
| 63 | LH_SomMot_32 | 0.00328 | 0.18730 | 0.18402 | 9.999e−5 | 0.0022762 |
| 59 | LH_SomMot_28 | 0.00035 | 0.18121 | 0.18086 | 0.00039996 | 0.00324605 |
| 255 | RH_SomMot_25 | 0.02707 | 0.20668 | 0.17961 | 9.999e−5 | 0.0022762 |
| 321 | RH_Limbic_OFC_3 | −0.00238 | 0.17686 | 0.17924 | 9.999e−5 | 0.0022762 |
| 265 | RH_SomMot_35 | 0.01624 | 0.19363 | 0.17739 | 0.00029997 | 0.00324605 |
| 56 | LH_SomMot_25 | 0.02803 | 0.20329 | 0.17526 | 0.00039996 | 0.00324605 |
| 251 | RH_SomMot_21 | 0.02503 | 0.19819 | 0.17316 | 9.999e−5 | 0.0022762 |
| 257 | RH_SomMot_27 | 0.02134 | 0.19351 | 0.17217 | 0.00049995 | 0.003535 |
| 108 | LH_SalVentAttn_Med_2 | 0.03548 | 0.20687 | 0.17138 | 0.00039996 | 0.00324605 |
| 278 | RH_DorsAttn_Post_8 | 0.01383 | 0.18423 | 0.17040 | 0.00039996 | 0.00324605 |
| 168 | LH_Default_PFC_3 | 0.00317 | 0.16920 | 0.16603 | 0.00089991 | 0.00521687 |
| 316 | RH_SalVentAttn_Med_6 | 0.02757 | 0.19358 | 0.16601 | 9.999e−5 | 0.0022762 |
| 343 | RH_Cont_PFCl_3 | 0.01728 | 0.18274 | 0.16545 | 0.00039996 | 0.00324605 |
| 389 | RH_Default_PFCm_11 | 0.04992 | 0.21436 | 0.16444 | 0.00039996 | 0.00324605 |
| 336 | RH_Cont_Par_5 | 0.01054 | 0.17477 | 0.16423 | 0.00039996 | 0.00324605 |
| 324 | RH_Limbic_OFC_6 | −0.00013 | 0.16356 | 0.16369 | 9.999e−5 | 0.0022762 |
| 386 | RH_Default_PFCm_8 | 0.03441 | 0.19602 | 0.16161 | 9.999e−5 | 0.0022762 |
| 132 | LH_Cont_Par_6 | −0.00249 | 0.15814 | 0.16063 | 0.00039996 | 0.00324605 |
| 60 | LH_SomMot_29 | −0.00007 | 0.15974 | 0.15982 | 0.00019998 | 0.00324605 |
| 388 | RH_Default_PFCm_10 | 0.04376 | 0.20296 | 0.15920 | 9.999e−5 | 0.0022762 |
| 342 | RH_Cont_PFCl_2 | 0.01994 | 0.17881 | 0.15887 | 0.00049995 | 0.003535 |
| 390 | RH_Default_PFCm_12 | 0.05247 | 0.21067 | 0.15821 | 0.00039996 | 0.00324605 |
| 310 | RH_SalVentAttn_PFCl_1 | 0.03812 | 0.19599 | 0.15787 | 0.00039996 | 0.00324605 |
| 188 | LH_Default_PFC_23 | 0.04362 | 0.19953 | 0.15591 | 0.00039996 | 0.00324605 |
| 105 | LH_SalVentAttn_FrOper_9 | −0.00185 | 0.15389 | 0.15574 | 0.00049995 | 0.003535 |
| 359 | RH_Cont_Cing_2 | 0.05017 | 0.20382 | 0.15365 | 9.999e−5 | 0.0022762 |
| 51 | LH_SomMot_20 | −0.00097 | 0.15204 | 0.15301 | 0.00039996 | 0.00324605 |
| 348 | RH_Cont_PFCl_8 | 0.03128 | 0.18385 | 0.15257 | 0.00039996 | 0.00324605 |
| 68 | LH_SomMot_37 | 0.00428 | 0.15677 | 0.15250 | 0.00059994 | 0.00398064 |
| 235 | RH_SomMot_5 | 0.02070 | 0.17302 | 0.15232 | 0.00039996 | 0.00324605 |
| 382 | RH_Default_PFCm_4 | 0.01871 | 0.16947 | 0.15076 | 9.999e−5 | 0.0022762 |
| 253 | RH_SomMot_23 | 0.02813 | 0.17834 | 0.15021 | 0.00109989 | 0.0059684 |
| 53 | LH_SomMot_22 | −0.00074 | 0.14854 | 0.14928 | 0.00019998 | 0.00324605 |
| 138 | LH_Cont_PFCl_5 | 0.00400 | 0.15248 | 0.14849 | 9.999e−5 | 0.0022762 |
| 181 | LH_Default_PFC_16 | 0.00472 | 0.15311 | 0.14839 | 0.00039996 | 0.00324605 |
| 239 | RH_SomMot_9 | 0.02308 | 0.17032 | 0.14724 | 9.999e−5 | 0.0022762 |
| 236 | RH_SomMot_6 | −0.00123 | 0.14578 | 0.14701 | 9.999e−5 | 0.0022762 |
| 135 | LH_Cont_PFCl_2 | 0.00494 | 0.15145 | 0.14651 | 0.00049995 | 0.003535 |
| 137 | LH_Cont_PFCl_4 | 0.00375 | 0.14890 | 0.14514 | 0.00039996 | 0.00324605 |
| 345 | RH_Cont_PFCl_5 | 0.03055 | 0.17542 | 0.14487 | 0.00039996 | 0.00324605 |
| 89 | LH_DorsAttn_FEF_4 | 0.02642 | 0.17110 | 0.14468 | 0.00039996 | 0.00324605 |
| 312 | RH_SalVentAttn_Med_2 | 0.03814 | 0.18245 | 0.14431 | 0.00049995 | 0.003535 |
| 142 | LH_Cont_PFCl_9 | 0.01224 | 0.15576 | 0.14353 | 0.00039996 | 0.00324605 |
| 304 | RH_SalVentAttn_FrOper_3 | −0.00212 | 0.13951 | 0.14163 | 0.00169983 | 0.00819195 |
| 184 | LH_Default_PFC_19 | 0.01742 | 0.15865 | 0.14123 | 0.00039996 | 0.00324605 |
| 337 | RH_Cont_Par_6 | 0.00777 | 0.14819 | 0.14042 | 0.00029997 | 0.00324605 |
| 32 | LH_SomMot_1 | 0.00520 | 0.14518 | 0.13998 | 9.999e−5 | 0.0022762 |
| 95 | LH_SalVentAttn_ParOper_4 | −0.00157 | 0.13715 | 0.13872 | 9.999e−5 | 0.0022762 |
| 25 | LH_Vis_25 | 0.00904 | 0.14743 | 0.13839 | 0.0009999 | 0.00561066 |
| 129 | LH_Cont_Par_3 | −0.00233 | 0.13590 | 0.13823 | 0.00039996 | 0.00324605 |
| 186 | LH_Default_PFC_21 | 0.03009 | 0.16612 | 0.13602 | 0.00039996 | 0.00324605 |
| 355 | RH_Cont_PFCl_15 | 0.06000 | 0.19587 | 0.13588 | 9.999e−5 | 0.0022762 |
| 214 | RH_Vis_14 | 0.01519 | 0.15101 | 0.13582 | 0.00019998 | 0.00324605 |
| 263 | RH_SomMot_33 | 0.00780 | 0.14336 | 0.13555 | 0.00059994 | 0.00398064 |
| 380 | RH_Default_PFCm_2 | 0.00920 | 0.14442 | 0.13522 | 0.00029997 | 0.00324605 |
| 131 | LH_Cont_Par_5 | −0.00244 | 0.13262 | 0.13506 | 0.00039996 | 0.00324605 |
| 173 | LH_Default_PFC_8 | 0.00455 | 0.13873 | 0.13418 | 0.00019998 | 0.00324605 |
| 309 | RH_SalVentAttn_FrOper_8 | 0.02393 | 0.15714 | 0.13321 | 0.00049995 | 0.003535 |
| 62 | LH_SomMot_31 | 0.00154 | 0.13468 | 0.13314 | 0.00059994 | 0.00398064 |
| 364 | RH_Default_Par_3 | 0.01078 | 0.14319 | 0.13241 | 9.999e−5 | 0.0022762 |
| 351 | RH_Cont_PFCl_11 | 0.05131 | 0.18369 | 0.13237 | 0.00039996 | 0.00324605 |
| 178 | LH_Default_PFC_13 | 0.01699 | 0.14591 | 0.12892 | 0.00039996 | 0.00324605 |
| 276 | RH_DorsAttn_Post_6 | 0.03123 | 0.16011 | 0.12888 | 9.999e−5 | 0.0022762 |
| 180 | LH_Default_PFC_15 | 0.00655 | 0.13530 | 0.12875 | 0.00039996 | 0.00324605 |
| 240 | RH_SomMot_10 | 0.01614 | 0.14461 | 0.12847 | 9.999e−5 | 0.0022762 |
| 303 | RH_SalVentAttn_FrOper_2 | 0.01114 | 0.13884 | 0.12770 | 0.00609939 | 0.0232042 |
| 54 | LH_SomMot_23 | 0.00848 | 0.13585 | 0.12737 | 0.00069993 | 0.00439418 |
| 344 | RH_Cont_PFCl_4 | 0.02611 | 0.15205 | 0.12594 | 0.00049995 | 0.003535 |
| 268 | RH_SomMot_38 | 0.00722 | 0.13305 | 0.12583 | 0.0019998 | 0.00944256 |
| 352 | RH_Cont_PFCl_12 | 0.05493 | 0.18051 | 0.12558 | 0.00039996 | 0.00324605 |
| 250 | RH_SomMot_20 | 0.05203 | 0.17727 | 0.12524 | 9.999e−5 | 0.0022762 |
| 302 | RH_SalVentAttn_FrOper_1 | −0.00026 | 0.12458 | 0.12484 | 0.00039996 | 0.00324605 |
| 24 | LH_Vis_24 | 0.01697 | 0.14117 | 0.12421 | 0.00129987 | 0.00672761 |
| 384 | RH_Default_PFCm_6 | 0.02023 | 0.14432 | 0.12409 | 0.00059994 | 0.00398064 |
| 12 | LH_Vis_12 | 0.03581 | 0.15835 | 0.12254 | 0.00119988 | 0.00631516 |
| 224 | RH_Vis_24 | 0.00154 | 0.12353 | 0.12199 | 0.00239976 | 0.0109793 |
| 349 | RH_Cont_PFCl_9 | 0.04314 | 0.16508 | 0.12194 | 0.00049995 | 0.003535 |
| 340 | RH_Cont_PFCv_1 | 0.01364 | 0.13551 | 0.12186 | 0.00289971 | 0.0126664 |
| 170 | LH_Default_PFC_5 | −0.00241 | 0.11942 | 0.12182 | 0.00059994 | 0.00398064 |
| 130 | LH_Cont_Par_4 | −0.00215 | 0.11860 | 0.12075 | 0.00089991 | 0.00521687 |
| 333 | RH_Cont_Par_2 | 0.01805 | 0.13705 | 0.11899 | 0.00039996 | 0.00324605 |
| 93 | LH_SalVentAttn_ParOper_2 | 0.00480 | 0.12326 | 0.11846 | 0.00249975 | 0.0112529 |
| 134 | LH_Cont_PFCl_1 | 0.01027 | 0.12775 | 0.11748 | 0.00119988 | 0.00631516 |
| 139 | LH_Cont_PFCl_6 | 0.00653 | 0.12381 | 0.11728 | 0.00049995 | 0.003535 |
| 256 | RH_SomMot_26 | 0.01418 | 0.13075 | 0.11657 | 0.00149985 | 0.00740667 |
| 74 | LH_DorsAttn_Post_6 | −0.00250 | 0.11253 | 0.11503 | 0.00069993 | 0.00439418 |
| 366 | RH_Default_Par_5 | 0.00212 | 0.11485 | 0.11273 | 9.999e−5 | 0.0022762 |
| 67 | LH_SomMot_36 | 0.00229 | 0.11478 | 0.11250 | 0.00429957 | 0.017347 |
| 269 | RH_SomMot_39 | 0.01938 | 0.13071 | 0.11134 | 0.00409959 | 0.0167574 |
| 385 | RH_Default_PFCm_7 | 0.01657 | 0.12704 | 0.11047 | 0.00079992 | 0.00479609 |
| 116 | LH_Limbic_OFC_3 | 0.00023 | 0.11050 | 0.11027 | 0.00139986 | 0.00702439 |
| 183 | LH_Default_PFC_18 | 0.03078 | 0.14081 | 0.11003 | 0.00049995 | 0.003535 |
| 165 | LH_Default_Temp_17 | −0.00207 | 0.10643 | 0.10850 | 0.00049995 | 0.003535 |
| 298 | RH_SalVentAttn_TempOccPar_5 | 0.01759 | 0.12572 | 0.10813 | 0.00079992 | 0.00479609 |
| 334 | RH_Cont_Par_3 | 0.00099 | 0.10881 | 0.10781 | 0.00059994 | 0.00398064 |
| 48 | LH_SomMot_17 | −0.00183 | 0.10545 | 0.10728 | 0.00479952 | 0.019008 |
| 241 | RH_SomMot_11 | 0.01290 | 0.12018 | 0.10728 | 0.00079992 | 0.00479609 |
| 127 | LH_Cont_Par_1 | 0.00315 | 0.10972 | 0.10657 | 0.00179982 | 0.00859982 |
| 179 | LH_Default_PFC_14 | 0.01288 | 0.11862 | 0.10573 | 0.00129987 | 0.00672761 |
| 353 | RH_Cont_PFCl_13 | 0.04972 | 0.15537 | 0.10565 | 0.00049995 | 0.003535 |
| 118 | LH_Limbic_OFC_5 | −0.00242 | 0.10255 | 0.10496 | 0.00089991 | 0.00521687 |
| 94 | LH_SalVentAttn_ParOper_3 | −0.00223 | 0.10255 | 0.10478 | 0.00079992 | 0.00479609 |
| 335 | RH_Cont_Par_4 | 0.01944 | 0.12280 | 0.10336 | 0.00109989 | 0.0059684 |
| 161 | LH_Default_Temp_13 | 0.00437 | 0.10691 | 0.10254 | 0.00039996 | 0.00324605 |
| 133 | LH_Cont_Temp_1 | 0.02082 | 0.12314 | 0.10232 | 0.00039996 | 0.00324605 |
| 90 | LH_DorsAttn_PrCv_1 | −0.00045 | 0.10170 | 0.10216 | 0.00169983 | 0.00819195 |
| 128 | LH_Cont_Par_2 | −0.00236 | 0.09972 | 0.10208 | 0.00119988 | 0.00631516 |
| 363 | RH_Default_Par_2 | 0.00291 | 0.10466 | 0.10175 | 0.00359964 | 0.0152944 |
| 148 | LH_Cont_PFCmp_1 | 0.03437 | 0.13603 | 0.10166 | 0.0009999 | 0.00561066 |
| 15 | LH_Vis_15 | 0.03273 | 0.13364 | 0.10091 | 0.0039996 | 0.0164448 |
| 103 | LH_SalVentAttn_FrOper_7 | 0.00165 | 0.10056 | 0.09891 | 0.00239976 | 0.0109793 |
| 33 | LH_SomMot_2 | 0.00293 | 0.10115 | 0.09823 | 0.00949905 | 0.0328362 |
| 176 | LH_Default_PFC_11 | 0.01046 | 0.10863 | 0.09818 | 0.00289971 | 0.0126664 |
| 220 | RH_Vis_20 | 0.00746 | 0.10501 | 0.09755 | 0.00379962 | 0.0158553 |
| 172 | LH_Default_PFC_7 | −0.00178 | 0.09357 | 0.09535 | 0.00159984 | 0.00783138 |
| 332 | RH_Cont_Par_1 | 0.00975 | 0.10503 | 0.09528 | 0.00059994 | 0.00398064 |
| 140 | LH_Cont_PFCl_7 | 0.00306 | 0.09796 | 0.09490 | 0.00239976 | 0.0109793 |
| 361 | RH_Cont_PFCmp_2 | 0.05129 | 0.14575 | 0.09447 | 0.00119988 | 0.00631516 |
| 175 | LH_Default_PFC_10 | 0.00418 | 0.09767 | 0.09348 | 0.00229977 | 0.010626 |
| 280 | RH_DorsAttn_Post_10 | 0.00461 | 0.09768 | 0.09308 | 0.00409959 | 0.0167574 |
| 30 | LH_Vis_30 | 0.00253 | 0.09409 | 0.09156 | 0.0109989 | 0.0368825 |
| 147 | LH_Cont_Cing_2 | 0.02754 | 0.11888 | 0.09134 | 0.00039996 | 0.00324605 |
| 296 | RH_SalVentAttn_TempOccPar_3 | 0.01354 | 0.10436 | 0.09082 | 0.00069993 | 0.00439418 |
| 383 | RH_Default_PFCm_5 | 0.01607 | 0.10587 | 0.08980 | 0.00379962 | 0.0158553 |
| 36 | LH_SomMot_5 | −0.00040 | 0.08937 | 0.08976 | 0.00149985 | 0.00740667 |
| 71 | LH_DorsAttn_Post_3 | 0.02219 | 0.11118 | 0.08899 | 0.00249975 | 0.0112529 |
| 80 | LH_DorsAttn_Post_12 | −0.00221 | 0.08367 | 0.08588 | 0.00429957 | 0.017347 |
| 136 | LH_Cont_PFCl_3 | 0.00208 | 0.08770 | 0.08562 | 0.00249975 | 0.0112529 |
| 182 | LH_Default_PFC_17 | 0.02317 | 0.10860 | 0.08543 | 0.00109989 | 0.0059684 |
| 50 | LH_SomMot_19 | −0.00011 | 0.08283 | 0.08294 | 0.00819918 | 0.0290604 |
| 299 | RH_SalVentAttn_TempOccPar_6 | 0.01606 | 0.09893 | 0.08287 | 0.0029997 | 0.0130422 |
| 358 | RH_Cont_Cing_1 | 0.02963 | 0.11248 | 0.08286 | 0.00039996 | 0.00324605 |
| 96 | LH_SalVentAttn_TempOcc_1 | 0.00793 | 0.08958 | 0.08165 | 0.00639936 | 0.024019 |
| 300 | RH_SalVentAttn_TempOccPar_7 | 0.01596 | 0.09709 | 0.08113 | 0.00249975 | 0.0112529 |
| 285 | RH_DorsAttn_Post_15 | −0.00059 | 0.08014 | 0.08073 | 0.0039996 | 0.0164448 |
| 391 | RH_Default_PFCm_13 | 0.05799 | 0.13803 | 0.08003 | 0.0009999 | 0.00561066 |
| 52 | LH_SomMot_21 | −0.00163 | 0.07707 | 0.07869 | 0.0121988 | 0.0396708 |
| 225 | RH_Vis_25 | 0.00616 | 0.08437 | 0.07820 | 0.0149985 | 0.0473993 |
| 109 | LH_SalVentAttn_Med_3 | 0.01119 | 0.08909 | 0.07789 | 0.00169983 | 0.00819195 |
| 317 | RH_SalVentAttn_Med_7 | 0.00659 | 0.08421 | 0.07763 | 0.00129987 | 0.00672761 |
| 84 | LH_DorsAttn_Post_16 | −0.00249 | 0.07302 | 0.07551 | 0.00909909 | 0.0316884 |
| 387 | RH_Default_PFCm_9 | 0.01722 | 0.09244 | 0.07523 | 0.00109989 | 0.0059684 |
| 243 | RH_SomMot_13 | 0.01199 | 0.08520 | 0.07320 | 0.00469953 | 0.0186913 |
| 28 | LH_Vis_28 | 0.00975 | 0.08268 | 0.07294 | 0.0118988 | 0.0392885 |
| 6 | LH_Vis_6 | 0.04369 | 0.11643 | 0.07275 | 0.00419958 | 0.0170418 |
| 339 | RH_Cont_Temp_2 | −0.00246 | 0.07000 | 0.07247 | 0.0152985 | 0.048076 |
| 212 | RH_Vis_12 | 0.01457 | 0.08677 | 0.07220 | 0.00339966 | 0.0146673 |
| 150 | LH_Default_Temp_2 | 0.01640 | 0.08814 | 0.07174 | 0.0149985 | 0.0473993 |
| 81 | LH_DorsAttn_Post_13 | −0.00175 | 0.06912 | 0.07087 | 0.00669933 | 0.0248781 |
| 20 | LH_Vis_20 | 0.04594 | 0.11496 | 0.06902 | 0.0019998 | 0.00944256 |
| 39 | LH_SomMot_8 | −0.00034 | 0.06854 | 0.06887 | 0.00289971 | 0.0126664 |
| 234 | RH_SomMot_4 | 0.00792 | 0.07674 | 0.06882 | 0.00229977 | 0.010626 |
| 29 | LH_Vis_29 | 0.00438 | 0.07268 | 0.06830 | 0.0142986 | 0.0454438 |
| 378 | RH_Default_PFCv_4 | 0.02651 | 0.09356 | 0.06705 | 0.0130987 | 0.0421083 |
| 164 | LH_Default_Temp_16 | −0.00248 | 0.06455 | 0.06703 | 0.00289971 | 0.0126664 |
| 35 | LH_SomMot_4 | −0.00026 | 0.06619 | 0.06645 | 0.0009999 | 0.00561066 |
| 34 | LH_SomMot_3 | 0.00537 | 0.07039 | 0.06502 | 0.00849915 | 0.0298965 |
| 274 | RH_DorsAttn_Post_4 | −0.00177 | 0.06302 | 0.06479 | 0.00469953 | 0.0186913 |
| 205 | RH_Vis_5 | 0.00931 | 0.07397 | 0.06466 | 0.0128987 | 0.0416087 |
| 159 | LH_Default_Temp_11 | 0.01139 | 0.07528 | 0.06389 | 0.00749925 | 0.0270591 |
| 193 | LH_Default_PCC_4 | 0.00209 | 0.06531 | 0.06322 | 0.00459954 | 0.0183982 |
| 98 | LH_SalVentAttn_FrOper_2 | 0.00310 | 0.06329 | 0.06019 | 0.00629937 | 0.0238033 |
| 301 | RH_SalVentAttn_PrC_1 | 0.04247 | 0.10161 | 0.05914 | 0.0109989 | 0.0368825 |
| 288 | RH_DorsAttn_Post_18 | 0.00521 | 0.06420 | 0.05899 | 0.0112989 | 0.0377979 |
| 318 | RH_SalVentAttn_Med_8 | 0.05402 | 0.11271 | 0.05869 | 0.00719928 | 0.0262132 |
| 311 | RH_SalVentAttn_Med_1 | 0.05240 | 0.10945 | 0.05705 | 0.0151985 | 0.0478154 |
| 400 | RH_Default_PCC_9 | 0.01172 | 0.06864 | 0.05692 | 0.010199 | 0.0349109 |
| 346 | RH_Cont_PFCl_6 | 0.03582 | 0.09206 | 0.05623 | 0.0155984 | 0.0486908 |
| 9 | LH_Vis_9 | 0.04032 | 0.09491 | 0.05459 | 0.0156984 | 0.0488939 |
| 167 | LH_Default_PFC_2 | 0.00059 | 0.05501 | 0.05442 | 0.00349965 | 0.0149375 |
| 97 | LH_SalVentAttn_FrOper_1 | 0.00882 | 0.06034 | 0.05152 | 0.00929907 | 0.0322245 |
| 217 | RH_Vis_17 | 0.04371 | 0.09499 | 0.05129 | 0.0059994 | 0.0228549 |
| 198 | LH_Default_PCC_9 | 0.02109 | 0.07110 | 0.05001 | 0.0120988 | 0.0394832 |
| 163 | LH_Default_Temp_15 | −0.00220 | 0.04751 | 0.04971 | 0.0121988 | 0.0396708 |
| 13 | LH_Vis_13 | 0.05017 | 0.09755 | 0.04738 | 0.0120988 | 0.0394832 |
| 292 | RH_DorsAttn_FEF_3 | 0.05353 | 0.09823 | 0.04470 | 0.00889911 | 0.0311469 |
| 82 | LH_DorsAttn_Post_14 | 0.01247 | 0.05626 | 0.04379 | 0.0128987 | 0.0416087 |
| 189 | LH_Default_PFC_24 | 0.03731 | 0.07627 | 0.03896 | 0.0117988 | 0.0390505 |

Table S9. Continued. D. Significant regional gains of receptor-informed CAP depth-profile models.

| **CAP-4: 15 / 400 significant parcels (3.75%)** | | | | | | |
| --- | --- | --- | --- | --- | --- | --- |
| **ROI ID** | **Atlas parcel** | **Baseline adjusted R²** | **Full model adjusted R²** | **Gain (Δadj. R²)** | **Spin p** | **FDR q** |
| 246 | RH_SomMot_16 | 0.01765 | 0.30666 | 0.28901 | 0.00039996 | 0.00324605 |
| 60 | LH_SomMot_29 | 0.10207 | 0.33978 | 0.23771 | 9.999e−5 | 0.0022762 |
| 134 | LH_Cont_PFCl_1 | 0.04072 | 0.22586 | 0.18514 | 9.999e−5 | 0.0022762 |
| 66 | LH_SomMot_35 | 0.07133 | 0.24994 | 0.17861 | 0.00039996 | 0.00324605 |
| 64 | LH_SomMot_33 | 0.13391 | 0.29770 | 0.16379 | 0.00049995 | 0.003535 |
| 244 | RH_SomMot_14 | 0.03990 | 0.20049 | 0.16058 | 0.00929907 | 0.0322245 |
| 253 | RH_SomMot_23 | 0.00987 | 0.16922 | 0.15935 | 0.00039996 | 0.00324605 |
| 45 | LH_SomMot_14 | 0.00897 | 0.16374 | 0.15477 | 9.999e−5 | 0.0022762 |
| 262 | RH_SomMot_32 | 0.05954 | 0.20138 | 0.14184 | 9.999e−5 | 0.0022762 |
| 264 | RH_SomMot_34 | 0.04004 | 0.17414 | 0.13410 | 9.999e−5 | 0.0022762 |
| 57 | LH_SomMot_26 | 0.03563 | 0.15140 | 0.11576 | 0.0009999 | 0.00561066 |
| 196 | LH_Default_PCC_7 | 0.04100 | 0.14614 | 0.10514 | 0.0142986 | 0.0454438 |
| 397 | RH_Default_PCC_6 | 0.08421 | 0.18680 | 0.10259 | 0.0113989 | 0.0379962 |
| 257 | RH_SomMot_27 | 0.19150 | 0.28099 | 0.08949 | 0.0120988 | 0.0394832 |
| 167 | LH_Default_PFC_2 | 0.00881 | 0.08050 | 0.07169 | 0.0119988 | 0.0394327 |

Table S9. Continued. E. Significant regional gains of receptor-informed CAP depth-profile models.

| **CAP-5: 187 / 400 significant parcels (46.75%)** | | | | | | |
| --- | --- | --- | --- | --- | --- | --- |
| **ROI ID** | **Atlas parcel** | **Baseline adjusted R²** | **Full model adjusted R²** | **Gain (Δadj. R²)** | **Spin p** | **FDR q** |
| 134 | LH_Cont_PFCl_1 | 0.11954 | 0.51225 | 0.39271 | 9.999e−5 | 0.0022762 |
| 233 | RH_SomMot_3 | 0.00040 | 0.38159 | 0.38119 | 9.999e−5 | 0.0022762 |
| 59 | LH_SomMot_28 | 0.06038 | 0.42341 | 0.36303 | 0.00039996 | 0.00324605 |
| 51 | LH_SomMot_20 | 0.02335 | 0.38479 | 0.36144 | 9.999e−5 | 0.0022762 |
| 64 | LH_SomMot_33 | 0.08020 | 0.43986 | 0.35967 | 9.999e−5 | 0.0022762 |
| 48 | LH_SomMot_17 | 0.01478 | 0.37325 | 0.35847 | 9.999e−5 | 0.0022762 |
| 116 | LH_Limbic_OFC_3 | 0.11522 | 0.46935 | 0.35413 | 9.999e−5 | 0.0022762 |
| 55 | LH_SomMot_24 | 0.06749 | 0.42161 | 0.35412 | 9.999e−5 | 0.0022762 |
| 53 | LH_SomMot_22 | 0.04021 | 0.39400 | 0.35380 | 9.999e−5 | 0.0022762 |
| 251 | RH_SomMot_21 | 0.01900 | 0.36654 | 0.34754 | 9.999e−5 | 0.0022762 |
| 66 | LH_SomMot_35 | 0.07655 | 0.40415 | 0.32759 | 0.00039996 | 0.00324605 |
| 259 | RH_SomMot_29 | 0.06542 | 0.39203 | 0.32661 | 0.00039996 | 0.00324605 |
| 255 | RH_SomMot_25 | 0.03595 | 0.35988 | 0.32393 | 9.999e−5 | 0.0022762 |
| 60 | LH_SomMot_29 | 0.07229 | 0.39595 | 0.32366 | 9.999e−5 | 0.0022762 |
| 270 | RH_SomMot_40 | 0.08866 | 0.40080 | 0.31214 | 9.999e−5 | 0.0022762 |
| 68 | LH_SomMot_37 | 0.07388 | 0.38509 | 0.31121 | 9.999e−5 | 0.0022762 |
| 354 | RH_Cont_PFCl_14 | −0.00138 | 0.30867 | 0.31005 | 0.00039996 | 0.00324605 |
| 63 | LH_SomMot_32 | 0.10337 | 0.41233 | 0.30895 | 0.00039996 | 0.00324605 |
| 262 | RH_SomMot_32 | 0.10570 | 0.40879 | 0.30309 | 0.00039996 | 0.00324605 |
| 264 | RH_SomMot_34 | 0.08075 | 0.38010 | 0.29936 | 9.999e−5 | 0.0022762 |
| 241 | RH_SomMot_11 | 0.00171 | 0.30037 | 0.29866 | 9.999e−5 | 0.0022762 |
| 257 | RH_SomMot_27 | 0.06723 | 0.36189 | 0.29466 | 0.00039996 | 0.00324605 |
| 260 | RH_SomMot_30 | 0.07798 | 0.36729 | 0.28931 | 0.00039996 | 0.00324605 |
| 254 | RH_SomMot_24 | 0.02351 | 0.31157 | 0.28806 | 9.999e−5 | 0.0022762 |
| 389 | RH_Default_PFCm_11 | 0.04096 | 0.32810 | 0.28714 | 0.00039996 | 0.00324605 |
| 365 | RH_Default_Par_4 | 0.04475 | 0.33106 | 0.28631 | 0.00039996 | 0.00324605 |
| 341 | RH_Cont_PFCl_1 | 0.11486 | 0.39976 | 0.28490 | 9.999e−5 | 0.0022762 |
| 62 | LH_SomMot_31 | 0.06635 | 0.35067 | 0.28431 | 0.00039996 | 0.00324605 |
| 265 | RH_SomMot_35 | 0.07810 | 0.35901 | 0.28091 | 0.00039996 | 0.00324605 |
| 49 | LH_SomMot_18 | 0.03818 | 0.31791 | 0.27973 | 9.999e−5 | 0.0022762 |
| 50 | LH_SomMot_19 | 0.02044 | 0.29029 | 0.26985 | 0.00039996 | 0.00324605 |
| 56 | LH_SomMot_25 | 0.02498 | 0.29160 | 0.26662 | 9.999e−5 | 0.0022762 |
| 324 | RH_Limbic_OFC_6 | 0.13867 | 0.40435 | 0.26568 | 0.00039996 | 0.00324605 |
| 384 | RH_Default_PFCm_6 | 0.04090 | 0.30212 | 0.26122 | 0.00039996 | 0.00324605 |
| 355 | RH_Cont_PFCl_15 | 0.00136 | 0.26243 | 0.26107 | 9.999e−5 | 0.0022762 |
| 57 | LH_SomMot_26 | 0.04056 | 0.29915 | 0.25859 | 0.00039996 | 0.00324605 |
| 45 | LH_SomMot_14 | 0.00167 | 0.25946 | 0.25778 | 0.00039996 | 0.00324605 |
| 33 | LH_SomMot_2 | 0.00365 | 0.25874 | 0.25509 | 0.00039996 | 0.00324605 |
| 268 | RH_SomMot_38 | 0.08115 | 0.33389 | 0.25274 | 9.999e−5 | 0.0022762 |
| 256 | RH_SomMot_26 | 0.05085 | 0.30356 | 0.25271 | 0.00039996 | 0.00324605 |
| 103 | LH_SalVentAttn_FrOper_7 | 0.01992 | 0.26905 | 0.24913 | 0.00039996 | 0.00324605 |
| 263 | RH_SomMot_33 | 0.07480 | 0.32182 | 0.24703 | 9.999e−5 | 0.0022762 |
| 390 | RH_Default_PFCm_12 | 0.00924 | 0.25129 | 0.24205 | 0.00039996 | 0.00324605 |
| 74 | LH_DorsAttn_Post_6 | 0.02646 | 0.26510 | 0.23864 | 9.999e−5 | 0.0022762 |
| 348 | RH_Cont_PFCl_8 | 0.09834 | 0.33622 | 0.23788 | 0.00039996 | 0.00324605 |
| 364 | RH_Default_Par_3 | 0.04545 | 0.28164 | 0.23619 | 0.00039996 | 0.00324605 |
| 253 | RH_SomMot_23 | 0.03694 | 0.26669 | 0.22975 | 0.00039996 | 0.00324605 |
| 151 | LH_Default_Temp_3 | 0.00275 | 0.23048 | 0.22773 | 9.999e−5 | 0.0022762 |
| 150 | LH_Default_Temp_2 | −0.00246 | 0.22434 | 0.22680 | 0.00039996 | 0.00324605 |
| 234 | RH_SomMot_4 | 0.00103 | 0.22678 | 0.22575 | 0.00039996 | 0.00324605 |
| 278 | RH_DorsAttn_Post_8 | 0.07269 | 0.29772 | 0.22503 | 0.00039996 | 0.00324605 |
| 129 | LH_Cont_Par_3 | 0.04481 | 0.26868 | 0.22386 | 0.00039996 | 0.00324605 |
| 386 | RH_Default_PFCm_8 | 0.08119 | 0.30461 | 0.22343 | 9.999e−5 | 0.0022762 |
| 168 | LH_Default_PFC_3 | 0.12003 | 0.34234 | 0.22231 | 0.00039996 | 0.00324605 |
| 385 | RH_Default_PFCm_7 | 0.08783 | 0.30991 | 0.22209 | 9.999e−5 | 0.0022762 |
| 246 | RH_SomMot_16 | 0.01333 | 0.23389 | 0.22056 | 9.999e−5 | 0.0022762 |
| 128 | LH_Cont_Par_2 | 0.02850 | 0.24729 | 0.21879 | 0.00039996 | 0.00324605 |
| 67 | LH_SomMot_36 | 0.08367 | 0.30223 | 0.21856 | 9.999e−5 | 0.0022762 |
| 171 | LH_Default_PFC_6 | 0.11124 | 0.32851 | 0.21727 | 9.999e−5 | 0.0022762 |
| 118 | LH_Limbic_OFC_5 | 0.12982 | 0.34269 | 0.21287 | 0.00039996 | 0.00324605 |
| 249 | RH_SomMot_19 | 0.02624 | 0.23903 | 0.21279 | 0.00049995 | 0.003535 |
| 236 | RH_SomMot_6 | 0.02024 | 0.23275 | 0.21251 | 9.999e−5 | 0.0022762 |
| 146 | LH_Cont_Cing_1 | −0.00139 | 0.20906 | 0.21045 | 0.00039996 | 0.00324605 |
| 322 | RH_Limbic_OFC_4 | 0.06140 | 0.26998 | 0.20858 | 9.999e−5 | 0.0022762 |
| 363 | RH_Default_Par_2 | 0.05127 | 0.25984 | 0.20857 | 0.00039996 | 0.00324605 |
| 133 | LH_Cont_Temp_1 | 0.01885 | 0.22720 | 0.20835 | 9.999e−5 | 0.0022762 |
| 132 | LH_Cont_Par_6 | 0.04910 | 0.25641 | 0.20731 | 0.00039996 | 0.00324605 |
| 187 | LH_Default_PFC_22 | 0.00943 | 0.21498 | 0.20555 | 0.00039996 | 0.00324605 |
| 54 | LH_SomMot_23 | 0.09190 | 0.29655 | 0.20465 | 9.999e−5 | 0.0022762 |
| 165 | LH_Default_Temp_17 | 0.05759 | 0.26199 | 0.20440 | 0.00039996 | 0.00324605 |
| 108 | LH_SalVentAttn_Med_2 | 0.00016 | 0.20042 | 0.20026 | 0.00039996 | 0.00324605 |
| 162 | LH_Default_Temp_14 | 0.05464 | 0.25486 | 0.20022 | 9.999e−5 | 0.0022762 |
| 160 | LH_Default_Temp_12 | 0.04738 | 0.24567 | 0.19829 | 9.999e−5 | 0.0022762 |
| 371 | RH_Default_Temp_5 | 0.00276 | 0.20027 | 0.19752 | 9.999e−5 | 0.0022762 |
| 333 | RH_Cont_Par_2 | 0.04597 | 0.24347 | 0.19750 | 9.999e−5 | 0.0022762 |
| 252 | RH_SomMot_22 | 0.02517 | 0.22184 | 0.19668 | 0.00039996 | 0.00324605 |
| 335 | RH_Cont_Par_4 | 0.03362 | 0.22731 | 0.19369 | 0.00039996 | 0.00324605 |
| 167 | LH_Default_PFC_2 | 0.10486 | 0.29831 | 0.19345 | 9.999e−5 | 0.0022762 |
| 383 | RH_Default_PFCm_5 | 0.11236 | 0.30449 | 0.19213 | 9.999e−5 | 0.0022762 |
| 312 | RH_SalVentAttn_Med_2 | −0.00082 | 0.19056 | 0.19138 | 9.999e−5 | 0.0022762 |
| 351 | RH_Cont_PFCl_11 | 0.02985 | 0.21963 | 0.18978 | 0.00039996 | 0.00324605 |
| 239 | RH_SomMot_9 | 0.00720 | 0.19672 | 0.18952 | 0.00039996 | 0.00324605 |
| 338 | RH_Cont_Temp_1 | 0.00392 | 0.19153 | 0.18761 | 0.00069993 | 0.00439418 |
| 337 | RH_Cont_Par_6 | 0.07127 | 0.25676 | 0.18550 | 9.999e−5 | 0.0022762 |
| 154 | LH_Default_Temp_6 | 0.00892 | 0.19365 | 0.18472 | 0.00019998 | 0.00324605 |
| 106 | LH_SalVentAttn_PFCl_1 | 0.07323 | 0.25789 | 0.18466 | 0.00039996 | 0.00324605 |
| 235 | RH_SomMot_5 | −0.00125 | 0.18120 | 0.18245 | 0.00029997 | 0.00324605 |
| 316 | RH_SalVentAttn_Med_6 | 0.08399 | 0.26316 | 0.17916 | 9.999e−5 | 0.0022762 |
| 72 | LH_DorsAttn_Post_4 | 0.05603 | 0.23416 | 0.17812 | 9.999e−5 | 0.0022762 |
| 362 | RH_Default_Par_1 | 0.02993 | 0.20668 | 0.17675 | 0.00049995 | 0.003535 |
| 380 | RH_Default_PFCm_2 | 0.12440 | 0.30094 | 0.17654 | 0.00039996 | 0.00324605 |
| 164 | LH_Default_Temp_16 | 0.06965 | 0.24041 | 0.17076 | 0.00039996 | 0.00324605 |
| 248 | RH_SomMot_18 | 0.01629 | 0.18696 | 0.17067 | 0.00039996 | 0.00324605 |
| 240 | RH_SomMot_10 | 0.03181 | 0.20240 | 0.17059 | 0.00039996 | 0.00324605 |
| 34 | LH_SomMot_3 | 0.01132 | 0.18157 | 0.17025 | 0.00039996 | 0.00324605 |
| 8 | LH_Vis_8 | 0.06175 | 0.23108 | 0.16933 | 0.00039996 | 0.00324605 |
| 188 | LH_Default_PFC_23 | 0.00431 | 0.17294 | 0.16863 | 0.00019998 | 0.00324605 |
| 294 | RH_SalVentAttn_TempOccPar_1 | 0.03326 | 0.20160 | 0.16834 | 0.00049995 | 0.003535 |
| 269 | RH_SomMot_39 | 0.06200 | 0.22902 | 0.16701 | 0.00259974 | 0.0115728 |
| 130 | LH_Cont_Par_4 | 0.07397 | 0.24024 | 0.16627 | 9.999e−5 | 0.0022762 |
| 342 | RH_Cont_PFCl_2 | 0.08783 | 0.25315 | 0.16531 | 9.999e−5 | 0.0022762 |
| 336 | RH_Cont_Par_5 | 0.06215 | 0.22685 | 0.16470 | 9.999e−5 | 0.0022762 |
| 332 | RH_Cont_Par_1 | 0.05593 | 0.22049 | 0.16456 | 9.999e−5 | 0.0022762 |
| 368 | RH_Default_Temp_2 | −0.00125 | 0.16266 | 0.16391 | 0.00019998 | 0.00324605 |
| 163 | LH_Default_Temp_15 | 0.03704 | 0.20043 | 0.16339 | 0.00049995 | 0.003535 |
| 343 | RH_Cont_PFCl_3 | 0.10823 | 0.27091 | 0.16268 | 0.00039996 | 0.00324605 |
| 271 | RH_DorsAttn_Post_1 | 0.02454 | 0.18699 | 0.16245 | 0.00059994 | 0.00398064 |
| 122 | LH_Limbic_TempPole_4 | −0.00129 | 0.16106 | 0.16235 | 0.00179982 | 0.00859982 |
| 161 | LH_Default_Temp_13 | 0.07279 | 0.23499 | 0.16219 | 0.00039996 | 0.00324605 |
| 152 | LH_Default_Temp_4 | −0.00163 | 0.16004 | 0.16167 | 0.00049995 | 0.003535 |
| 131 | LH_Cont_Par_5 | 0.06138 | 0.22174 | 0.16037 | 0.00049995 | 0.003535 |
| 339 | RH_Cont_Temp_2 | 0.02039 | 0.17985 | 0.15945 | 0.00039996 | 0.00324605 |
| 330 | RH_Limbic_TempPole_6 | 0.00335 | 0.16202 | 0.15867 | 0.0049995 | 0.0196059 |
| 70 | LH_DorsAttn_Post_2 | 0.00560 | 0.16376 | 0.15816 | 0.00059994 | 0.00398064 |
| 366 | RH_Default_Par_5 | 0.06464 | 0.22256 | 0.15792 | 9.999e−5 | 0.0022762 |
| 149 | LH_Default_Temp_1 | 0.01030 | 0.16768 | 0.15738 | 0.00049995 | 0.003535 |
| 273 | RH_DorsAttn_Post_3 | 0.03039 | 0.18583 | 0.15545 | 0.00039996 | 0.00324605 |
| 39 | LH_SomMot_8 | 0.00294 | 0.15804 | 0.15510 | 0.00039996 | 0.00324605 |
| 32 | LH_SomMot_1 | −0.00234 | 0.15239 | 0.15473 | 0.00039996 | 0.00324605 |
| 321 | RH_Limbic_OFC_3 | 0.05244 | 0.20497 | 0.15253 | 0.00039996 | 0.00324605 |
| 180 | LH_Default_PFC_15 | 0.08670 | 0.23858 | 0.15188 | 0.00039996 | 0.00324605 |
| 209 | RH_Vis_9 | 0.05551 | 0.20731 | 0.15180 | 0.00019998 | 0.00324605 |
| 345 | RH_Cont_PFCl_5 | 0.07266 | 0.22379 | 0.15113 | 0.00039996 | 0.00324605 |
| 391 | RH_Default_PFCm_13 | −0.00004 | 0.15044 | 0.15047 | 0.00079992 | 0.00479609 |
| 184 | LH_Default_PFC_19 | 0.04650 | 0.19658 | 0.15008 | 0.00059994 | 0.00398064 |
| 69 | LH_DorsAttn_Post_1 | 0.00473 | 0.15416 | 0.14944 | 0.00259974 | 0.0115728 |
| 138 | LH_Cont_PFCl_5 | 0.09884 | 0.24811 | 0.14927 | 0.00039996 | 0.00324605 |
| 299 | RH_SalVentAttn_TempOccPar_6 | 0.02013 | 0.16899 | 0.14886 | 0.00059994 | 0.00398064 |
| 352 | RH_Cont_PFCl_12 | 0.00472 | 0.15356 | 0.14883 | 0.00149985 | 0.00740667 |
| 183 | LH_Default_PFC_18 | 0.04673 | 0.19520 | 0.14847 | 9.999e−5 | 0.0022762 |
| 373 | RH_Default_Temp_7 | 0.01301 | 0.15831 | 0.14530 | 0.00089991 | 0.00521687 |
| 387 | RH_Default_PFCm_9 | 0.07543 | 0.21756 | 0.14213 | 0.00019998 | 0.00324605 |
| 96 | LH_SalVentAttn_TempOcc_1 | 0.01996 | 0.16114 | 0.14118 | 0.00089991 | 0.00521687 |
| 181 | LH_Default_PFC_16 | 0.03654 | 0.17688 | 0.14035 | 0.00219978 | 0.0102828 |
| 388 | RH_Default_PFCm_10 | 0.03829 | 0.17789 | 0.13960 | 0.00029997 | 0.00324605 |
| 310 | RH_SalVentAttn_PFCl_1 | 0.07114 | 0.21029 | 0.13915 | 0.00039996 | 0.00324605 |
| 178 | LH_Default_PFC_13 | 0.09581 | 0.23417 | 0.13836 | 0.00039996 | 0.00324605 |
| 243 | RH_SomMot_13 | 0.01749 | 0.15547 | 0.13799 | 0.00119988 | 0.00631516 |
| 399 | RH_Default_PCC_8 | 0.07339 | 0.20722 | 0.13384 | 0.00019998 | 0.00324605 |
| 47 | LH_SomMot_16 | 0.00459 | 0.13711 | 0.13252 | 0.00639936 | 0.024019 |
| 186 | LH_Default_PFC_21 | 0.00194 | 0.13431 | 0.13237 | 0.00649935 | 0.0242642 |
| 297 | RH_SalVentAttn_TempOccPar_4 | 0.02385 | 0.15200 | 0.12814 | 0.00649935 | 0.0242642 |
| 52 | LH_SomMot_21 | 0.04045 | 0.16781 | 0.12736 | 0.00229977 | 0.010626 |
| 300 | RH_SalVentAttn_TempOccPar_7 | 0.01718 | 0.14402 | 0.12684 | 0.00109989 | 0.0059684 |
| 176 | LH_Default_PFC_11 | 0.11688 | 0.24210 | 0.12522 | 0.00029997 | 0.00324605 |
| 135 | LH_Cont_PFCl_2 | 0.09317 | 0.21562 | 0.12245 | 0.00029997 | 0.00324605 |
| 274 | RH_DorsAttn_Post_4 | 0.07506 | 0.19735 | 0.12229 | 9.999e−5 | 0.0022762 |
| 93 | LH_SalVentAttn_ParOper_2 | 0.04355 | 0.16372 | 0.12017 | 0.00129987 | 0.00672761 |
| 276 | RH_DorsAttn_Post_6 | 0.04080 | 0.16003 | 0.11923 | 0.0039996 | 0.0164448 |
| 35 | LH_SomMot_4 | −0.00228 | 0.11654 | 0.11882 | 0.00639936 | 0.024019 |
| 81 | LH_DorsAttn_Post_13 | 0.08800 | 0.20646 | 0.11846 | 9.999e−5 | 0.0022762 |
| 311 | RH_SalVentAttn_Med_1 | 0.01302 | 0.12962 | 0.11661 | 9.999e−5 | 0.0022762 |
| 127 | LH_Cont_Par_1 | 0.08220 | 0.19766 | 0.11546 | 0.00419958 | 0.0170418 |
| 173 | LH_Default_PFC_8 | 0.12224 | 0.23641 | 0.11417 | 0.00039996 | 0.00324605 |
| 318 | RH_SalVentAttn_Med_8 | 0.00318 | 0.11449 | 0.11131 | 0.00569943 | 0.0218309 |
| 179 | LH_Default_PFC_14 | 0.09307 | 0.20404 | 0.11098 | 0.00049995 | 0.003535 |
| 305 | RH_SalVentAttn_FrOper_4 | 0.00301 | 0.11324 | 0.11022 | 0.010399 | 0.0353363 |
| 148 | LH_Cont_PFCmp_1 | 0.01518 | 0.12478 | 0.10960 | 0.0121988 | 0.0396708 |
| 137 | LH_Cont_PFCl_4 | 0.09024 | 0.19981 | 0.10957 | 0.0019998 | 0.00944256 |
| 334 | RH_Cont_Par_3 | 0.08139 | 0.19059 | 0.10920 | 0.00359964 | 0.0152944 |
| 326 | RH_Limbic_TempPole_2 | 0.00396 | 0.11265 | 0.10869 | 0.0141986 | 0.0452802 |
| 36 | LH_SomMot_5 | 0.02591 | 0.13246 | 0.10655 | 0.00419958 | 0.0170418 |
| 400 | RH_Default_PCC_9 | 0.08464 | 0.19079 | 0.10615 | 0.00049995 | 0.003535 |
| 142 | LH_Cont_PFCl_9 | 0.03756 | 0.14214 | 0.10458 | 0.00529947 | 0.0205805 |
| 340 | RH_Cont_PFCv_1 | 0.05840 | 0.15871 | 0.10031 | 0.00389961 | 0.0161762 |
| 216 | RH_Vis_16 | 0.06103 | 0.15855 | 0.09752 | 0.00359964 | 0.0152944 |
| 95 | LH_SalVentAttn_ParOper_4 | 0.01767 | 0.11478 | 0.09712 | 0.00419958 | 0.0170418 |
| 170 | LH_Default_PFC_5 | 0.06118 | 0.15788 | 0.09670 | 0.00049995 | 0.003535 |
| 396 | RH_Default_PCC_5 | 0.09685 | 0.19138 | 0.09454 | 0.00349965 | 0.0149375 |
| 195 | LH_Default_PCC_6 | 0.08395 | 0.17383 | 0.08988 | 0.00369963 | 0.015554 |
| 41 | LH_SomMot_10 | 0.01513 | 0.10483 | 0.08971 | 0.00549945 | 0.021298 |
| 222 | RH_Vis_22 | 0.07817 | 0.16702 | 0.08886 | 0.0049995 | 0.0196059 |
| 177 | LH_Default_PFC_12 | 0.07017 | 0.15449 | 0.08432 | 0.00409959 | 0.0167574 |
| 377 | RH_Default_PFCv_3 | 0.05665 | 0.13869 | 0.08204 | 0.00259974 | 0.0115728 |
| 145 | LH_Cont_pCun_2 | 0.09186 | 0.16963 | 0.07777 | 0.00879912 | 0.0308741 |
| 357 | RH_Cont_pCun_2 | 0.08256 | 0.15998 | 0.07742 | 0.0059994 | 0.0228549 |
| 285 | RH_DorsAttn_Post_15 | 0.10258 | 0.17836 | 0.07578 | 0.0153985 | 0.048174 |
| 196 | LH_Default_PCC_7 | 0.05709 | 0.13275 | 0.07566 | 0.0119988 | 0.0394327 |
| 197 | LH_Default_PCC_8 | 0.06974 | 0.14523 | 0.07549 | 0.0156984 | 0.0488939 |
| 144 | LH_Cont_pCun_1 | 0.10018 | 0.17508 | 0.07490 | 0.0146985 | 0.046662 |
| 308 | RH_SalVentAttn_FrOper_7 | 0.00318 | 0.07249 | 0.06932 | 0.0136986 | 0.0438858 |
| 82 | LH_DorsAttn_Post_14 | 0.08886 | 0.15504 | 0.06618 | 0.0120988 | 0.0394832 |
| 382 | RH_Default_PFCm_4 | 0.10946 | 0.17221 | 0.06275 | 0.00719928 | 0.0262132 |
| 200 | LH_Default_PCC_11 | 0.07567 | 0.13619 | 0.06052 | 0.00559944 | 0.0216254 |
| 381 | RH_Default_PFCm_3 | 0.10585 | 0.16476 | 0.05890 | 0.00429957 | 0.017347 |
| 29 | LH_Vis_29 | 0.10561 | 0.16276 | 0.05715 | 0.0151985 | 0.0478154 |
| 107 | LH_SalVentAttn_Med_1 | 0.01643 | 0.05657 | 0.04014 | 0.0049995 | 0.0196059 |

Table S9. Continued. F. Significant regional gains of receptor-informed CAP depth-profile models.

| **CAP-6: 46 / 400 significant parcels (11.50%)** | | | | | | |
| --- | --- | --- | --- | --- | --- | --- |
| **ROI ID** | **Atlas parcel** | **Baseline adjusted R²** | **Full model adjusted R²** | **Gain (Δadj. R²)** | **Spin p** | **FDR q** |
| 296 | RH_SalVentAttn_TempOccPar_3 | −0.00007 | 0.19921 | 0.19928 | 0.00039996 | 0.00324605 |
| 366 | RH_Default_Par_5 | 0.00014 | 0.19106 | 0.19092 | 0.00039996 | 0.00324605 |
| 239 | RH_SomMot_9 | 0.00143 | 0.18985 | 0.18842 | 9.999e−5 | 0.0022762 |
| 265 | RH_SomMot_35 | 0.01261 | 0.19917 | 0.18655 | 0.00019998 | 0.00324605 |
| 257 | RH_SomMot_27 | 0.02953 | 0.16809 | 0.13856 | 0.00139986 | 0.00702439 |
| 224 | RH_Vis_24 | 0.00844 | 0.14658 | 0.13813 | 0.00459954 | 0.0183982 |
| 280 | RH_DorsAttn_Post_10 | 0.00198 | 0.13144 | 0.12946 | 0.00039996 | 0.00324605 |
| 388 | RH_Default_PFCm_10 | −0.00190 | 0.12671 | 0.12862 | 9.999e−5 | 0.0022762 |
| 15 | LH_Vis_15 | 0.05630 | 0.17037 | 0.11407 | 0.00379962 | 0.0158553 |
| 248 | RH_SomMot_18 | 0.00046 | 0.11359 | 0.11314 | 9.999e−5 | 0.0022762 |
| 64 | LH_SomMot_33 | 0.07658 | 0.18955 | 0.11296 | 0.00269973 | 0.0119231 |
| 262 | RH_SomMot_32 | −0.00247 | 0.11018 | 0.11265 | 0.00039996 | 0.00324605 |
| 48 | LH_SomMot_17 | 0.02587 | 0.13428 | 0.10841 | 0.00029997 | 0.00324605 |
| 36 | LH_SomMot_5 | 0.00002 | 0.10647 | 0.10645 | 0.00039996 | 0.00324605 |
| 251 | RH_SomMot_21 | 0.03295 | 0.13839 | 0.10543 | 0.00619938 | 0.0235526 |
| 47 | LH_SomMot_16 | 0.01730 | 0.12125 | 0.10395 | 0.00049995 | 0.003535 |
| 273 | RH_DorsAttn_Post_3 | 0.00625 | 0.10912 | 0.10287 | 0.00689931 | 0.0254856 |
| 43 | LH_SomMot_12 | 0.01457 | 0.11557 | 0.10100 | 0.00649935 | 0.0242642 |
| 161 | LH_Default_Temp_13 | −0.00195 | 0.09778 | 0.09973 | 0.00069993 | 0.00439418 |
| 175 | LH_Default_PFC_10 | 0.00101 | 0.09831 | 0.09730 | 0.00349965 | 0.0149375 |
| 193 | LH_Default_PCC_4 | 0.00060 | 0.09118 | 0.09058 | 0.0113989 | 0.0379962 |
| 50 | LH_SomMot_19 | 0.00334 | 0.09384 | 0.09050 | 9.999e−5 | 0.0022762 |
| 235 | RH_SomMot_5 | 0.00587 | 0.09486 | 0.08898 | 0.00059994 | 0.00398064 |
| 16 | LH_Vis_16 | 0.04116 | 0.13008 | 0.08892 | 0.0126987 | 0.0412488 |
| 252 | RH_SomMot_22 | 0.04912 | 0.13618 | 0.08706 | 0.00639936 | 0.024019 |
| 343 | RH_Cont_PFCl_3 | 0.01537 | 0.10184 | 0.08647 | 0.0137986 | 0.0441052 |
| 289 | RH_DorsAttn_Post_19 | 0.03748 | 0.12360 | 0.08613 | 0.00119988 | 0.00631516 |
| 351 | RH_Cont_PFCl_11 | 0.06222 | 0.14695 | 0.08473 | 0.0089991 | 0.0313792 |
| 129 | LH_Cont_Par_3 | −0.00191 | 0.08261 | 0.08452 | 0.00569943 | 0.0218309 |
| 62 | LH_SomMot_31 | −0.00103 | 0.07995 | 0.08097 | 0.00049995 | 0.003535 |
| 340 | RH_Cont_PFCv_1 | −0.00220 | 0.07744 | 0.07964 | 0.00849915 | 0.0298965 |
| 197 | LH_Default_PCC_8 | 0.02946 | 0.10846 | 0.07899 | 0.00929907 | 0.0322245 |
| 352 | RH_Cont_PFCl_12 | 0.07657 | 0.15541 | 0.07884 | 0.009999 | 0.0342683 |
| 80 | LH_DorsAttn_Post_12 | 0.01231 | 0.08543 | 0.07312 | 0.00019998 | 0.00324605 |
| 132 | LH_Cont_Par_6 | 0.00364 | 0.07527 | 0.07163 | 0.0133987 | 0.0429739 |
| 393 | RH_Default_PCC_2 | −0.00160 | 0.06650 | 0.06810 | 0.0154985 | 0.0484327 |
| 39 | LH_SomMot_8 | −0.00195 | 0.06551 | 0.06745 | 0.0109989 | 0.0368825 |
| 146 | LH_Cont_Cing_1 | 0.05697 | 0.12361 | 0.06665 | 0.00189981 | 0.00904671 |
| 145 | LH_Cont_pCun_2 | −0.00010 | 0.06540 | 0.06550 | 0.0119988 | 0.0394327 |
| 333 | RH_Cont_Par_2 | 0.01501 | 0.08045 | 0.06544 | 0.0107989 | 0.0364301 |
| 359 | RH_Cont_Cing_2 | 0.10486 | 0.16228 | 0.05742 | 0.0109989 | 0.0368825 |
| 400 | RH_Default_PCC_9 | −0.00174 | 0.05198 | 0.05371 | 0.0089991 | 0.0313792 |
| 238 | RH_SomMot_8 | −0.00247 | 0.05002 | 0.05250 | 0.00149985 | 0.00740667 |
| 63 | LH_SomMot_32 | 0.04599 | 0.09225 | 0.04626 | 0.00939906 | 0.0325307 |
| 35 | LH_SomMot_4 | 0.00440 | 0.04276 | 0.03835 | 0.0069993 | 0.0256855 |
| 316 | RH_SalVentAttn_Med_6 | 0.00152 | 0.03767 | 0.03615 | 0.00519948 | 0.0202765 |

Table S9. Continued. G. Significant regional gains of receptor-informed CAP depth-profile models.

| **CAP-7: 152 / 400 significant parcels (38.00%)** | | | | | | |
| --- | --- | --- | --- | --- | --- | --- |
| **ROI ID** | **Atlas parcel** | **Baseline adjusted R²** | **Full model adjusted R²** | **Gain (Δadj. R²)** | **Spin p** | **FDR q** |
| 62 | LH_SomMot_31 | 0.00186 | 0.28183 | 0.27997 | 0.00039996 | 0.00324605 |
| 270 | RH_SomMot_40 | 0.00696 | 0.28342 | 0.27646 | 9.999e−5 | 0.0022762 |
| 64 | LH_SomMot_33 | 0.00143 | 0.27403 | 0.27259 | 0.00039996 | 0.00324605 |
| 263 | RH_SomMot_33 | −0.00057 | 0.26524 | 0.26582 | 9.999e−5 | 0.0022762 |
| 268 | RH_SomMot_38 | 0.00156 | 0.26057 | 0.25900 | 9.999e−5 | 0.0022762 |
| 259 | RH_SomMot_29 | 0.01843 | 0.26774 | 0.24930 | 0.00039996 | 0.00324605 |
| 256 | RH_SomMot_26 | −0.00242 | 0.24002 | 0.24245 | 0.00039996 | 0.00324605 |
| 262 | RH_SomMot_32 | 0.00338 | 0.24267 | 0.23929 | 0.00039996 | 0.00324605 |
| 68 | LH_SomMot_37 | 0.00462 | 0.23122 | 0.22660 | 0.00039996 | 0.00324605 |
| 59 | LH_SomMot_28 | −0.00049 | 0.22558 | 0.22608 | 0.00039996 | 0.00324605 |
| 51 | LH_SomMot_20 | −0.00179 | 0.22268 | 0.22447 | 0.00039996 | 0.00324605 |
| 257 | RH_SomMot_27 | −0.00032 | 0.22147 | 0.22179 | 0.00039996 | 0.00324605 |
| 53 | LH_SomMot_22 | −0.00147 | 0.21992 | 0.22139 | 9.999e−5 | 0.0022762 |
| 269 | RH_SomMot_39 | 0.00531 | 0.21735 | 0.21204 | 0.00039996 | 0.00324605 |
| 251 | RH_SomMot_21 | 0.00885 | 0.21806 | 0.20920 | 0.00049995 | 0.003535 |
| 255 | RH_SomMot_25 | 0.01439 | 0.21732 | 0.20293 | 0.00039996 | 0.00324605 |
| 265 | RH_SomMot_35 | 0.00014 | 0.20023 | 0.20009 | 0.00049995 | 0.003535 |
| 60 | LH_SomMot_29 | −0.00029 | 0.18934 | 0.18963 | 0.0009999 | 0.00561066 |
| 50 | LH_SomMot_19 | 0.00260 | 0.19213 | 0.18954 | 0.00039996 | 0.00324605 |
| 57 | LH_SomMot_26 | 0.00827 | 0.19446 | 0.18619 | 0.00039996 | 0.00324605 |
| 239 | RH_SomMot_9 | 0.00057 | 0.18632 | 0.18574 | 9.999e−5 | 0.0022762 |
| 252 | RH_SomMot_22 | 0.00806 | 0.19243 | 0.18438 | 9.999e−5 | 0.0022762 |
| 354 | RH_Cont_PFCl_14 | 0.04798 | 0.23054 | 0.18256 | 0.00039996 | 0.00324605 |
| 254 | RH_SomMot_24 | 0.01244 | 0.19021 | 0.17777 | 9.999e−5 | 0.0022762 |
| 365 | RH_Default_Par_4 | 0.00760 | 0.18462 | 0.17702 | 0.00089991 | 0.00521687 |
| 63 | LH_SomMot_32 | −0.00033 | 0.17132 | 0.17165 | 0.00079992 | 0.00479609 |
| 171 | LH_Default_PFC_6 | −0.00249 | 0.16879 | 0.17127 | 0.00039996 | 0.00324605 |
| 384 | RH_Default_PFCm_6 | −0.00248 | 0.16472 | 0.16720 | 9.999e−5 | 0.0022762 |
| 236 | RH_SomMot_6 | 0.00097 | 0.16439 | 0.16341 | 9.999e−5 | 0.0022762 |
| 36 | LH_SomMot_5 | 0.00254 | 0.16214 | 0.15960 | 0.00039996 | 0.00324605 |
| 264 | RH_SomMot_34 | 0.01877 | 0.17706 | 0.15828 | 0.00269973 | 0.0119231 |
| 343 | RH_Cont_PFCl_3 | 0.01286 | 0.17112 | 0.15826 | 0.00039996 | 0.00324605 |
| 132 | LH_Cont_Par_6 | −0.00244 | 0.15542 | 0.15787 | 0.00119988 | 0.00631516 |
| 49 | LH_SomMot_18 | 0.01419 | 0.17017 | 0.15598 | 0.00039996 | 0.00324605 |
| 224 | RH_Vis_24 | 0.00838 | 0.16256 | 0.15418 | 0.00369963 | 0.015554 |
| 324 | RH_Limbic_OFC_6 | −0.00093 | 0.15320 | 0.15413 | 0.00109989 | 0.0059684 |
| 321 | RH_Limbic_OFC_3 | −0.00001 | 0.15272 | 0.15274 | 0.00179982 | 0.00859982 |
| 363 | RH_Default_Par_2 | 0.01043 | 0.16172 | 0.15129 | 0.00039996 | 0.00324605 |
| 276 | RH_DorsAttn_Post_6 | 0.02488 | 0.17459 | 0.14972 | 0.00019998 | 0.00324605 |
| 366 | RH_Default_Par_5 | 0.00457 | 0.14927 | 0.14470 | 9.999e−5 | 0.0022762 |
| 142 | LH_Cont_PFCl_9 | 0.01165 | 0.15566 | 0.14401 | 0.00049995 | 0.003535 |
| 95 | LH_SalVentAttn_ParOper_4 | −0.00246 | 0.13864 | 0.14110 | 0.0009999 | 0.00561066 |
| 273 | RH_DorsAttn_Post_3 | 0.01281 | 0.15179 | 0.13898 | 0.00039996 | 0.00324605 |
| 56 | LH_SomMot_25 | 0.02298 | 0.16146 | 0.13848 | 9.999e−5 | 0.0022762 |
| 129 | LH_Cont_Par_3 | −0.00219 | 0.13548 | 0.13767 | 0.00049995 | 0.003535 |
| 137 | LH_Cont_PFCl_4 | 0.00443 | 0.14146 | 0.13702 | 0.00139986 | 0.00702439 |
| 146 | LH_Cont_Cing_1 | 0.00091 | 0.13549 | 0.13458 | 0.00039996 | 0.00324605 |
| 336 | RH_Cont_Par_5 | 0.01451 | 0.14797 | 0.13346 | 0.00129987 | 0.00672761 |
| 48 | LH_SomMot_17 | −0.00067 | 0.13199 | 0.13266 | 0.00259974 | 0.0115728 |
| 278 | RH_DorsAttn_Post_8 | 0.01811 | 0.15013 | 0.13203 | 0.00089991 | 0.00521687 |
| 345 | RH_Cont_PFCl_5 | 0.03263 | 0.16178 | 0.12916 | 0.00139986 | 0.00702439 |
| 337 | RH_Cont_Par_6 | 0.01090 | 0.13908 | 0.12818 | 0.00229977 | 0.010626 |
| 333 | RH_Cont_Par_2 | 0.01312 | 0.14113 | 0.12801 | 0.00049995 | 0.003535 |
| 352 | RH_Cont_PFCl_12 | 0.02841 | 0.15579 | 0.12738 | 0.00039996 | 0.00324605 |
| 296 | RH_SalVentAttn_TempOccPar_3 | 0.00850 | 0.13405 | 0.12555 | 0.00049995 | 0.003535 |
| 362 | RH_Default_Par_1 | 0.00635 | 0.13190 | 0.12555 | 0.00109989 | 0.0059684 |
| 310 | RH_SalVentAttn_PFCl_1 | 0.03867 | 0.16214 | 0.12347 | 0.00159984 | 0.00783138 |
| 66 | LH_SomMot_35 | −0.00250 | 0.11760 | 0.12009 | 0.00649935 | 0.0242642 |
| 15 | LH_Vis_15 | 0.06186 | 0.18193 | 0.12006 | 0.00809919 | 0.0288521 |
| 197 | LH_Default_PCC_8 | 0.00242 | 0.12013 | 0.11771 | 0.00229977 | 0.010626 |
| 332 | RH_Cont_Par_1 | 0.01000 | 0.12767 | 0.11768 | 0.0009999 | 0.00561066 |
| 253 | RH_SomMot_23 | 0.02558 | 0.14253 | 0.11695 | 0.00529947 | 0.0205805 |
| 355 | RH_Cont_PFCl_15 | 0.08506 | 0.20197 | 0.11691 | 0.00069993 | 0.00439418 |
| 139 | LH_Cont_PFCl_6 | 0.00898 | 0.12583 | 0.11685 | 0.00479952 | 0.019008 |
| 340 | RH_Cont_PFCv_1 | −0.00030 | 0.11497 | 0.11526 | 0.00169983 | 0.00819195 |
| 55 | LH_SomMot_24 | 0.03111 | 0.14431 | 0.11320 | 0.00019998 | 0.00324605 |
| 89 | LH_DorsAttn_FEF_4 | 0.07471 | 0.18757 | 0.11286 | 0.00239976 | 0.0109793 |
| 334 | RH_Cont_Par_3 | 0.00195 | 0.11401 | 0.11206 | 0.00079992 | 0.00479609 |
| 180 | LH_Default_PFC_15 | 0.00747 | 0.11950 | 0.11203 | 0.00719928 | 0.0262132 |
| 161 | LH_Default_Temp_13 | −0.00099 | 0.11034 | 0.11133 | 0.00129987 | 0.00672761 |
| 67 | LH_SomMot_36 | 0.00194 | 0.11313 | 0.11120 | 0.00269973 | 0.0119231 |
| 130 | LH_Cont_Par_4 | −0.00250 | 0.10763 | 0.11013 | 0.00759924 | 0.0273495 |
| 184 | LH_Default_PFC_19 | 0.03464 | 0.14451 | 0.10987 | 0.00789921 | 0.0282475 |
| 359 | RH_Cont_Cing_2 | 0.02643 | 0.13595 | 0.10953 | 0.00059994 | 0.00398064 |
| 173 | LH_Default_PFC_8 | 0.00575 | 0.11519 | 0.10944 | 0.00759924 | 0.0273495 |
| 386 | RH_Default_PFCm_8 | 0.03222 | 0.14060 | 0.10838 | 0.010399 | 0.0353363 |
| 12 | LH_Vis_12 | 0.06968 | 0.17490 | 0.10523 | 0.00979902 | 0.0336653 |
| 351 | RH_Cont_PFCl_11 | 0.02086 | 0.12590 | 0.10504 | 0.00119988 | 0.00631516 |
| 192 | LH_Default_PCC_3 | 0.01091 | 0.11459 | 0.10368 | 0.0105989 | 0.0358418 |
| 240 | RH_SomMot_10 | −0.00055 | 0.10293 | 0.10348 | 0.00039996 | 0.00324605 |
| 133 | LH_Cont_Temp_1 | 0.01831 | 0.12089 | 0.10258 | 0.00149985 | 0.00740667 |
| 138 | LH_Cont_PFCl_5 | −0.00047 | 0.10200 | 0.10247 | 0.00079992 | 0.00479609 |
| 260 | RH_SomMot_30 | 0.02603 | 0.12837 | 0.10235 | 0.00039996 | 0.00324605 |
| 388 | RH_Default_PFCm_10 | −0.00174 | 0.10048 | 0.10223 | 0.00129987 | 0.00672761 |
| 316 | RH_SalVentAttn_Med_6 | −0.00224 | 0.09978 | 0.10202 | 0.00039996 | 0.00324605 |
| 135 | LH_Cont_PFCl_2 | −0.00162 | 0.10006 | 0.10167 | 0.00289971 | 0.0126664 |
| 170 | LH_Default_PFC_5 | −0.00251 | 0.09862 | 0.10114 | 0.00409959 | 0.0167574 |
| 84 | LH_DorsAttn_Post_16 | 0.00964 | 0.10945 | 0.09981 | 0.00109989 | 0.0059684 |
| 274 | RH_DorsAttn_Post_4 | −0.00031 | 0.09924 | 0.09955 | 0.00169983 | 0.00819195 |
| 344 | RH_Cont_PFCl_4 | 0.02740 | 0.12594 | 0.09853 | 0.00679932 | 0.0251827 |
| 294 | RH_SalVentAttn_TempOccPar_1 | 0.00846 | 0.10630 | 0.09784 | 0.00089991 | 0.00521687 |
| 248 | RH_SomMot_18 | −0.00250 | 0.09501 | 0.09751 | 0.00039996 | 0.00324605 |
| 94 | LH_SalVentAttn_ParOper_3 | −0.00210 | 0.09518 | 0.09729 | 0.00629937 | 0.0238033 |
| 349 | RH_Cont_PFCl_9 | 0.04507 | 0.14219 | 0.09712 | 0.00809919 | 0.0288521 |
| 285 | RH_DorsAttn_Post_15 | 0.00133 | 0.09801 | 0.09667 | 0.00209979 | 0.00986479 |
| 35 | LH_SomMot_4 | 0.00585 | 0.10211 | 0.09626 | 9.999e−5 | 0.0022762 |
| 280 | RH_DorsAttn_Post_10 | 0.00472 | 0.10071 | 0.09599 | 0.0019998 | 0.00944256 |
| 82 | LH_DorsAttn_Post_14 | 0.00209 | 0.09580 | 0.09371 | 0.00039996 | 0.00324605 |
| 74 | LH_DorsAttn_Post_6 | −0.00196 | 0.09081 | 0.09278 | 0.0115988 | 0.038571 |
| 196 | LH_Default_PCC_7 | −0.00234 | 0.09000 | 0.09235 | 0.00779922 | 0.0279972 |
| 400 | RH_Default_PCC_9 | −0.00059 | 0.09169 | 0.09229 | 0.00139986 | 0.00702439 |
| 131 | LH_Cont_Par_5 | −0.00109 | 0.09104 | 0.09213 | 0.00779922 | 0.0279972 |
| 118 | LH_Limbic_OFC_5 | −0.00086 | 0.09119 | 0.09204 | 0.0109989 | 0.0368825 |
| 6 | LH_Vis_6 | 0.04720 | 0.13894 | 0.09174 | 0.0116988 | 0.0388113 |
| 249 | RH_SomMot_19 | 0.00165 | 0.09323 | 0.09158 | 0.00249975 | 0.0112529 |
| 179 | LH_Default_PFC_14 | 0.01297 | 0.10446 | 0.09149 | 0.0149985 | 0.0473993 |
| 128 | LH_Cont_Par_2 | −0.00180 | 0.08966 | 0.09147 | 0.00229977 | 0.010626 |
| 165 | LH_Default_Temp_17 | −0.00107 | 0.08926 | 0.09032 | 0.0029997 | 0.0130422 |
| 47 | LH_SomMot_16 | 0.00888 | 0.09903 | 0.09015 | 0.00059994 | 0.00398064 |
| 390 | RH_Default_PFCm_12 | 0.00845 | 0.09853 | 0.09008 | 0.00239976 | 0.0109793 |
| 282 | RH_DorsAttn_Post_12 | 0.00254 | 0.09249 | 0.08995 | 0.010399 | 0.0353363 |
| 387 | RH_Default_PFCm_9 | 0.01793 | 0.10782 | 0.08989 | 0.0137986 | 0.0441052 |
| 160 | LH_Default_Temp_12 | −0.00153 | 0.08720 | 0.08874 | 0.00489951 | 0.0193493 |
| 288 | RH_DorsAttn_Post_18 | 0.00996 | 0.09854 | 0.08859 | 0.0049995 | 0.0196059 |
| 81 | LH_DorsAttn_Post_13 | −0.00097 | 0.08645 | 0.08742 | 0.0039996 | 0.0164448 |
| 32 | LH_SomMot_1 | −0.00244 | 0.08435 | 0.08679 | 0.00739926 | 0.026802 |
| 183 | LH_Default_PFC_18 | 0.02829 | 0.11413 | 0.08584 | 0.0119988 | 0.0394327 |
| 272 | RH_DorsAttn_Post_2 | −0.00178 | 0.08277 | 0.08455 | 0.00539946 | 0.0209397 |
| 71 | LH_DorsAttn_Post_3 | 0.01894 | 0.10157 | 0.08263 | 0.00729927 | 0.0265084 |
| 96 | LH_SalVentAttn_TempOcc_1 | 0.00065 | 0.08318 | 0.08252 | 0.00349965 | 0.0149375 |
| 214 | RH_Vis_14 | −0.00241 | 0.07944 | 0.08185 | 0.00389961 | 0.0161762 |
| 298 | RH_SalVentAttn_TempOccPar_5 | 0.01827 | 0.09752 | 0.07925 | 0.0039996 | 0.0164448 |
| 233 | RH_SomMot_3 | −0.00204 | 0.07553 | 0.07757 | 0.00679932 | 0.0251827 |
| 145 | LH_Cont_pCun_2 | −0.00140 | 0.07449 | 0.07589 | 0.00449955 | 0.0181277 |
| 187 | LH_Default_PFC_22 | 0.01108 | 0.08629 | 0.07521 | 0.00709929 | 0.0259504 |
| 341 | RH_Cont_PFCl_1 | 0.00528 | 0.07991 | 0.07462 | 0.00419958 | 0.0170418 |
| 300 | RH_SalVentAttn_TempOccPar_7 | 0.01688 | 0.09130 | 0.07441 | 0.0120988 | 0.0394832 |
| 20 | LH_Vis_20 | 0.02410 | 0.09701 | 0.07291 | 0.00969903 | 0.0333628 |
| 39 | LH_SomMot_8 | −0.00015 | 0.07255 | 0.07270 | 0.00509949 | 0.0199143 |
| 286 | RH_DorsAttn_Post_16 | 0.01425 | 0.08614 | 0.07189 | 0.00349965 | 0.0149375 |
| 371 | RH_Default_Temp_5 | 0.00734 | 0.07749 | 0.07015 | 0.0029997 | 0.0130422 |
| 80 | LH_DorsAttn_Post_12 | −0.00073 | 0.06878 | 0.06951 | 0.00739926 | 0.026802 |
| 318 | RH_SalVentAttn_Med_8 | 0.10359 | 0.17211 | 0.06852 | 0.0059994 | 0.0228549 |
| 175 | LH_Default_PFC_10 | −0.00126 | 0.06721 | 0.06847 | 0.0141986 | 0.0452802 |
| 116 | LH_Limbic_OFC_3 | −0.00250 | 0.06420 | 0.06670 | 0.00849915 | 0.0298965 |
| 241 | RH_SomMot_11 | 0.00261 | 0.06875 | 0.06614 | 0.00669933 | 0.0248781 |
| 364 | RH_Default_Par_3 | 0.00521 | 0.07122 | 0.06601 | 0.00629937 | 0.0238033 |
| 399 | RH_Default_PCC_8 | −0.00095 | 0.06460 | 0.06555 | 0.0129987 | 0.041883 |
| 358 | RH_Cont_Cing_1 | 0.00791 | 0.07320 | 0.06529 | 0.0117988 | 0.0390505 |
| 181 | LH_Default_PFC_16 | 0.01328 | 0.07814 | 0.06485 | 0.0157984 | 0.0491506 |
| 261 | RH_SomMot_31 | 0.00729 | 0.07070 | 0.06341 | 0.00109989 | 0.0059684 |
| 200 | LH_Default_PCC_11 | −0.00248 | 0.06076 | 0.06323 | 0.00689931 | 0.0254856 |
| 234 | RH_SomMot_4 | 0.00480 | 0.06698 | 0.06218 | 0.009999 | 0.0342683 |
| 69 | LH_DorsAttn_Post_1 | −0.00243 | 0.05882 | 0.06125 | 0.0120988 | 0.0394832 |
| 373 | RH_Default_Temp_7 | 0.00966 | 0.07037 | 0.06072 | 0.00389961 | 0.0161762 |
| 164 | LH_Default_Temp_16 | −0.00244 | 0.05647 | 0.05891 | 0.0150985 | 0.0476079 |
| 65 | LH_SomMot_34 | 0.03118 | 0.08838 | 0.05720 | 0.0009999 | 0.00561066 |
| 61 | LH_SomMot_30 | 0.03325 | 0.08665 | 0.05340 | 0.00259974 | 0.0115728 |
| 277 | RH_DorsAttn_Post_7 | 0.00617 | 0.05880 | 0.05263 | 0.00069993 | 0.00439418 |
| 235 | RH_SomMot_5 | 0.00132 | 0.05014 | 0.04883 | 0.00269973 | 0.0119231 |
| 34 | LH_SomMot_3 | −0.00245 | 0.03802 | 0.04047 | 0.00809919 | 0.0288521 |
| 258 | RH_SomMot_28 | −0.00243 | 0.03514 | 0.03757 | 0.00839916 | 0.0296566 |

Regional results underlying Figure 3 are listed for CAP-1–CAP-7. For each CAP, the distance-only baseline and the distance-plus-receptor model were used to explain the seed parcel’s CAP depth-profile similarity to all 400 cortical parcels, using normative shared GM–WM activation maps across WM depths of 1–7 mm. The additional predictor was similarity of the 19-receptor/transporter fingerprints.

Gain was defined as Δadjusted R² = full-model adjusted R² − baseline adjusted R². Included parcels had positive gains and BH-FDR q < 0.05 across all 2,800 CAP-by-parcel tests, based on 10,000 one-sided spatial-spin permutations. All 900 significant CAP–parcel entries are reported; a parcel may be significant for more than one CAP. Entries are sorted by decreasing gain within each CAP.

Parcel IDs and labels follow the Schaefer 400-parcel, 7-network atlas used in the analysis. The common label prefix “7Networks_” is omitted. LH/RH, left/right hemisphere; Vis, visual; SomMot, somatomotor; DorsAttn, dorsal attention; SalVentAttn, salience/ventral attention; Limbic, limbic; Cont, control; Default, default-mode network. R² and gain are shown to five decimals; p and q are shown to six significant digits. Selection uses unrounded values.

Table S10. Yeo7 network distribution of significant receptor-model gains. Significant parcels were assigned to Yeo7 networks using their Schaefer 400-parcel atlas labels. Left and right hemispheres were pooled. The same positive-gain and BH-FDR q < 0.05 criteria as Figure 3 and Table S9 were used. A. Significant parcels by network, n (% of the CAP’s significant parcels).

| **Network** | **CAP-1** | **CAP-2** | **CAP-3** | **CAP-4** | **CAP-5** | **CAP-6** | **CAP-7** |
| --- | --- | --- | --- | --- | --- | --- | --- |
| VIS | 13 (10.3) | 7 (3.9) | 18 (9.2) | 0 (0.0) | 5 (2.7) | 3 (6.5) | 6 (3.9) |
| SMN | 33 (26.2) | 44 (24.7) | 47 (24.0) | 11 (73.3) | 54 (28.9) | 19 (41.3) | 51 (33.6) |
| DAN | 15 (11.9) | 17 (9.6) | 15 (7.7) | 0 (0.0) | 12 (6.4) | 4 (8.7) | 19 (12.5) |
| SVAN | 16 (12.7) | 26 (14.6) | 26 (13.3) | 0 (0.0) | 18 (9.6) | 2 (4.3) | 10 (6.6) |
| LIM | 0 (0.0) | 5 (2.8) | 4 (2.0) | 0 (0.0) | 8 (4.3) | 0 (0.0) | 4 (2.6) |
| CON | 31 (24.6) | 42 (23.6) | 42 (21.4) | 1 (6.7) | 35 (18.7) | 10 (21.7) | 30 (19.7) |
| DMN | 18 (14.3) | 37 (20.8) | 44 (22.4) | 3 (20.0) | 55 (29.4) | 8 (17.4) | 32 (21.1) |
| **Total** | **126 (100)** | **178 (100)** | **196 (100)** | **15 (100)** | **187 (100)** | **46 (100)** | **152 (100)** |

For each CAP, the denominator is its total number of significant parcels; network counts sum to the CAP total.

Table S10. Yeo7 network distribution of significant receptor-model gains. B. Significant parcels as a percentage of all parcels in each network (%).

| **Network (total N)** | **CAP-1** | **CAP-2** | **CAP-3** | **CAP-4** | **CAP-5** | **CAP-6** | **CAP-7** |
| --- | --- | --- | --- | --- | --- | --- | --- |
| VIS (61) | 21.3 | 11.5 | 29.5 | 0.0 | 8.2 | 4.9 | 9.8 |
| SMN (77) | 42.9 | 57.1 | 61.0 | 14.3 | 70.1 | 24.7 | 66.2 |
| DAN (46) | 32.6 | 37.0 | 32.6 | 0.0 | 26.1 | 8.7 | 41.3 |
| SVAN (47) | 34.0 | 55.3 | 55.3 | 0.0 | 38.3 | 4.3 | 21.3 |
| LIM (26) | 0.0 | 19.2 | 15.4 | 0.0 | 30.8 | 0.0 | 15.4 |
| CON (52) | 59.6 | 80.8 | 80.8 | 1.9 | 67.3 | 19.2 | 57.7 |
| DMN (91) | 19.8 | 40.7 | 48.4 | 3.3 | 60.4 | 8.8 | 35.2 |

The denominator is the total number of atlas parcels in the corresponding network, shown in parentheses in the first column. Network sizes sum to 400.

Notes. Panel A describes the network composition of significant model gains; Panel B describes the within-network significant fraction and accounts for unequal network sizes in its denominator. Percentages are rounded to one decimal place, so Panel A may not sum to exactly 100% after rounding. These are descriptive counts and proportions, not tests of network enrichment. Significance was determined from 10,000 one-sided spatial-spin permutations followed by BH-FDR correction across 7 CAPs × 400 parcels (2,800 tests).

VIS, visual; SMN, somatomotor; DAN, dorsal attention; SVAN, salience/ventral attention; LIM, limbic; CON, control; DMN, default mode; CAP, coactivation pattern.

Table S11. Significant receptor associations of normative shared CAP phenotypes.

| **Phenotype** | **CAP** | **Receptor / transporter** | **Spearman ρ** | **Spin p** | **q** |
| --- | --- | --- | --- | --- | --- |
| A | CAP-1 | MOR | 0.589 | 0.0011 | 0.0183 |
| A | CAP-2 | MOR | −0.576 | 0.0025 | 0.0314 |
| A | CAP-3 | 5HT2a | 0.376 | 0.0060 | 0.0469 |
| A | CAP-3 | NET | −0.418 | 1.00e−3 | 0.0183 |
| A | CAP-4 | 5HT2a | −0.519 | 4.00e−4 | 0.0133 |
| A | CAP-4 | 5HT4 | −0.482 | 4.00e−4 | 0.0133 |
| A | CAP-4 | 5HTT | 0.395 | 0.0017 | 0.0251 |
| A | CAP-4 | CB1 | −0.343 | 0.0038 | 0.0389 |
| A | CAP-4 | MOR | −0.502 | 6.00e−4 | 0.0160 |
| A | CAP-6 | 5HT2a | −0.470 | 4.00e−4 | 0.0133 |
| A | CAP-6 | 5HT4 | −0.438 | 0.0034 | 0.0377 |
| A | CAP-6 | CB1 | −0.415 | 0.0051 | 0.0452 |
| A | CAP-6 | MOR | −0.517 | 8.00e−4 | 0.0177 |
| A | CAP-6 | NET | 0.390 | 0.0058 | 0.0469 |
| A | CAP-7 | 5HT2a | 0.433 | 4.00e−4 | 0.0133 |
| A | CAP-7 | MOR | 0.472 | 0.0026 | 0.0314 |
| A | CAP-7 | NET | −0.388 | 0.0044 | 0.0418 |
| L | CAP-2 | 5HT2a | 0.424 | 0.0044 | 0.0488 |
| L | CAP-2 | 5HT4 | 0.424 | 0.0013 | 0.0432 |
| L | CAP-2 | M1 | 0.302 | 9.00e−4 | 0.0432 |
| L | CAP-2 | MOR | 0.402 | 0.0011 | 0.0432 |
| L | CAP-2 | mGluR5 | 0.368 | 0.0041 | 0.0488 |
| L | CAP-5 | 5HT2a | 0.353 | 0.0026 | 0.0473 |
| L | CAP-5 | 5HT4 | 0.382 | 1.00e−3 | 0.0432 |
| L | CAP-5 | CB1 | 0.301 | 0.0032 | 0.0473 |
| L | CAP-5 | MOR | 0.357 | 0.0040 | 0.0488 |
| L | CAP-7 | 5HT4 | −0.317 | 0.0024 | 0.0473 |
| L | CAP-7 | M1 | −0.259 | 0.0030 | 0.0473 |
| L | CAP-7 | mGluR5 | −0.291 | 0.0030 | 0.0473 |

Notes. All 29 significant associations in Figure 4A–B are listed (17 for A; 12 for L). A is across-depth mean shared GM–WM activation and L is the linear depth slope, estimated from covariate-adjusted NC maps. Spearman correlations were evaluated across 400 parcels using 10,000 spatial-spin permutations and two-sided tests. BH-FDR correction was applied across 7 CAPs × 19 receptors (133 tests) separately for A and L. Receptor maps are independent normative atlas measurements. Full results for all 266 A/L associations and 931 depth-specific associations, including nonsignificant tests, are supplied in Normative_receptor_AL.csv and Normative_receptor_depth.csv.

Table S12. Globally significant PLS components of normative shared CAP organization.

| **Target** | **PLS** | **ΔY (%)** | **Spin p** | **Global q** | **Local n** | **CV R² cumulative** | **ΔCV R²** | **Figure** |
| --- | --- | --- | --- | --- | --- | --- | --- | --- |
| CAP-4 / A | 1 | 29.92 | 4.00e−4 | 0.0076 | 69 | 0.245 | 0.245 | Fig. 4C |
| CAP-4 / A | 2 | 13.18 | 0.0049 | 0.0465 | 5 | 0.351 | 0.106 | — |
| CAP-5 / A | 6 | 2.39 | 0.0057 | 0.0475 | 56 | 0.227 | −0.030 | — |
| CAP-5 / A | 7 | 2.41 | 0.0016 | 0.0304 | 0 | 0.228 | 0.000 | — |
| CAP-5 / A | 13 | 0.01 | 0.0075 | 0.0475 | 0 | 0.255 | 0.002 | — |
| CAP-6 / A | 1 | 26.63 | 0.0013 | 0.0247 | 24 | 0.191 | 0.191 | Fig. 4E |
| CAP-1 / L | 2 | 9.72 | 0.0022 | 0.0418 | 0 | 0.087 | 0.078 | Fig. S17A |
| CAP-2 / L | 1 | 20.96 | 0.0021 | 0.0399 | 0 | 0.179 | 0.179 | Fig. 4D |
| CAP-2 / L | 6 | 2.04 | 0.0052 | 0.0494 | 0 | 0.303 | 0.018 | — |
| CAP-5 / L | 1 | 16.60 | 0.0026 | 0.0494 | 0 | 0.094 | 0.094 | Fig. 4F |
| CAP-2 / 1 mm | 1 | 32.02 | 0.0011 | 0.0209 | 21 | 0.274 | 0.274 | — |
| CAP-3 / 1 mm | 1 | 20.83 | 8.00e−4 | 0.0152 | 38 | 0.071 | 0.071 | Fig. S17B |
| CAP-5 / 1 mm | 6 | 3.01 | 8.00e−4 | 0.0152 | 0 | 0.086 | −0.007 | — |
| CAP-7 / 1 mm | 1 | 25.23 | 0.0012 | 0.0228 | 34 | 0.175 | 0.175 | Fig. S17C |

Notes. All 14 components meeting global q < 0.05 among the 399 tested components (7 CAPs × 3 targets × 19 components) are included. A, across-depth mean; L, linear depth slope; 1 mm, activation at 1-mm WM depth. ΔY is the incremental percentage of target variance explained. Global q is the BH-adjusted spatial-null p value across 19 components within each CAP–target model, not across all 399 tests. Local n is the number of significant neighborhood centers out of 400 after within-component FDR correction; zero does not negate global significance. CV uses five spatial folds. Cumulative CV R² evaluates all components up to the listed one; ΔCV R² is its increment over the preceding model and can be negative despite global significance. A dash in the Figure column indicates that the component is not shown in the main or supplementary figures. All 399 component statistics, including nonsignificant results and bootstrap summaries, are supplied in PLS_all_components.csv.

Table S13. Significant co-occurrence cross-tissue state-pair differences at 1-mm WM depth.

| **CAP pair** | **NC, n** | **MDD, n** | **Hedges’ g [95% CI]** | **t** | **p** | **q** |
| --- | --- | --- | --- | --- | --- | --- |
| 1 / 1 † | 1130 | 1404 | −0.248 [−0.331, −0.164] | −5.80 | 7.67e−9 | 1.42e−7 |
| 2 / 2 † | 1130 | 1404 | −0.259 [−0.342, −0.176] | −6.10 | 1.20e−9 | 4.45e−8 |
| 2 / 3 | 1130 | 1404 | −0.106 [−0.193, −0.020] | −2.41 | 0.0161 | 0.0496 |
| 2 / 6 | 1130 | 1404 | −0.130 [−0.214, −0.045] | −3.02 | 0.0026 | 0.0106 |
| 3 / 3 | 1130 | 1404 | 0.147 [0.065, 0.230] | 3.52 | 4.47e−4 | 0.0021 |
| 3 / 5 | 1130 | 1404 | 0.177 [0.094, 0.260] | 4.16 | 3.29e−5 | 2.03e−4 |
| 4 / 4 | 1130 | 1404 | 0.158 [0.076, 0.241] | 3.76 | 1.76e−4 | 9.31e−4 |
| 4 / 5 | 1130 | 1404 | 0.199 [0.119, 0.280] | 4.87 | 1.18e−6 | 1.09e−5 |
| 4 / 7 | 1130 | 1404 | 0.109 [0.028, 0.191] | 2.63 | 0.0085 | 0.0285 |
| 5 / 1 † | 1130 | 1404 | −0.213 [−0.297, −0.129] | −4.98 | 6.78e−7 | 8.36e−6 |
| 6 / 5 | 1130 | 1404 | 0.181 [0.099, 0.263] | 4.35 | 1.41e−5 | 1.04e−4 |
| 7 / 4 | 1130 | 1404 | 0.115 [0.032, 0.198] | 2.71 | 0.0068 | 0.0253 |

Notes. Pairs are source CAP → target CAP; for simultaneous co-occurrence, read GM CAP / WM CAP. For directional asymmetry, pairs share source and target state labels across the two directions. Panels B–D use one-frame-lag conditional probabilities. Effects are MDD minus NC and are adjusted for age, sex, education and site after ComBat; confidence intervals are for Hedges’ g. Tables S13 and S14 list all 53 pairs with q < 0.05 across the four Figure 5 panels. BH-FDR was applied separately within each 49-pair family. † marks |g| > 0.20 (the heatmap’s red outline criterion). Thresholding uses unrounded estimates. Sample sizes can vary with source-state availability. All 196 pairs, including nonsignificant results, are supplied in Cross_tissue_1mm_all_pairs.csv.

Table S14. Group differences in lagged GM–WM state coupling at 1-mm WM depth. A. Directional asymmetry (WM-leading minus GM-leading).

| **CAP pair** | **NC, n** | **MDD, n** | **Hedges’ g [95% CI]** | **t** | **p** | **q** |
| --- | --- | --- | --- | --- | --- | --- |
| 1 → 4 | 1125 | 1387 | −0.114 [−0.198, −0.031] | −2.69 | 0.0073 | 0.0446 |
| 3 → 5 | 1130 | 1404 | −0.133 [−0.216, −0.049] | −3.12 | 0.0019 | 0.0191 |
| 5 → 1 † | 1130 | 1404 | 0.200 [0.115, 0.285] | 4.63 | 3.80e−6 | 9.32e−5 |
| 5 → 3 | 1130 | 1404 | 0.118 [0.035, 0.202] | 2.79 | 0.0054 | 0.0409 |
| 5 → 5 † | 1130 | 1404 | −0.204 [−0.287, −0.121] | −4.83 | 1.42e−6 | 6.98e−5 |
| 5 → 6 | 1130 | 1404 | 0.116 [0.033, 0.198] | 2.76 | 0.0058 | 0.0409 |
| 6 → 5 | 1130 | 1404 | −0.131 [−0.214, −0.048] | −3.10 | 0.0019 | 0.0191 |
| 6 → 6 | 1130 | 1404 | 0.135 [0.052, 0.219] | 3.18 | 0.0015 | 0.0191 |

Notes. Pairs are source CAP → target CAP; for simultaneous co-occurrence, read GM CAP / WM CAP. For directional asymmetry, pairs share source and target state labels across the two directions. Panels B–D use one-frame-lag conditional probabilities. Effects are MDD minus NC and are adjusted for age, sex, education and site after ComBat; confidence intervals are for Hedges’ g. Tables S13 and S14 list all 53 pairs with q < 0.05 across the four Figure 5 panels. BH-FDR was applied separately within each 49-pair family. † marks |g| > 0.20 (the heatmap’s red outline criterion). Thresholding uses unrounded estimates. Sample sizes can vary with source-state availability. All 196 pairs, including nonsignificant results, are supplied in Cross_tissue_1mm_all_pairs.csv.

Table S14. Continued. B. WM to GM.

| **CAP pair** | **NC, n** | **MDD, n** | **Hedges’ g [95% CI]** | **t** | **p** | **q** |
| --- | --- | --- | --- | --- | --- | --- |
| 1 → 2 | 1127 | 1392 | 0.166 [0.097, 0.234] | 4.73 | 2.42e−6 | 5.92e−5 |
| 2 → 1 | 1125 | 1396 | 0.115 [0.033, 0.197] | 2.75 | 0.0060 | 0.0296 |
| 2 → 6 | 1125 | 1396 | 0.113 [0.028, 0.199] | 2.61 | 0.0092 | 0.0377 |
| 3 → 2 † | 1130 | 1404 | −0.264 [−0.347, −0.181] | −6.25 | 4.83e−10 | 2.37e−8 |
| 3 → 3 | 1130 | 1404 | 0.148 [0.066, 0.230] | 3.53 | 4.25e−4 | 0.0042 |
| 3 → 5 | 1130 | 1404 | −0.136 [−0.219, −0.053] | −3.22 | 0.0013 | 0.0091 |
| 4 → 6 | 1130 | 1404 | −0.144 [−0.227, −0.060] | −3.37 | 7.59e−4 | 0.0062 |
| 5 → 1 | 1130 | 1404 | −0.192 [−0.274, −0.109] | −4.54 | 5.77e−6 | 9.42e−5 |
| 5 → 4 | 1130 | 1404 | 0.116 [0.034, 0.198] | 2.78 | 0.0054 | 0.0296 |
| 6 → 2 | 1130 | 1404 | −0.191 [−0.275, −0.107] | −4.46 | 8.45e−6 | 1.04e−4 |
| 6 → 6 | 1130 | 1404 | 0.115 [0.033, 0.196] | 2.75 | 0.0059 | 0.0296 |
| 7 → 1 | 1130 | 1404 | −0.110 [−0.192, −0.028] | −2.63 | 0.0085 | 0.0377 |

Notes. Pairs are source CAP → target CAP; for simultaneous co-occurrence, read GM CAP / WM CAP. For directional asymmetry, pairs share source and target state labels across the two directions. Panels B–D use one-frame-lag conditional probabilities. Effects are MDD minus NC and are adjusted for age, sex, education and site after ComBat; confidence intervals are for Hedges’ g. Tables S13 and S14 list all 53 pairs with q < 0.05 across the four Figure 5 panels. BH-FDR was applied separately within each 49-pair family. † marks |g| > 0.20 (the heatmap’s red outline criterion). Thresholding uses unrounded estimates. Sample sizes can vary with source-state availability. All 196 pairs, including nonsignificant results, are supplied in Cross_tissue_1mm_all_pairs.csv.

Table S14. Continued. C. GM to WM.

| **CAP pair** | **NC, n** | **MDD, n** | **Hedges’ g [95% CI]** | **t** | **p** | **q** |
| --- | --- | --- | --- | --- | --- | --- |
| 1 → 1 | 1126 | 1394 | −0.160 [−0.243, −0.077] | −3.79 | 1.55e−4 | 0.0015 |
| 1 → 4 | 1126 | 1394 | 0.111 [0.028, 0.194] | 2.63 | 0.0086 | 0.0222 |
| 2 → 1 † | 1126 | 1400 | 0.248 [0.176, 0.319] | 6.79 | 1.35e−11 | 6.62e−10 |
| 3 → 1 | 1130 | 1404 | −0.128 [−0.214, −0.041] | −2.90 | 0.0037 | 0.0115 |
| 3 → 2 † | 1130 | 1404 | −0.219 [−0.302, −0.137] | −5.20 | 2.19e−7 | 3.57e−6 |
| 3 → 3 | 1130 | 1404 | 0.149 [0.067, 0.232] | 3.54 | 4.03e−4 | 0.0022 |
| 3 → 7 | 1130 | 1404 | −0.151 [−0.234, −0.067] | −3.55 | 3.87e−4 | 0.0022 |
| 4 → 2 | 1130 | 1404 | −0.128 [−0.211, −0.044] | −3.00 | 0.0027 | 0.0089 |
| 4 → 5 | 1130 | 1404 | 0.128 [0.045, 0.210] | 3.03 | 0.0025 | 0.0086 |
| 4 → 6 | 1130 | 1404 | −0.110 [−0.193, −0.027] | −2.60 | 0.0093 | 0.0228 |
| 5 → 1 † | 1130 | 1404 | −0.276 [−0.360, −0.193] | −6.47 | 1.17e−10 | 2.87e−9 |
| 5 → 3 | 1130 | 1404 | −0.115 [−0.198, −0.031] | −2.68 | 0.0074 | 0.0201 |
| 5 → 5 | 1130 | 1404 | 0.196 [0.113, 0.280] | 4.62 | 4.09e−6 | 5.01e−5 |
| 5 → 6 | 1130 | 1404 | −0.158 [−0.242, −0.074] | −3.68 | 2.38e−4 | 0.0019 |
| 5 → 7 | 1130 | 1404 | 0.100 [0.017, 0.183] | 2.37 | 0.0178 | 0.0414 |
| 6 → 1 | 1130 | 1404 | −0.137 [−0.221, −0.052] | −3.18 | 0.0015 | 0.0056 |
| 6 → 2 | 1130 | 1404 | −0.144 [−0.227, −0.061] | −3.39 | 7.02e−4 | 0.0029 |
| 6 → 3 | 1130 | 1404 | 0.144 [0.060, 0.227] | 3.39 | 7.13e−4 | 0.0029 |
| 6 → 5 | 1130 | 1404 | 0.151 [0.069, 0.234] | 3.58 | 3.44e−4 | 0.0022 |
| 6 → 7 | 1130 | 1404 | −0.121 [−0.203, −0.038] | −2.86 | 0.0042 | 0.0122 |
| 7 → 1 | 1130 | 1404 | −0.142 [−0.225, −0.060] | −3.40 | 6.84e−4 | 0.0029 |

Notes. Pairs are source CAP → target CAP; for simultaneous co-occurrence, read GM CAP / WM CAP. For directional asymmetry, pairs share source and target state labels across the two directions. Panels B–D use one-frame-lag conditional probabilities. Effects are MDD minus NC and are adjusted for age, sex, education and site after ComBat; confidence intervals are for Hedges’ g. Tables S13 and S14 list all 53 pairs with q < 0.05 across the four Figure 5 panels. BH-FDR was applied separately within each 49-pair family. † marks |g| > 0.20 (the heatmap’s red outline criterion). Thresholding uses unrounded estimates. Sample sizes can vary with source-state availability. All 196 pairs, including nonsignificant results, are supplied in Cross_tissue_1mm_all_pairs.csv.

Table S15. State-conditional mismatch and frame-weighted sensitivity analysis.

| **Anchor / CAP** | **Primary significant depths** | **Primary max \|g\|** | **Primary interaction q** | **Weighted significant depths** | **Weighted max \|g\|** | **Weighted interaction q** |
| --- | --- | --- | --- | --- | --- | --- |
| GM / CAP-1 | None | 0.114 | 0.0687 | 3, 4 | 0.121 | 0.1578 |
| GM / CAP-2 | None | 0.095 | 0.4494 | None | 0.037 | 0.1272 |
| GM / CAP-3 | 4 | 0.123 | 0.0248 | None | 0.110 | 0.0280 |
| GM / CAP-4 | None | 0.040 | 0.3857 | None | 0.047 | 0.1595 |
| GM / CAP-5 | 2, 3, 4, 6, 7 | 0.140 | 0.2864 | 2, 3, 4, 6, 7 | 0.142 | 0.1272 |
| GM / CAP-6 | 1, 2, 3, 5 | 0.165 | 0.0721 | 1, 2, 3 | 0.154 | 0.0580 |
| GM / CAP-7 | None | 0.081 | 0.0248 | None | 0.078 | 0.0070 |
| WM / CAP-1 | None | 0.096 | 0.4494 | None | 0.032 | 0.6661 |
| WM / CAP-2 | 7 | 0.129 | 0.2113 | 2, 3, 7 | 0.111 | 0.1260 |
| WM / CAP-3 | None | 0.096 | 0.3217 | None | 0.100 | 0.2272 |
| WM / CAP-4 | None | 0.113 | 0.6682 | None | 0.113 | 0.6661 |
| WM / CAP-5 | 1, 2, 3 | 0.145 | 0.4494 | 1, 2, 3 | 0.153 | 0.1595 |
| WM / CAP-6 | None | 0.073 | 0.0687 | None | 0.045 | 0.0944 |
| WM / CAP-7 | None | 0.062 | 0.0326 | None | 0.065 | 0.0070 |

Notes. The fully adjusted model included age, sex, education and site. Primary analyses required at least 20 anchor-state frames; sensitivity analyses required at least one frame and weighted observations by available anchor frames, with weights capped at the 95th percentile. Significant depths (mm) satisfy BH-FDR q < 0.05 across 98 anchor-state-by-depth contrasts within each analysis and covariate model. Max |g| is the largest absolute adjusted Hedges’ g across all seven depths, not a pooled effect. Interaction q values were corrected across 14 anchor-state models within each analysis and covariate model. “None” means no depth survived correction. Sample sizes vary by anchor, state and depth. All 392 depth-specific contrasts and 56 interaction tests, including site-adjusted sensitivity models and sample sizes, are supplied in State_contrasts.csv and State_interactions.csv.

**Table S16. Group differences in GM-WM state coordination across depths.**

| **Outcome** | **AUC (g)** | **AUC (q)** | **b₁ (g)** | **b₁ (q)** | **Interaction (F)** | **Interaction (q)** |
| --- | --- | --- | --- | --- | --- | --- |
| Matched-state concordance | −0.148 | **4.69e−4** | −0.086 | 0.0785 | 7.27 | **1.79e−7** |
| Mismatch fraction | 0.148 | **4.69e−4** | 0.086 | 0.0785 | 7.27 | **1.79e−7** |
| Matching above chance | −0.205 | **4.22e−6** | −0.097 | 0.0785 | 7.72 | **1.79e−7** |
| Cohen’s κ | −0.181 | **3.04e−5** | −0.099 | 0.0785 | 7.44 | **1.79e−7** |
| Chance matching | 0.270 | **8.42e−10** | 0.068 | 0.1456 | 6.53 | **1.05e−6** |
| Normalized mutual information | −0.183 | **3.04e−5** | −0.004 | 0.9340 | 1.58 | 0.1483 |
| Adjusted Rand index | −0.172 | **6.43e−5** | 0.022 | 0.7003 | 4.41 | **2.26e−4** |

Notes. All seven outcomes are reported for 2,534 participants (1,404 MDD; 1,130 NC) across WM depths of 1-7 mm. AUC is the depth-normalized area under the curve; b₁ is the linear coefficient from each participant’s quadratic depth fit. Their group comparisons used OLS with participant-clustered, small-sample-corrected (HC1-equivalent) robust covariance, adjusted for age, sex, education and site. Hedges’ g is oriented MDD minus NC. Diagnosis × categorical-depth interactions used participant-clustered robust OLS with the same covariates (F₆,₂₅₃₃). BH-FDR correction was applied across seven outcomes separately for AUC, b₁ and interaction tests. Bold q values indicate q < 0.05. GM, gray matter; WM, white matter; MDD, major depressive disorder; NC, normal controls.

Table S17. Depth-specific adjusted group differences in state coordination.

| **Outcome** | **Depth (mm)** | **NC mean** | **MDD mean** | **MDD − NC [95% CI]** | **q** |
| --- | --- | --- | --- | --- | --- |
| Matched-state concordance | 1 | 0.6518 | 0.6421 | −0.0097 [−0.0137, −0.0057] | 1.73e−5 |
| Matched-state concordance | 2 | 0.6170 | 0.6087 | −0.0083 [−0.0125, −0.0040] | 2.70e−4 |
| Matched-state concordance | 3 | 0.5860 | 0.5765 | −0.0095 [−0.0140, −0.0050] | 7.77e−5 |
| Matched-state concordance | 4 | 0.4702 | 0.4610 | −0.0093 [−0.0135, −0.0050] | 7.64e−5 |
| Matched-state concordance | 5 | 0.5469 | 0.5399 | −0.0070 [−0.0117, −0.0023] | 0.0047 |
| Matched-state concordance | 6 | 0.5473 | 0.5431 | −0.0042 [−0.0089, 0.0006] | 0.0985 |
| Matched-state concordance | 7 | 0.4344 | 0.4361 | 0.0017 [−0.0023, 0.0056] | 0.4115 |
| Mismatch fraction | 1 | 0.3482 | 0.3579 | 0.0097 [0.0057, 0.0137] | 1.73e−5 |
| Mismatch fraction | 2 | 0.3830 | 0.3913 | 0.0083 [0.0040, 0.0125] | 2.70e−4 |
| Mismatch fraction | 3 | 0.4140 | 0.4235 | 0.0095 [0.0050, 0.0140] | 7.77e−5 |
| Mismatch fraction | 4 | 0.5298 | 0.5390 | 0.0093 [0.0050, 0.0135] | 7.64e−5 |
| Mismatch fraction | 5 | 0.4531 | 0.4601 | 0.0070 [0.0023, 0.0117] | 0.0047 |
| Mismatch fraction | 6 | 0.4527 | 0.4569 | 0.0042 [−0.0006, 0.0089] | 0.0985 |
| Mismatch fraction | 7 | 0.5656 | 0.5639 | −0.0017 [−0.0056, 0.0023] | 0.4115 |
| Matching above chance | 1 | 0.4949 | 0.4817 | −0.0132 [−0.0177, −0.0086] | 1.15e−7 |
| Matching above chance | 2 | 0.4603 | 0.4482 | −0.0121 [−0.0169, −0.0074] | 9.81e−7 |
| Matching above chance | 3 | 0.4295 | 0.4163 | −0.0132 [−0.0181, −0.0083] | 2.64e−7 |
| Matching above chance | 4 | 0.3171 | 0.3044 | −0.0127 [−0.0173, −0.0081] | 2.16e−7 |
| Matching above chance | 5 | 0.3912 | 0.3803 | −0.0109 [−0.0159, −0.0059] | 2.55e−5 |
| Matching above chance | 6 | 0.3919 | 0.3836 | −0.0083 [−0.0133, −0.0033] | 0.0014 |
| Matching above chance | 7 | 0.2838 | 0.2827 | −0.0011 [−0.0051, 0.0028] | 0.5756 |

Notes. All seven outcomes at all seven depths are reported, including nonsignificant contrasts. Participant-clustered robust OLS included diagnosis, categorical depth, their interaction, age, sex, education and site (2,534 participants). Means are model-adjusted estimates. Differences and confidence bounds are on the original outcome scale, not the Hedges’ g scale. BH-FDR correction was applied across seven depths separately within each outcome. Table S16 gives the across-depth AUC, quadratic-fit b₁ and interaction summaries. Site-adjusted sensitivity estimates and unrounded p values, standard errors and test statistics are provided in the accompanying source CSV files.

Table S17. Depth-specific adjusted group differences in state coordination. Continued.

| **Outcome** | **Depth (mm)** | **NC mean** | **MDD mean** | **MDD − NC [95% CI]** | **q** |
| --- | --- | --- | --- | --- | --- |
| Cohen’s κ | 1 | 0.5865 | 0.5732 | −0.0133 [−0.0184, −0.0083] | 1.40e−6 |
| Cohen’s κ | 2 | 0.5453 | 0.5333 | −0.0120 [−0.0173, −0.0067] | 1.88e−5 |
| Cohen’s κ | 3 | 0.5087 | 0.4952 | −0.0136 [−0.0191, −0.0080] | 4.01e−6 |
| Cohen’s κ | 4 | 0.3741 | 0.3604 | −0.0137 [−0.0189, −0.0084] | 1.40e−6 |
| Cohen’s κ | 5 | 0.4629 | 0.4520 | −0.0109 [−0.0166, −0.0052] | 2.74e−4 |
| Cohen’s κ | 6 | 0.4636 | 0.4560 | −0.0076 [−0.0134, −0.0019] | 0.0111 |
| Cohen’s κ | 7 | 0.3342 | 0.3339 | −0.0003 [−0.0049, 0.0044] | 0.9148 |
| Chance matching | 1 | 0.1570 | 0.1603 | 0.0033 [0.0022, 0.0044] | 2.94e−9 |
| Chance matching | 2 | 0.1569 | 0.1604 | 0.0036 [0.0025, 0.0047] | 4.11e−10 |
| Chance matching | 3 | 0.1566 | 0.1600 | 0.0034 [0.0024, 0.0045] | 8.40e−10 |
| Chance matching | 4 | 0.1533 | 0.1564 | 0.0031 [0.0021, 0.0041] | 2.48e−9 |
| Chance matching | 5 | 0.1558 | 0.1594 | 0.0036 [0.0025, 0.0046] | 1.49e−10 |
| Chance matching | 6 | 0.1556 | 0.1593 | 0.0037 [0.0027, 0.0048] | 5.16e−11 |
| Chance matching | 7 | 0.1508 | 0.1532 | 0.0023 [0.0014, 0.0032] | 4.39e−7 |
| Normalized mutual information | 1 | 0.5472 | 0.5352 | −0.0120 [−0.0167, −0.0072] | 6.04e−6 |
| Normalized mutual information | 2 | 0.4920 | 0.4805 | −0.0116 [−0.0164, −0.0067] | 9.14e−6 |
| Normalized mutual information | 3 | 0.4501 | 0.4384 | −0.0118 [−0.0167, −0.0068] | 9.14e−6 |
| Normalized mutual information | 4 | 0.4243 | 0.4133 | −0.0110 [−0.0159, −0.0060] | 2.26e−5 |
| Normalized mutual information | 5 | 0.4107 | 0.4013 | −0.0093 [−0.0143, −0.0044] | 3.38e−4 |
| Normalized mutual information | 6 | 0.4003 | 0.3922 | −0.0081 [−0.0131, −0.0030] | 0.0020 |
| Normalized mutual information | 7 | 0.3847 | 0.3771 | −0.0076 [−0.0126, −0.0027] | 0.0026 |
| Adjusted Rand index | 1 | 0.4252 | 0.4087 | −0.0165 [−0.0231, −0.0099] | 7.75e−6 |
| Adjusted Rand index | 2 | 0.3677 | 0.3539 | −0.0138 [−0.0200, −0.0075] | 2.97e−5 |
| Adjusted Rand index | 3 | 0.3254 | 0.3114 | −0.0140 [−0.0201, −0.0079] | 1.64e−5 |
| Adjusted Rand index | 4 | 0.2900 | 0.2765 | −0.0136 [−0.0194, −0.0078] | 1.64e−5 |
| Adjusted Rand index | 5 | 0.2826 | 0.2725 | −0.0100 [−0.0158, −0.0042] | 1.00e−3 |
| Adjusted Rand index | 6 | 0.2760 | 0.2688 | −0.0072 [−0.0130, −0.0013] | 0.0159 |
| Adjusted Rand index | 7 | 0.2556 | 0.2488 | −0.0069 [−0.0125, −0.0013] | 0.0159 |

Notes. All seven outcomes at all seven depths are reported, including nonsignificant contrasts. Participant-clustered robust OLS included diagnosis, categorical depth, their interaction, age, sex, education and site (2,534 participants). Means are model-adjusted estimates. Differences and confidence bounds are on the original outcome scale, not the Hedges’ g scale. BH-FDR correction was applied across seven depths separately within each outcome. Table S16 gives the across-depth AUC, quadratic-fit b₁ and interaction summaries. Site-adjusted sensitivity estimates and unrounded p values, standard errors and test statistics are provided in the accompanying source CSV files.

Table S18. Significant receptor associations of MDD-related GM and WM CAP effects.

| **Tissue** | **CAP** | **Receptor/transporter** | **Spearman ρ** | **Spin p** | **q** |
| --- | --- | --- | --- | --- | --- |
| GM | CAP-1 | 5HT2a | −0.370 | 4.00e−4 | 0.0199 |
| GM | CAP-1 | MOR | −0.425 | 6.00e−4 | 0.0199 |
| GM | CAP-2 | 5HT2a | 0.403 | 4.00e−4 | 0.0199 |
| GM | CAP-2 | MOR | 0.425 | 6.00e−4 | 0.0199 |
| GM | CAP-2 | mGluR5 | 0.328 | 0.0014 | 0.0310 |
| GM | CAP-4 | VAChT | −0.360 | 0.0012 | 0.0310 |
| WM | CAP-1 | 5HTT | 0.392 | 9.00e−4 | 0.0239 |
| WM | CAP-6 | 5HT2a | 0.355 | 4.00e−4 | 0.0199 |
| WM | CAP-6 | 5HT4 | 0.323 | 6.00e−4 | 0.0199 |
| WM | CAP-6 | 5HTT | −0.351 | 4.00e−4 | 0.0199 |
| WM | CAP-7 | 5HT4 | −0.349 | 6.00e−4 | 0.0199 |
| WM | CAP-7 | 5HTT | 0.288 | 0.0024 | 0.0456 |
| WM | CAP-7 | MOR | −0.303 | 0.0016 | 0.0355 |

Notes. All 13 significant associations in Figure 6J–K are listed (6 GM; 7 WM). Each correlation relates a signed parcel-wise Hedges’ g map (MDD minus NC) for phenotype A to an independent normative receptor/transporter map; it is not a group comparison of receptor density. A is the mean activation across the seven depth conditions. Parcel-wise group models adjusted for age, sex, education, mean framewise displacement and site; model-specific sample sizes are in Table S2. Spearman correlations used 400 corresponding parcels and 10,000 two-sided spatial-spin tests. BH-FDR was applied across 133 CAP-by-receptor tests separately for GM and WM. Absolute-g sensitivity analyses used separate correction families and yielded no q < 0.05 associations in either tissue. Complete signed- and absolute-g results, and the seven-depth results related to Figure S22, are supplied in Disease_receptor_A.csv and Disease_receptor_depth.csv.

**Supplementary Figures**

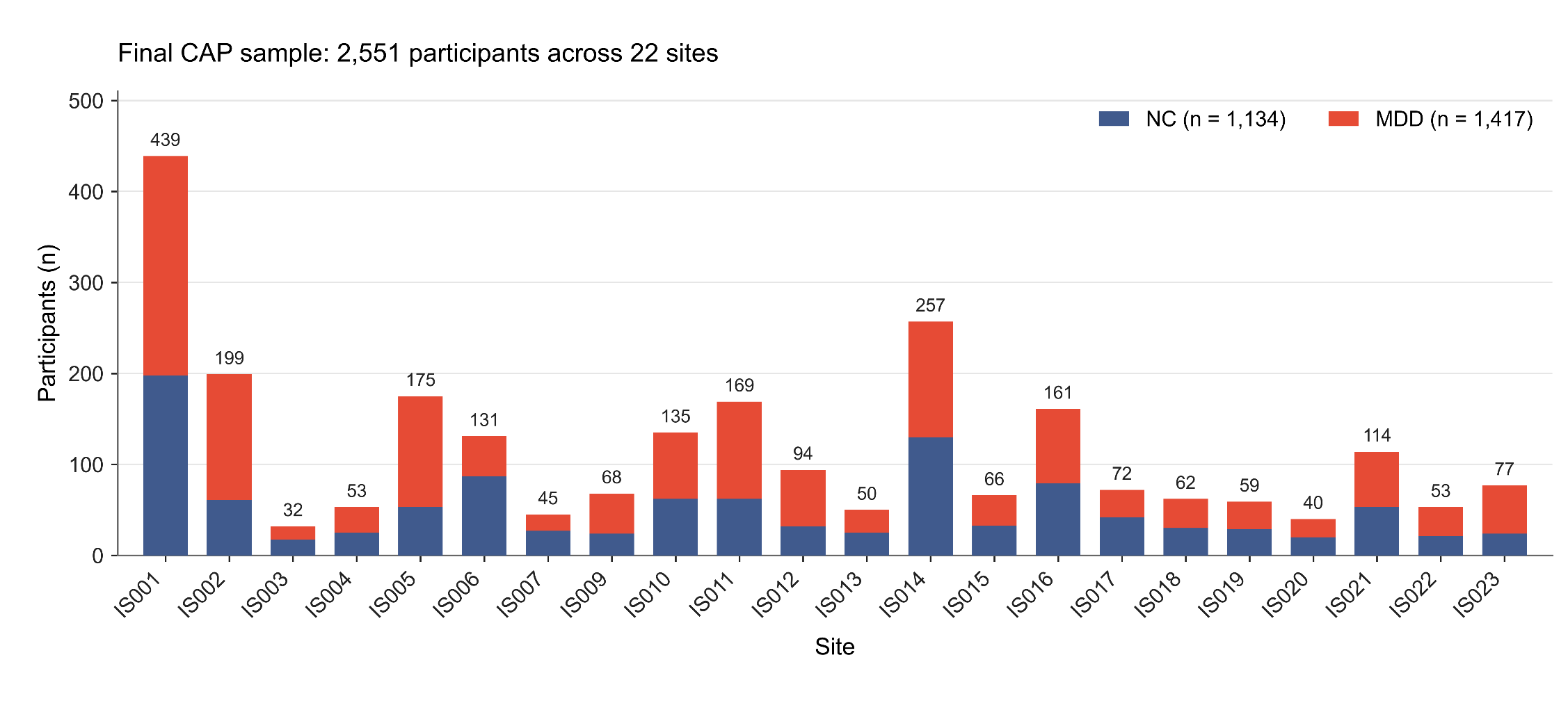

Figure S1. Site-wise composition of the final CAP cohort. Stacked bars show the numbers of normal controls (NC; blue) and participants with major depressive disorder (MDD; red) at each of the 22 sites after study-specific quality control and time-series availability checks. The final cohort comprised 2,551 participants (1,417 MDD and 1,134 NC); numbers above bars indicate site totals. IS008 is absent from the final CAP cohort.

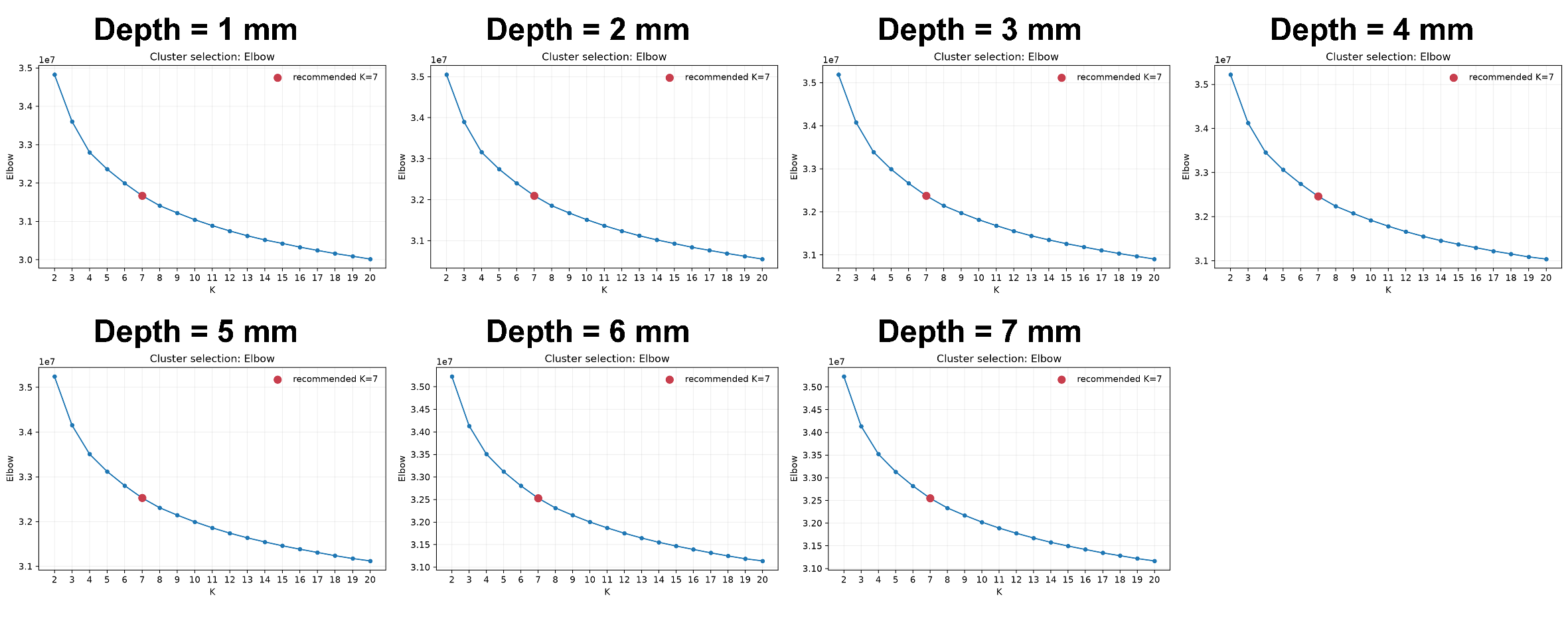

Figure S2. Elbow criterion for selecting the number of clusters in k-means clustering at each of the seven WM sampling depths.

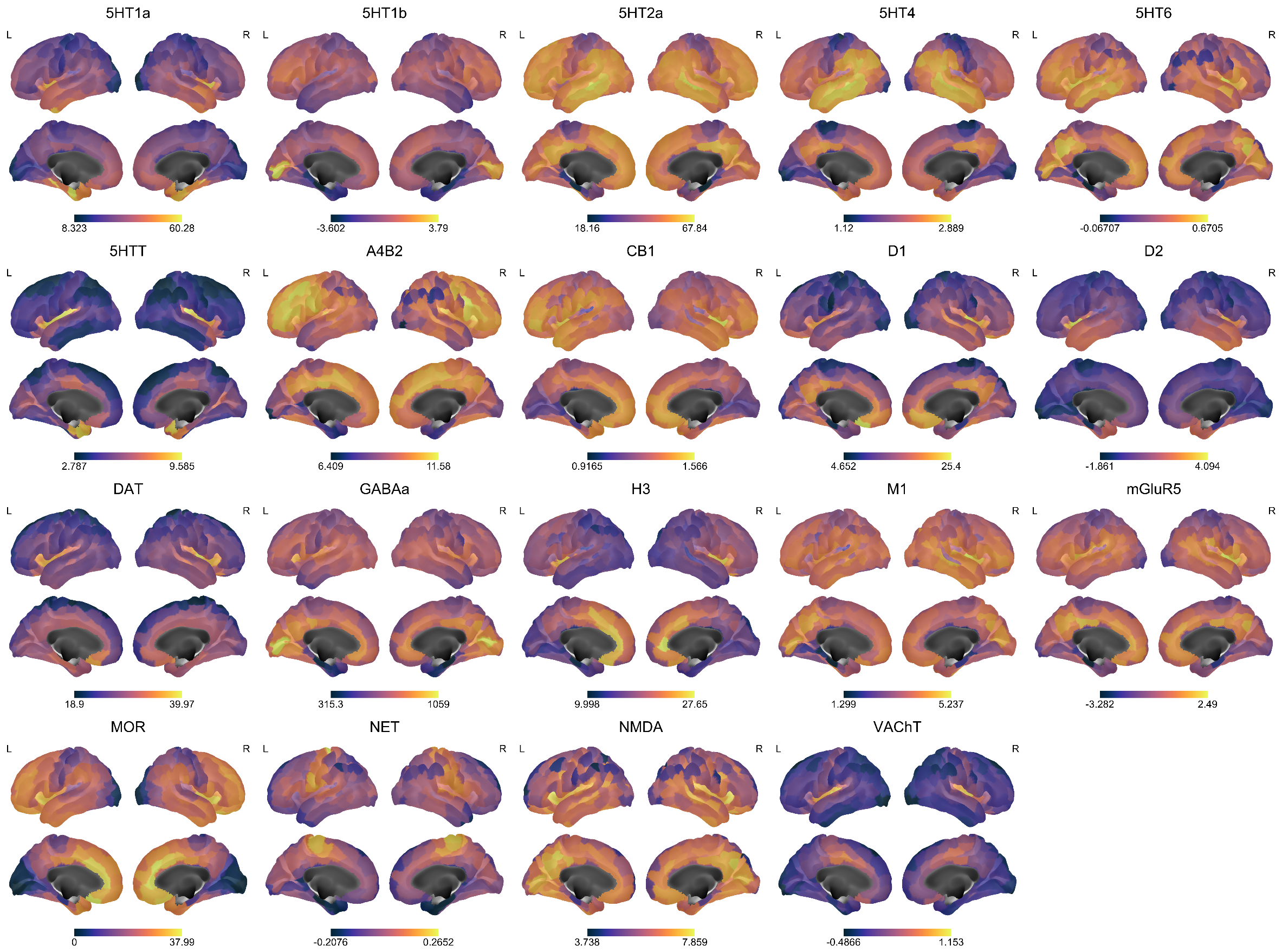

Figure S3. Spatial distributions of 19 neurotransmitter receptors and transporters. Regional receptor and transporter values are displayed using the 400-parcel Schaefer atlas on the fsLR cortical surface. Each panel shows the lateral views of the left and right hemispheres in the upper row and the corresponding medial views in the lower row. Colors progress from dark blue/purple to yellow with increasing values. Each receptor or transporter is displayed on its own color scale, with the colorbar beneath the panel indicating the minimum and maximum values across parcels. L, left; R, right.

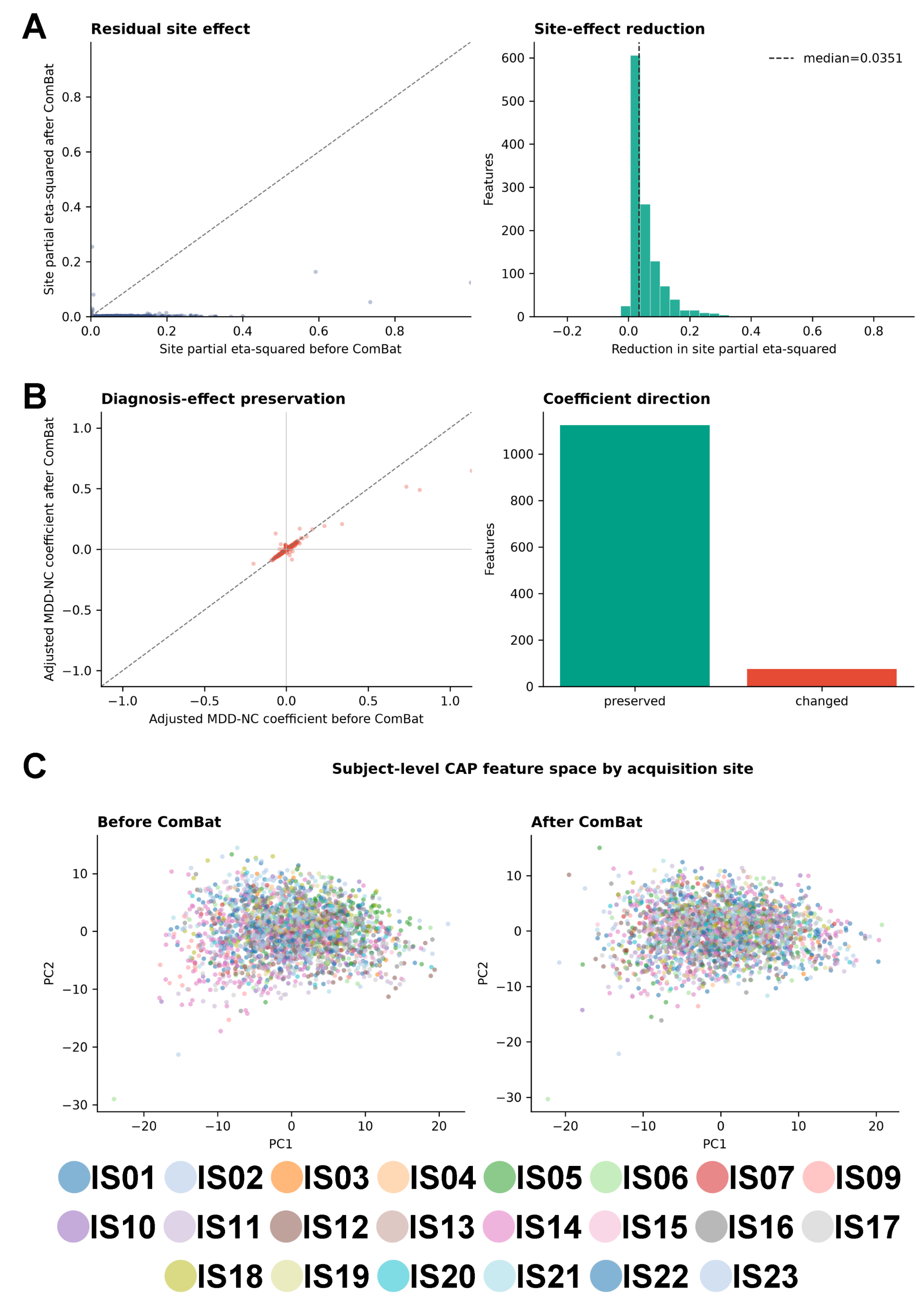

Figure S4. Evaluation of ComBat harmonization of subject-level coactivation pattern features. **(A)** Acquisition-site effects before and after ComBat harmonization, quantified by partial eta-squared (η²ₚ). The scatterplot compares site-effect sizes before and after harmonization; points below the dashed identity line indicate reduced site effects. The histogram shows the distribution of feature-wise reductions, calculated as η²ₚ before minus η²ₚ after ComBat. The vertical dashed line marks the median reduction (0.0351). **(B)** Preservation of diagnosis-related effects. The scatterplot compares covariate-adjusted MDD–NC regression coefficients before and after harmonization, with the dashed identity line indicating unchanged coefficients. The bar chart summarizes the number of features with preserved or changed coefficient directions. Each point in the scatterplots in A and B represents one CAP-derived feature. **(C)** Subject-level CAP feature distributions along the first two principal components before (left) and after (right) ComBat harmonization. Each point represents one participant, and colors indicate acquisition sites, as identified in the legend. CAP, coactivation pattern; ComBat, combating batch effects; MDD, major depressive disorder; NC, normal control; PC, principal component.

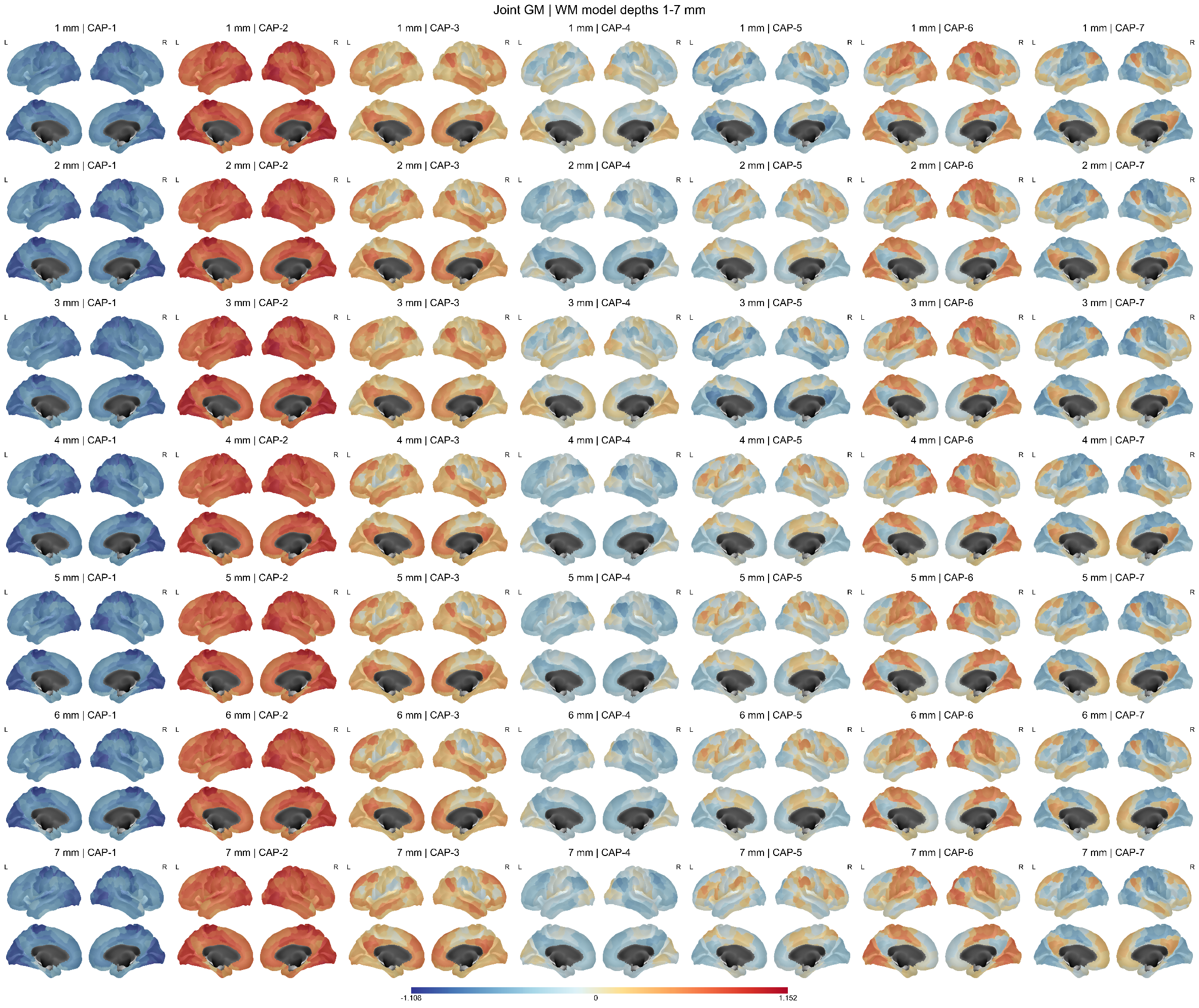

Figure S5. Gray matter components of joint coactivation patterns across white matter sampling depths. Gray matter components of seven CAPs identified by joint clustering of GM and WM signals are shown. Columns represent CAP-1 to CAP-7, and rows represent analyses with WM sampling depths increasing from 1 to 7 mm below the GM–WM boundary. GM sampling was fixed at 1 mm outward from the boundary in all analyses; therefore, the row labels denote the associated WM sampling depth. Regional centroid values are displayed using the 400-parcel Schaefer atlas. Within each panel, lateral views of the left and right hemispheres are shown above the corresponding medial views. All CAP maps share a common zero-centered color mapping, with blue and red indicating negative and positive centroid values, respectively. A single colorbar is shared across all CAPs and depth conditions within the figure. L, left; R, right.

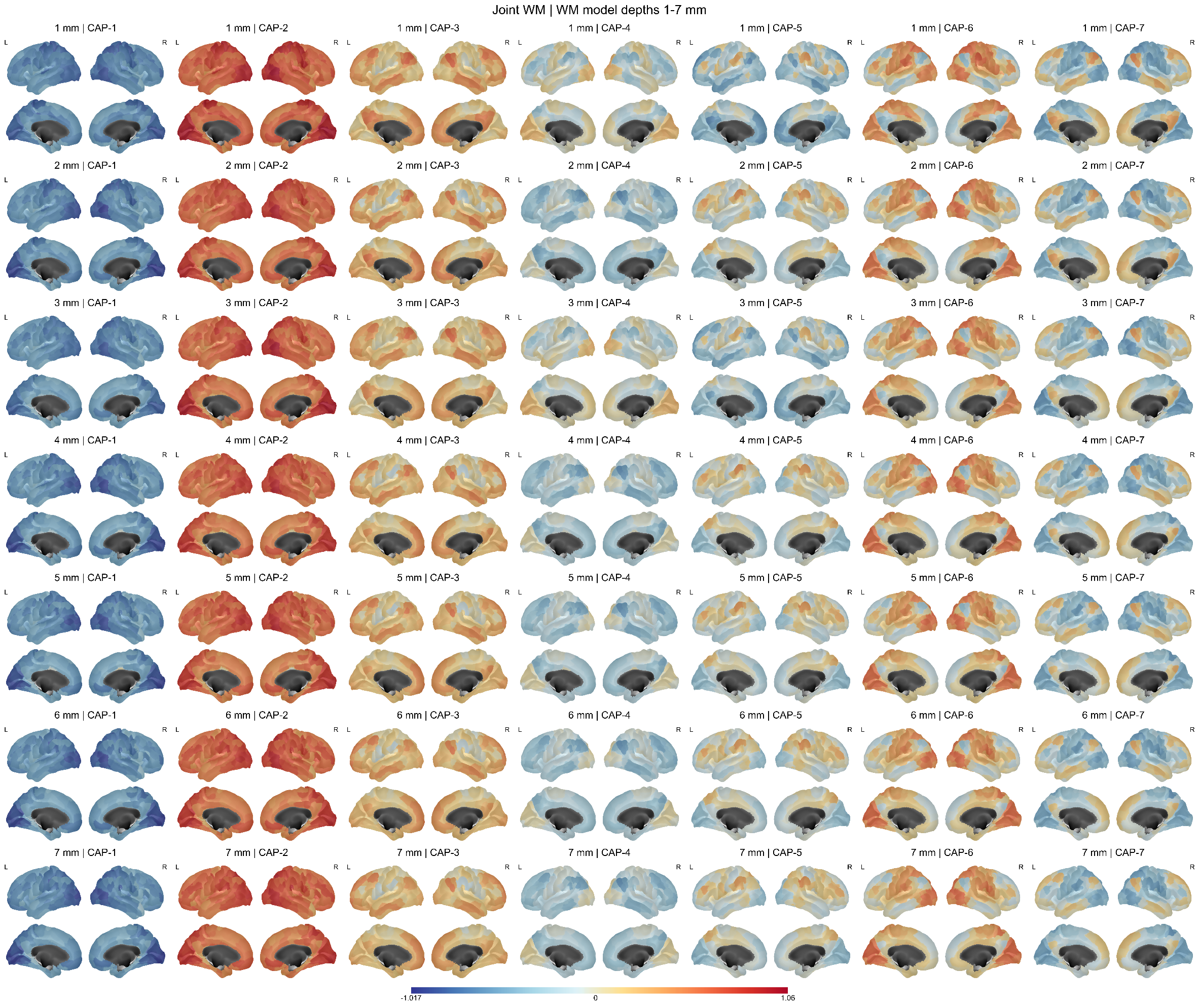

Figure S6. White matter components of joint coactivation patterns across sampling depths. White matter components of seven CAPs identified by joint clustering of GM and WM signals are shown. Columns represent CAP-1 to CAP-7, and rows represent WM sampling depths of 1–7 mm inward from the GM–WM boundary along the cortical surface normals. GM signals were sampled at a fixed position 1 mm outward from the boundary. For visualization, each WM parcel’s centroid value is displayed on its corresponding cortical parcel in the 400-parcel Schaefer atlas. Within each panel, lateral views of the left and right hemispheres are shown above the corresponding medial views. All CAP maps share a common zero-centered color mapping, with blue and red indicating negative and positive centroid values, respectively. A single colorbar is shared across all CAPs and depth conditions within the figure. L, left; R, right.

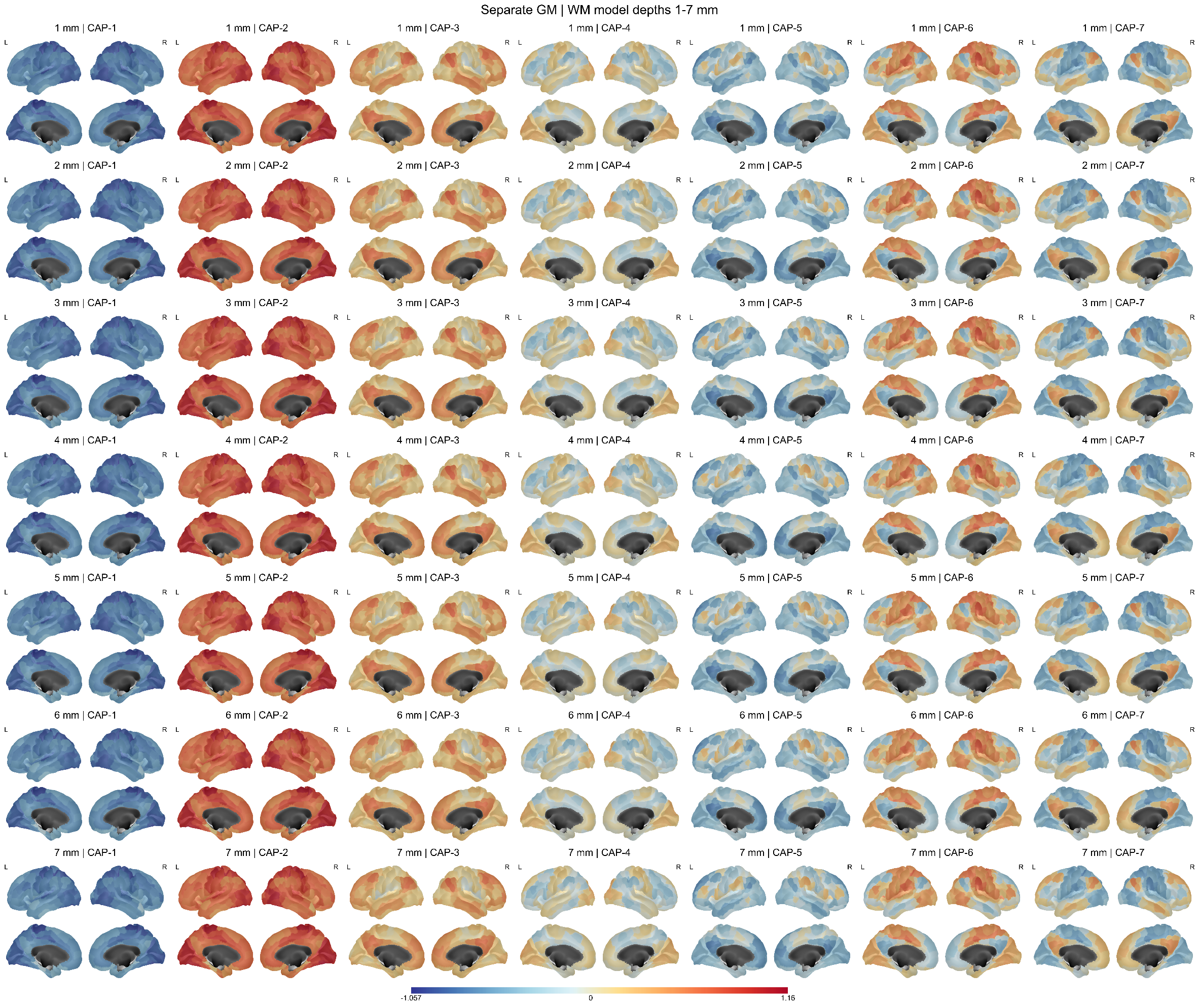

Figure S7. Gray matter coactivation patterns estimated separately from white matter across depth-specific analyses. Spatial centroid maps of seven gray matter CAPs, estimated by clustering GM signals separately from WM signals, are shown. Columns represent CAP-1 to CAP-7. Rows correspond to the analysis runs associated with WM sampling depths of 1–7 mm; GM sampling remained fixed at 1 mm outward from the GM–WM boundary in every run. Regional centroid values are displayed using the 400-parcel Schaefer atlas. Within each panel, lateral views of the left and right hemispheres are shown above the corresponding medial views. All CAP maps share a common zero-centered color mapping, with blue and red indicating negative and positive centroid values, respectively. A single colorbar is shared across all CAPs and depth conditions within the figure. L, left; R, right.

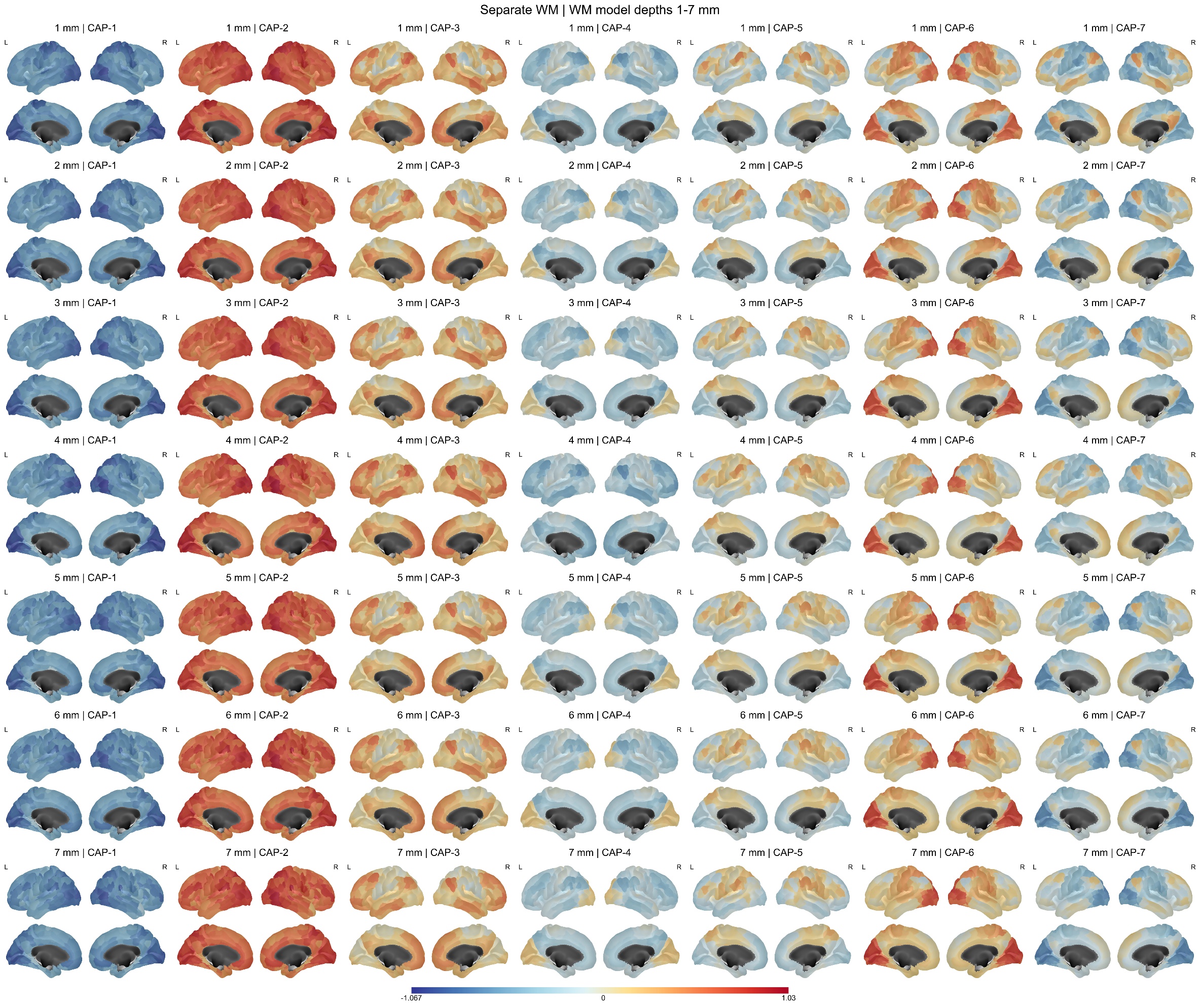

Figure S8. White matter coactivation patterns estimated separately from gray matter across sampling depths. Spatial centroid maps of seven white matter CAPs, estimated by clustering WM signals separately from gray matter (GM) signals at each sampling depth, are shown. Columns represent CAP-1 to CAP-7, and rows represent WM sampling depths increasing from 1 to 7 mm inward from the GM–WM boundary along the cortical surface normals. For visualization, each WM parcel’s centroid value is displayed on its corresponding cortical parcel in the 400-parcel Schaefer atlas. Within each panel, lateral views of the left and right hemispheres are shown above the corresponding medial views. All CAP maps share a common zero-centered color mapping, with blue and red indicating negative and positive centroid values, respectively. A single colorbar is shared across all CAPs and depth conditions within the figure. L, left; R, right.

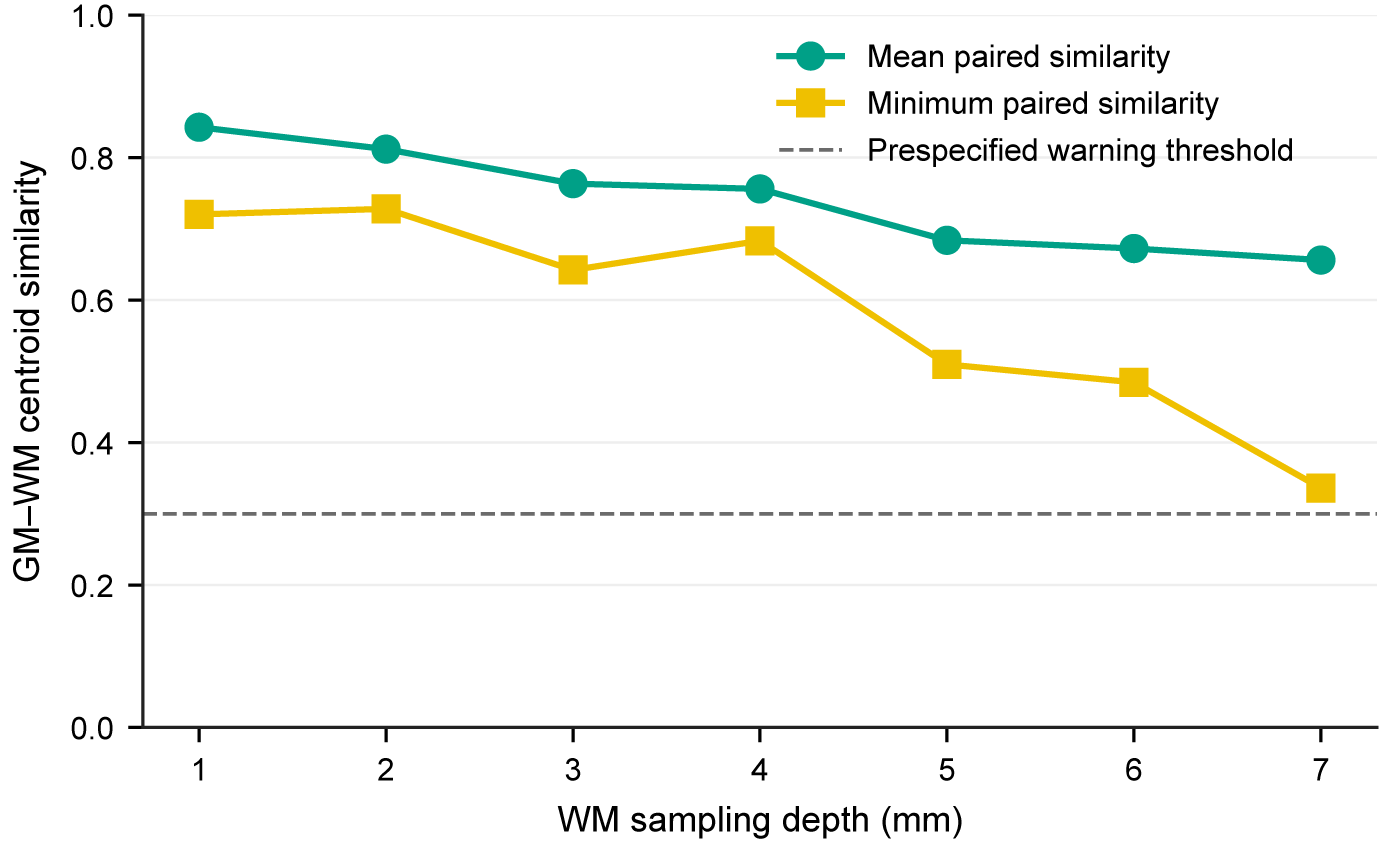

Figure S9. GM-WM CAP matching quality across white matter sampling depths. Coactivation patterns (CAPs) estimated separately from gray matter (GM) and white matter (WM) were paired at each WM sampling depth using a one-to-one Hungarian assignment that maximized spatial Pearson correlations between their centroid maps across 400 cortical parcels. GM sampling was fixed at 1 mm outside the GM–WM boundary, whereas WM sampling extended from 1 to 7 mm inward. Teal circles and yellow squares indicate the mean and minimum centroid similarities, respectively, across the seven matched CAP pairs at each depth. The gray dashed line denotes the predefined quality-control warning threshold (r = 0.30), rather than a statistical significance threshold. Mean GM–WM centroid similarity decreased with greater WM depth, while the minimum paired similarity remained above the warning threshold at all depths.

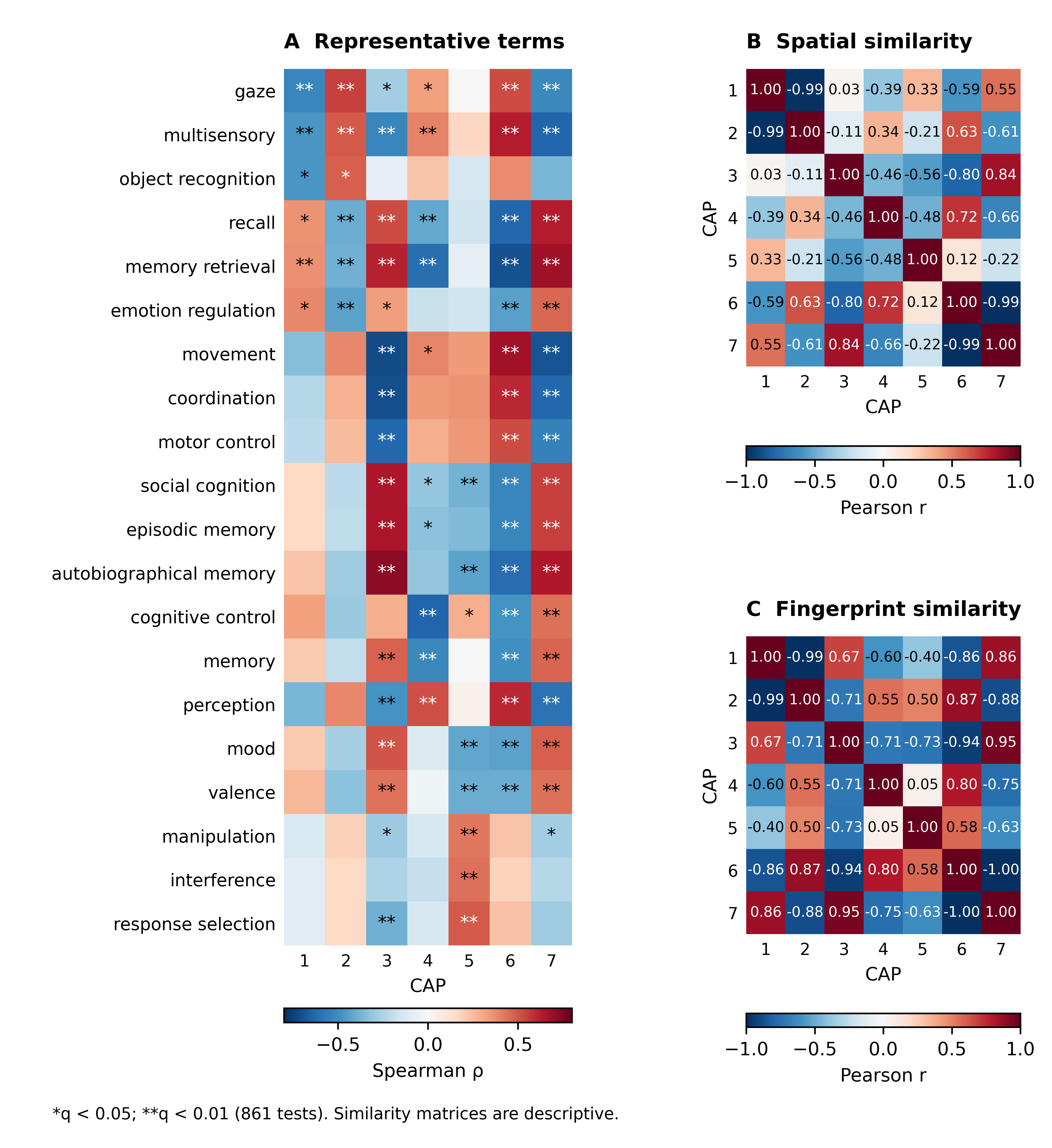

Figure S10. Meta-analytic functional fingerprints and similarities of shared GM–WM CAPs. (A) Signed Spearman correlations between the adjusted NC phenotype A maps and 20 representative Neurosynth association-test maps across 400 cortical parcels. Terms are the union of the three strongest positive and three strongest negative correlations for each CAP. Asterisks indicate two-sided, null-mean-centered spatial-spin significance after BH-FDR correction across all 861 comparisons (7 CAPs × 123 terms; 10,000 spins): *q < 0.05, **q < 0.01. Counts of significant terms among all 123 were 17, 19, 55, 40, 29, 51, and 56 for CAP1–CAP7, respectively (Table S5). (B) Pearson spatial correlations between the 400-parcel CAP maps. (C) Pearson correlations between the complete, unthresholded 123-term coefficient vectors. Panels B and C are descriptive; no significance testing was applied. Positive and negative values indicate similar and opposing profiles, respectively. All 123 terms are shown in Figure S11 and Table S6.

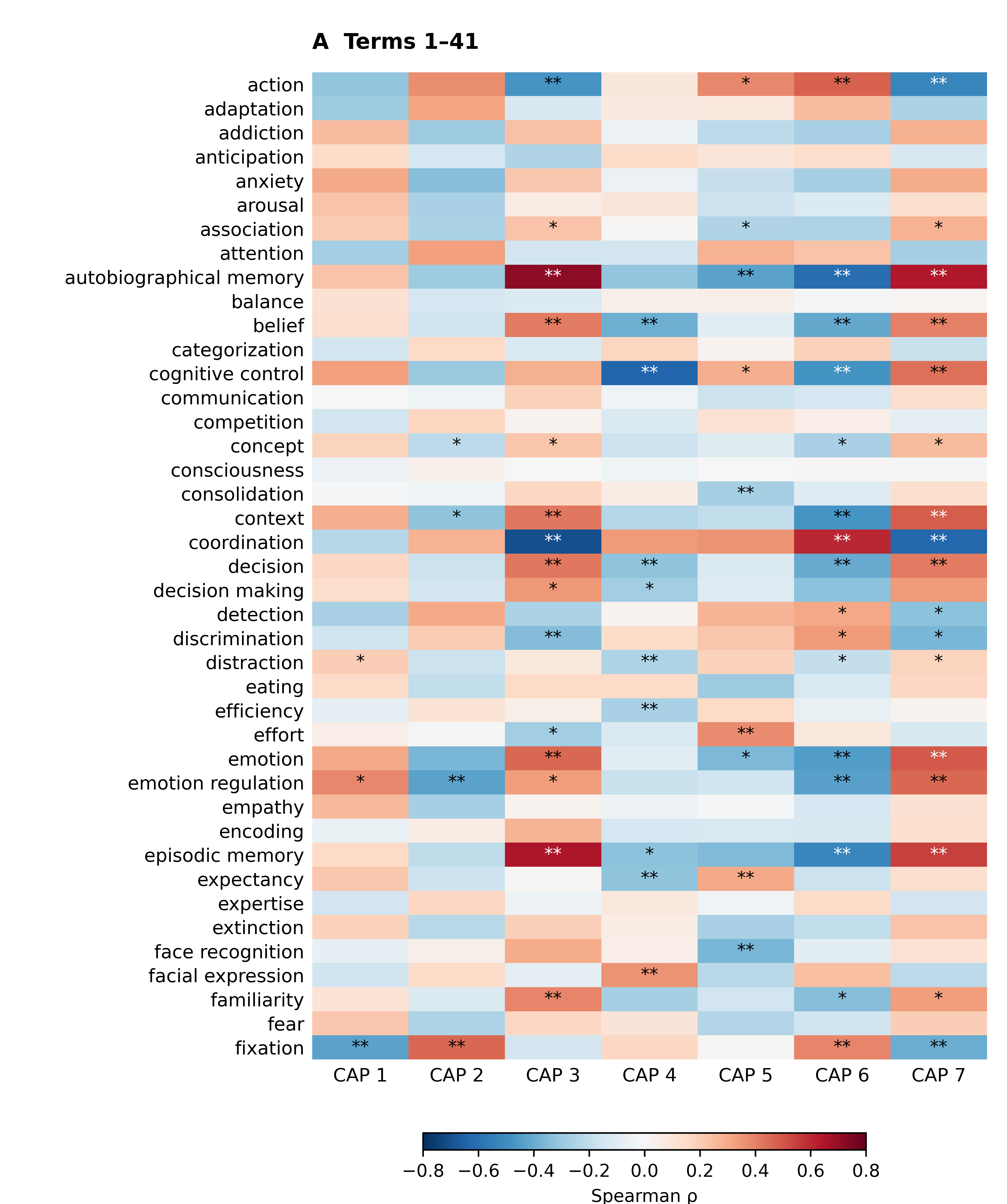

Figure S11. Complete 123-term functional profiles of shared GM–WM CAPs. (A) Terms 1–41 in alphabetical order. Colors show signed Spearman correlations between the adjusted NC phenotype A maps and Neurosynth association-test maps across 400 parcels. Panels A–C share a color scale. All correlations are displayed; cells without asterisks do not meet the significance criterion. *q < 0.05, **q < 0.01, based on 10,000 two-sided, null-mean-centered spatial spins and BH-FDR correction across all 861 CAP-by-term tests. Numerical estimates are reported in Table S6.

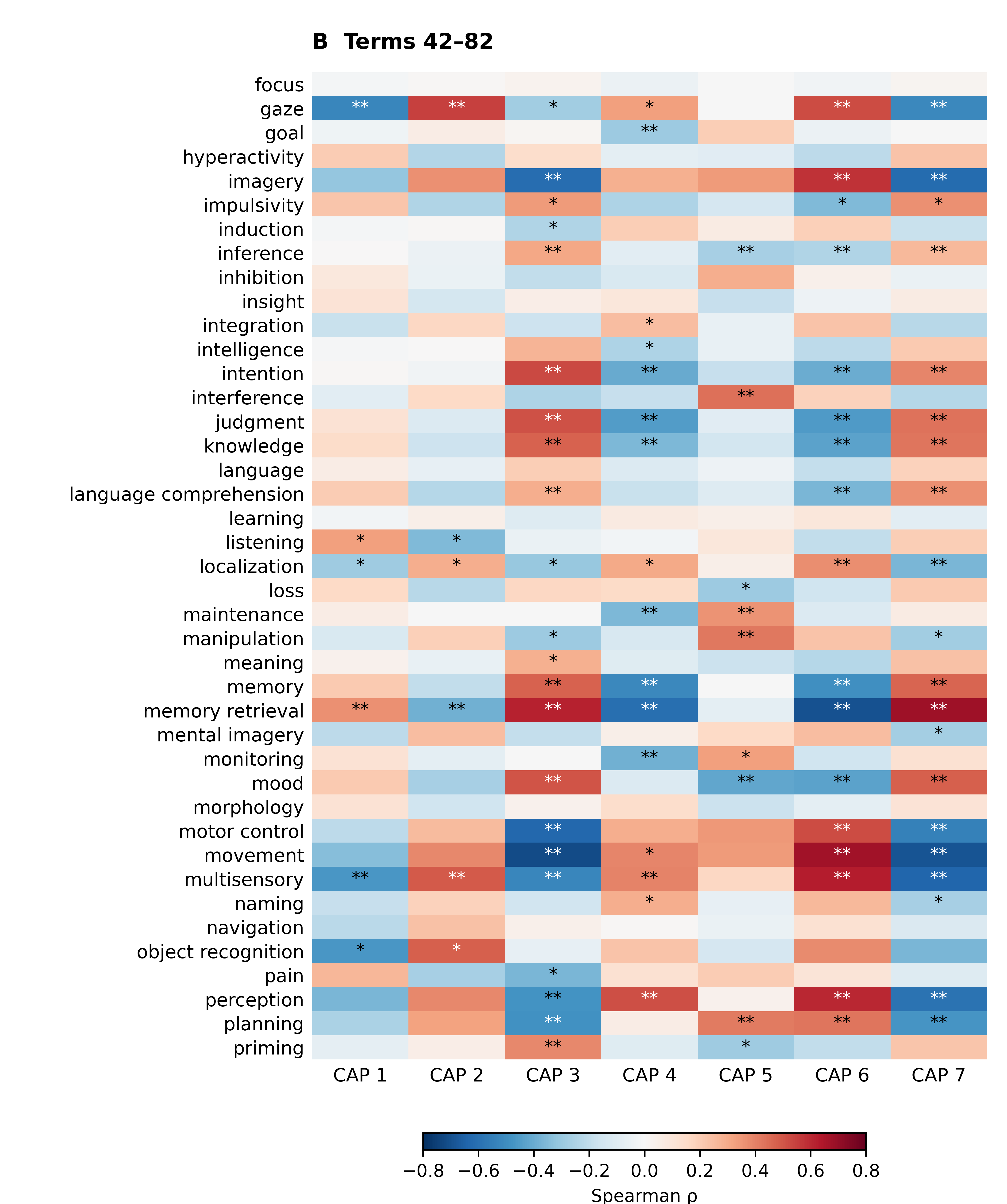

Figure S11. continued. (B) Terms 42–82 in alphabetical order. Colors show signed Spearman correlations between the adjusted NC phenotype A maps and Neurosynth association-test maps across 400 parcels. Panels A–C share a color scale. All correlations are displayed; cells without asterisks do not meet the significance criterion. *q < 0.05, **q < 0.01, based on 10,000 two-sided, null-mean-centered spatial spins and BH-FDR correction across all 861 CAP-by-term tests. Numerical estimates are reported in Table S6.

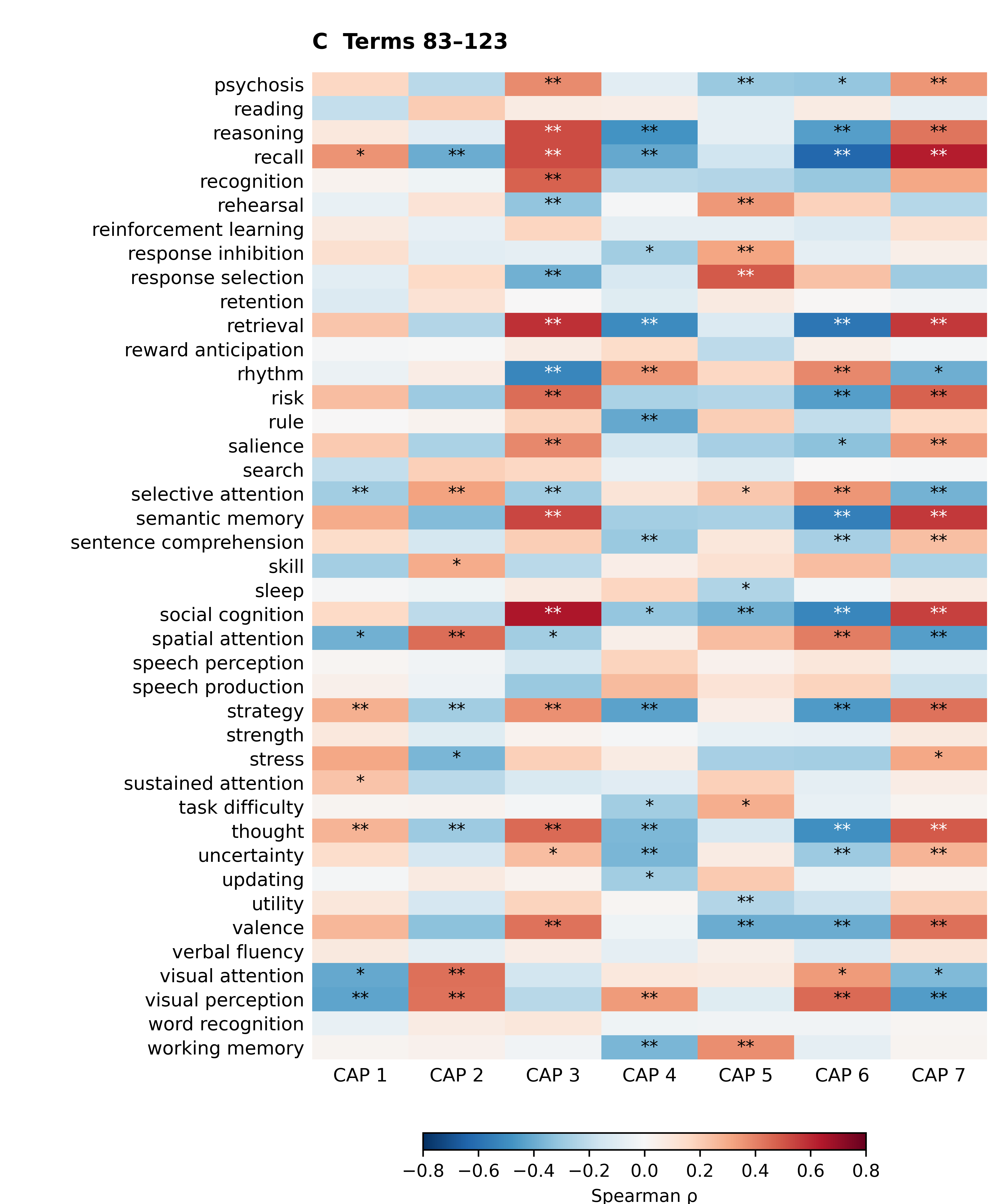

Figure S11. continued. (C) Terms 83–123 in alphabetical order. Colors show signed Spearman correlations between the adjusted NC phenotype A maps and Neurosynth association-test maps across 400 parcels. Panels A–C share a color scale. All correlations are displayed; cells without asterisks do not meet the significance criterion. *q < 0.05, **q < 0.01, based on 10,000 two-sided, null-mean-centered spatial spins and BH-FDR correction across all 861 CAP-by-term tests. Numerical estimates are reported in Table S6.

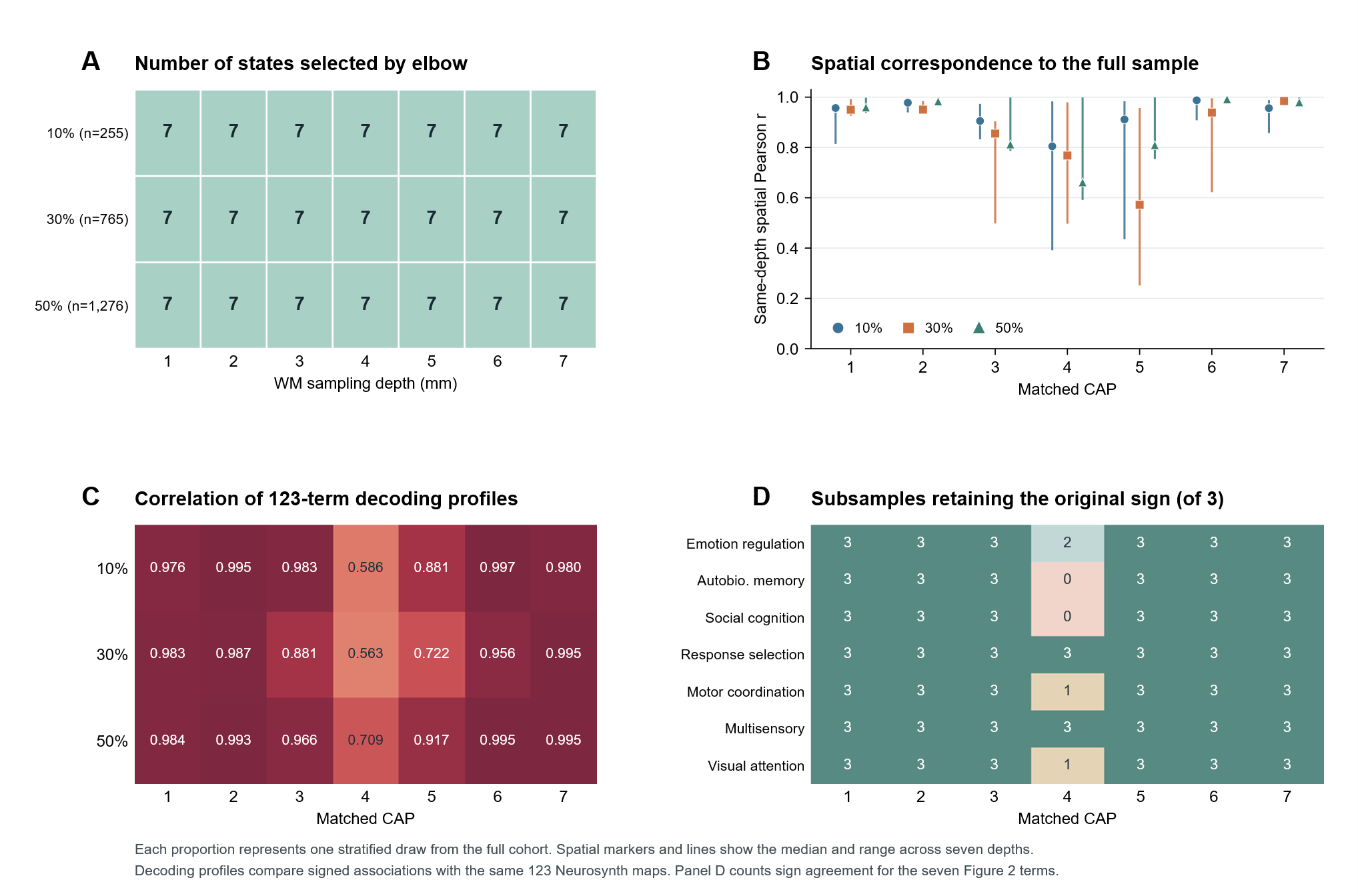

Figure S12. Sensitivity of joint GM–WM CAPs and their functional annotations to participant subsampling. One site- and diagnosis-stratified sample was drawn at each proportion of the full cohort (n = 2,551). (A) Number of clusters selected by the elbow criterion at each WM depth. (B) Signed Pearson correlation between each subsample centroid and its corresponding full-cohort centroid at the same depth. Symbols indicate medians, and lines span the range across the seven depths. (C) Spearman correlations between the subsample and full-cohort decoding profiles across 123 Neurosynth terms for each CAP. Profiles were derived from the adjusted NC mean shared-activation maps averaged across 1–7 mm (phenotype A). (D) Number of subsamples, from zero to three, in which each of the seven representative term associations had the same sign as in the full cohort. Table S7 gives the numerical results.

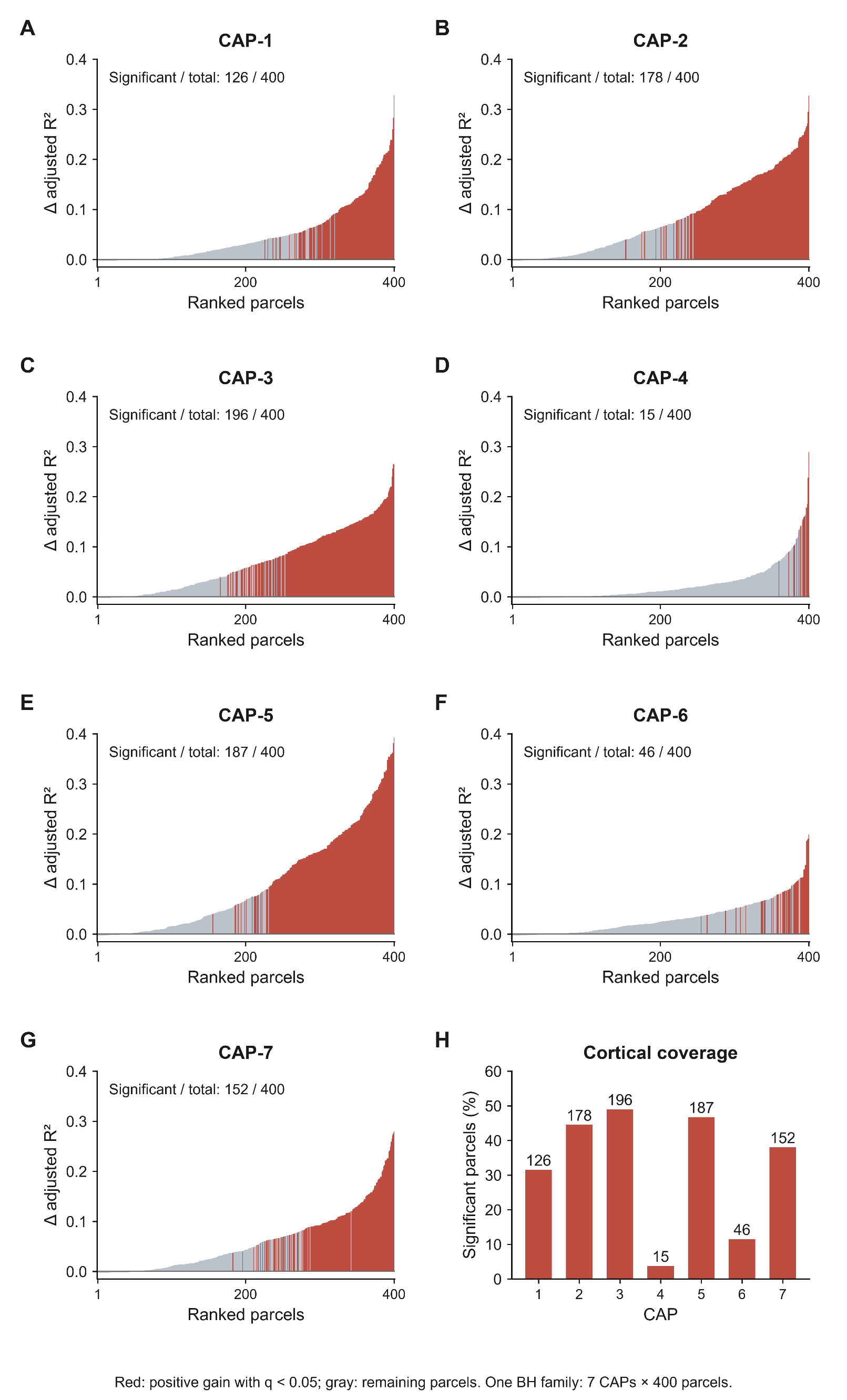

Figure S13. Regional explanatory gains from receptor information across joint CAPs. (A-G) Cortical parcels ranked by the increase in adjusted R² attributable to receptor information beyond spatial distance for CAP1-7. Red indicates significant positive gains; gray indicates remaining parcels. Significance was assessed using Benjamini-Hochberg FDR correction across all 2,800 CAP-parcel tests (q < 0.05). Significant parcel counts are shown within each panel. (H) Percentage of cortical parcels showing significant positive gains, with corresponding counts above the bars.

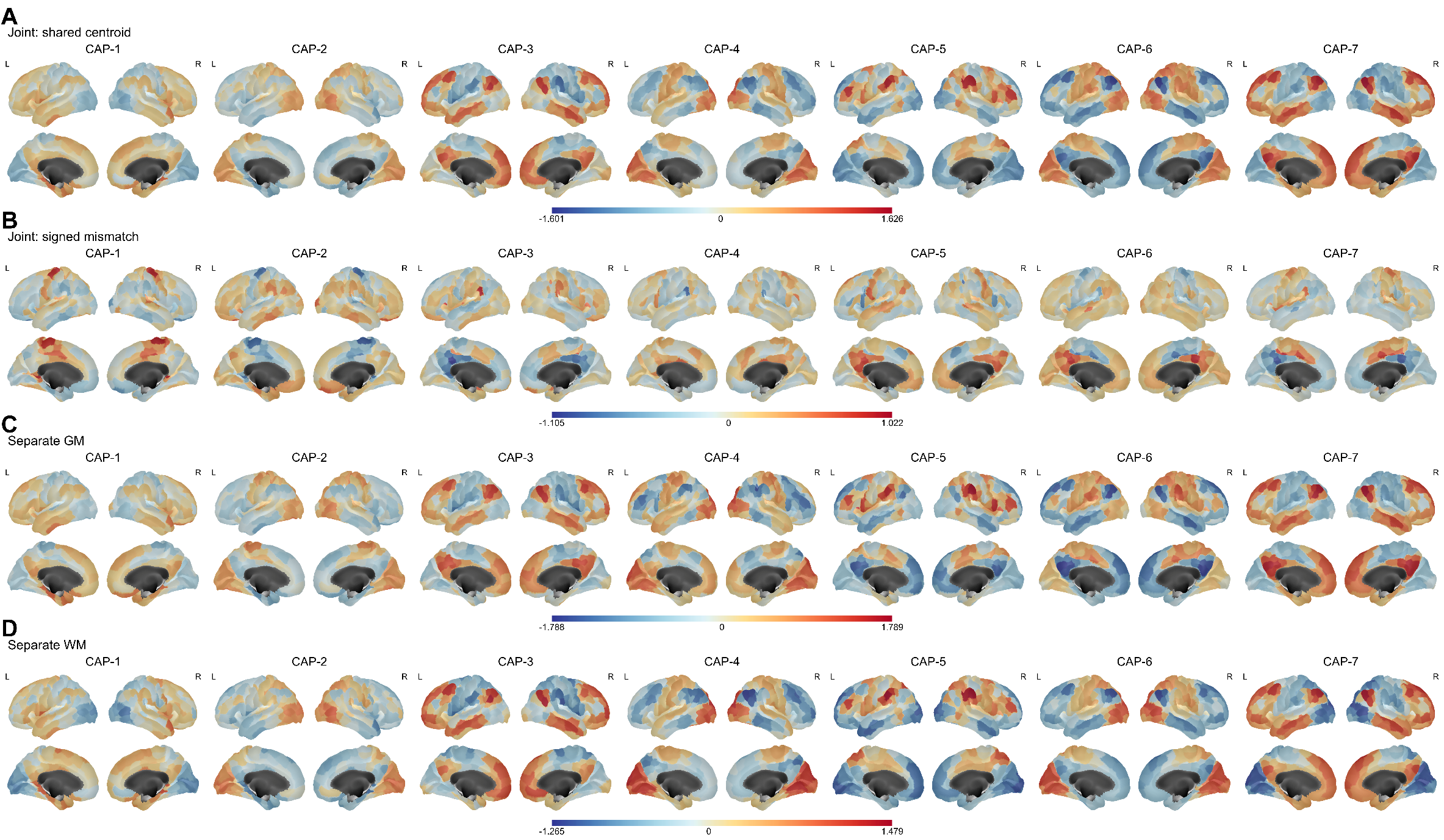

Figure S14. Spatial distribution of depth-averaged boundary CAP phenotypes in healthy controls (phenotype A). Maps show covariate-adjusted mean values in healthy controls. For each participant, phenotype A was calculated by averaging the ComBat-harmonized regional measure across white matter (WM) sampling depths of 1-7 mm. (A) Shared centroid from the joint gray matter (GM)-WM model, derived from the mean of standardized GM and WM activation. (B) Signed tissue difference from the joint model, defined as standardized WM minus GM activation. (C, D) Activation from the separate GM and WM models, respectively. Columns represent coactivation patterns CAP-1-CAP-7. GM sampling remained fixed at 1 mm outside the GM-WM boundary. Values for the 400 Schaefer parcels are displayed on the corresponding cortical surface parcels, with lateral views above and medial views below for both hemispheres. Warm and cool colors indicate positive and negative phenotype values, respectively. Each panel shares one colorbar across its seven CAPs, with limits determined by the minimum and maximum values within that panel. Maps are displayed without significance thresholding.

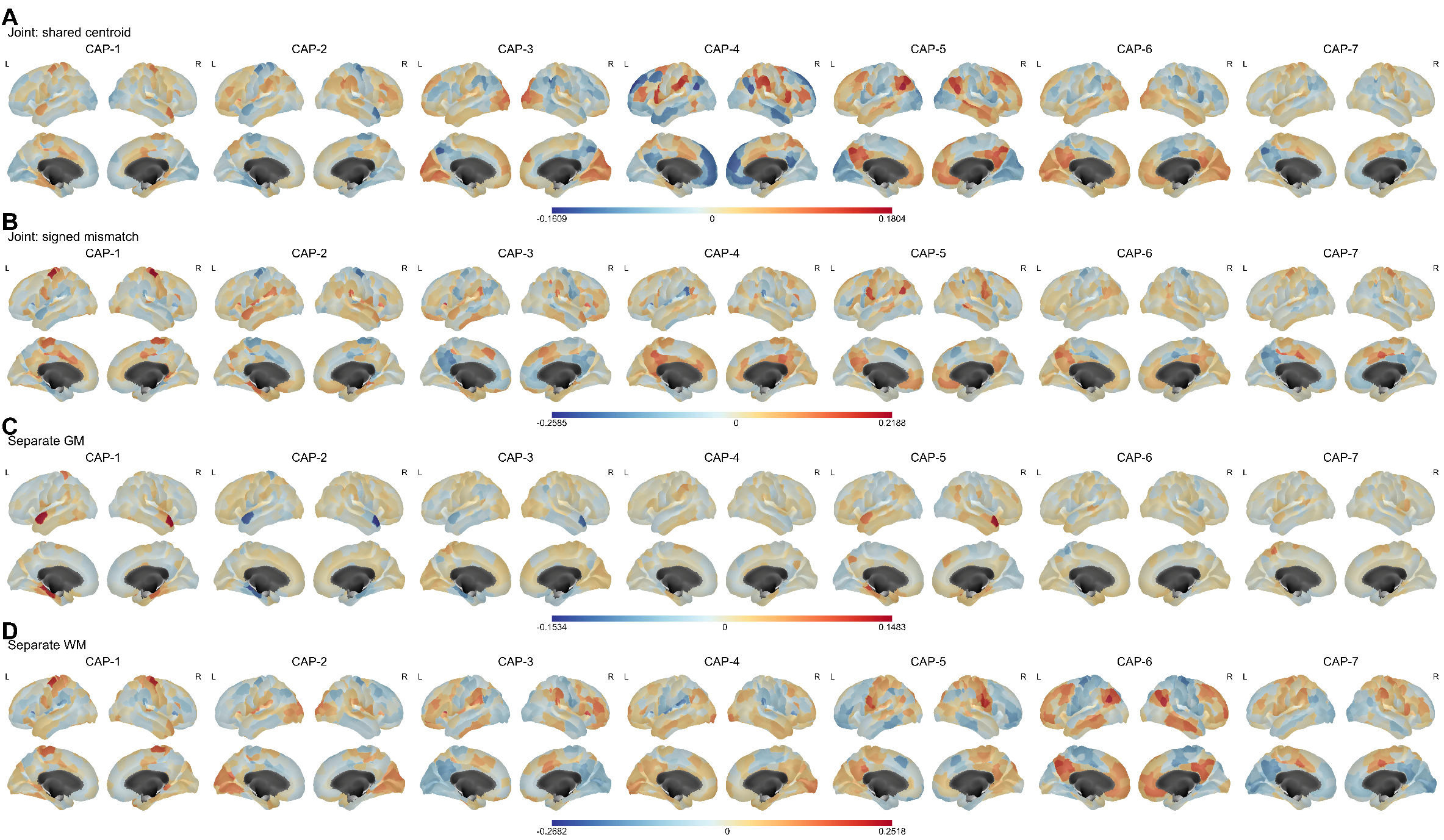

Figure S15. Spatial distribution of linear depth gradients in boundary CAP phenotypes in healthy controls (phenotype L). Maps show covariate-adjusted mean values of phenotype L in healthy controls. For each participant, phenotype L was calculated as the linear slope of the ComBat-harmonized regional measure against white matter (WM) sampling depth across 1-7 mm, expressed per millimeter. (A) Depth gradient of the shared centroid from the joint gray matter (GM)–WM model. (B) Depth gradient of the joint-model signed tissue difference, defined as standardized WM minus GM activation. (C, D) Depth gradients of activation from the separate GM and WM models, respectively. Columns represent coactivation patterns CAP-1-CAP-7. GM sampling remained fixed at 1 mm outside the GM-WM boundary. Values for the 400 Schaefer parcels are displayed on the corresponding cortical surface parcels, with lateral views above and medial views below for both hemispheres. Warm and cool colors indicate increasing and decreasing values of the corresponding measure with greater WM depth, respectively. Each panel shares one colorbar across its seven CAPs, with limits determined separately for each panel. Maps are displayed without significance thresholding.

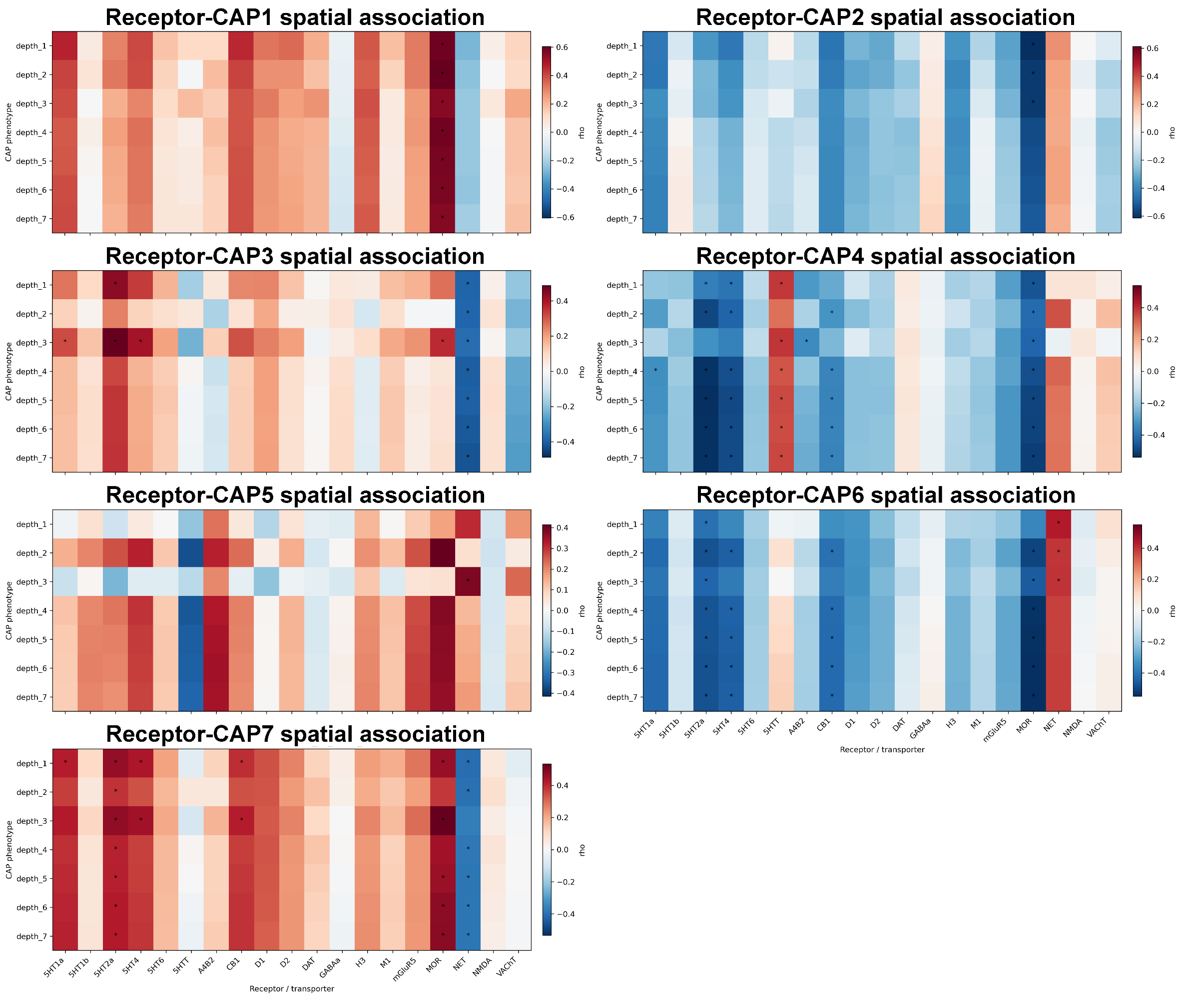

Figure S16. Spatial associations between neurotransmitter receptor maps and joint CAP activation across depths. Heatmaps show Spearman correlations (ρ) between healthy-control normative joint CAP shared-centroid maps and 19 receptor/transporter maps across 400 cortical parcels. Each panel represents one CAP, with rows indicating WM sampling depths of 1-7 mm. Red and blue indicate positive and negative correlations, respectively; color scales are panel-specific. Asterisks indicate significance based on 10,000 spatial-spin permutations and BH-FDR correction across CAP-receptor comparisons within each depth (q < 0.05).

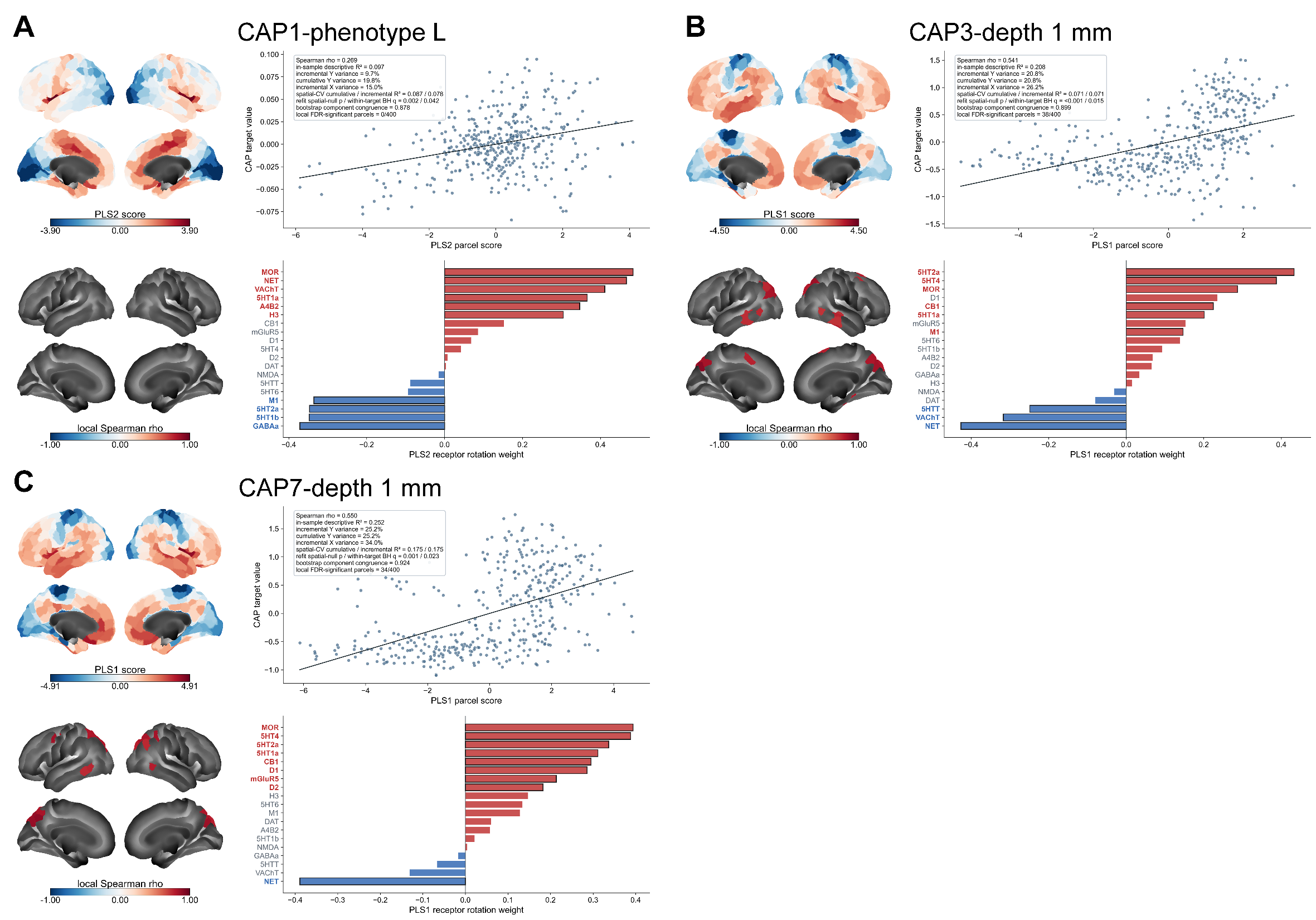

Figure S17. Additional receptor-CAP associations identified by partial least squares analysis. Results are shown for (A) CAP-1 phenotype L (linear depth slope; PLS2), (B) CAP-3 at 1 mm (PLS1), and (C) CAP-7 at 1 mm (PLS1). Within each panel, upper brain maps display parcel-wise PLS scores, and scatterplots show their association with the corresponding healthy-control normative CAP phenotype across 400 cortical parcels. Lower brain maps display local Spearman correlations surviving FDR correction (q < 0.05); no parcel survived in panel A. Bar plots show receptor rotation weights, with red and blue indicating positive and negative weights, respectively. Colored receptor labels and outlined bars indicate bootstrap FDR significance (q < 0.05). Scatterplot annotations report model fit, explained variance, spatial cross-validation, and local significance.

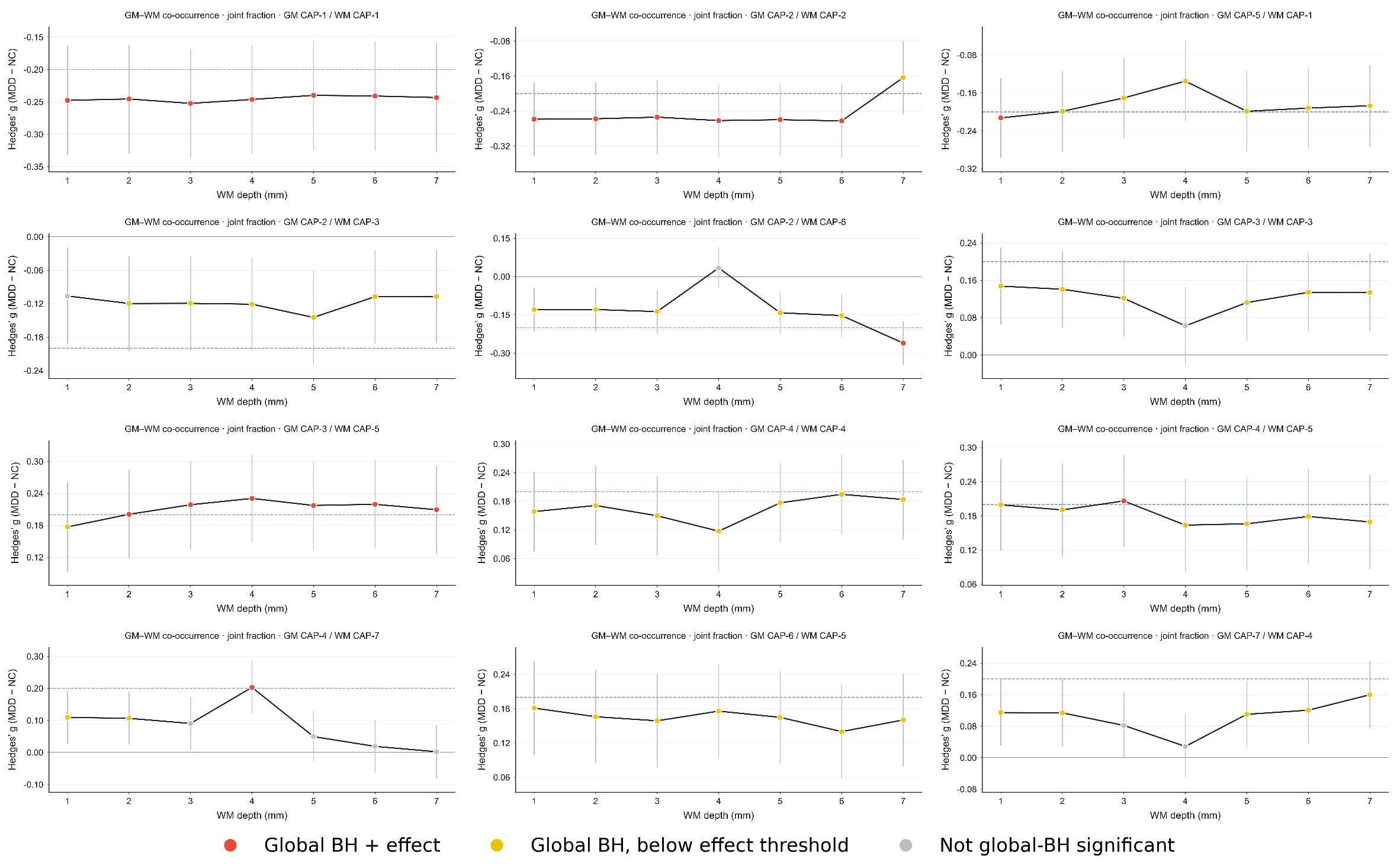

Figure S18. Cross-depth group differences in GM-WM CAP co-occurrence. Hedges’ g (MDD - NC) is shown across WM depths of 1-7 mm for selected CAP pairs. Co-occurrence denotes the fraction of frames jointly assigned to the indicated GM and WM states. Estimates were adjusted for age, sex, education, and site; error bars indicate 95% confidence intervals. Red points indicate global BH-FDR q < 0.05 and |g| > 0.20; yellow points indicate q < 0.05 and |g| ≤ 0.20; gray points indicate q ≥ 0.05. Blue dashed lines mark the ±0.20 effect-size thresholds where visible.

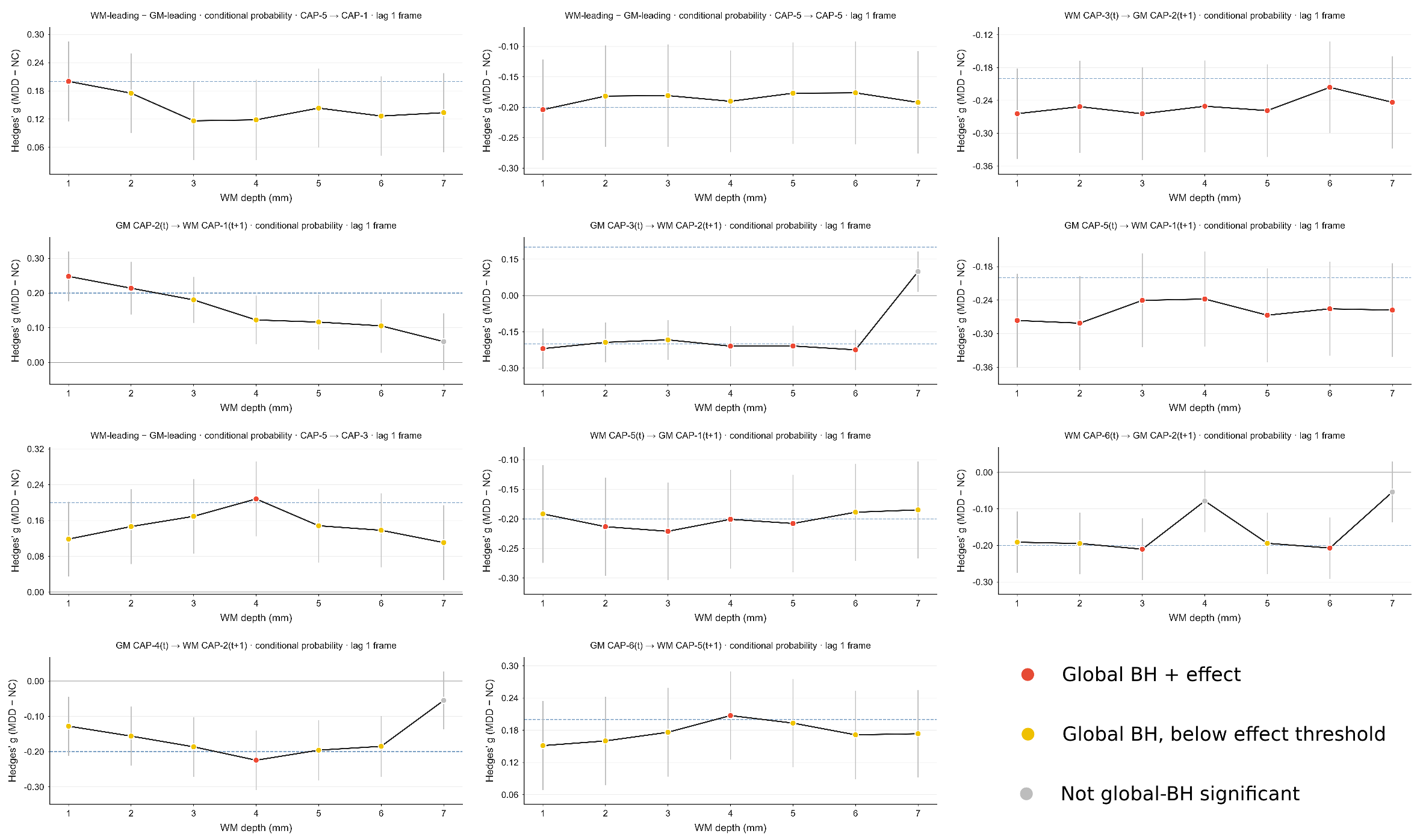

Figure S19. Cross-depth group differences in lagged GM-WM CAP coupling. Hedges’ g (MDD - NC) is shown across WM depths of 1-7 mm for selected directional conditional probabilities and their asymmetry (WM-leading minus GM-leading), evaluated at a one-frame lag. Arrows identify the source and subsequent target states. Estimates were adjusted for age, sex, education, and site; error bars indicate 95% confidence intervals. Red points indicate global BH-FDR q < 0.05 and |g| > 0.20; yellow points indicate q < 0.05 and |g| ≤ 0.20; gray points indicate q ≥ 0.05. Blue dashed lines mark the ±0.20 effect-size thresholds where visible.

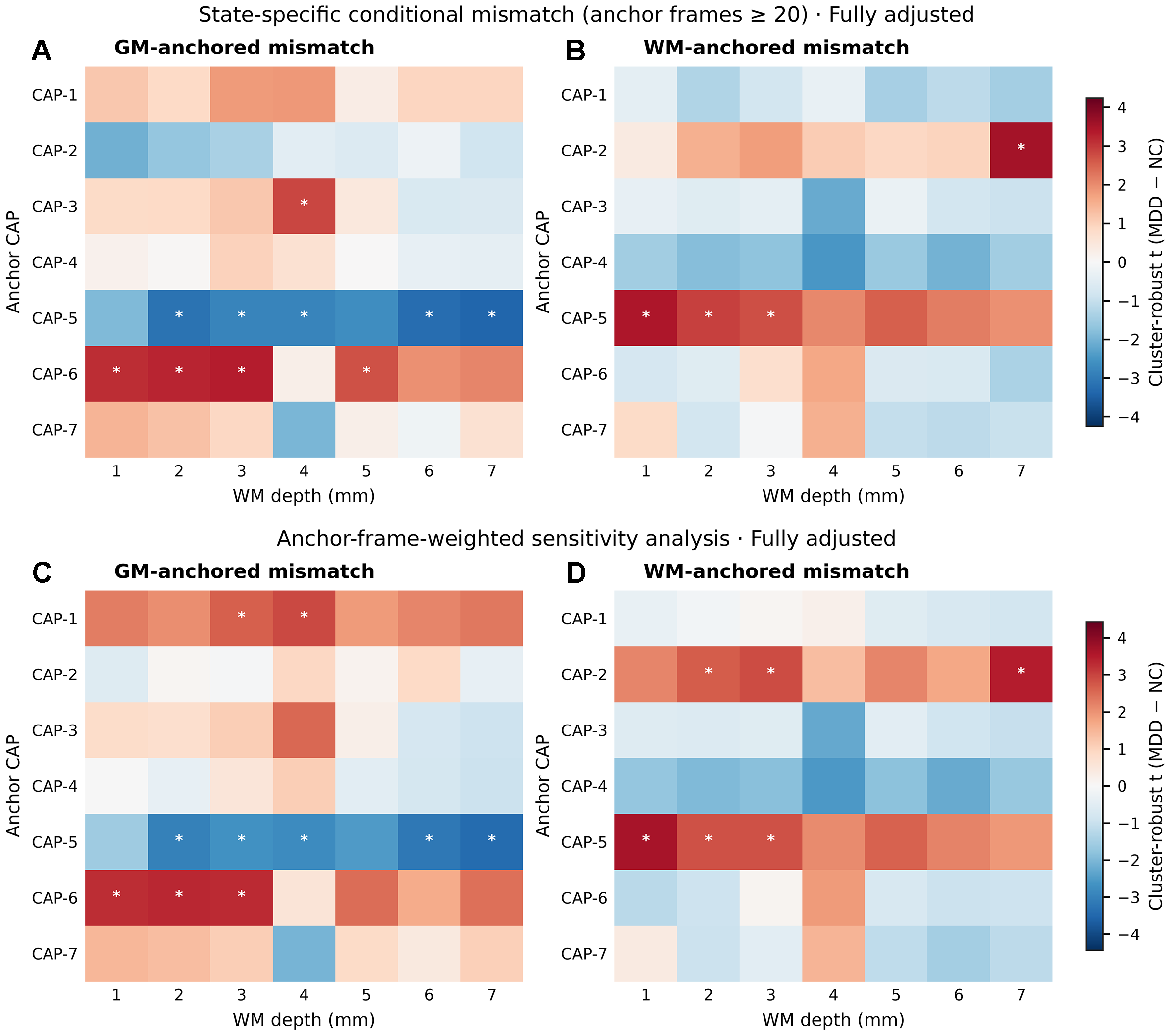

Figure S20. State-specific gray-white matter mismatch across sampling depths. Conditional mismatch denotes the probability of a nonmatching state in the other tissue given the anchor CAP. (A, B) GM- and WM-anchored analyses requiring ≥20 anchor-state frames. (C, D) Corresponding sensitivity analyses weighted by anchor-frame counts, retaining ≥1 frame and capping weights at the 95th percentile. Colors indicate cluster-robust t-statistics for MDD-NC differences, adjusted for age, sex, education, and site; red and blue indicate higher and lower mismatch in MDD, respectively. Asterisks denote Benjamini-Hochberg FDR q < 0.05 across 98 contrasts separately within each analysis.

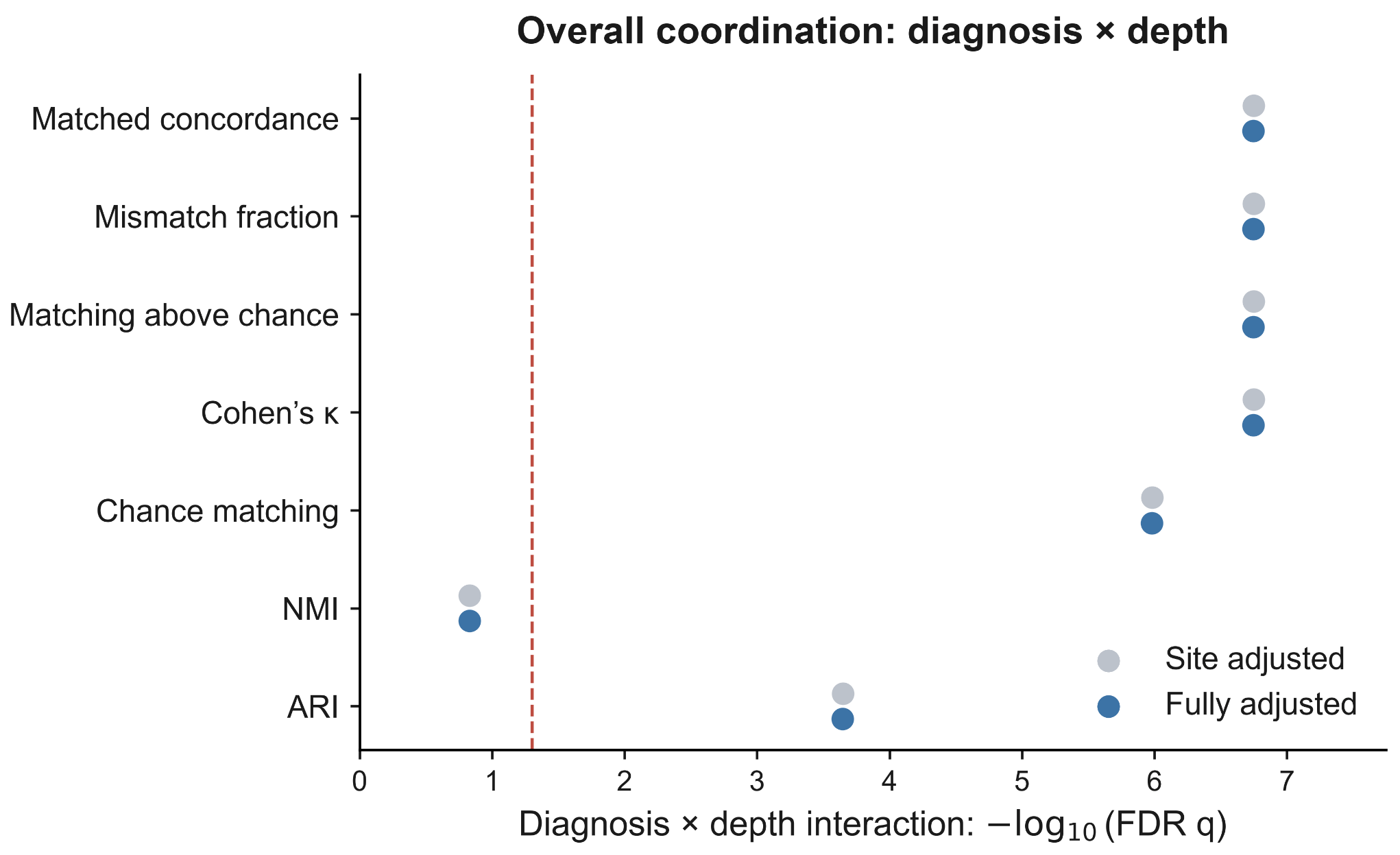

Figure S21. Diagnosis-by-depth interactions in overall gray-white matter coordination. Omnibus interaction tests across white matter depths of 1-7 mm are shown for seven coordination measures under site-adjusted and fully adjusted models. Fully adjusted models additionally include age, sex, and education. Points show -log10(FDR q), with correction across seven outcomes separately for each model; the dashed line marks q = 0.05. Participant-clustered robust covariance accounts for repeated depth measurements. NMI, normalized mutual information; ARI, adjusted Rand index.

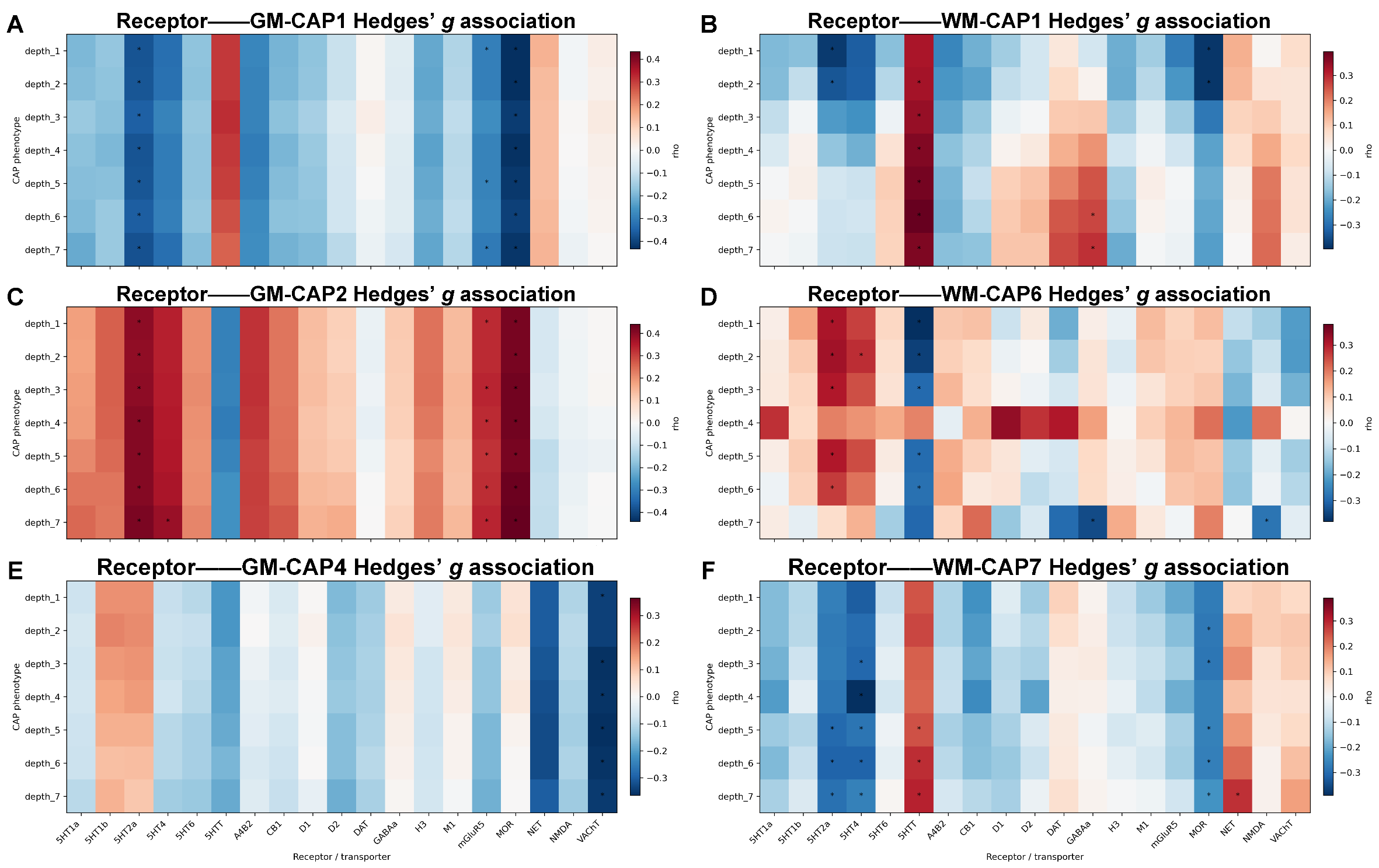

Figure S22. Depth-specific receptor associations with GM and WM CAP group differences. Heatmaps show Spearman correlations between signed Hedges’ g maps (MDD − NC) and 19 normative receptor/transporter maps across 400 corresponding parcels. Panels display (A) GM CAP-1, (B) WM CAP-1, (C) GM CAP-2, (D) WM CAP-6, (E) GM CAP-4, and (F) WM CAP-7. Rows represent WM depth conditions of 1–7 mm; GM sampling remained fixed at 1 mm outside the boundary. Group effects were adjusted for age, sex, education, mean framewise displacement, and site. Red and blue indicate positive and negative correlations, respectively; color scales are panel-specific. Asterisks indicate significance from 10,000 spatial-spin permutations with BH-FDR correction across all 7 CAPs × 19 receptors within each tissue representation and depth (q < 0.05).
